# A blood atlas of COVID-19 defines hallmarks of disease severity and specificity

**DOI:** 10.1101/2021.05.11.21256877

**Authors:** COvid-19 Multi-omics Blood ATlas (COMBAT) Consortium, David J Ahern, Zhichao Ai, Mark Ainsworth, Chris Allan, Alice Allcock, Azim Ansari, Carolina V Arancibia-Carcamo, Dominik Aschenbrenner, Moustafa Attar, J. Kenneth Baillie, Eleanor Barnes, Rachael Bashford-Rogers, Archana Bashyal, Sally Beer, Georgina Berridge, Amy Beveridge, Sagida Bibi, Tihana Bicanic, Luke Blackwell, Paul Bowness, Andrew Brent, Andrew Brown, John Broxholme, David Buck, Katie L Burnham, Helen Byrne, Susana Camara, Ivan Candido Ferreira, Philip Charles, Wentao Chen, Yi-Ling Chen, Amanda Chong, Elizabeth Clutterbuck, Mark Coles, Christopher P Conlon, Richard Cornall, Adam P Cribbs, Fabiola Curion, Emma E Davenport, Neil Davidson, Simon Davis, Calliope Dendrou, Julie Dequaire, Lea Dib, James Docker, Christina Dold, Tao Dong, Damien Downes, Alexander Drakesmith, Susanna J Dunachie, David A Duncan, Chris Eijsbouts, Robert Esnouf, Alexis Espinosa, Rachel Etherington, Benjamin Fairfax, Rory Fairhead, Hai Fang, Shayan Fassih, Sally Felle, Maria Fernandez Mendoza, Ricardo Ferreira, Roman Fischer, Thomas Foord, Aden Forrow, John Frater, Anastasia Fries, Veronica Gallardo Sanchez, Lucy Garner, Clementine Geeves, Dominique Georgiou, Leila Godfrey, Tanya Golubchik, Maria Gomez Vazquez, Angie Green, Hong Harper, Heather A Harrington, Raphael Heilig, Svenja Hester, Jennifer Hill, Charles Hinds, Clare Hird, Ling-Pei Ho, Renee Hoekzema, Benjamin Hollis, Jim Hughes, Paula Hutton, Matthew Jackson, Ashwin Jainarayanan, Anna James-Bott, Kathrin Jansen, Katie Jeffery, Elizabeth Jones, Luke Jostins, Georgina Kerr, David Kim, Paul Klenerman, Julian C Knight, Vinod Kumar, Piyush Kumar Sharma, Prathiba Kurupati, Andrew Kwok, Angela Lee, Aline Linder, Teresa Lockett, Lorne Lonie, Maria Lopopolo, Martyna Lukoseviciute, Jian Luo, Spyridoula Marinou, Brian Marsden, Jose Martinez, Philippa Matthews, Michalina Mazurczyk, Simon McGowan, Stuart McKechnie, Adam Mead, Alexander J Mentzer, Yuxin Mi, Claudia Monaco, Ruddy Montadon, Giorgio Napolitani, Isar Nassiri, Alex Novak, Darragh O'Brien, Daniel O'Connor, Denise O'Donnell, Graham Ogg, Lauren Overend, Inhye Park, Ian Pavord, Yanchun Peng, Frank Penkava, Mariana Pereira Pinho, Elena Perez, Andrew J Pollard, Fiona Powrie, Bethan Psaila, T. Phuong Quan, Emmanouela Repapi, Santiago Revale, Laura Silva-Reyes, Jean-Baptiste Richard, Charlotte Rich-Griffin, Thomas Ritter, Christine S Rollier, Matthew Rowland, Fabian Ruehle, Mariolina Salio, Stephen N Sansom, Alberto Santos Delgado, Tatjana Sauka-Spengler, Ron Schwessinger, Giuseppe Scozzafava, Gavin Screaton, Anna Seigal, Malcolm G Semple, Martin Sergeant, Christina Simoglou Karali, David Sims, Donal Skelly, Hubert Slawinski, Alberto Sobrinodiaz, Nikolaos Sousos, Lizzie Stafford, Lisa Stockdale, Marie Strickland, Otto Sumray, Bo Sun, Chelsea Taylor, Stephen Taylor, Adan Taylor, Supat Thongjuea, Hannah Thraves, John A Todd, Adriana Tomic, Orion Tong, Amy Trebes, Dominik Trzupek, Felicia A Tucci, Lance Turtle, Irina Udalova, Holm Uhlig, Erinke van Grinsven, Iolanda Vendrell, Marije Verheul, Alexandru Voda, Guanlin Wang, Lihui Wang, Dapeng Wang, Peter Watkinson, Robert Watson, Michael Weinberger, Justin Whalley, Lorna Witty, Katherine Wray, Luzheng Xue, Hing Yuen Yeung, Zixi Yin, Rebecca K Young, Jonathan Youngs, Ping Zhang, Yasemin-Xiomara Zurke

## Abstract

Treatment of severe COVID-19 is currently limited by clinical heterogeneity and incomplete understanding of potentially druggable immune mediators of disease. To advance this, we present a comprehensive multi-omic blood atlas in patients with varying COVID-19 severity and compare with influenza, sepsis and healthy volunteers. We identify immune signatures and correlates of host response. Hallmarks of disease severity revealed cells, their inflammatory mediators and networks as potential therapeutic targets, including progenitor cells and specific myeloid and lymphocyte subsets, features of the immune repertoire, acute phase response, metabolism and coagulation. Persisting immune activation involving AP-1/p38MAPK was a specific feature of COVID-19. The plasma proteome enabled sub-phenotyping into patient clusters, predictive of severity and outcome. Tensor and matrix decomposition of the overall dataset revealed feature groupings linked with disease severity and specificity. Our systems-based integrative approach and blood atlas will inform future drug development, clinical trial design and personalised medicine approaches for COVID-19.

## Introduction

The pathophysiology associated with severe acute respiratory syndrome coronavirus 2 (SARS-CoV-2) reflects a complex interplay between virus induced lung pathology and maladaptive host immune responses (Kuri-Cervantes et al., 2020; Mathew et al., 2020; Tay et al., 2020; Vabret et al., 2020). Severe COVID-19 is characterised by hypoxia, with the risk of rapid deterioration which may require intensive care support and, in some patients, progression to acute respiratory distress syndrome, multiorgan failure and death. Predisposing factors include age, gender, ethnicity, obesity and comorbidities. Currently, opportunities for biomarker-led timed and targeted precision medicine approaches are limited by an incomplete understanding of pathogenesis and heterogeneity among patients with severe disease (Wynants et al., 2020). A dysregulated hyperinflammatory state occurs in some individuals (Moore and June, 2020), consistent with reported benefits from glucocorticoids (dexamethasone) and inhibitors of the IL-6 receptor (tocilizumab/sarilumab) and Janus kinases (baricitinib) in severe disease (Gordon et al., 2021; Horby et al., 2021a; Horby et al., 2021b; Kalil et al., 2021). Nevertheless, blood-derived signatures of COVID-19 severity are diverse, including evidence of immune suppression, myeloid dysfunction, lymphopenia, interferon driven immunopathology, T cell activation as well as exhaustion, and immune senescence (Bost et al., 2021; Chen and Wherry, 2020; Diao et al., 2020; Hadjadj et al., 2020; Mann et al., 2020; Schulte-Schrepping et al., 2020). In the lung, widespread neutrophil and macrophage infiltration, T cell cytokine production and alveolitis are seen with features of altered redox balance, endothelial damage and thrombosis (Grant et al., 2021). Here, through the COvid-19 Multi-omics Blood ATlas (COMBAT) consortium, we characterise COVID-19 of varying severity and establish shared and specific features in comparison with severe influenza and non-SARS-CoV-2 sepsis. We demonstrate the informativeness of data-driven, systems biology approaches to identify cells, mediators and pathways that are hallmarks of increasing severity, and to define potential therapeutic targets and biomarkers of the variable individual response to SARS-CoV-2 infection.

## Results

### Clinical features, severity metrices and disease stratification in COVID-19

To characterise the peripheral blood response to COVID-19 we analysed a prospective cohort of adult patients with confirmed SARS-CoV-2 presenting to clinical services at the start of the United Kingdom pandemic (February-March 2020). We recruited 116 hospitalised COVID-19 patients following informed consent at a single site (Oxford University Hospitals) (Table S2). The overall mortality rate was 23.3%. Samples were collected during the acute admission and, in survivors from 28 days after discharge (convalescent samples). We compared these patients with community COVID-19 cases in the recovery phase (never admitted to hospital), age-matched healthy volunteers, influenza cases requiring mechanical ventilation (critically ill receiving intensive care), and all-cause sepsis patients (hospitalised encompassing severe and critical disease) recruited prior to the pandemic (Figure 1A; Table S2; Star Methods).

**Figure 1.**
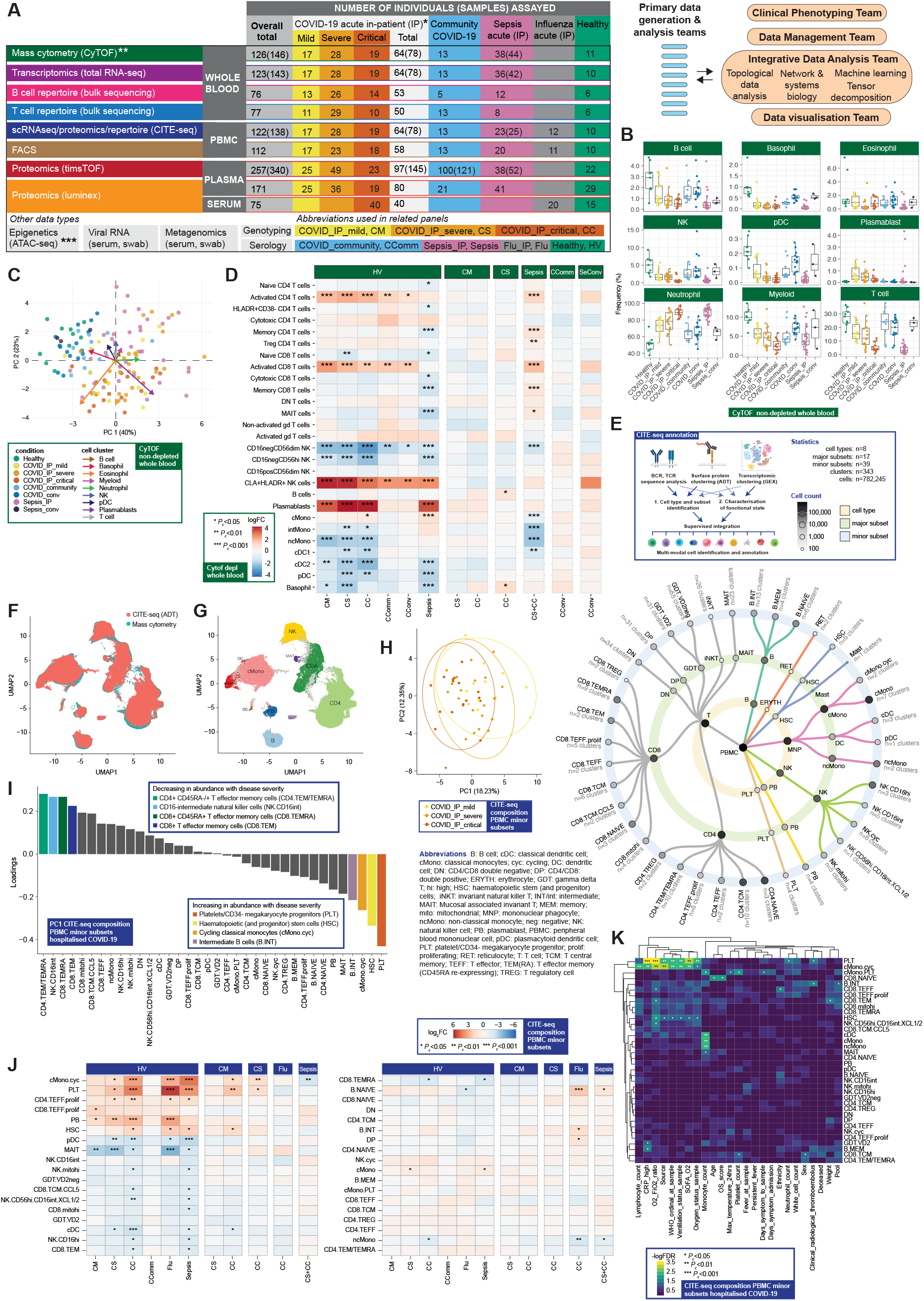
Complementary single cell compositional approaches reveal variance in specific cell populations by clinical group and severity. (A) Study design, assay modalities and workflow. Table shows number of patients assayed, with number of samples in brackets where more than one sample assayed. *WHO severity categories show number of patients at time of sampling **single paired convalescent sample assayed for n=16 COVID-19 and n=3 sepsis patients; ***10 samples assayed (8 samples for paired acute-convalescent COVID-19 and 2 healthy). (B-G) Major cell populations in whole blood for clinical comparator groups assayed by single cell mass cytometry (CyTOF) showing (B,C) for non-depleted samples (3,893,390 cells) (B) cell frequencies (C) PCA with arrows indicating drivers of variation by cell population and (D) differential abundance analysis of CD66^+^ depleted whole blood (7,118,158 cells assayed) by patient group (E) multimodal identification and annotation of PBMC subpopulations based on curated intersection of CITE-seq data (F,G) UMAP showing joint visualisation of CITE-seq and CyTOF datasets (using CITE-seq data as reference) (F) overlaid CITE-seq (ADT) and mass cytometry (G) cell annotations transferred between datasets and coloured by cell type where concordant (94.5% of cells) or discordant (grey). (H-K) CITE-seq compositional analysis of minor subsets (H,I) PCA for hospitalised COVID-19 cases (H) PC1 vs PC2 (I) analysis of loadings of minor subsets on PC1. (J) Differential abundance analysis between clinical groups (K) Covariate analysis for clinical, demographic and experimental variables for hospitalised COVID-19 cases (with BH adjusted ANOVA test for significance). See Figure S1.

We computed a correlation matrix of all clinical features for our hospitalised COVID-19 cohort, including severity scores and surrogate markers of illness response, to inform rational use in downstream cellular and molecular phenotyping (Figure S1A-C; Table S2; Star Methods). There were positive correlations (spearman’s rho >0.5) between markers of severity based on WHO severity scales, oxygenation status, ventilation status, Sequential Organ Failure Assessment (SOFA) oxygenation score, vasopressor use, length of intensive care unit (ICU) stay, days on mechanical ventilation, C-reactive protein (CRP) and neutrophil count (Figure S1C). We further investigated covariance of patient and clinical features using unsupervised machine learning to understand clinically derived patient groupings (Star Methods). We found that the optimal cluster number for hospitalised cases was 2 or 3, and these showed broad concordance to the first released WHO categorical criteria, namely mild (no requirement for supplemental oxygen), severe (oxygen saturation SaO_2_ <=93% on air but not requiring mechanical ventilation) and critical (requiring mechanical ventilation) (Figure S1D). This clustering finding persisted when we restricted the analysis to acute measures of physiology and clinical biomarkers, where the main correlates of consensus clustering related to ventilation status (Figure S1E-F). Accordingly, hereinafter we refer to WHO categorical criteria as our primary hospitalised COVID-19 severity comparator groups.

### COVID-19 severity is associated with differences in abundance of diverse immune cell populations

We first investigated changes in cellular composition associated with COVID-19 severity. In this differential abundance analysis, we used one sample per patient. For patients with more than one time point of sampling, the sample closest to onset of maximal disease as defined by clinical features at the time of sampling was selected. In this ‘prioritised sample set’, we found increased neutrophil, and reduced T and B lymphocytes, measured in terms of both relative and absolute counts, in patients with more severe/critical disease, together with reduced myeloid, dendritic cell (DC), natural killer (NK) cell and basophil populations when we analysed whole blood using mass cytometry (Figures 1A,B and S1G-J; STAR Methods; Methods S1). These changes were also seen in patients with severe non-SARS-CoV-2 sepsis. Plasmablasts were increased in COVID-19 (Figure 1B). In community cases and convalescent COVID-19 samples, the abundance of major cell types was broadly comparable to healthy volunteers although differences in neutrophil and mononuclear phagocyte cell frequency persisted. To reduce dimensionality and identify correlates of variance in the data between patient groups, we performed principal component analysis (PCA). This showed separation along PC1 driven by the frequency of neutrophils and plasmablasts (increased) and of plasmacytoid DCs (pDC) and basophils (reduced) (Figure 1C).

To characterise lymphocyte and mononuclear phagocyte populations with greater resolution, we performed mass cytometry after granulocyte (CD66^+^) depletion for a second aliquot from the same blood draw (Star Methods). COVID-19 infection was associated with differential abundance of specific populations of monocytes, NK cells and plasmablasts, together with activated and cytotoxic CD8^+^ T cells (Figure 1D). We validated and further characterised these differences using Cellular Indexing of Transcriptomes and Epitopes by Sequencing (CITEseq) (Figure 1A). We annotated peripheral blood mononuclear cell (PBMC) types, subsets and clusters by combining information from single-cell RNA-sequencing (scRNAseq, GEX), cell surface protein quantification (192 antibody panel) and B/T cell V(D)J repertoire profiling (Figure 1E; STAR Methods; Methods S2). These annotations showed a high predicted concordance with those from mass cytometry (Figures 1F,G and S1K,L,M). We found that cellular composition differed by patient group and by severity in hospitalised COVID-19 patients (Figures 1H-K and S1N-Q). Performing PCA for all groups, we found that PC1, which explained 21.5% of the variance, was associated with group membership (*P_c_* = 2.01×10^-15^, ANOVA) (Figure S1N; Methods S2). When hospitalised COVID-19 cases were analysed alone, the first PC was associated with group membership, oxygenation status (SaO_2_/FiO_2_ ratio, SOFA oxygenation, and ventilation and oxygen status), and lymphocyte count (all *P_c_* <0.05) (Figure 1H; Methods S2). The cell subsets with the largest negative loadings on PC1 (higher abundance in more severe disease) were platelets/CD34^-^ megakaryocyte progenitors, haemopoietic (and progenitor) stem cells (HSCs), and cycling classical monocytes (cMono.cyc) (Figure 1I).

Consistent with these findings, empirical Bayes analysis of differential abundance also demonstrated significant differences in the same cell populations in COVID-19 compared with age/gender-matched healthy volunteers, together with reduced DC, T, and NK cell subsets, particularly in COVID-19 critical cases (Figure 1J). Among hospitalised COVID-19 cases, we found that a higher abundance of platelets/CD34^-^ megakaryocyte progenitors and cycling cMono was associated with oxygenation status (SaO_2_/FiO_2_, SOFA oxygenation score, ventilation status), severity (WHO ordinal), and CRP (all *P_c_* <0.01). Higher platelet/CD34^-^ megakaryocyte progenitor abundance was also associated with the occurrence of thromboembolism during hospitalisation (*P_c_* = 0.036, ANOVA) (Figure 1K).

### Whole blood hallmarks of COVID-19 and severity involving neutrophils, progenitor cells, lymphocyte exhaustion, clotting, immunoglobulin and the interferon response

We next defined global signatures of the host response to COVID-19 by performing whole blood total RNA-sequencing (RNA-seq) (Figure 1A; STAR Methods). We investigated overall variance in gene expression using the prioritised sample set. The first two PCs separated samples by cohort, from healthy volunteers through increasing severity among COVID-19 cases and patients with sepsis (Figure 2A). We then restricted the analysis to hospitalised COVID-19 patients. Patients differed by severity group including 28-day mortality on PC1 (P=2.34×10^-6^, Kruskal-Wallis test) (Figure 2B), which also correlated with other measures of severity and differential cell count (Figure S2A). Topological data analysis, using the mapper algorithm with PC1 as the filter function, captures the geometric structure of the high-dimensional data and shows the connectivity of COVID-19 cases, with colour corresponding to a gradient of severity (Figure 2B; Star Methods). We found that genes with the highest loadings for PC1 were strongly enriched for immune system function, notably neutrophil degranulation (fold change (FC) 4.23, FDR 2.4×10^-15^), PD-1 signaling (FC 21.5, FDR 9.2×10^-12^) (consistent with lymphocyte exhaustion), antimicrobial peptides (FC 10.8 FDR 9.2×10^-7^), and clotting cascade (FC 10.6 FDR 2.1×10^-5^). PC2 gene loadings showed strong enrichment for interferon signaling (FC 10.7, FDR 3×10^-33^) including key viral response network genes (*IFI1-3*, *IFI6*, *IFI44*, *IFIT3*, and *OAS1-3*) and specific immunoglobulin heavy and lambda genes (Figure S2B).

**Figure 2.**
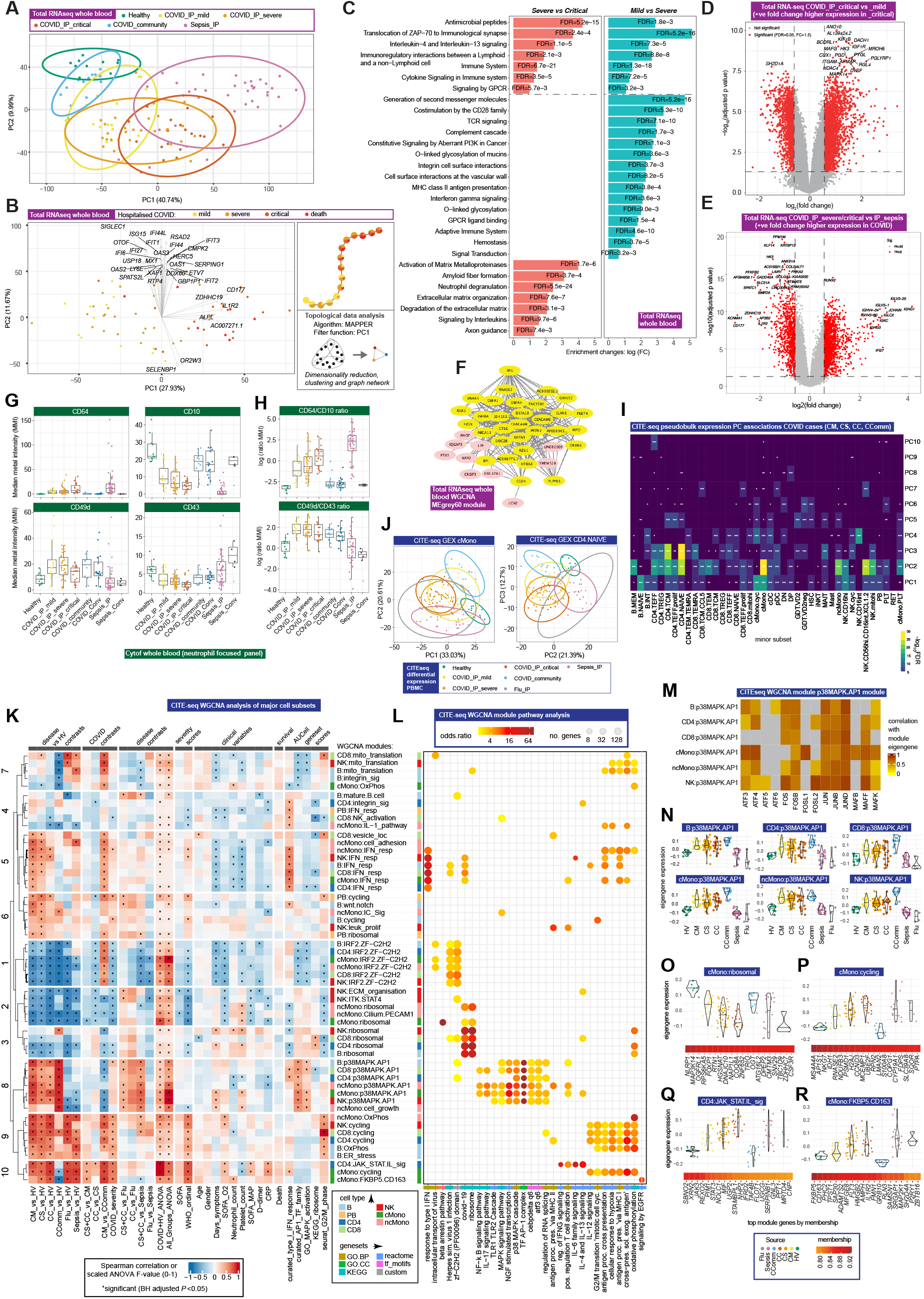
Signatures of COVID-19 response from transcriptomics. (A-E) Whole blood total RNAseq (A,B) PCA (A) all clinical comparator groups (B) hospitalised COVID-19 patients including topological data analysis (C) pathway enrichment by severity of COVID-19 (D,E) differential gene expression (D) critical vs mild COVID-19 (E) COVID-19 vs sepsis. (F) Intramodular hub genes for weighted gene correlation network analysis (WGCNA) whole blood RNAseq module grey60. (G,H) Neutrophil cell surface proteins assayed by mass cytometry shown by (G) marker (H) ratio of markers. (I,J) CITEseq gene expression analysis of PBMC (I) association of PCs of expression variance with minor subset cell clusters in patients with COVID-19 (J) PC plots of gene expression in classical monocytes and naive CD4+ T cells. (K-V) WGCNA CITEseq gene expression for major cell types (K) association of module eigengenes with disease contrasts, clinical severity scores and variables, survival and gene set scores (*all significant associations shown, BH adjusted *P* < 0.05 in individual tests) (L) pathway enrichment (M) module eigengene correlation with AP-1 family genes (N) p38MAPK.AP-1 module eigengene expression across patient groups (O-R) eigengene expression and top eigengene-gene correlations (O) ribosomal module in cMono (P) cycling module in cMono (Q) JAK-STAT.interleukin module in CD4 (R) FKB5.CD163 module in cMono. See Figure S2.

Among hospitalised COVID-19 patients, the greatest distinction between mild and more severe disease involved T cell receptor (TCR) signaling, and for severe vs critical disease, neutrophil degranulation, activation of metalloproteases, antimicrobial peptides and interleukin signaling (Figure 2C,D). When we incorporated cell proportion into the differential expression model, the number of differentially expressed genes was reduced but the fold changes from both models remained well correlated (between critical and mild disease), with comparable pathway enrichment on analysis of severity as a quantitative trait (Figure S2C,D). In terms of specificity of the COVID-19 response, we found this was largely distinct from non-SARS-CoV-2 sepsis of comparable severity, as shown by PCA (Figure 2A). Specific features of COVID-19 compared to sepsis included a relative upregulation in COVID-19 of many immunoglobulin heavy/kappa/lambda genes, and unique pathway enrichments relating to cell proliferation and innate/adaptive immune function (Figures 2E and S2E).

Next, we identified modules of genes correlated with COVID-19 severity using weighted gene correlation network analysis (WGCNA) (Star Methods). We found the three modules that were most significantly correlated with severity (P<1×10^-10^); genes in these modules were enriched for, respectively, cellular and functional neutrophil gene signatures and neutrophil count (MEblue module); CD8^+^ T cell signatures and relative lymphopenia (MEturquoise module); and granulocyte and common myeloid progenitor cell gene signatures, neutrophil degranulation, antimicrobial peptides and defensins pathways (MEGrey60 module) (Figure S2F-J). The MEgrey60 module was more highly expressed in critical COVID-19 than sepsis (Figure S2J), and in this module the ETS transcription factor related gene *ERG,* which regulates lineage plasticity, showed the highest intramodule connectivity (Figure 2F). These features support the MEgrey60 module representing a mixture of progenitor cells that associate with severity, consistent with the large severity associated loadings seen for progenitor cell abundance in CITEseq (Figure 1H,I).

To better characterise the neutrophil populations in COVID-19, we then applied a myeloid-marker enriched mass cytometry panel to the same samples (Table S2, STAR Methods). We found evidence for the presence of immature neutrophils and neutrophil progenitors (pro-neutrophils) based on high expression of CD64 (Fc gamma receptor 1) and CD49d (integrin alpha 4), and decreased expression of CD10 (neutral endopeptidase) (Figure 2G) (Evrard et al., 2018; Kwok et al., 2020; Marini et al., 2017). CD64 expression was raised in severe/critical COVID-19 and further elevated in sepsis, together with increased PD-L1 (CD274) expression (Figure S2K). Using CD64:CD10 ratio as an index score for immature neutrophil presence we found association with the MEblue module eigengene that correlated with neutrophil count and function (Figure S2L). We further determined that neutrophil CD49d expression was elevated, while CD43 (leukosialin) was reduced, in COVID-19 patients but was largely unchanged in sepsis (Figure 2G). The CD49d:CD43 ratio remained high in convalescence (Figure 2H).

### Shared and cell-type-specific gene expression signatures of COVID-19 include those related to ZFN, ribosomal and cell-cycle genes and AP-1 and IFN signaling

We proceeded to investigate gene expression signatures in patients with COVID-19 using profiles from the CITEseq minor cell subsets (STAR Methods, Methods S2). PCA of all hospitalised and community COVID-19 cases (PBMC, prioritised sample set), revealed variance in gene expression across a wide range of cell subsets: cycling classical monocytes (cMono.cyc), naïve B cells and CD16^high^ NK cells contributed most to the first PC while lower PCs were associated with classical monocytes (cMono), memory B cells, CD4^+^ T cells (naïve, effector and effector memory), effector memory CD8^+^ T cells, NK cells (CD56^high^CD16^int^, mitochondrial^high^) and non-classical monocytes (ncMono) (Figure 2I). Inclusion of sepsis, influenza and healthy volunteers yielded similar results (Figure S2M) with cMono, plasmablasts, CD4^+^ T cells (naïve, effector and effector memory) and cycling NK cells contributing most to discriminating between groups by PCA (as exemplified in Figure 2J).

We then identified sets of differentially expressed genes between patient cohorts and tested their enrichment for curated pathways in major cell types (Figure S2N; STAR Methods) (Liberzon et al., 2015). Type I and type II interferon pathways were up-regulated in the less severe hospitalised COVID-19 patients across cell types. Redox state (reflected by MTORC1 signalling and oxidative phosphorylation) pathways were enriched across mononuclear phagocytes (MNP), T cells, NK cells and plasmablasts in more severe COVID-19, as were cell cycle (MYC, E2F targets, G2M checkpoint) pathways (except for MNP) while IL2-STAT5 pathway enrichment was found in more severe disease for T cells.

To further deconvolute biological pathways and cellular functions associated with COVID-19, we investigated gene expression for major cell subsets by applying WGCNA to the CITEseq dataset (all comparator groups) (Star Methods). Analysis of module co-variation identified five distinct module sets, shared to varying extents between CD4^+^ and CD8^+^ T cells, NK cells, B cells, plasmablasts, cMono and ncMono (Figures 2K and S2O; Methods S2). First, we found a set of type I IFN response modules with module eigengenes correlated with milder disease, better oxygenation status and earlier sampling from symptom onset across cell populations (Figure 2K,L). The second module set, discovered in all cell types except plasmablasts, showed strong enrichment for activator protein 1 (AP-1) (FOS, JUN, ATF family genes) and the p38MAPK cascade. The module eigengenes were highly expressed in all COVID-19 patient groups, including recovery phase community cases, were distinct from influenza and sepsis, and did not show a consistent relationship with severity or other clinical features (Figure 2K-N). The third module set (found in all cell types except plasmablasts) was enriched for classical (C2H2) zinc finger (ZNF) genes and contained *IRF2* and *IL16*. Expression of these eigengenes was lower in COVID-19 and influenza compared with healthy volunteers and sepsis cases (Figure 2K,L). The fourth set of modules showed enrichment for ribosomal proteins and the KEGG ‘COVID-19’ pathway; the top genes by membership relate to inflammasome function (including *NLRP1*, *MAP3K14* and *FOXP1)* and were negatively correlated with COVID-19 severity in monocytes (Figure 2K,L,O). Finally, we found a set of “cycling” modules based on pathway enrichments and membership of *MKI67* and *TOP2A*. These modules showed a weaker correlation across cell type. The cMono cycling module was enriched for stem cell differentiation and regulation of granulopoiesis, correlated with severity, and module genes included *S100A8/9* which encodes calprotectin, a known severity biomarker (Silvin et al., 2020) (Figure 2K,L,P). We also identified two cell-type-specific modules sharing very similar associations to severe disease. These comprised of a JAK-STAT/interleukin signaling module in CD4^+^ T cells, and an EGFR pathway enriched module in cMono (Figure 2K,L,Q,R). The top gene members of the EGFR module included *FKBP5,* a factor involved in stress response and glucocorticoid receptor sensitivity, and *CD163*, a scavanger receptor modulating induced innate immune response (Figure 2K,L,R).

### Transcriptomic and epigenetic signatures of severity in monocyte populations

Using the detail afforded by our high-resolution multi-modality dataset, we further investigated signatures of severity in specific mononuclear phagocyte populations. Consistent with the PCA loadings (Figure 1I), we observed an overall relative increase in the frequency of cMono and reduced intermediate monocytes (CD16^+^CD14^+^), ncMono (CD14^-^CD16^+^) and dendritic cells (pDC, cDC1 and cDC2), in hospitalised COVID-19 patients with more severe disease using mass cytometry (prioritised sample set) (Figure 3A,B) (Star Methods). With increasing disease severity we observed a shift in the phenotype of cMono to lower expression of HLA-DR, CD33 and CD11c, and evidence of proliferating monocytes based on expression of Ki67 and DNA abundance, with comparable changes in the sepsis patients (Figures 3A-C and S3A,B). Lower levels of pDCs and CD33^low^cDC2 were found in sepsis compared with severe/critical COVID-19 (Figure 3C).

**Figure 3.**
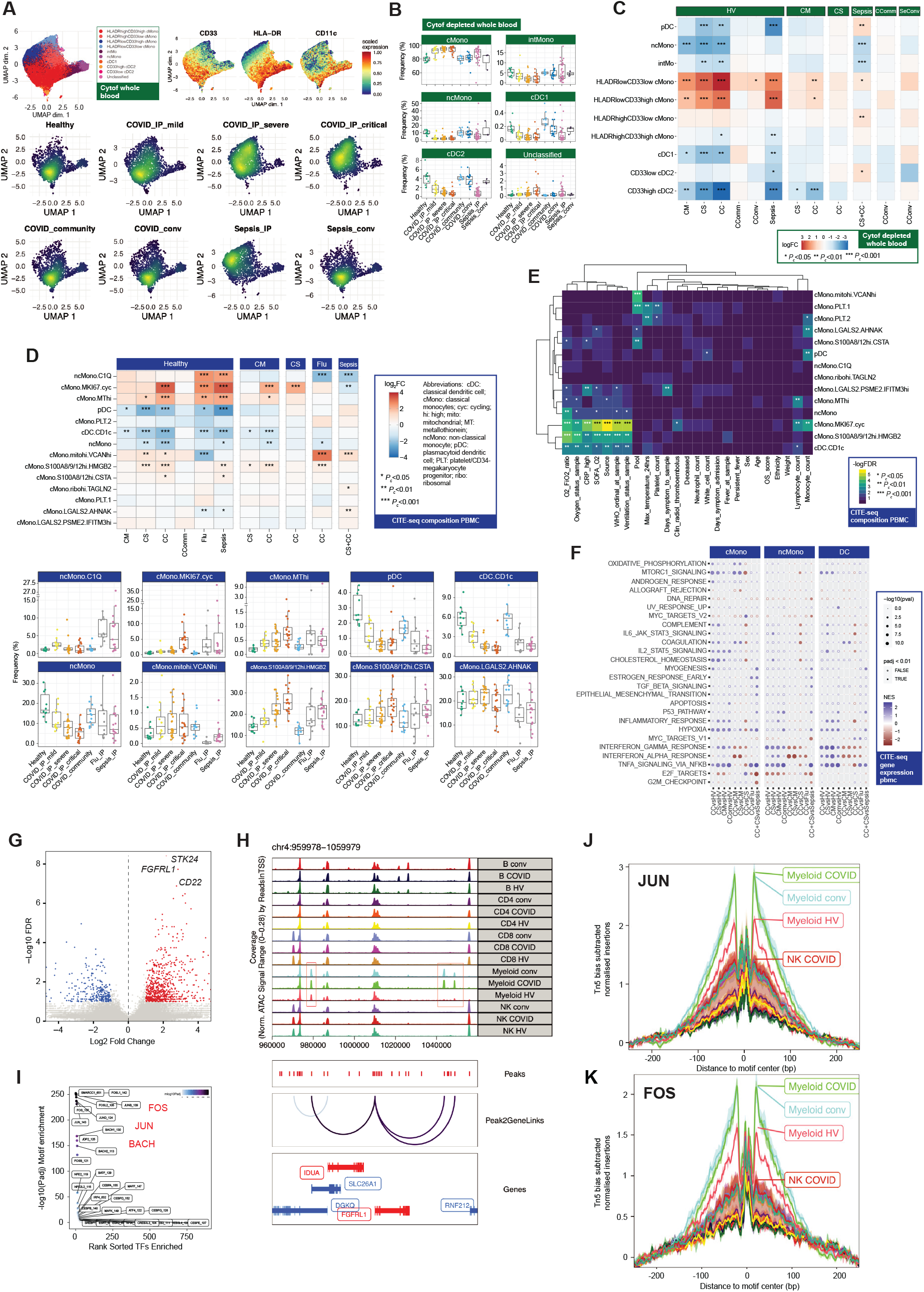
Changes in myeloid cells associated with COVID-19 severity. (A-C) Single cell mass cytometry (A) UMAP of overall myeloid cell clusters coloured by patient group and by Mean Metal Intensity (MMI) of HLA-DR, CD33 and CD11c (B) cell population frequencies by patient group (C) differential abundance analysis in patients vs healthy volunteers, and between disease categories (D-F) CITE-seq PBMC myeloid cell clusters (D-E) cell composition (D) differential abundance analysis with box plots of cell cluster frequency by patient group where abundance significantly differs relative to healthy volunteers (E) covariate analysis of abundance and clinical, demographic and experimental variables for hospitalised COVID-19 cases (with BH adjusted ANOVA test for significance) (F) scRNAseq MSigDB hallmark gene set enrichment for cMono, ncMono and DC. (G-K) scATACseq (G) differential chromatin accessibility in myeloid cells comparing acute COVID-19vs healthy volunteers (H) scATACseq tracks at *FGFRL1* locus comparing cell populations and condition (healthy, COVID-19 acute and convalescent) (I) differential motif enrichment in myeloid cells, acute COVID-19 vs healthy volunteers (J,K) transcription factor footprinting for myeloid enriched factors (J) JUN (K) FOS. See Figure S3.

Using CITEseq (prioritised sample set) (Star Methods) we found cycling cMono, and cMono with high expression of the anti-oxidant metallothionein genes (MT^hi^), were significantly elevated in critical COVID-19 cases, influenza and sepsis compared to healthy volunteers (Figure 3D).

*S100A8/9/12*^hi^ *HMGB2*-expressing cMono correlated with COVID-19 severity, and were also increased in sepsis. cMono expressing *VCAN*, which is implicated in adhesion and cytokine release, were specifically increased in COVID-19 and reduced in influenza, whilst complement component *C1Q*-expressing ncMono were increased in influenza and sepsis, but not in COVID-19. Galectin-2 (*LGALS2*) and desmoyokin (*AHNAK*) expressing cMono were significantly reduced in influenza and sepsis but not in COVID-19. pDCs showed reduced abundance in more severe COVID-19, influenza and sepsis, as did CD1c^+^ cDCs. Consistent with the progressive changes in abundance according to COVID-19 severity, the frequencies of CD1c^+^ cDCs, and most significantly cycling cMono and *S100A8/9/12*^hi^ *HMGB2* expressing cMono, were associated with clinical variables relating to oxygenation and respiratory function in hospitalised cases (Figure 3E).

In terms of gene expression, the PCs that explained the greatest variance between hospitalised COVID-19 cases in MNP, specifically cMono and cDC, were associated with severity measures including WHO ordinal scale and ventilation status (Figure S3C,D). In addition to the IFN, cycling and EGFR pathways highlighted by WGCNA analysis, pathway enrichment analysis of differentially expressed genes in cMono showed enrichment for inflammatory response/TNF signaling in milder disease, hypoxia and IL2_STAT5 pathways across severity groups, and complement, coagulation and cholesterol metabolism in more severe disease (Figures 3F and S3E,F).

We proceeded to further investigate potential mechanistic drivers of disease by characterising the chromatin state of myeloid and other PBMC populations in COVID-19 patients using single-cell ATAC-seq (Figures 1A and S3G,H; Star Methods). Overall, 750 and 303 accessible sites were up- and down-regulated respectively in COVID-19 patients compared to healthy volunteers in myeloid cells (Figure 3G). Genes linked to top differentially open chromatin peaks included *STK24* (MAPK promoting apoptosis) and *FGFRL1* (cell adhesion promoting fibroblast growth factor receptor) (Figure 3H). We identified specific DNA binding motif enrichments in the differentially accessible sites. The most significant enrichments were found for AP-1, SW1/SNF and BACH transcription factor family members which are involved in chromatin remodelling and immunity (Figure 3I). In line with this observation, motif footprint analysis revealed an increased accessibility of genomic regions containing FOS and JUN motifs in COVID-19 patients relative to healthy volunteers in myeloid cells, a signal which was also seen in convalescence (Figure 3J,K).

### Severe COVID-19 is associated with clonal expansion of unmutated B cells, and activation of autoreactive B cells

We next determined the relationship of B cell composition, gene expression and the B cell receptor (BCR) repertoire with COVID-19 severity. Mass cytometry of whole blood (prioritised sample set) showed significant lymphopenia in COVID-19 with reduced overall frequency and number of B cells, predominantly naïve B cells, but an increase in terminally differentiated plasmablasts (significantly higher than in sepsis) (Figures 1B and 4A and S4A,B; STAR Methods). The greatest increase in switched memory CD11c^+^ B cells was in community COVID-19 cases while unswitched memory B cells and naïve CD11c^+^ B cells were higher in COVID-19 convalescent samples (Figure 4A). We observed a relatively high proportion of CLA^+^ plasmablasts in COVID-19 patients (Figure 4A) as previously observed in patients with respiratory infections (Seong et al., 2017). Analysis of GEX/ADT defined clusters revealed significant increases in plasmablasts in severe disease (Figures 4B-D and S4C,D; STAR Methods). Naïve CD1c^+^ naïve and cycling naïve B cells were reduced in COVID-19 but overall naïve B cells were significantly more reduced in influenza than COVID-19 of comparable severity (Figures 4D and S4C). In mild hospitalised COVID-19, only the IFN-responsive naïve B cell cluster increased significantly (Figure 4D).

**Figure 4.**
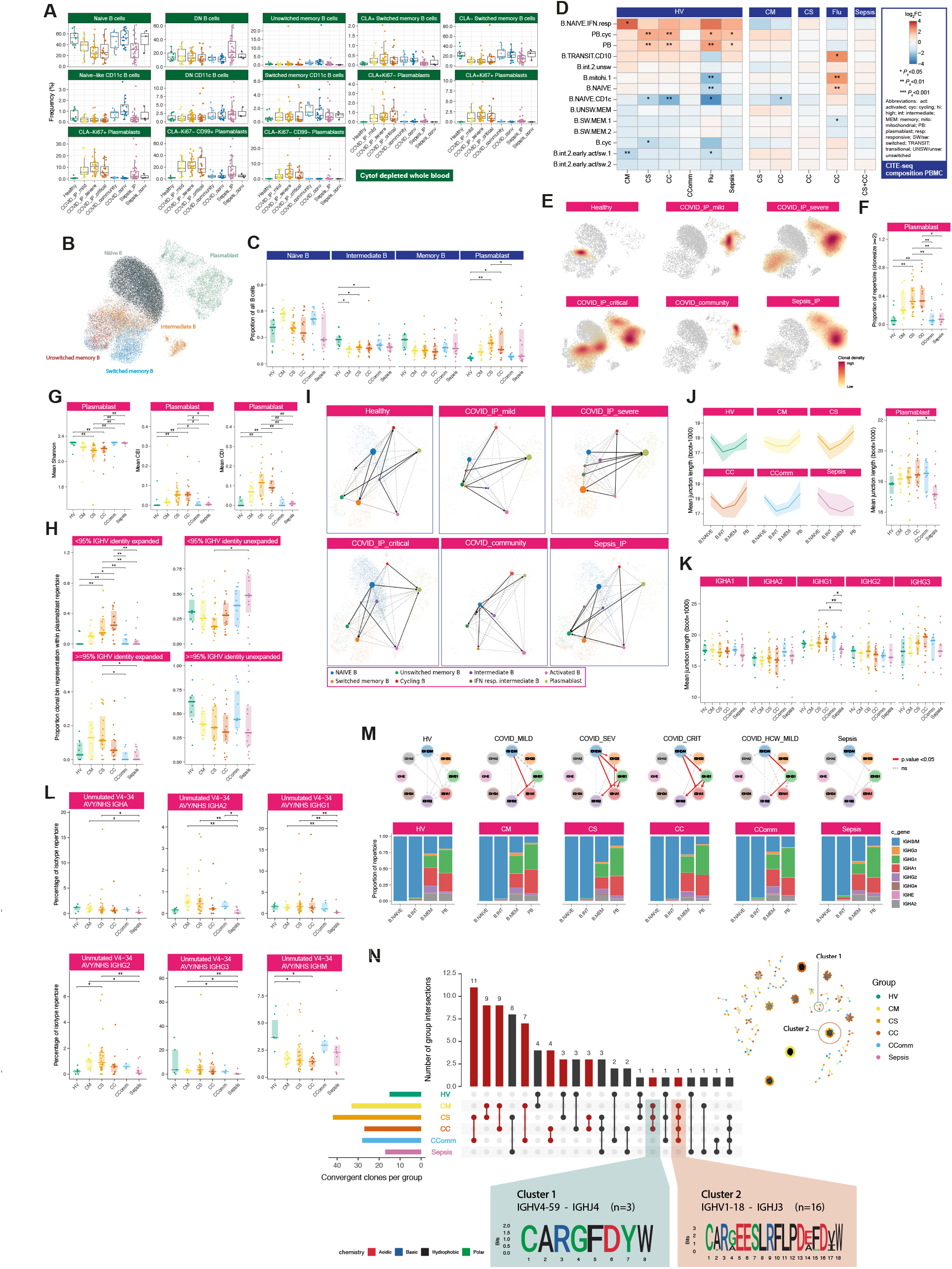
B lymphocytes show changes in composition and repertoire associated with COVID-19 severity. (A-D) Cell composition analysis comparing study groups (A) by single cell mass cytometry (B-D) by CITE-seq clustering (B) UMAP embedding with cluster identities (C) cluster proportions and (D) differential abundance. (E) Clonal density plots with Kernels density estimates overlaid onto UMAP embeddings by comparator group. (F) Plasmablast repertoire clonality. (G) Mean plasmablast diversity indices by comparator group; clonal expansion index, CEI, clonal diversification index, CDI. (H) Mutation and expansion proportions in plasmablast clone repertoire. (I) Partition-based graph abstraction (PAGA) plots of scRNA-seq by cell population and patient group (weighted edges indicate degree of cell cluster connectivity). (J,K) Junction lengths from resampled repertoires by patient group (J) per B cell cluster (left) and specifically in plasmablasts (right) (K) for immunoglobulin constant genes (L) IGHV4-34 AVY/NHS motif usage in unmutated VDJ sequences shown across IGH genes (bulk BCR-seq). (M) Class switch inference networks (RNA derived B cell repertoires) (top) Ig constant region genes per B cell cluster (single cell VDJ data) (bottom). (N) Clonal overlap across comparator groups with total number of convergent clones per group shown with clonal network depicting distribution of convergent clusters (inset) and sequence logos of COVID19 exclusive clusters (with IGHV/J usage and number of members). Significance * <0.05, ** 0.005 Kruskal Wallis. See Figure S4.

We then characterised differences involving the B cell immune repertoire using bulk VDJ sequencing of whole blood (Fig 1A) (1,206,531 filtered BCR sequences analysed) and single cells (CITEseq) (STAR Methods). As expected, plasmablasts showed the highest BCR expression in the CITEseq dataset (Figure S4E). Whereas in healthy volunteers clonal expansions appeared to predominate within the memory B cell population, in COVID-19 and sepsis patients, expansions were seen in plasmablasts, with severe and notably critical COVID-19 patients also harbouring clones within memory populations (Figure 4E). The clonal expansion in plasmablasts was statistically significant, and also showed a significant association with COVID-19 severity, in contrast to sepsis in which there was no significant change (Figures 4F,G and S4F).

Limited somatic hypermutation (SHM) of SARS-CoV-2 antibodies has been widely reported (Brouwer et al., 2020). We observed fewer somatic hypermutations in intermediate B cells in hospitalised mild COVID-19 but a severity-associated increase in number was found in plasmablasts and also seen in sepsis (Figure S4G). There were, however, COVID-19 specific differences in the proportion of expanded clones with few mutations (>95% IGHV identity, Figure 4H). RNA velocity analysis suggested a differentiation directionality between naïve B cells and plasmablasts in COVID-19 patients that was distinct from sepsis, and consistent with a predominant extrafollicular B cell response in COVID-19 (accumulating fewer SHMs) (Figure 4I). Moreover, we observed a higher number of shared clones between plasmablasts and intermediate or memory B cells in severe/critical COVID-19 patients, whereas sepsis patients exhibited higher clonal overlap between intermediate and memory B cells (Figure S4H,I).

These data together indicate that in severe/critical COVID-19 there is substantial expansion of unmutated B cells associated with plasmablast populations. We next explored the accompanying differences in B cell selection and tolerance. Firstly, in COVID-19 patients we observed increased BCR complementarity-determining region 3 (CDR3) lengths compared to sepsis (Figure 4J,K); such increases have been associated with antibody polyreactivity and autoimmunity (Meffre et al., 2001), notably within plasmablasts and IgG1^+^ B cells. Secondly, we found multiple differentially utilised IGHV/J genes between COVID-19 groups indicating differential B cell selection and/or expansion of naïve B cells, while the antigen experienced IgD/M mutated and class-switched B cell repertoire showed differentially utilised IGHV/J genes, revealing differential peripheral selection of B cells with increasing COVID-19 severity (Figure S4J-M). Thirdly, we tested whether B cells targeting autoantigen and red blood cell antigen are associated with COVID-19. We found that autoreactive IGHV4-34 BCRs, which are elevated in autoimmunity (Pascual et al., 1991), were significantly depleted in IGHD/M but elevated in class-switched B cells, most notably for the IGHA2 and IGHG2 B cells (Figure 4L) consistent with class-switching of these autoreactive B cells during the response to SARS-CoV-2. In further support of this observation, the degree of class-switching, inferred from the BCR sequencing data (Bashford-Rogers et al., 2019), was significantly elevated between IgD/M and IgG1 and IgA1, and finally to IgG2 in COVID-19 patients (Figure 4M). No detectable differences in either IGHV4-34 autoreactive BCR levels or class-switching were observed in sepsis cases.

Previous reports indicate an unexpectedly high level of BCR convergence between unrelated COVID-19 patients (Galson et al., 2020). We also found clonal sharing within and between COVID-19 severity groups (Figures 4N and S4N-P). Comparing to known receptor-binding domain antibodies, we observed that most highly similar patient BCRs have a plasmablast phenotype (Figure S4N). Overall, our data indicate that the plasmablast expansions in severe COVID-19 include high levels of broadly auto-reactive B cells, consistent with an emerging role for B-cell driven immune pathology (Wang et al., 2020).

### COVID-19 severity correlates with specific T and NK cell populations and features relating to cell cycle, redox state and exhaustion

We proceeded to investigate T and NK cell function in COVID-19 and its relationship with severity. Whole blood mass cytometry showed that both activated CD4^+^ and CD8^+^ T cells were increased in frequency in all COVID-19 patient groups, remaining elevated in convalescence (Figure 5A; STAR Methods). The proportion of CD27^+^ activated CD4^+^ T cells was higher than in sepsis, while CD56^+^ cytotoxic CD8^+^ T cell frequency was reduced (Figure S5A). We characterised the T cell subsets using markers of activation, proliferation and exhaustion. While these markers were comparably expressed in activated CD4^+^ T cells across acute COVID-19 cases, they increased in CD8^+^ T cells with disease severity (Figure 5B). Multicolour flow cytometry showed differential chemokine receptor expression in the overall memory CD4^+^ T cell population (Figure S5B; STAR Methods) and increased expression of the inhibitory receptor TIM3 in activated CD8^+^ T cells (Figure S5C). CLA^+^ HLADR^+^ NK cells were increased in all COVID-19 cases (Figure S5D). We also found evidence of significant changes in innate-like lymphocytic cell populations with increasing COVID-19 severity including MAIT cells, with evidence from mass cytometry of a gradient of involvement across severity in terms of cell activation (% CD69^+^ MAIT cells, Figure 5C).

**Figure 5.**
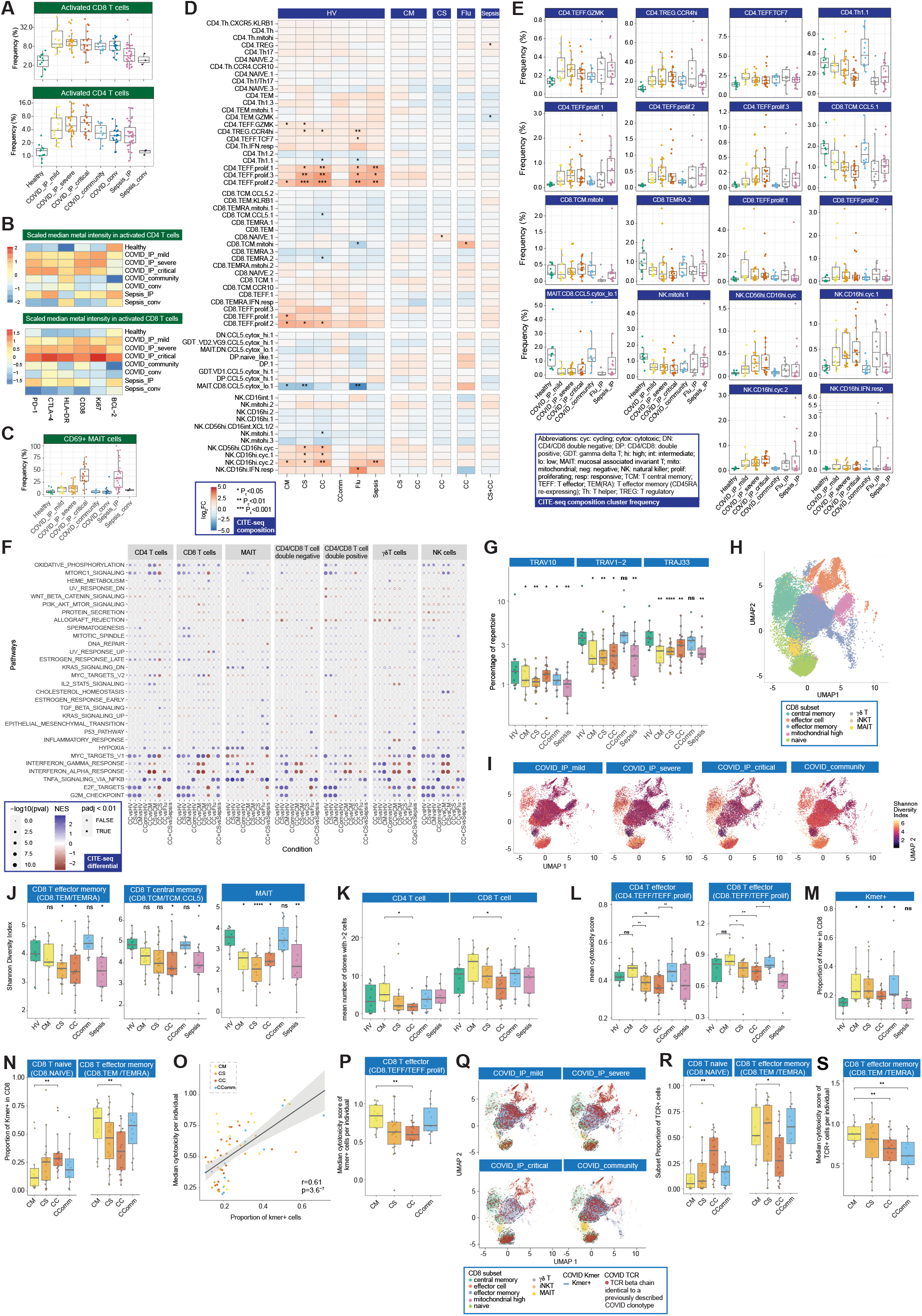
Dynamic changes in T lymphocyte and NK composition and repertoire associated with COVID-19 severity. (A-C) Single cell mass cytometry whole blood (A-B) activated CD4 and CD8 T lymphocytes (A) frequency (B) median metal intensity of specific markers (C) frequency of activated MAIT cells (D,E) CITEseq profiling CD4^+^ CD8^+^ T and NK cell clusters (D) differential abundance and (E) frequency between comparator groups. (F) scRNAseq MSigDB hallmark gene set enrichment for T cell populations (G) TRAV and TRAJ repertoire analysis. (H,I) UMAP of CD8^+^ T cells and associated clusters (H) used in repertoire analysis (I) Shannon Diversity Index of CD8^+^ T cells by patent group. (J) Boxplots of Shannon Diversity Index for specific cell populations by comparator group. (K) Number of enlarged clones by comparator group in CD4^+^ and CD8^+^ subsets. (L) Mean cytotoxicity score by comparator group. (M) Proportion of CD8^+^ T cells carrying TCR containing COVID-19 associated Kmers. (N) Frequency of COVID-19 Kmer positive cells in CD8^+^ naïve and effector memory cells. (O) Correlation of COVID-19 Kmer containing CD8^+^ T cells per individual with median cytotoxicity score. (P) Cytotoxicity of CD8^+^ T effector cells positive for a COVID-19 associated Kmer across patient groups. (Q) UMAP of CD8^+^ T cells by patient group indicating density of COVID-19 Kmer positive cells (blue dashed line) and cells with previously described COVID-19 clonotype. (R) Proportion of COVID-19 known clonotype matching cells in CD8^+^ naïve and effector memory cells. (S) Cytotoxicity of CD8^+^ T effector memory cells with clonotypes matching published COVID-19 clonotypes. Wilcoxon Test age and sample size adjusted linear model used *P<0.05, **P<0.01, ***P<0.001. See Figure S5.

Complementing these findings, our analysis of the CITEseq data showed an increase in cycling and activated CD4^+^ and CD8^+^ T and NK cell populations in hospitalised COVID-19 cases, including CCR4^hi^ Tregs (CD4.TREG.CCR4hi) (Figure 5D,E; STAR Methods). Conversely, we observed a decrease in CD4^+^ Th1, CCL5^+^ CD8^+^ T central memory (CD8.TCM.CCL5), CD45RA^+^ CD8^+^ T effector memory (CD8.TEMRA), and NK cells with high mitochondrial gene expression (NK.mitohi). There was little compositional variation in these cell populations by severity or clinical covariates (Figure S5E). However, we found that, in the gene expression analysis of hospitalised COVID-19 patients, the largest component of variance (PC1) was associated with oxygenation status in CD4^+^ T effector (CD4.TEFF) and CD8^+^ T effector memory (CD8.TEM) cells (Figure S5F-H). Activated NK cells (CD56^high^CD16^low^ *XCL1/2* expressing) (NK.CD56hi.CD16int.XCL1.2) showed a strong association with WHO ordinal and oxygenation/ventilation status (Figure S5I). In terms of differential gene expression pathway enrichment, we found cell cycle and redox state pathways were enriched in more severe hospitalised COVID-19 cases across CD4^+^, CD8^+^ and NK cells; interferon pathways were enriched in less severe disease; and TNF signaling was most enriched in community cases vs healthy volunteers (Figure 5F). MAIT cells showed enrichment for TNF signalling and KRAS across COVID-19 groups while γδ T cells showed a greater enrichment for cell cycle pathways (Figure 5F).

### Reduced diversity in CD8^+^ T cell populations on repertoire analysis

To further investigate the effect of disease on T cell subsets, with reference to antigen recognition and clonality, we integrated TCR sequencing data performed across the same cell subsets. Given we saw clonotypes present across populations, we merged subsets to provide power for downstream clonal analysis (STAR Methods). For semi-invariant T cells, differences observed with severity and disease group by cell cluster were supported by consistent changes in TCR alpha variable (TRAV) gene usage. We found that hospitalized COVID-19 and sepsis cases displayed significant reductions in the percentage of repertoire occupied by TRAV10, specific to invariant natural killer T cells, and TRAV1-2 and TRAJ33 usage, in keeping with reductions in MAIT cells (Figure 5G).

To better understand the relationship between COVID-19 and T cell clonality, we calculated Shannon diversity indices across clones based on the beta chain, controlling for age. While CD4^+^ subsets showed higher diversity than CD8^+^ subsets, differences with disease severity were only seen in CD8^+^ T cells (Figures 5H,I and S5J,K; STAR Methods). Across disease states, accounting for age, CD8^+^ T effector memory (CD8.TEM/TEMRA), CD8^+^ T central memory (CD8.TCM/CD8.TCM.CCL5) and MAIT cell diversity was reduced in COVID-19 severe and critical disease with comparable changes in sepsis (Figure 5J).

Recent evidence suggests that effective CD8^+^ T cell responses involve increased numbers of expanded clones (Fairfax et al., 2020). Consistent with this, we found that hospitalised COVID-19 patients with mild disease had higher numbers of expanded clones in both CD4^+^ and CD8^+^ subsets (Figure 5K) and the mean clone size was higher within the CD8^+^ subset (Figure S5L). In keeping with the observation that expanded CD8 T cell clones show increased expression of cytotoxicity markers (Watson et al., 2020a), using a composite gene score for cytotoxicity we found that the number of expanded clones correlated with the average cytotoxicity score across all cells for that individual in both CD4^+^ T effector (CD4.TEFF/TEFF.prolif) and CD8^+^ T effector (CD8.TEFF/TEFF.prolif) populations (Figure S5M), and was higher in mild and community COVID-19 cases with reduced cytotoxicity observed in critical and severe disease (Figure 5L).

To further explore whether COVID-19 leads to generalized signatures of antigen presentation with reciprocal effects on TCR sequence and corresponding CDR3 usage, we devised an approach to identify COVID-19 associated amino acid (aa) sequences (Kmers of 4 aa) within the beta chain CDR3 region (STAR Methods). These were compared with chains from healthy volunteers and sepsis patients to exclude sequences non-specifically associated with infection (Figure S5N). We identified 125, 4 aa Kmers (referred to as COVSeqs) enriched in COVID-19 (Bonferroni corrected *P* <0.05 versus both groups), the vast majority being observed in CD8^+^ T cells (Figure S5O), with the proportion of cells with TCRs containing at least one COVSeq in the beta chain specifically increased in all COVID-19 patients (Figure 5M). In hospitalised patients we found a lower proportion of CD8^+^ T effector memory cells with COVSeq containing TCRs with increasing disease severity (Figure 5N). Critical disease was associated with naïve CD8^+^ T cells containing COVSeqs, indicating failure of the SARS-CoV-2 reactive cells in critical patients to expand into the effector phenotype, or possibly a distinct redistribution of the expanded cells.

Further supporting functionality of the COVID-19 Kmer containing cells, we observed a significant correlation between the proportion of COVSeq containing cells and the median cytotoxicity of cells per individual within the COVID-19 patients (Figure 5O). Notably, COVSeq positive CD8^+^ T effector cells from critical patients showed significantly reduced cytotoxicity compared to those from patients with mild disease (Figure 5P).

Finally, we addressed whether using previously published COVID-19 associated beta chain clonotypes could further resolve variation in the T cell response according to disease severity (STAR Methods). We observed many cells carrying such TCRs across the COVID-19 patients, often overlapping COVSeq-containing cells. Notably, the distribution of these cells across clusters varied markedly according to COVID-19 disease state (Figure 5Q). Replicating the observations with COVSeq-positive cells, CD8^+^ T effector memory cells were relatively depleted for COVID-19 clonotypes in critical disease (Figure 5R,S).

### Correlates of severity and disease specificity in the COVID-19 plasma proteome involve acute phase proteins, metabolic processes and markers of tissue injury

We aimed to complement our multi-modal cellular profiling with analysis of the COVID-19 plasma proteome. To do this we performed high-throughput liquid chromatography with tandem mass spectrometry (LC-MS-MS), producing data for 105 proteins on 257 individuals (340 samples) after QC (Figure 1A; Table S2; STAR Methods). We found differences by severity and aetiology on unsupervised hierarchical clustering (Figure S6A), PCA (Figure 6A) and supervised correlation analysis (Figure S6B). Severe disease, reflected in PC loadings (Figure 6B), was associated with increased acute-phase proteins and complement system proteins, including recognised biomarkers of inflammation (SAA1, SAA2 and CRP), complement membrane attack complex components (C5, C6, C9 and CFB), and functionally related protein families such as protease inhibitors (SERPINA3, SERPINA1 and ITIH3) and serum amyloid P-component (APCS) (Figure S6C). We also found differential protein abundance involving markers of tissue injury and necrosis, notably reduced extracellular actin scavenger plasma gelsolin (GSN); increased fibrinogens (FGA, FGB and FGG); and an increase in proteins implicated in IL-6 mediated inflammation (LGALS3BP, LRG1, LBP, HP and ITIH4). We further identified protein clusters based on the protein-protein interaction network, including a large cluster enriched for biological processes involving cholesterol transport and fibrin blood clots within which individual proteins showed positive and negative correlations with PC1 (disease severity) and two smaller clusters enriched for cytolysis and complement activation, both showing negative correlations for all constituent proteins with PC1 (Figure 6C), thus positively correlating with disease severity.

**Figure 6.**
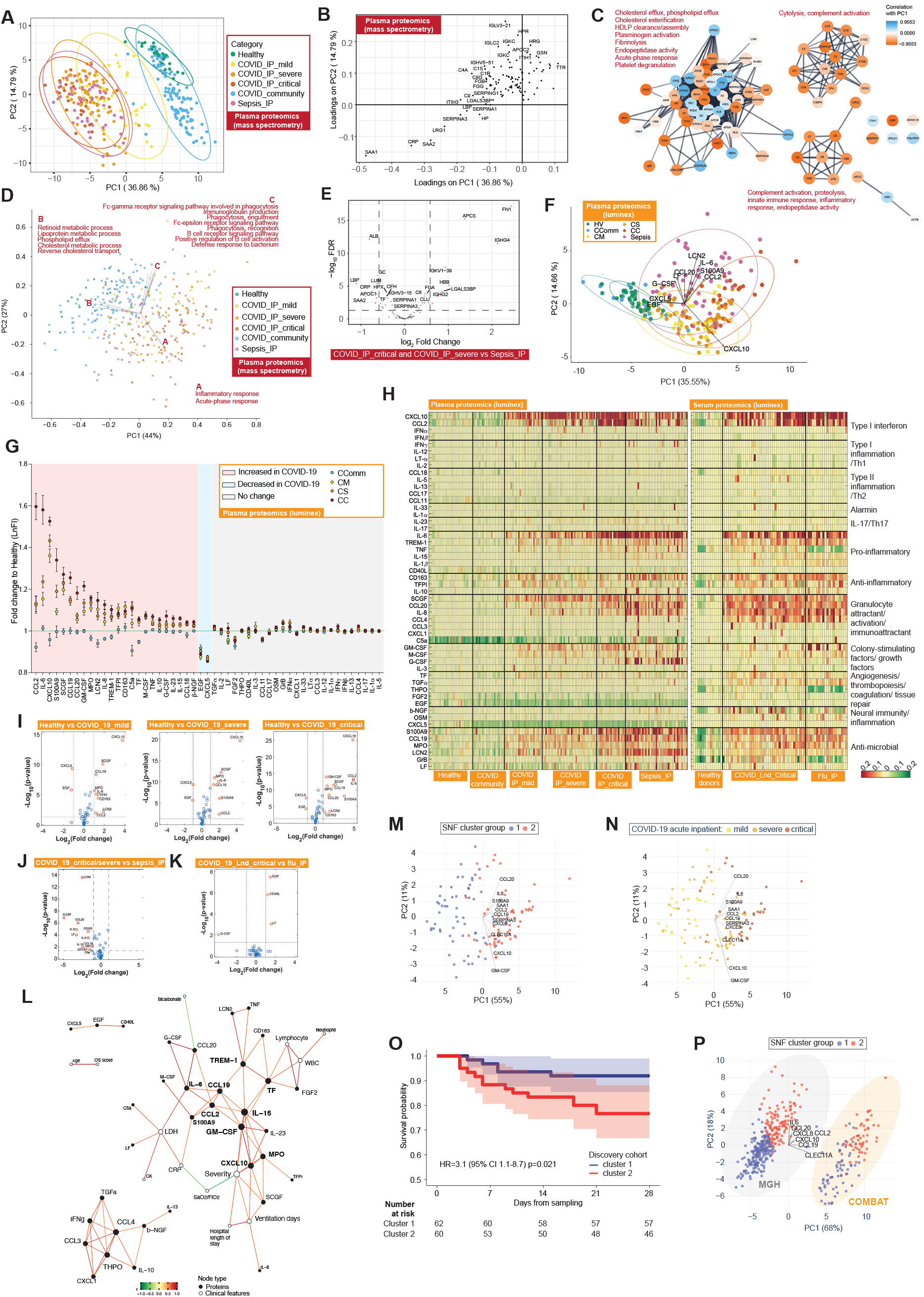
Plasma protein COVID-19 signatures and subphenotypes. (A-E) HT-LC-MS/MS mass spectrometry of plasma proteins (A) PCA all samples (B) proteins contributing to PC loadings, the more negative loading values indicating higher positive correlation with disease severity (C) clusters based on the protein-protein interaction network with enriched GOBP terms (D) functional PCA (E) differential protein abundance COVID-19 severe or critical vs sepsis. (F-L) Luminex blood proteins (F) PCA all plasma samples (G) summary of differential abundance of plasma proteins (H) heat map showing abundance of plasma and serum proteins by disease groups (I-K) Volcano plots comparing differential abundance of plasma proteins (I) COVID-19 severity groups vs healthy volunteers (J) critical/severe COVID-19 vs sepsis (K) critical COVID-19 vs influenza (L) network of clinical feature−protein correlations in COVID−19 patients and healthy volunteers based on highly correlated events (r^2^ >0.7 or <-0.5). (M-O) Similarity network fusion for hospitalised COVID-19 patients (SNF) from COMBAT (discovery) data using networks derived from sample-by-sample similarity matrix for mass spectrometry and luminex assays of plasma proteins. Coloured by (M) cluster group (N) severity. (O) Kaplan-Meier survival plot by SNF cluster group (95% CIs shaded) (HR, hazard ratio calculated using Cox proportional hazard model). (P) Mass General Hospital (Olink) validation data and COMBAT (discovery) cohorts showing groups. See Figure S6.

We proceeded to a functional PCA, generating a vector of biological process enrichment scores from single-sample Gene Set Enrichment Analysis (ssGSEA) derived from ranked intensities of the identified proteins. This revealed the main processes associated with differences between samples were acute-phase response and inflammation, metabolic (retinoid and lipoprotein) and cholesterol transport (Figure 6D). Reduced levels of proteins associated with lipoprotein and cholesterol metabolism included apolipoproteins A-I, A-II, C-I and C-II (APOA1/2 and APOC1/2) and transthyretin (TTR), consistent with their downregulation in systemic inflammation and differences in metabolic state specifically associated with disease severity. This was further evident on pairwise comparisons, with mild hospitalised COVID-19 patients differing from healthy volunteers in metabolic processes and vesicle transport of retinoid, cholesterol, lipoproteins, and fat-soluble vitamins; and from community cases by higher levels of complement activation and coagulation (Figure S6D). Severe COVID-19 patients differed from mild and from critically ill patients in processes relating to platelet degranulation and neutrophil degranulation respectively (Figure S6E-F). When we compared severe and critical COVID-19 with sepsis, 19 out of 105 proteins showed changes specific to COVID-19 (FDR<0.05, FC>1.5), enriched in acute-phase response, complement activation, and receptor-mediated endocytosis (Figure 6E).

### Plasma cytokine and chemokine profiling shows evidence for involvement of key inflammatory mediators

To further characterise inflammatory mediators of the response to SARS-CoV-2 and the biology of potential therapeutic targets, we analysed 51 circulating cytokine and chemokine proteins using the Luminex assay for 171 individuals (Figure 1A; Table S2; STAR Methods). There was clear clustering of hospitalised COVID-19 cases by severity on PCA of plasma protein abundance by severity while community cases overlapped with heathy controls and sepsis cases clustered separately (Figure 6F). The major proteins contributing to these axes of variance between groups were CXCL10, CXCL5, EGF, CCL2, S100A9, IL6, LCN2, CCL20, LF and G-CSF (Figure 6F). Overall, we found 49% (25 of 51) analytes were significantly differentially abundant in plasma from COVID-19 cases vs healthy volunteers (Figures 6G-I and S6G). Amongst these, CCL2, CCL19, CCL20, CXCL10, GM-CSF, IL-6, IL-8, IL-15, S100A9 and SCGF (all increased abundance) were strongly correlated with severity in hospitalised COVID-19 patients (r^2^>0.5, *P* <0.001).

We further compared with sepsis and influenza to investigate disease specificity and found that the plasma levels of G-CSF, IL-8, LF, CD163, LCN2, CCL20, IL-6, IL-10, CCL4, CCL19, TNF and C5a were lower in critical and severe COVID-19 than sepsis (Figure 6J). Compared with influenza, serum EGF, LF and CD40L were higher in serum from patients with critical COVID-19 while G-CSF was lower (Figure 6K). We then investigated protein-protein correlation network relationships of assayed plasma cytokines and chemokines. This identified S100A9, M-CSF and CCL2/19 as nodal proteins. When we performed protein-clinical trait correlation network analysis for COVID-19 severity we found strong correlations between clinical features (CRP, SaO2/FiO2 and ventilation days) and specific nodal proteins (GM-CSF, CXCL10, TREM-1, CCL2/19, TF, IL-6/15, MPO and S100A9) at the centre of the network (Figure 6L).

### Plasma proteome variation identifies patient sub-phenotypes of differing disease severity

We next investigated the utility of plasma proteins for patient sub-phenotyping within hospitalised COVID-19 cases (n=122 samples) by integrating the LC-MS-MS and Luminex datasets using Similarity Network Fusion (SNF) (Wang et al., 2014) (STAR Methods). We first constructed a sample-by-sample similarity matrix from which we derived a network for each of the two data types. Analysing these individually in an unsupervised manner with spectral clustering, we could only discriminate a minority of cases (the most mild from all others). However, when we fused these networks into a single similarity network that maximised shared and complementary information, we discovered two clusters that separated mild and critical cases, and discriminated within the severe cases to assign similar numbers to each of the two clusters (Figure 6M,N). We identified 11 proteins as the main discriminatory features distinguishing the clusters (mutual information score >= 0.15) (Figure 6M). When we compared the two clusters, we found that clinical measures of disease severity were significantly different including inspired oxygen concentration and SOFA oxygen score (t-test *P_c_* <0.05) (Figure S6H) and that membership of cluster two was associated with higher 28-day mortality (Figure 6O). The predictive protein set spanned key inflammatory mediators, including the cytokines and chemokines IL-6, IL-8 (CXCL8), CCL2, CCL19, CCL20 and CXCL10 together with S100A9 (calprotectin), the acute phase proteins serum amyloid protein (SAA1) and protease inhibitor (SERPINA3), GM-CSF and the C-type lectin CLEC11A.

We validated the clusters in an independent acute hospitalised COVID-19 cohort from Boston assayed using a different technology, targeted proteomics by Olink (Filbin et al., 2020) (STAR Methods). We found that clustering analysis, using the 7 out of 11 predictive proteins for which data was available, identified two optimal clusters (Figures 6P and S6I). These showed a clear relationship with measures of disease severity, including WHO ordinal score (maximum) (Figure S6J), and patients in cluster 1 had significantly lower mortality at 28 days (5/164=3.0%) compared with cluster 2 (33/105=31.4%) (Chi-squared test *P* <0.0001), validating the findings from our discovery cohort. We then explored the specificity of the clusters by extending the approach to include a combination of hospitalised COVID-19 and sepsis patients from COMBAT. This revealed three clusters, two corresponding to the clusters seen with COVID-19 cases analysed alone, indicating a high level of specificity (Figure S6K,L). Features that separated COVID-19 and sepsis included lipocalin2 (LCN2) and CCL20, which were elevated in sepsis, and CXCL10, APCS and fibronectin (FN1) which were higher in COVID-19.

### Supervised machine learning identifies predictive protein biomarkers for disease severity

We complemented our findings by using machine learning to combine the two proteomics data types with whole blood total RNAseq to determine which features were predictive of disease severity (WHO category at the time of sampling) and their relative informativeness (Figure S7A; STAR Methods). We first identified assay-type-specific feature scoring PCs to reduce dimensionality for a training sample set, and then determined the six maximally informative PCs (Figure 7A) and the genes or proteins maximally contributing to loadings for each PC. After feature elimination based on performance, we found the minimal set of cross-modality features to predict severity were the acute phase proteins SAA2 and CRP, an immunoglobulin (IGHG4), chemokines (CCL20 and CCL2), IL-6 and complement component C5a (Figures 7B and S7B); the combined performance of these features in the hold-out validation set showed a balanced accuracy of 75-80% to predict WHO category group (Figure S7C). We also used machine learning to search for features that distinguish hospitalised COVID-19 patients from sepsis. A multi-omic set of 81 features was discovered using SIMON (Tomic et al., 2021) (STAR Methods) (AUC 0.85 95% CI 0.59-1), identifying specific differentially abundant genes, proteins (including FCN1 and APCS as higher in COVID-19) and significant pathway enrichment for hematopoietic cell lineage and the renin-angiotensin system (Figure S7D,E).

**Figure 7.**
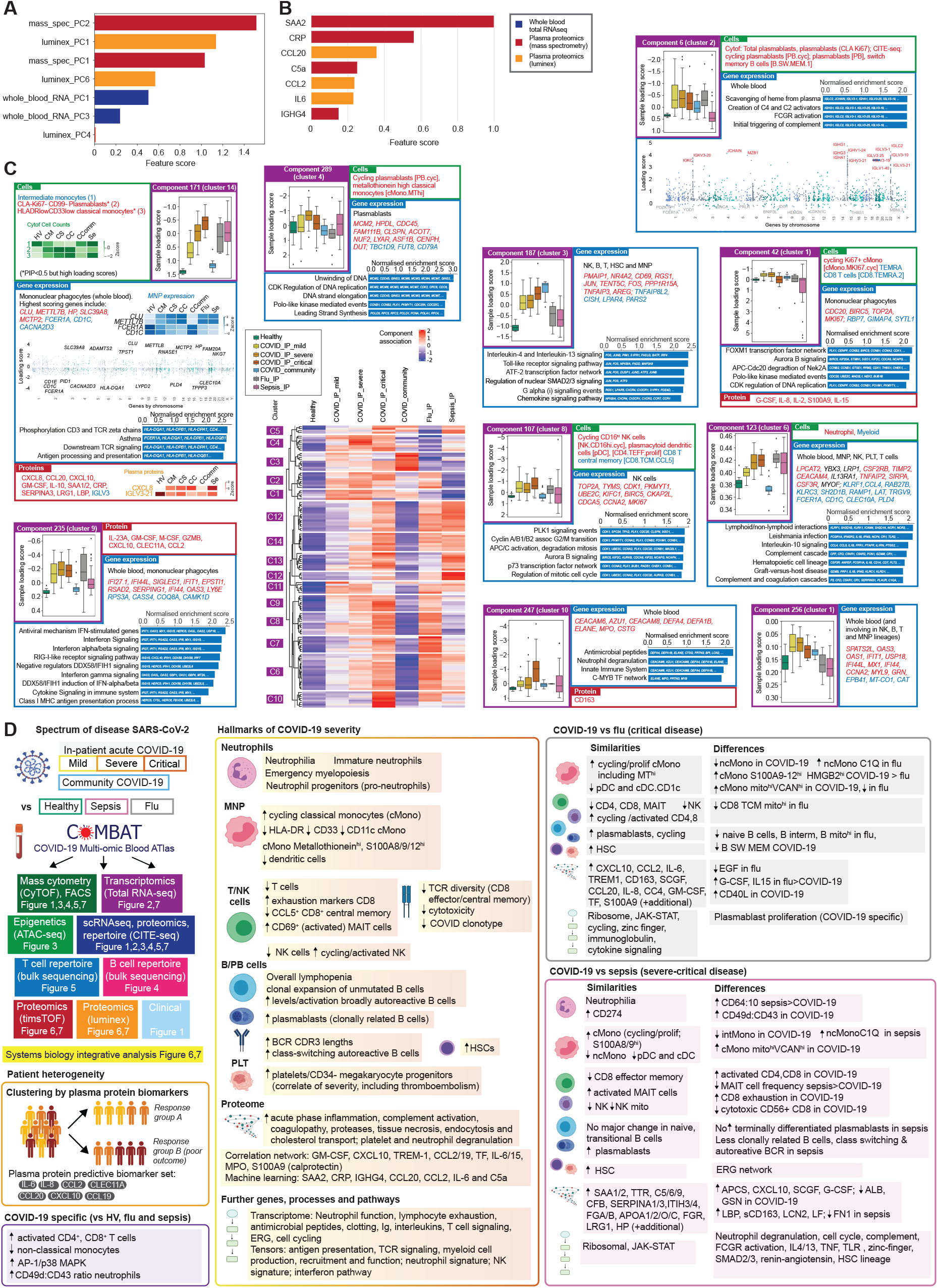
Integrative approaches define hallmarks of COVID-19 response. (A,B) Machine learning for COVID-19 severity showing average feature score of (A) highest-scoring features (PCs) (B) final feature set. (C) Tensor and matrix decomposition across multi-omic datasets (CITEseq, whole blood RNAseq, mass cytometry, luminex and mass spectrometry) for 152 samples showing clustering of COVID-19 associated components (k-means clustering of row-scaled median sample loadings) and relationship with disease comparator groups, with examples of components showing component number and cluster membership; and sample loading scores across comparator groups and features (cells, gene expression, proteins) whose variance contributes to that component. For gene expression, cell type and highest scoring genes listed (red upregulated, blue downregulated) together with top pathway enrichment (FDR <0.05) with pathway genes listed within bars (features shown or included in pathway analysis where posterior inclusion probability >0.5). More detail provided for component 171 as exemplar. (D) Overview figure summarising key findings from this study including hallmarks of COVID-19 severity, shared and specific features with influenza and sepsis, and patient stratification. See Figure S7.

### Integrated hallmarks of COVID-19 severity and specificity

We next sought to understand the immune response to COVID-19 across all assay types and samples using a multi-omics tensor approach (Chang et al., 2021; Fanaee-T and Thoresen, 2018; Taguchi, 2017), specifically the sparse decomposition of arrays (SDA) algorithm (Hore et al., 2016). We analysed 152 samples assayed for cellular composition, gene expression and plasma proteomics, and identified 381 latent SDA components, each comprising vectors of scores (loadings) that indicate the contribution of individual cell types, genes or proteins linked by that component, and thereby offer insights into shared mechanism (Figure S7F; Table S3; STAR Methods). We then identified components associated with specific clinical covariates, severity or patient group, noting that while in some instances such as gender there was a single associated component (involving differential sex chromosome gene expression across cell types, component 2), typically several components were associated (Figure S7G,H). The strongest association with COVID-19 severity was component 171 (*P_c_* 5.9×10^-14^, rho 0.74 Spearman) which was unusual in having a high feature contribution from plasma proteins whereas gene expression contributed most to the majority of the other components (Figure S7I). Component 171 involved myeloid cell production, recruitment and function. Features contributing to loading scores with high posterior inclusion probability included raised plasma chemokines involved in chemotaxis and activation (CXCL8, CXCL10 and CCL20) and GM-CSF together with acute phase activating proteins (SAA1/2 and SERPINA3), LRG1 and LBP (Figure 7C); reduced abundance of intermediate monocytes; high expression of cell stress chaperone *CLU* and methyltransferase *METTL7B*, and downregulation of IgE receptor and multiple HLA class II genes, and pathway enrichment for antigen presentation, TCR signaling and asthma.

To further delineate COVID-19 associated SDA components, we performed pairwise contrasts and analysis of variance involving COVID-19 patient groups. Overall 130 of 381 components were significantly associated with COVID-19 versus healthy volunteers (Figure 7C and S7J). To identify which of these components were informative for severity and how they may be shared or specific for COVID-19, we clustered their median loadings across the different disease groups (Figure 7C). Components associated with mild and severe but not critical disease included component 42 (features of monocyte/granulocyte proliferation and function, elevated plasma proteins G-CSF, IL-2, IL-8 and IL-15, and enrichment of cell division related pathways); and component 256 (including upregulation of interferon response genes and down regulation of genes such as catalase and cytochrome c oxidase) which was specific to COVID-19 cases (Figure 7C). Other components strongly associated with severe disease involved plasmablast proliferation, combined with increased MT^hi^ cMono and a clear DNA replication signature (component 289), or with widespread upregulation of immunoglobulin heavy/kappa/lambda genes, *JCHAIN* (regulating multimerization and mucosal secretion of IgM/IgA), and *MZB1* (involved in antibody secretion and integrin-mediated cell adhesion) (component 6), linking with possible antibody-dependent cellular toxicity (Figure 7C).

We found an innate response component specific to critical COVID-19 (component 247) with differential expression of granulocyte activation marker (*CEACAM8),* neutrophil elastase (*ELANE*) and defensins (*DEFA1B/4*) and increased soluble CD163 scavanger protein levels, reflected in pathway enrichment for neutrophil functions. Neutrophil related features were also found in component 123 associated with COVID-19 severity, influenza and sepsis (Figure 7C). Other SDA components had high loading scores associated with all hospitalised COVID-19 patients including significant upregulation of interferon pathway genes (component 235) and an NK-signature component with upregulation of cell cycle and proliferation genes (component 107); as well as with hospitalised and community COVID-19 patients (component 187) (Figure 7C). The latter was driven by differential expression in major PBMC lineages (highest loadings in NK, B and T cells) involving upregulation of key stress and activation response genes including immediate early response protein (*PMAIP1*), AP-1 transcription factor genes *FOS* and *JUN*, the early activation marker and metabolic reprogramming gene *CD69,* and *TNFAIP3*, which limits NFkB mediated inflammation. The cytokine-induced STAT inhibitor (*CISH*) and immune checkpoint regulator of inflammation and metabolism *TNFAIP8L2* were downregulated. Pathway enrichment was seen for type-2 inflammation (IL4/13), TLR signaling and the ATF-2 network (Figure 7C).

Overall, the latent component analysis identifies hallmarks of COVID-19 severity, and specificity with respect to sepsis and influenza. Our findings highlight key cellular populations such as proliferating monocytes and plasmablasts, and features of innate and adaptive mechanisms ranging from interferon signalling to myelopoiesis, immunoglobulin production and stress activation response signaling. The results prioritise and validate hallmarks seen on individual modality analysis such as AP-1, and generate hypotheses for how hallmarks may be related in terms of pathophysiology based on co-occurrence in a given component.

## Discussion

Our comprehensive multi-modal integrated approach, applied to multiple well-defined cohorts of patients and healthy volunteers, has defined blood hallmarks of COVID-19 severity and specificity involving particular immune cell populations and their development, components of innate and adaptive immunity, and connectivity with the inflammatory response (Figure 7D).

Hallmarks of severity involving myeloid related features include emergency myelopoiesis, immature neutrophils, increased HSC and platelet/CD34^-^ megakaryocyte progenitors, with the latter associated with thromboembolism. These findings substantiate and add granularity to previous reports (Bernardes et al., 2020; Stephenson et al., 2021). We find evidence that *ERG* is central to a gene network linked to these cell populations, encoding a transcription factor important in determining lineage plasticity, modulating inflammation and maintaining an anti-thrombotic environment (Yuan et al., 2009). We further identify hallmarks supporting the importance of mononuclear phagocyte dysfunction in severe disease (Bost et al., 2021; Mann et al., 2020; Schulte-Schrepping et al., 2020), namely proliferating cMono, and specific monocyte populations showing reduced HLA-DR, CD33 and CD11c expression, high expression of antioxidant metallothionein and S100A8/9/12 (calprotectin), together with reduced pDCs.

The frequency of specific T cell subsets, and their activation and exhaustion, has been previously implicated in severe COVID-19 (Chen and Wherry, 2020; Jouan et al., 2020; Parrot et al., 2020). We find evidence for increased numbers of activated CD8^+^ T cells and NK cell populations in COVID-19, and, with increasing severity, failure of clonal expansion in CD8^+^ T effector and central memory cells and depletion of COVID-19 clonotypes. We further find association with severity for exhaustion markers and specific activated NK and CD69^+^ MAIT cell populations. In terms of adaptive immunity (Brouwer et al., 2020; Galson et al., 2020), we find increased numbers of terminally differentiated plasmablasts, with expansion of unmutated B cells differing in selection and tolerance and a higher proportion of clonally related B cells.

Redox state and cell cycle associate with more severe disease across cell populations. Our data are also consistent with the importance of the hyperinflammatory state (Moore and June, 2020) and interferon response (Hadjadj et al., 2020; Lei et al., 2020) but as features of less critical disease and earlier phase of illness.

Our proteomic analysis has identified specific plasma cytokine and chemokine levels as biomarkers of severe disease with evidence for acute phase inflammation, complement activation/attack, fibrin clots, proteases, serum amyloid, tissue necrosis, receptor mediated endocytosis and cholesterol transport as hallmarks. Moreover, we have discovered plasma protein signatures that can be used to stratify acute hospitalised COVID-19 cases into disease sub-phenotypes, with cluster membership informative for response state and associated with differential 28-day mortality. We have validated our finding in an independent dataset using a predictive set of seven plasma proteins (cytokines IL-6, IL-8; chemokines CCL2, CCL19, CCL20, CXCL10; and C-type lectin CLEC11A, a key growth factor for primitive haematopoietic progenitor cells). Patient stratification is important given the observed clinical heterogeneity within severe COVID-19. Such variability has historically been a major confounder of clinical trials for targeted immune therapy in other severe infections (Davies et al., 2018; Marshall, 2014).

This work has demonstrated the informativeness of a multi-linear tensor approach to stratify and interpret the complexity of multi-omic datasets. Application of the SDA algorithm was able to identify latent components of variance that joined together signals from across cellular, gene expression and plasma protein measurements. For example, the most significant tensor component associated with COVID-19 severity involves myeloid chemotaxis and activation, the acute phase response, HLA class II downregulation and TCR signaling. Thus, our dataset provides a useful resource from which to develop other approaches for identifying multi-modal signals and associated mechanistic insights, such as those leveraging algebraic systems biology (Gross et al., 2016), multi-layer networks (Kivela et al., 2014), topological data analysis (Camara, 2017) or tensor clustering (Seigal et al., 2019).

Considering features specific to COVID-19, we have found that the ratio of integrin alpha 4 (CD49d) to leukosialin (CD43) is specifically elevated across COVID-19 patients, including into recovery and in convalescence, suggesting that this ratio may be an informative index score for neutrophil activities specific to COVID-19. Reduction in CD43 expression leads to neutrophil retention in the bloodstream and increased adherence to vessel walls (Woodman et al., 1998), and may be linked to the enhanced thrombosis observed in COVID-19 patients.

Our epigenetic, gene expression and integrative SDA analyses have all identified the AP-1 p38 MAPK pathway upregulation as a specific feature of COVID-19 disease across different immune subsets. Combined with evidence of proliferation and cytokine response in these populations, this supports systemic immune activation and proliferation as a hallmark specific to COVID-19. Moreover, AP-1 is a pioneer transcription factor that reversibly imprints the senescence enhancer landscape following stress and can be modulated to reverse T cell exhaustion (Lynn et al., 2019; Martinez-Zamudio et al., 2020). This suggests a possible role for AP-1 in inappropriate chromatin remodelling and cellular activation/senescence in multiple cell types, with evidence of persisting differential chromatin accessibility, gene expression signatures and cell protein markers of activation that may contribute to both acute disease and post-COVID-19 syndrome. Our findings are in keeping with recent evidence of efficacy for baricitinib (Janus Kinase inhibitor) in improving recovery of hospitalised patients (Kalil et al., 2021). Baricitinib acts upstream of AP-1 (Zarrin et al., 2021) and controls macrophage inflammation and neutrophil recruitment in COVID-19 (Hoang et al., 2021).

Our findings involving immune activation and proliferation are of further relevance given the key nodal plasma cytokines, including GM-CSF and IL-6, we identify as hallmarks of disease severity in proteomic and integrative analysis, targeting of which could also benefit dysfunctional granulopoiesis and neutrophil subsets. This supports the efficacy of tocilizumab (IL-6 receptor inhibitor) in severe disease (Horby et al., 2021b), moreover we find upregulation of *IL6R* in more severe disease. Our data supports the use of inhibitors of GM-CSF in current clinical trials, use of anti-CCL2 (Gracia-Hernandez et al., 2020), repurposing of immune checkpoint inhibitors (Pezeshki and Rezaei, 2021) to restore/enhance CD8^+^ T cell cytotoxicity, and potential targets such as FGFRs, which are cofactors for early stage viral infection upregulated by MERS-CoV2 leading to lung damage and proposed as a therapeutic target (Hondermarck et al., 2020).

Establishing optimal biomarkers of response and relevant sample collection, as well as timely availability of results, are important considerations in current immunomodulatory trial design in COVID-19. We find the widely used clinical biomarker CRP has predictive utility as a component of discriminating biomarker marker sets for severity. Total RNAseq of whole blood stabilised at the bedside, a tractable sample type for collection in a pandemic situation, is highly informative for understanding variance in disease severity while use of small volumes of rapidly fixed whole blood for later FACS and mass cytometry gives complementary and highly granular resolution of the cellular immune response state and identifies specific cell populations important in severe disease.

An important question is how the hallmarks of severe disease in COVID-19 relate to non-SARS-CoV-2 sepsis (Beltran-Garcia et al., 2020; Olwal et al., 2021). We have compared patients with severe or critical illness, not only revealing shared features relating to emergency myelopoiesis and progenitors but also identifying discriminating neutrophil markers (CD49d:CD43 ratio) in COVID-19 and specific ncMono and cMono phenotypes in sepsis (Figure 7D). Lymphocyte exhaustion is proposed as a common mechanism in COVID-19 and sepsis (Boomer et al., 2012; Diao et al., 2020). We find overall higher levels of CD4^+^ and CD8^+^ T cell activation, and more marked changes in cell markers, in COVID-19, suggesting a greater degree of CD8^+^ T cell exhaustion. While we find no major changes in naïve and transitional B cells in either condition, in sepsis we observed an absence of terminally differentiated plasmablasts and fewer clonally related B cells, class switching and autoreactive BCRs. We have further identified plasma proteins discriminating COVID-19 and sepsis, for example lipopolysaccharide binding protein, lactoferrin and lipocalin 2; and differential pathways including neutrophil degranulation, complement, AP-1/p38MAPK, TLR, renin-angiotensin and the HSC lineage. We note that COVID-19 induced down regulation of *ACE2* may be involved in the modulation of both the renin-angiotensin system and activation of AP-1/p38MAPK signaling (Grimes and Grimes, 2020).

Similarly, while differences in target cells and control of viral replication are recognised between COVID-19 and influenza, shared immune responses and mechanisms are also reported (Flerlage et al., 2021). Our data builds on previous blood immune signatures (Lee et al., 2020; Mudd et al., 2020; Zhu et al., 2020) to gain cellular and proteomic insights into the more severe form of these infections. We find similar changes in frequency of major immune cell populations between COVID-19 and influenza for HSC, the majority of T and NK cell populations, classical/cycling monocytes and plasmablasts; but differences for many B cell populations (differential naïve, intermediate and memory B cell response) as well as specific cell subsets notably ncMono subpopulations (Figure 7D). While the bulk of the circulating proteomic response is shared, specific differences involved EGF, G-CSF and IL-15. We further found extensive sharing of pathways and networks including modules involving JAK-STAT and zinc finger proteins, as well as differences, most notably the enrichment of AP-1/MAPK specifically in COVID-19.

In conclusion, our multi-omic integrated blood atlas comprehensively delineates the host immune response in COVID-19 from the start of the UK pandemic, prior to clinical trial-led implementation of approved treatments or vaccination. This provides the community a unique reference resource for replication and meta-analysis, to interpret datasets generated from interventional trials and includes tools for direct visualisation (https://mlv.combat.ox.ac.uk/).

Integrative approaches such as we have applied here are essential to better differentiate COVID-19 patients according to disease severity, underlying pathophysiology and infectious aetiology. This will be important as we seek novel therapeutic targets and the opportunity for a precision medicine approach to treatment that is appropriately timed and targeted to those patients most likely to benefit from a particular intervention.

## Data Availability

Data from our study can be explored interactively through a web portal: https://mlv.combat.ox.ac.uk/

## Consortia

Members (Table S1).

## Acknowledgements

Funding support and declaration of interests (Table S1), author contributions (Table S1, Methods S3).

## Supplementary figure legends

**Figure S1.**
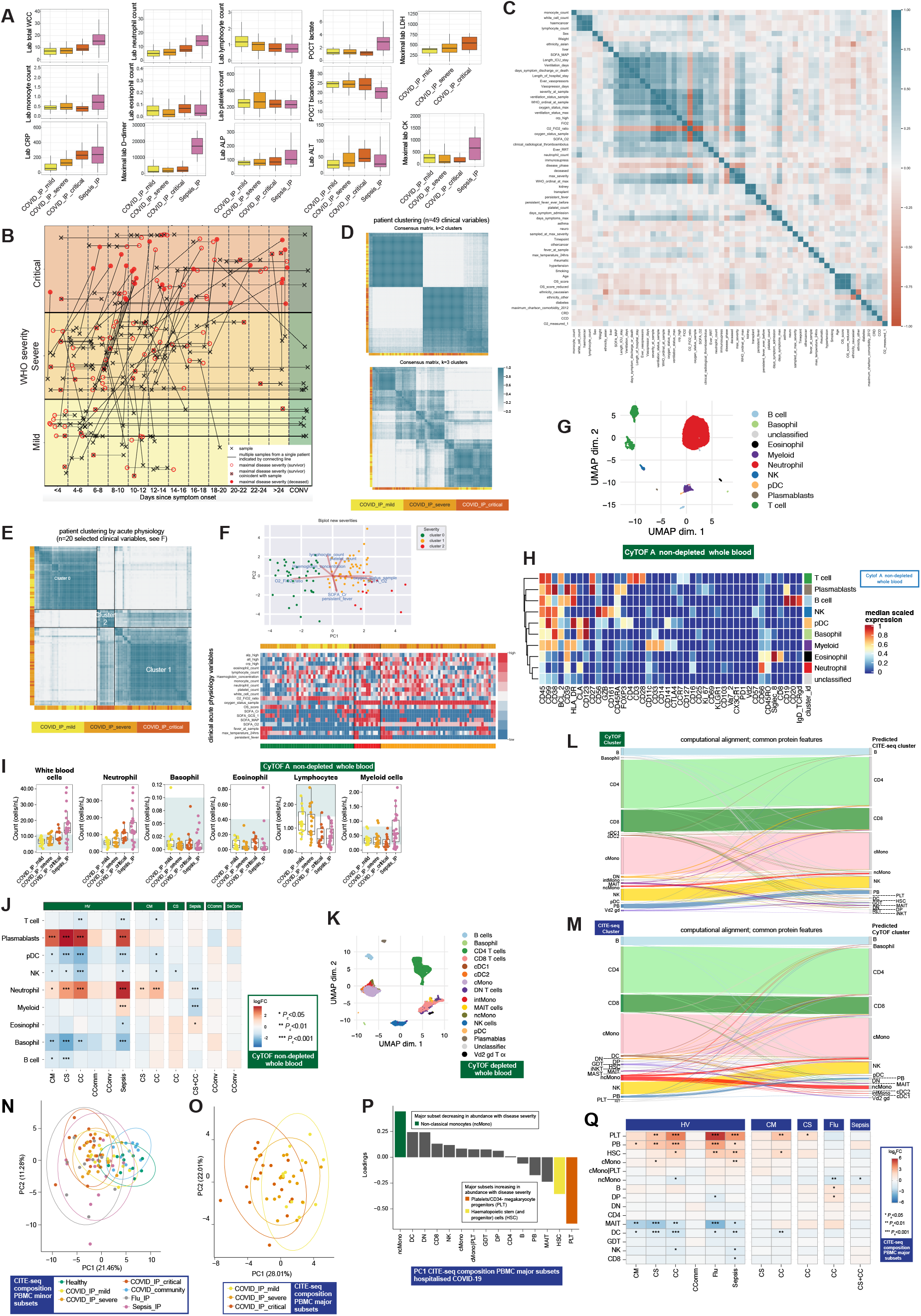
Study cohorts, clinical covariates and single cell compositional approaches, related to Figure 1. (A-F) Clinical covariates and features for studied cohorts. (A) Admission samples for hospitalised COVID-19 (n=116) and sepsis (n=58) including total and differential cell count and clinically assayed biomarkers CRP, D-dimer, LDH, creatine kinase, ALT, ALP. (B) Overview of hospitalised COVID-19 sampling by time from symptom onset and WHO severity with maximal severity indicated. (C) Correlation matrix of clinical covariates and markers of response in hospitalised COVID-19 cases. (D,E) Unsupervised clustering of samples from hospitalised COVID-19 patients with consensus k-means clustering followed by hierarchical clustering on the consensus matrix based on (D) 49 clinical features (excluding WHO severity classifiers) (E) acute measures of physiology and clinical biomarkers of response without significant missingness (including measures of oxygenation requirements, blood cell counts, fever, ALT, CRP). (F) Biplot illustrating for PC1 and PC2 features driving clustering, and heat map of clinical features, coloured by three clusters identified in k-means clustering using acute measures of physiology and clinical biomarkers of response. (G-M) Stabilized whole blood (Cytodelics) from COVID-19 patients analysed by single cell resolution mass cytometry (Helios CyTOF system) (including matched samples collected during convalescence from 16 COVID-19 hospitalized patients). (G-J) Non-granulocyte depleted samples. A self-organising map algorithm (FlowSOM) resolved 25 clusters by consensus clustering for 3,893,390 cells after down sampling to a maximum of 40,000 cells. Clusters merged to identify broad immune cell populations (G) UMAP (H) clustering of major cell populations (y axis) by discriminating marker (x-axis) (I) cell counts (J) differential abundance analysis in patients compared to healthy volunteers, and different disease states clustering major cell populations. (K) Granulocyte (CD66^+^) depleted whole blood with down sampling to a maximum of 75,000 cells and 7,118,158 cells assayed showing UMAP (L,M) Plots demonstrating cross validation mass cytometry and CITEseq cell clusters. (N-Q) CITE-seq analysis of viability sorted peripheral blood mononuclear cells (PBMCs) from 140 samples profiled using the 10X Genomics platform. (N) PCA minor cell subsets for all patient groups. (O,P) PCA major subsets for hospitalised COVID-19 patients (O) PC1 vs PC2 (P) loadings of cell clusters on PC1. (Q) Differential abundance analysis in patients compared to healthy volunteers, and between disease categories for major cell subsets.

**Figure S2.**
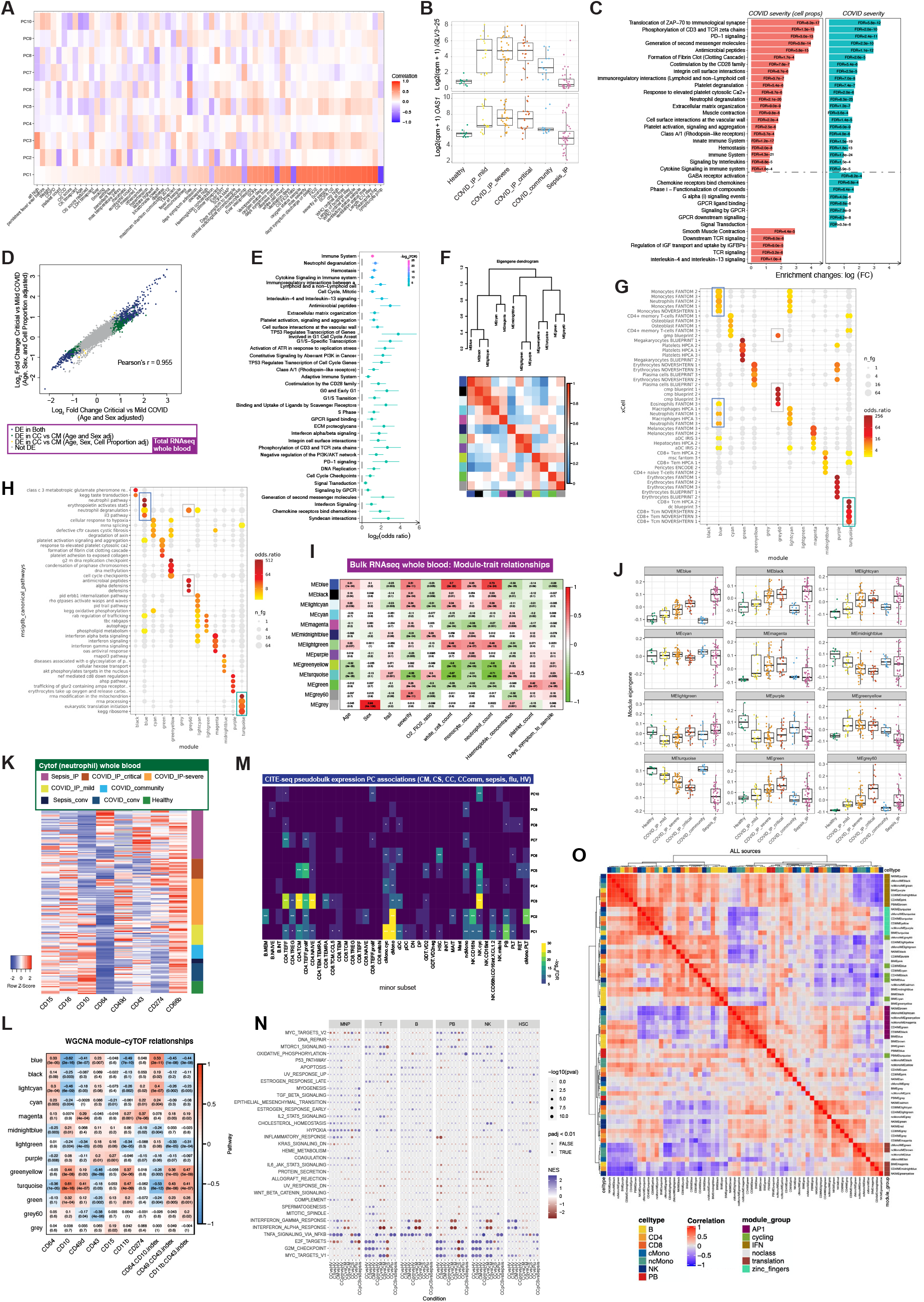
Signatures of COVID-19 severity revealed by bulk and single cell RNAseq, related to Figure 2. (A-E) Whole blood total RNAseq for hospitalised COVID-19 patients showing (A) matrix correlation of PCs with covariates (B) differentially expressed immunoglobulin lambda chain gene *IGLV3* and innate viral response gene *OAS1* (C) pathway enrichment on analysis of COVID-19 severity classes as a quantitative trait (mild=1, severe=2, critical=3) with and without inclusion of cell proportion (D) correlation plot showing the influence of cell proportion on detection of differentially expressed genes (E) pathway enrichment COVID-19 severe and critical vs sepsis using Reactome. (F-J) Weighted gene correlation network analysis (WGCNA) whole blood total RNAseq (F) dendrogram and heatmap of eigengene network arising from WGCNA with hierarchical clustering (G,H) enrichment of WGCNA modules using gene expression data for (G) 64 immune and stroma cell types (xCell) (H) msigdb canonical pathway genesets (I) heat map showing module trait relationships on WGCNA (blue, negative correlation; red, positive correlation) (J) module eigengene values plotted by patient group. (K) Neutrophil marker expression whole blood assayed by mass cytometry comparing across patient groups. (L) Correlations between whole blood total RNAseq WGCNA modules and neutrophil CyTOF markers. (M) Association p-values between principal components of pseudobulk GEX for specific cell clusters across all patients. (N) scRNAseq MSigDB hallmark gene set enrichment for cell types (Liberzon et al., 2015) (O) WGCNA analysis CITEseq gene expression for all samples showing correlation of eigengene expression (all modules in all cell types across all patient groups) with major module groups annotated.

**Figure S3.**
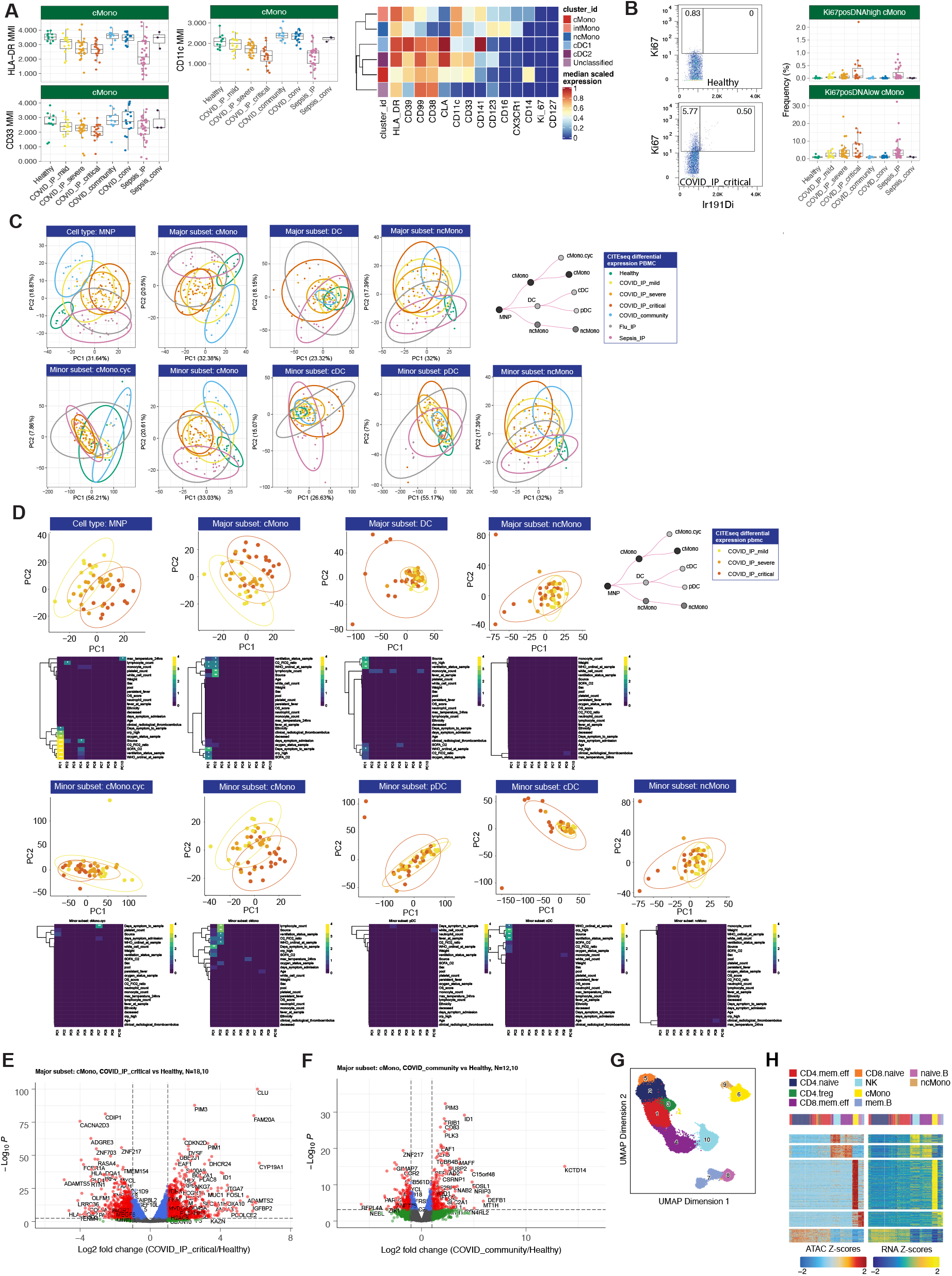
Changes in myeloid populations associated with COVID-19 severity, related to Figure 3. (A,B) Single cell mass cytometry (A) mean Metal Intensity (MMI) of HLA-DR, CD33 and CD11c in classical monocytes (cMono) (B) representative plot of Ki67^+^ expression and 191Iridium (DNA) labelling in a healthy volunteer and a COVID19 patient; two distinct population of Ki67^+^ proliferating cells were identified, one containing the same amount of DNA as Ki67^-^ cells (Ki67^+^DNA^low^) and a rarer population containing double the amount of DNA (Ki67^+^DNA^high^) which likely comprises proliferating cells in S, G2 and M phase. The boxplots describe the frequencies of Ki67^+^DNA^low^ and Ki67^+^DNA^high^ across different disease states. (C) PCA of CITE-seq gene expression in mononuclear phagocyte (MNP), major and minor cell subsets comparing all groups (COVID-19 IP mild, severe, critical; COVID-19 community; influenza; sepsis; healthy) (D) PCA and correlation with clinical covariates and severity measures for gene expression in MNP, major and minor cell subsets for acute hospitalised cases only (mild, severe, critical). (E,F) Differential gene expression in classical monocytes comparing (E) critical COVID-19 patients vs healthy volunteers and (F) COVID-19 community cases vs healthy volunteers with volcano plots showing significant genes (FDR < 0.01 and logFC > 2) in red. (G,H) scATACseq with cell lysis, nuclear extraction and tagmentation on viability sorted PBMC prior to single nuclei capture and sequencing. Data shown for 42,000 cells post QC (ArchR pipeline) for 8 COVID-19 samples (paired acute and convalescent) and 2 healthy volunteers with (G) label transfer (unconstrained method) to assign cell clusters based on CITEseq and (H) comparison of chromatin accessibility (scATACseq peaks linked to genes) to CITEseq gene expression.

**Figure S4.**
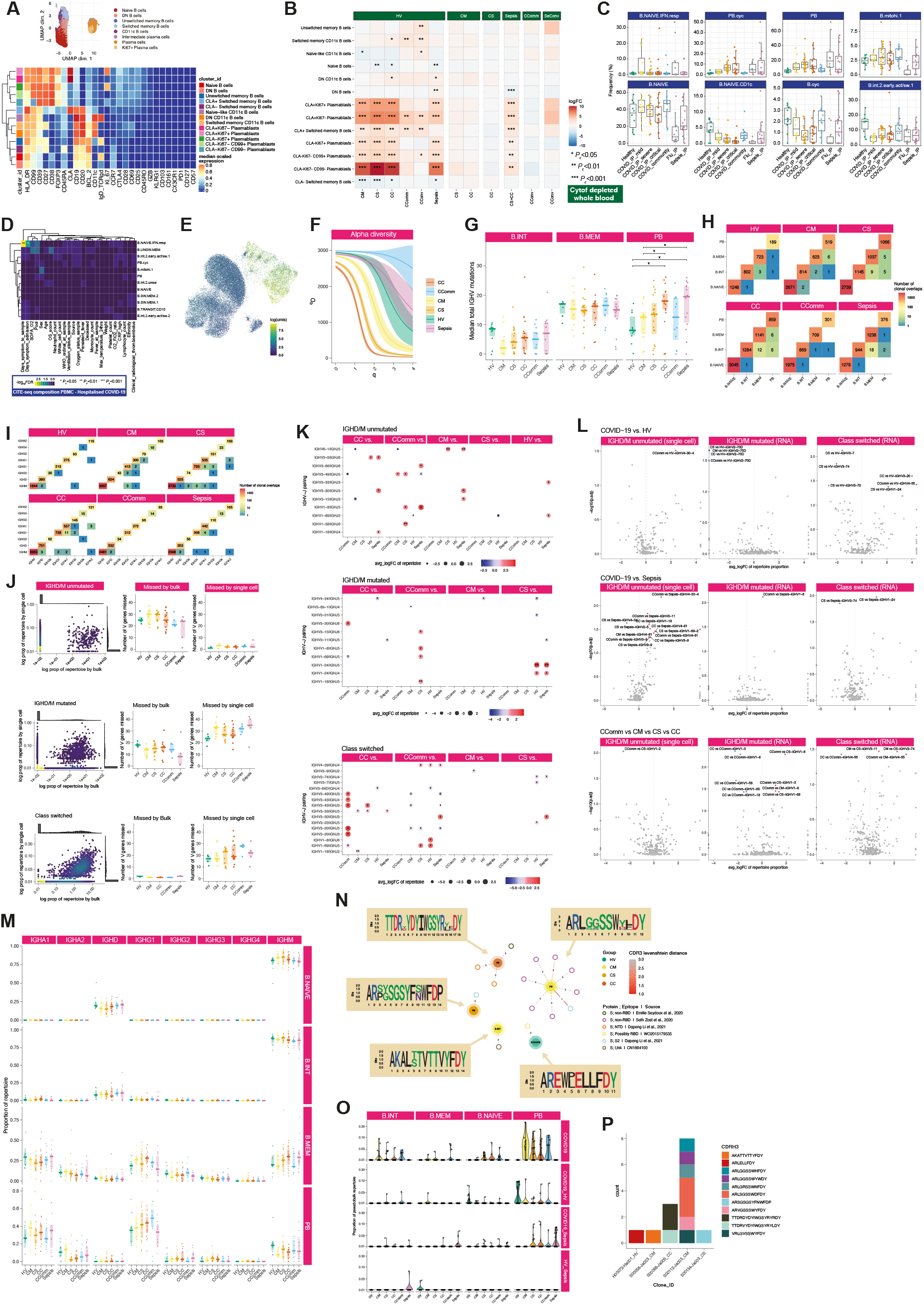
B lymphocytes show changes in composition and repertoire associated with COVID-19 severity, related to Figure 4. (A,B) Single cell mass cytometry (CyTOF) compositional analysis showing (A) UMAP and clustering of major cell populations (B) differential abundance analysis in patients compared to healthy volunteers, and between disease categories. (C,D) CITE-seq compositional analysis (C) differential abundance analysis of B and plasmablast cell clusters (D) covariate analysis of PBMC B and plasmablast cell cluster abundance and clinical, demographic and experimental variables in hospitalised COVID-19 cases (with BH adjusted ANOVA test for significance). (E) UMAP embedding of all B cells from single cell VDJ dataset. Colour scale depicts log total UMIs per VDJ sequence. (F) Alpha diversity (derived from (Gupta et al., 2015)). Bootstrapped alpha diversity curves are shown per study group across different orders of q. (G) IGHV total mutations across B cell subsets per study group (naïve B cells not shown, as no mutations). (H) Clonal overlaps across B cell clusters per study group. Numbers reflect binary detection events. (I) Clonal overlaps across constant region genes per study group. Numbers reflect binary detection events. (J) Correlation analysis of IGHV genes detected by single cell VDJ data and RNA VDJ data, within the (top) IGHD/M unmutated, (middle) IGHD/M mutated and (bottom) class-switched VDJ sequences. Scatter plots, with marginal histograms, depict log proportions of repertoires detected and missed by both datasets. Boxplots to the right depict number of IGHV genes missed by RNA (bulk) and single cell VDJ data. (K) Heavy chain V and J gene pairing across pairwise study group comparisons, only significant pairings shown. Dot sizes and colour depict average log2 fold change of proportion of repertoire. (L) Volcano plots of log2 fold change of IGHV gene usage of IGHD/M unmutated sequences (derived from single cell VDJ data), IGHD/M mutated sequences (derived from RNA VDJ data), class-switched sequences (derived from RNA VDJ data). Top panel depicts significant genes across pairwise comparisons of COVID19 groups vs. healthy volunteers. Middle panel depicts significant genes across COVID19 vs sepsis groups. Bottom panel depicts significant genes across COVID19 severity groups. All significant pairwise comparisons are derived from Dunn tests post Kruskal Wallis testing. (M) Proportion of constant region genes across B cell clusters per study group. (N) Sequence similarity network of VDJ sequences, from single cell VDJ data (central nodes), to published monoclonal antibodies (peripheral nodes; references and epitopes described in legend). Edges depict pairwise Levenshtein’s distance of CDR3s. CDR3 sequence logos are shown following multiple sequence alignment. (O) The proportion of B cells across each B cell cluster per disease group of sequences shared between patient groups (observed in at least 2 patients). (P) Cumulative bar chart of the frequencies of known SARS-CoV-2 binding BCRs per patient group. Each section represents an individual patient.

**Figure S5.**
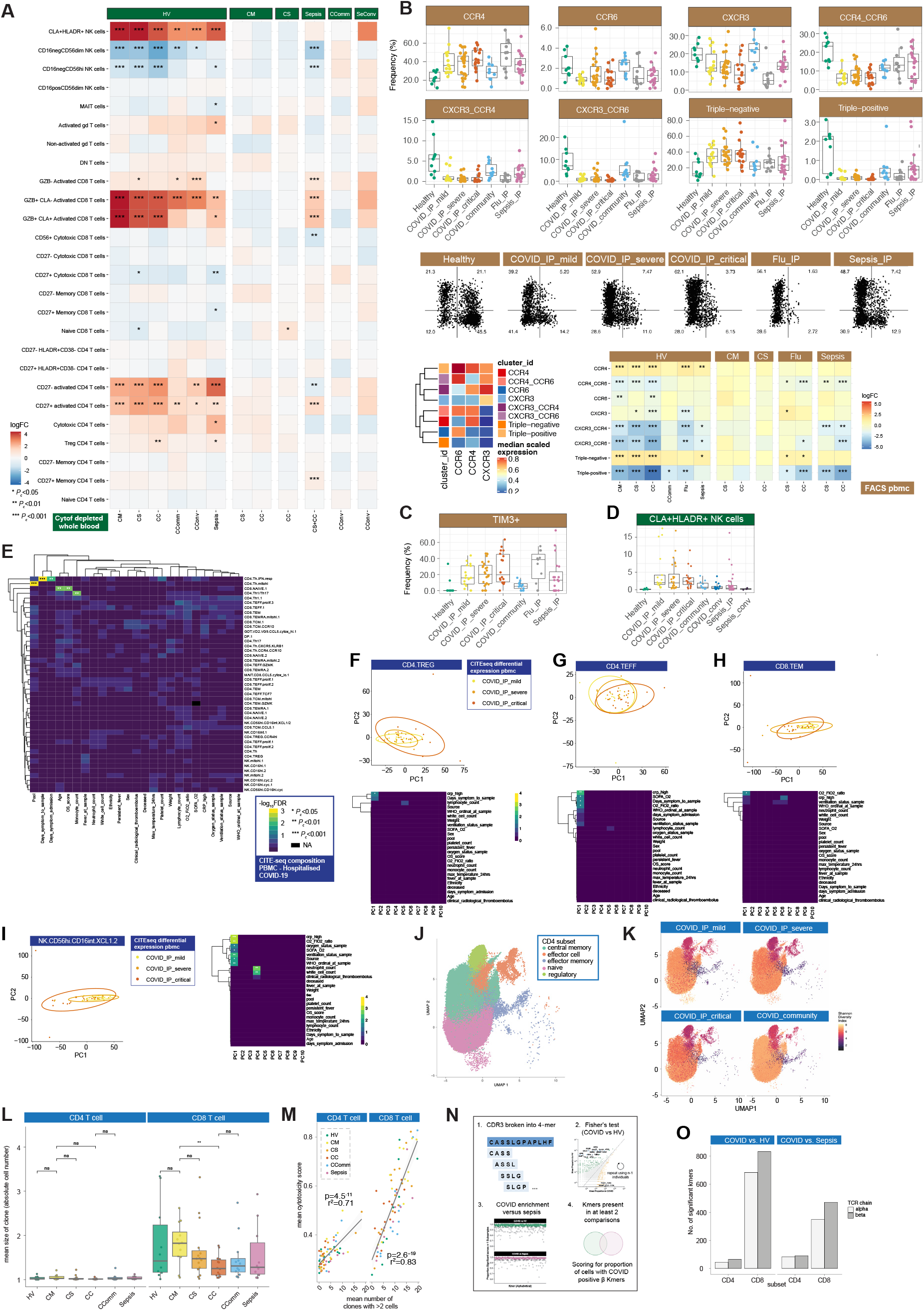
Dynamic changes in T lymphocyte and NK composition and repertoire associated with COVID-19 severity, related to. Figure 5. (A) Single cell mass cytometry (CyTOF) of whole blood showing differential abundance for T and NK cells. (B,C) Multicolour Flow Cytometry analysis of PBMC (B) boxplots, dotplots and heatmap describing the phenotype and frequency of subsets of memory CD4 T cell subsets defined based on the expression of CCR4, CCR6 and CXCR3 (C) frequency of TIM3^+^CD38^+^HLADR^+^ CD8^+^ T cells. (D) Single cell mass cytometry frequency of CLA^+^HLADR^+^NK cells. (E-I) CITEseq profiling of T and NK cell populations in PBMC (E) covariate analysis of cell abundance with clinical, demographic and experimental variables in hospitalised COVID-19 cases (with BH adjusted ANOVA test for significance) (F-I) PCA and correlation with clinical covariates and severity measures for gene expression in acute hospitalised cases (mild, severe, critical) in (F) CD4 regulatory T cells (G) CD4 effector T cells (H) CD8 effector memory T cells (I) activated NK cells. (J,K) UMAP of CD4+ T cells and associated clusters (J) used in repertoire analysis (K) indicating Shannon diversity of CD8 T cells by patent group. (L) Mean clone size CD4 and CD8. (M) Using a pre-defined cytotoxicity metric the overall cytotoxicity was calculated per individual for both the CD4 and CD8 subsets. For each individual the number of enlarged clones in these subsets was determined (defined as >2 cells with the same TCR chain). Mean cytotoxicity per individual is correlated with the number of expanded clones across each individual, irrespective of cohort origin (Pearson’s R2). (N) Illustration of the method used to identify CD3 Kmers associated with COVID-19 compared to cells from healthy volunteers and patients with sepsis. (O) Number of kmers comparing COVID-19 vs healthy volunteers and sepsis.

**Figure S6.**
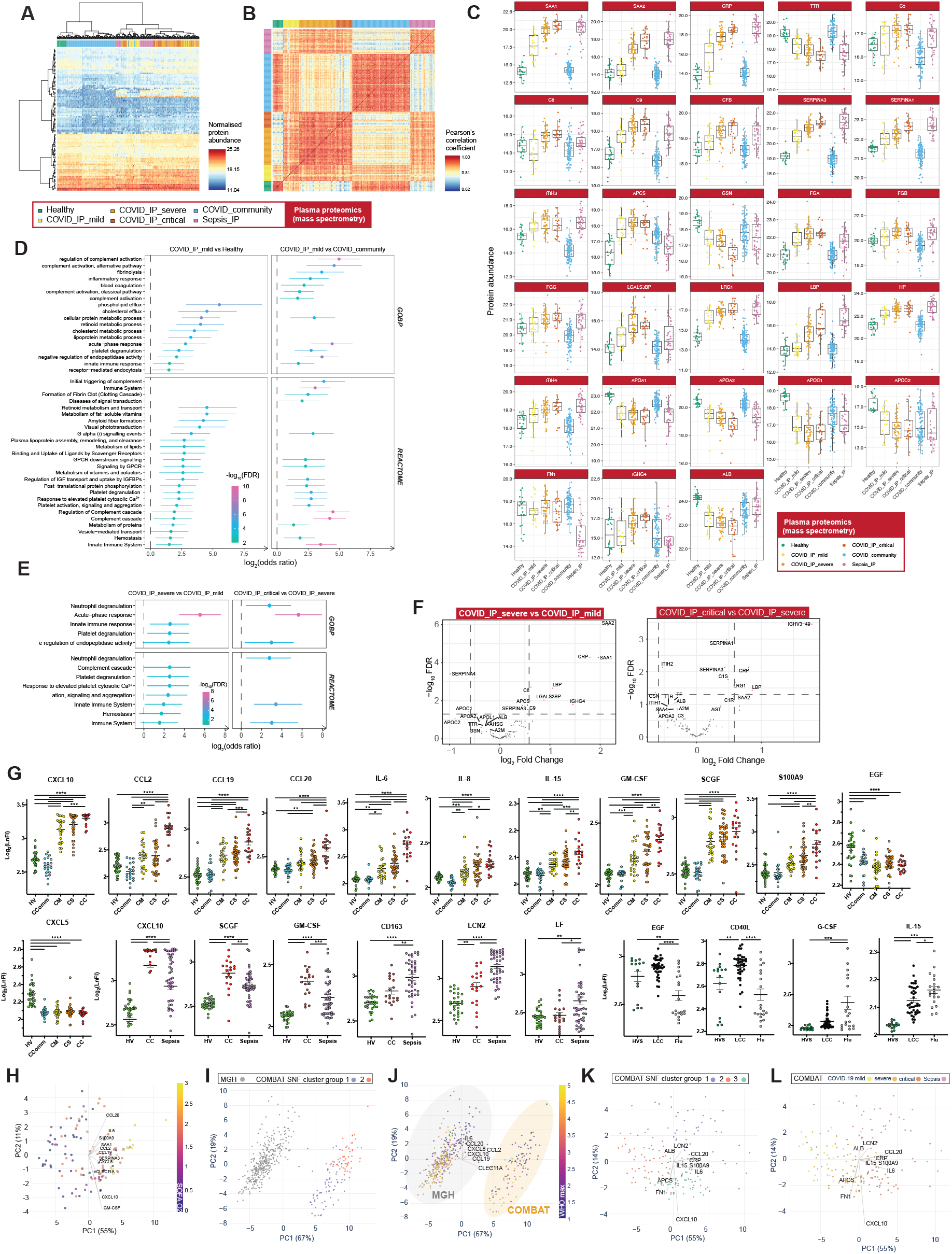
Plasma protein signatures and subphenotypes of COVID-19, related to Figure 6. (A-F) Plasma proteins assayed by HT-LC-MS/MS mass spectrometry (A) unsupervised clustering analysis all samples (B) supervised correlation analysis all samples (C) individual protein abundance across the comparator groups. Only one sample per patient at the maximal severity are plotted. (D,E) GOBP terms or Reactome pathways significantly enriched (FDR<0.05) in proteins differentially abundant contrasting samples from (D) mild hospitalised COVID-19 patients with those from healthy volunteers or from mild community COVID-19 cases and (E) severe vs mild or critical. (F) Pairwise contrasts, severe vs mild and critical vs severe COVID-19. (G) Individual protein abundance across the comparator groups assayed using luminex. (H) Similarity network fusion (SNF) for hospitalised COVID-19 patients from COMBAT (discovery) data using networks derived from sample-by-sample similarity matrix for mass spectrometry and luminex assays of plasma proteins, and shaded by SOFA_O2. (I,J) Mass General Hospital (Olink) validation data and COMBAT (discovery) cohorts shaded by (I) cluster group for COMBAT cohort only (J) shaded by WHO ordinal score (max severity) for all samples. (K,L) SNF clusters when analysing hospitalised COVID-19 and sepsis patients shaded by (K) cluster group (L) patient comparator group.*p< 0.05, **p< 0.01, ***p< 0.001, ****p< 0.0001 by One-way ANOVA with Tukey’s Multicomparison Test. Data are represented as mean ± SEM.

**Figure S7.**
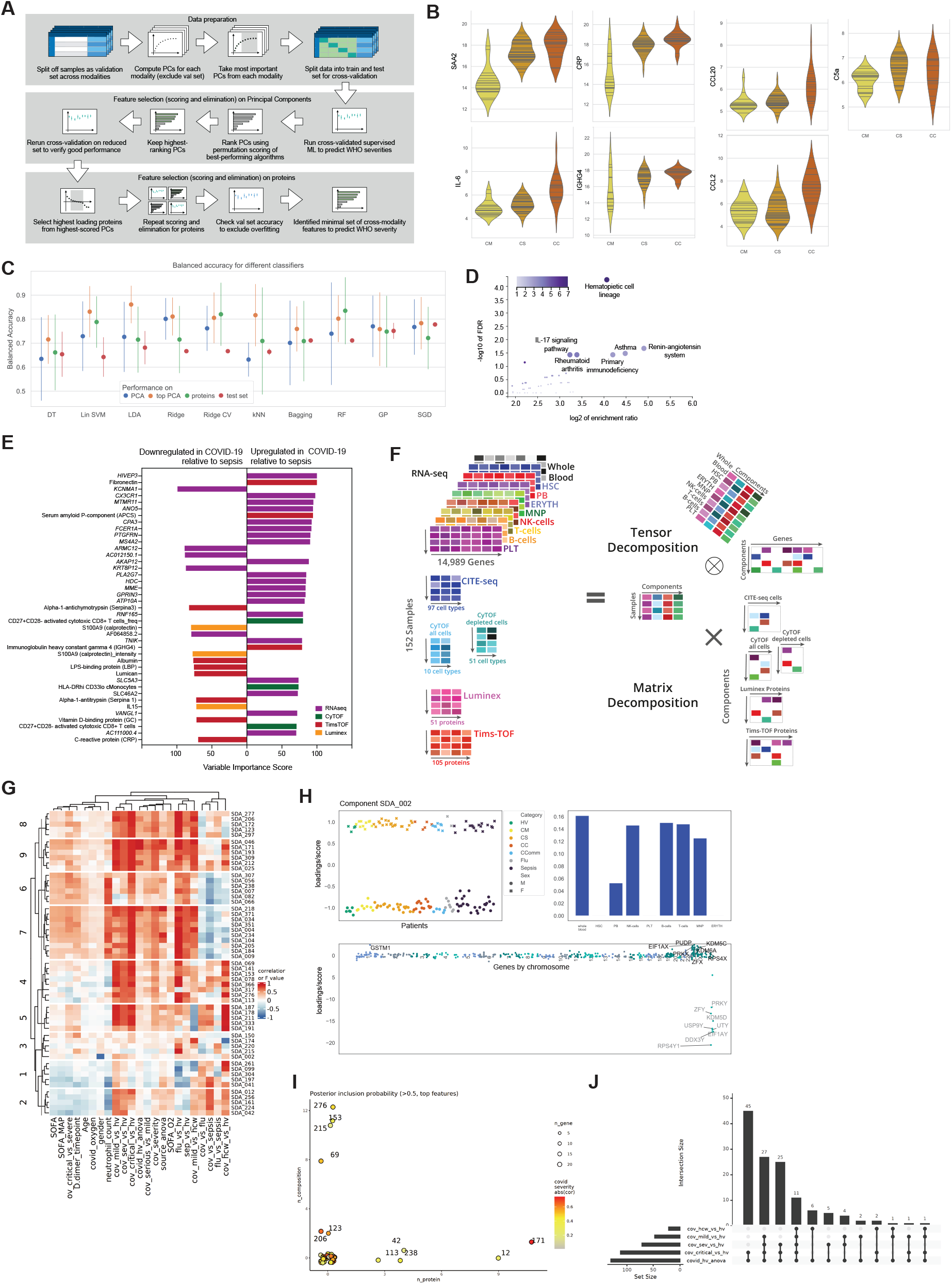
Integrative approaches define hallmarks of COVID-19 response, see Figure 7. (A-C) Machine learning feature selection for COVID-19 severity (A) summary of process followed (B) violin plots showing distribution of final selected predictive feature set across WHO severity groups (C) performance of the 10 best algorithms when run on all PCs, only the top-scored PCs, and the raw features extracted from the PCs (error bars are confidence intervals for the cross validation runs). We also show the accuracies from training the algorithms with the train+test sets and evaluating them on the validation set (averaged over 50 runs). (D,E) Machine learning to discriminate between sepsis and COVID-19 using plasma proteins, whole blood total RNAseq and mass cytometry as input variables in SIMON showing (D) enriched KEGG pathways on all features with variable importance score >50 (E) discriminating features with variable score >70. (F-J) Tensor analysis (F) tensor and matrix decomposition across multi-omic datasets showing datasets including 152 samples by 8 cell lineage clusters (scRNAseq, 22 missing samples) and whole blood (total RNAseq, 9 missing samples) by 14,989 genes; cell composition from CITEseq (152 samples by 64 pseudobulk cell types, 22 missing samples) and CyTOF (152 samples by 10 or 51 cell types, non-granulocyte depleted and depleted whole blood with 21 or 20 samples missing); and plasma proteins from luminex (152 samples by 51 proteins, 20 missing samples) and high throughput liquid chromatography with tandem mass spectrometry (152 samples by 105 proteins with 17 samples missing) (G) heat map summarising top components identified on pairwise contrasts involving clinical covariates, measures of severity and patient group (H) tensor component 2 showing loading scores and relationship with gender, with loading scores displayed for tensor involving differential gene expression cell lineage clusters and whole blood (I) feature types contributing to loading scores of the top components according to the posterior inclusion probability (J) component inclusion where significant on analysis of variance between COVID-19 source group and healthy volunteers (BH adjusted *P* <0.01) and absolute spearman’s *P* >= 0.5 (and BH adjusted *P* < 0.01) with at least one of the contrasts between the COVID-19 groups vs healthy volunteers.

## STAR Methods

### RESOURCE AVAILABILITY

#### Lead Contact

Further information and requests for resources and reagents should be directed to and will be fulfilled by Julian Knight.

#### Materials Availability

This study did not generate new unique reagents.

#### Data and Code Availability

Derived and processed data for all the datasets generated during this study and reported in this paper will be available including through this paper and the European Genome-phenome Archive (EGA). Web-based interfaces for visualising COMBAT datasets and outputs reported here are available at https://mlv.combat.ox.ac.uk/ and https://shiny.combat.ox.ac.uk. All code used for every algorithm followed in data processing and analysis is fully referenced within the specific methods text sections and Key Resources Table.

### EXPERIMENTAL MODEL AND SUBJECT DETAILS

#### Cohorts

The study was designed to allow deep molecular, multi-omic and immunological profiling of COVID-19 in peripheral blood at the outset of the pandemic in the United Kingdom during a public health emergency. Samples used were derived from multiple sources to allow comparison between adult patients with varying severities of COVID-19 and comparator disease or health states. A summary of the cohorts included in this study is provided below followed by a description of the methods used to define clinical parameters and handling of the blood samples. Demographic information and clinical phenotyping for all study subjects is summarised in **Table S2**, together with numbers of patients/samples assayed by cohort.

#### Hospitalised COVID-19 patients

Patients admitted to Oxford University Hospitals NHS Foundation Trust, UK were co-recruited into the Sepsis Immunomics study (a prospective observational cohort study applying an integrated immune -omic approach to understand why some patients have a severe response to infection), and the ISARIC/WHO Clinical Characterisation Protocol for Severe Emerging Infections, if they were found to have a syndrome consistent with COVID-19 and a positive test for SARS-CoV-2 using reverse transcriptase polymerase chain reaction (RT-PCR) from an upper respiratory tract (nose/throat) swab tested in accredited laboratories. Written informed consent was obtained from adults or personal/nominated consultees for patients lacking capacity, with retrospective consent obtained from the patient once capacity was regained. Ethical approval was given by the South Central–Oxford C Research Ethics Committee (REC) in England (Sepsis Immunomics REC reference 19/SC/0296; ISARIC WHO Clinical Characterisation Protocol for Severe Emerging Infections REC reference 13/SC/0149). Patients had whole blood sampled on days 1, 3 and 5 of either hospital or intensive care admission and were recruited during the SARS-COV-2 pandemic between 13 March and 28 April 2020. A selection of survivors were approached and asked to provide samples with consent under both research protocols at day 28 or more following discharge from hospital.

#### Healthcare workers with COVID-19

In order to provide a time of symptom matched set of samples from individuals with mild COVID-19 disease in the community, healthcare workers based at Oxford University Hospitals NHS Foundation Trust, UK with symptoms consistent with mild COVID-19 and a positive test for SARS-CoV-2 using RT-PCR from an upper respiratory tract (nose/throat) swab tested in an accredited lab were recruited into the Gastro-intestinal illness in Oxford: COVID substudy [Sheffield REC, reference: 16/YH/0247]. Individuals were consented and sampled at or after 7 days from the start of symptoms when the participants were returning to work.

#### Hospitalised patients with all-cause sepsis

In order to include samples from patients with sources of severe sepsis other than COVID-19, patients older than 18 years of age admitted to Oxford University Hospitals NHS Foundation Trust, UK with symptoms and signs of established sepsis (suspected infection with an acute change in total Sequential Organ Failure Assessment (SOFA) score of ≥2 points or a change in quick SOFA score by ≥2 points) were consented into the Sepsis Immunomics project [Oxford REC C, reference:19/SC/0296]) between 12 October 2019 and 13 March 2020. Patients were sampled on days 1, 3 and 5 of either hospital or intensive care admission and a selection of survivors were sampled, with additional consent at up to 6 months post-discharge.

#### Healthy volunteers

Following advertisement, interested individuals 55 years or over and self-reporting as healthy were consented and recruited into the Genetic diversity and gene expression in white blood cells study [South Central Oxford REC B, reference 06/Q1605/55]. Blood samples were collected on one occasion only.

#### Critically unwell patients with COVID-19 and influenza in London

In order to provide comparator samples from patients critically unwell with influenza, samples were included from patients ≥18 years old managed in an intensive care unit (ICU) for ≥24 hours requiring ventilator support with either a diagnosis of COVID-19 or severe influenza using the Aspergillosis in patients with severe influenza (AspiFlu ISRCTN51287266) study [Wales REC 5, reference 19/WA/0310]. Patients had blood sampled within 72 hours of enrolment that may occur within 7 days of admission onto ICU. Patients were recruited at one of three UK units including St George’s University Hospitals NHS Foundation Trust, Guy’s and St Thomas’ NHS Foundation Trust and King’s College Hospital NHS Foundation Trust. PCR diagnostics for influenza or SARS-CoV2 performed by accredited laboratories were used for diagnosis.

#### Clinical phenotyping

##### Clinical data capture

Healthy volunteers and healthcare workers with COVID-19 had age, sex and, where possible, self-reported ethnic background information collected. Information on previous medical history was not collected or stored for the healthy volunteer cohort but participants were invited on the basis of their self-reporting being ‘healthy’ and were deemed capable of self-presenting for study assessment. Healthcare workers were asked for the date of onset of symptoms (‘when they started to feel unwell’) and the number of days between this date and sampling was calculated for every participant. Any healthcare worker who was admitted to hospital or required oxygen had this information collected and retained and defined as maximal severity of illness using the World Health Organisation Criteria (see below).

A range of clinical data was collected and stored for downstream analysis using structured methodology from the hospitalised patients included in the study (COVID-19, all-cause sepsis and influenza).

#### Patient demographics

Sex and ethnicity were captured using electronic healthcare records (EHR). Age was calculated at the time of sampling or maximal severity of illness using dates of birth registered in EHR.

#### Patient medical history and risk factors

Smoking status was derived from clinical clerking or direct patient or next-of-kin questioning where possible. Estimated or calculated weight was available for the vast majority of patients. In London COVID-19 and influenza patients, body-mass index (BMI) was calculated through availability of estimated or measured height. In Oxford patients, height was rarely available and so BMI was estimated using sex and weight and correlation with measured BMI was acceptable in cases where BMI was available. BMI of 25 or over was defined as overweight. Index of multiple deprivation (IMD) quintile and Charlson comorbidity indices (2012 definitions) were derived from the EHR in Oxford and were not available for London patients. Previous medical problems in patients admitted to Oxford hospitals were defined using NHS UK Read Codes Clinical Terms Version 3 and converted into ICD10 codes using ‘TRUD’ (https://isd.digital.nhs.uk/trud3/user/guest/group/0/pack/9). Pre-existing medical conditions defined through ICD10 were defined through the consensus of at least one clinical opinion and in cases where there was uncertainty if a code was assigned to an acute presentation, pre-existing conditions were only assigned if they were present in the first episode of an admission, and not if they were only present in later admission episodes. For patients recruited in London, pre-existing diseases were defined by the study teams using clinical records and patient or relative questioning.

After defining pre-existing ICD10 codes or reported diagnoses, patients were classified as having the following conditions: hypertension, chronic respiratory disease (not explicitly available for London patients), asthma (not explicitly available for London patients but chronic obstructive pulmonary disease was), chronic cardiovascular disease, diabetes, haematological malignancy, other malignancy, liver disease, neurological disease (not available for London patients), chronic kidney disease, solid organ transplant, rheumatological condition (not available for London patients), significant immunosuppression (not available for London patients) and stroke / dementia (not available for London patients). An OpenSafely (OS) score was calculated for all patients using these classifications combined with BMI and IMD (Williamson et al., 2020). An adjusted OS score was calculated for the London cases that did not include some missing pre-morbid conditions and IMD quintile and the correlation was very high with the standard OS score (r=0.98).

#### Admission and disease timescales

Length of hospital stay was defined using hospital records for all hospitalised patients. Length of intensive care admission and duration of vasopressor, ventilation or, where appropriate extracorporeal membrane oxygenation (ECMO) and/or renal replacement therapy were all defined for hospitalised patients using EHR. All intervention and maximal severity timepoints were defined according to the date of onset of symptoms for each patient. This was defined by at least 2 independent clinicians through review of the clinical notes or direct questioning of the patient according to any unusual symptoms related to the current clinical condition. COVID-19 was defined by presence of at least one symptom consistent with COVID-19 and a positive microbiological test. Patients without symptoms were not approached for recruitment. Time between symptom onset and sampling was measured in days. All London patients were classified as critical requiring mechanical ventilation. No specific physiological observations were available for the London patients at the time of sampling and were only available at the time of admission onto ICU for calculation of the Acute Physiology and Chronic Health Evaluation (APACHE) and SOFA scores. Physiological observations were available from EHR for all hospitalised Oxford patients and were defined on the day of sampling at midday or the closest time before or after midday for each patient and timepoint including oxygen saturation, delivered fraction of inspired oxygen (either exact or estimated depending on delivery method: 0.24 for nasal cannula, 0.28 for simple face mask, 0.8 for non-rebreathe mask), pulse rate and blood pressure. Oxygen saturation: fraction of inspired oxygen ratio (SaO2:FiO2) was calculated as a proxy for partial pressure of oxygen:FiO2 ratio in the absence of complete arterial blood sampling for all patients. Days of oxygen therapy were defined in relation to sampling and maximal illness and total hospital stay for hospitalised COVID-19 patients where 2 or more timepoints of oxygen therapy were recorded. Fever was defined as 38.0°C and, if present at midday, was defined at present at time of sampling. Highest measured temperature 24 hours prior to midday was also captured using EHR for every timepoint and the presence of ‘persistent fever’ more measures of temperature ≥38°C in 24 hours before midday than temperatures recorded <38°C was also recorded for all sampling timepoints in hospitalised Oxford patients. In those patients where persistent fever was observed the time in days between onset of symptoms and end of persistent fever was calculated.

Maximal severity was defined for Oxford COVID-19 patients using SaO2:FiO2 ratio and the date of lowest value was defined through manual inspection of longitudinal physiological parameters available through EHR. The individual levels of SaO2, method and exact or estimated concentration of oxygen delivered and additional recruitment strategies such as paralysis or proning were captured. In cases where death occurred, this was defined as maximum severity of illness and time between symptom onset and maximum severity was defined thereafter.

All together this information was used to define WHO categorical (mild as no oxygen, severe as ≤93% oxygen saturation, and critical as requiring mechanical ventilation) and ordinal scales (https://www.who.int/blueprint/priority-diseases/key-action/COVID-19_Treatment_Trial_Design_Master_Protocol_synopsis_Final_18022020.pdf) for each timepoint of sampling and maximal severity of illness and these times were all aligned in days from onset of symptoms. Severity was also classified according to the WHO Ordinal scale (https://www.who.int/blueprint/priority-diseases/key-action/COVID-19_Treatment_Trial_Design_Master_Protocol_synopsis_Final_18022020.pdf) and Sequential Organ Failure Assessment score (Ferreira et al., 2001) together with the extent of lung disease and compromise (SOFA oxygenation score).

#### Other clinical and therapeutic tests and interventions

Full blood count differentials (haemoglobin concentration, platelet count, total white cell, neutrophil, lymphocyte, monocyte and eosinophil counts), highest lactate, C-reactive protein (CRP), alkaline phosphatase (ALP), alanine transaminase (ALT) and lowest bicarbonate were captured at the time of sampling for all hospitalised COVID, and sepsis patients using EHR. Equivalent results at the time of sampling were not available for London patients. D-dimer, lactate dehydrogenase (LDH) creatine kinase (CK) levels at the time of admission, sampling and maximal severity of illness were available for hospitalised Oxford patients but not London patients. Decisions on maximal levels of care based on senior lead clinician decision (frequently with consensus from senior intensive care clinicians) was captured and patients were defined as frail if the plan was not to offer multi-organ support.

Patients with computerised tomography (CT) images of the lungs and thorax had images assessed independently by three radiology or respiratory experts to define ground-glass or consolidation patterns of lung involvement, presence or absence of pulmonary embolus and extent of involvement.

The clinical suspicion and/or radiological confirmation of occurrence of any major arterial (e.g. stroke) or venous thromboembolic (deep venous thrombosis, pulmonary embolus) event was captured for each hospitalised Oxford patient and defined as a single binary outcome. Radiological diagnosis of pulmonary embolus was defined separately but compared to this definition to ensure consistency.

Co-prescription of agents hypothesised or proven to be effective in the management or treatment of COVID-19 were logged for all patients with COVID-19. These agents included dexamethasone, remdesivir, interferon 1beta, tociluzumab, anakinra, azithromycin, convalescent plasma or other experimental immunomodulatory agents.

#### Inclusion / Exclusion criteria and Early Stage Matching

Patients <18 years old or those with active malignancy or receiving significant immunosuppression (greater than an equivalent of 40mg once a day of prednisolone) prior to admission, or those with a clear alternative cause for symptoms and hospital presentation were excluded from analyses. For most modalities, samples were prioritised following a stepwise algorithm to match for age, sex and severity of illness. The steps for matching were as follows:

Select severe/critical hospitalized COVID-19 patients first

1. As many critical samples were identified at the earliest point of their maximal severity and samples from patients sampled during severe disease (not defined as frail) were matched in terms of age, sex and time between onset of symptoms and sampling as closely as possible.
2. Rejection sampling was used to select individuals from the comparator groups (mild COVID-19 including both hospitalised and community healthcare workers), healthy volunteers, influenza and all-cause sepsis) to match confounder distributions.

Some downstream analyses required multiple samples from the same individuals transiting through different disease states. In cases where data was available from multiple samples for the same individual a cross-sectional analysis involved prioritising a single sample based on closeness to maximal severity of illness.

### METHOD DETAILS

#### Blood sample processing

Whole blood from hospitalised Oxford patients, healthcare workers and healthy volunteers were sampled into Tempus tubes (Life Technologies Corporation) for extraction of whole blood total RNA for sequencing or DNA for genotyping or sequencing, or EDTA buffered vacutainers (Fisher Scientific) for processing within 4 hours of sampling. Processing consisted of mixing of whole blood in a 1:1 ratio with Cytodelics (Cytodelics), and then fractionated using Leucosep (Griener Bio-One) into peripheral blood mononuclear cells (PBMC) and EDTA-buffered plasma. PBMCs and plasma were frozen and thawed in batches for specific experiments to minimise the risk of batch effects. Tempus tubes were frozen at -80°C until extraction of RNA performed in batches.

#### Immune cell profiling using mass cytometry

##### Sample processing and antigen staining

On the day of staining, Cytodelics stabilised samples were thawed, processed to remove red blood cells and fixed using Whole Blood Processing Kit (Cytodelics) as per manufacturer instruction. Two control samples from 2 healthy volunteers (Control A and Control B) were included to each batch to assess batch to batch variations. Fixed cells were distributed in 96 deep well V-bottomed plate, washed once with CSB containing Benzonase, and then barcoded using the Cell-ID 20-Plex Pd Barcoding Kit (Fluidigm).

Briefly, cells were washed once with Barcode Perm Buffer, resuspended in Barcode Perm Buffer supplemented with Heparin to reduce nonspecific eosinophil staining artifacts (Rahman et al., 2016) and loaded with half the amount of metal barcodes recommended by the manufacturer. After 30m incubation at room temperature, cells were washed twice in CSB, pooled and counted. 40-70% of cells in whole blood are CD66+ granulocytes, and their frequency can increase to up to >95% in septic patients. To maintain both the possibility to measure changes in the frequency of granulocytes and to analyse with a reasonable efficiency mononuclear cells in samples from COVID-19 and sepsis patients, each batch was split in two aliquots. The first aliquot was stained with no enrichment/depletion done (i.e. unaltered). The second aliquot was enriched for mononuclear cells using a granulocytes depletion kit based on the magnetic separation of CD66+ cells using beads coated with anti-CD66 antibodies (EasySep; 17882). Both granulocyte depleted and non-depleted samples were first stained with the surface antibody cocktail for 30m at room temperature), washed with CSB, then fixed with Nuclear Antigen Staining Buffer and permeabilized with Nuclear Antigen Staining Perm before staining with an intracellular antibody cocktail. After staining at room temperature for 45m cells were washed in CSB buffer, and fixed in 1.6% FA solution before overnight incubation with Cell-ID™ Intercalator–Ir. On the day of acquisition, cells were washed once in CSB buffer supplemented with Benzonase, resuspended in water and run at an acquisition rate of <300 events/second and acquired on a Helios CyTOF machine. An additional aliquot of whole blood was thawed and stained for those samples where yield was too low after staining and acquisition. In total 31 samples were acquired twice (in different batches).

#### Flow cytometry

For flow cytometry we analysed one aliquot of the same PBMC samples at the same time they were being processed for 10x Genomics CITEseq. Each aliquot was split into three (to have 5-10×10^5^ cells/staining) and distributed in 96 well V-bottom plates. Cells were spun at 500g for 10 min, then pellets stained form 10min at RT in PBS with 15μl live dead Aqua, diluted as per manufacturer’s instructions) supplemented with 165μg/well human Ig (Gammanorm, Octopharma), to block non-specific Fc receptors binding. Cells were washed with 200μl FACS buffer (PBS 10% FCS), 10min 300g and pellets were stained with each antibody mix in Brilliant Stain Buffer (BD Biosciences), 20min at RT. Cells were with 200μl FACS buffer, 10min 300g and fixed in IC Fixation Buffer (Thermo Scientific) diluted 1:2 in PBS, total 200 μl/well. Cells were fixed a minimum of 30min and a maximum of overnight before being acquired at the BD Symphony X50.

#### Whole blood total RNAseq

Total RNA-seq was performed with libraries prepared by Oxford Genomics Centre with the NEBNext Ultra II Directional RNA Library Prep Kit for Illumina after rRNA and globin depletion kits. Libraries sequenced as a single pool of 144 samples (124 patients) on one NovaSeq S4 flow cell (4 lanes) with a target of 50M 100bp read pairs per sample. Due to limited material availability influenza samples were sequenced used a small-bulk RNA-sequencing method in order to facilitate genetic demultiplexing of the CITE-seq data.

#### Bulk BCR and TCR sequencing

Bulk isotype-resolved B cell receptor sequencing was performed on RNA from whole blood using a protocol adapted from (Bashford-Rogers et al., 2019), with moderations including the use of a more sensitive reverse transcriptase (Superscript IV) and primer concentrations (primer sequences provided in **Table S4**). Bulk TCR sequencing was based on the same protocol adapted from (De Mattos-Arruda et al., 2019), in which the TRB primers were redesigned to capture the full productive repertoire (primer sequences provided in **Table S4**). Sequencing libraries were prepared using Illumina protocols and sequenced using 300bp paired-ended sequencing on a MiSeq (Illumina).

#### 10x Genomics Chromium CITEseq

To minimise batch effects, cryopreserved PBMCs were processed in ten batches of 14 samples per batch, with each batch containing at least one sample from each comparator group and similar distribution of patients age and sex across the batches. After thawing, all 14 samples were mixed together to form a single pool (at a ratio that yielded the same number of live cells pooled per sample based on live/dead counting) followed by viability staining (7-AAD dye, BioLegend 420404) and live/dead sorting on a BD FACSAria Fusion sorter.

Post sort, each pool was incubated with FcX block (BioLegend) for ten minutes on ice, washed, and stained with a 192 TotalSeq-C antibody panel (BioLegend 99814) for 30 minutes on ice. Cells were washed three times in PBS + 1% BSA, counted on the BioRad TC20 Automated Cell Counter and loaded onto the 10X Genomics Chip G at 50,000 cells per channel. Each pool was loaded across seven channels.

#### scVDJ-seq data generation

scVDJ data was generated on the same cells as the scCITE-seq and GEX datasets. Cell Ranger software (v3.1.0) was used to process the Chromium scRNA-seq output data. The FASTQ files were filtered per sequence library plex (i.e. per-pool) using the 10x Genomics index-hopping-filter (https://support.10xgenomics.com/docs/index-hopping-filter) that implements a strategy to mitigate the known index hopping issue with the Illumina machines that use patterned flow cells. The IMGT’s reference genome was used as a reference for the BCR and TCR VDJ libraries. Cell Ranger was also used to filter read alignments, cell barcodes and UMIs, excluding those droplets with low numbers of reads (e.g. erythrocytes or droplets with encapsulating ambient RNA from dying cells).

#### 10x Genomics scATACseq

Similar to the CITE-seq approach, cryopreserved PBMCs were processed in two batches of five samples per batch with each batch containing at least one sample from each comparator group. Each sample underwent viability staining (7-AAD dye, BioLegend 420404) followed by live/dead sorting on a BD FACSAria Fusion sorter. Post sort, five samples of an equal number of cells were mixed together to form a single pool, and each pool underwent cell lysis and nuclear extraction according to the 10X demonstrated protocol Nuclei Isolation for Single Cell ATAC Sequencing CG000169 Rev D. Briefly, 200,000 cells from each sample were added to form the pool and cell lysis was performed with 100 μl chilled Lysis Buffer (10 mM Tris-HCl (pH 7.4), 10 mM NaCl, 3 mM MgCl_2_, 0.1% Tween-20, 0.1% Nonidet P40 Substitute, 0.01% digitonin and 1% BSA) for three minutes on ice. The lysis reaction was quenched with 1 ml chilled Wash Buffer (10 mM Tris-HCl (pH 7.4), 10 mM NaCl, 3 mM MgCl2, 0.1% Tween-20 and 1% BSA) and the nuclei were centrifuged (500*g* for 5 min at 4 °C). After removal of supernatant, the nuclei were resuspended in chilled diluted nuclei buffer (10X Genomics; 2000153) and concentration determined with the Biorad TC20 Automated Cell Counter. Each pool of nuclei was loaded across four channels of the 10X Genomics Chip E (15,000 nuclei per channel).

#### Tims-TOF mass spectrometry

##### Sample preparation of non-depleted plasma/serum for LC-MS/MS analysis

###### Samples

Plasma and serum samples were stored at -80°C then thawed overnight at 4°C before use.

###### Protein precipitation and protein digestion using S-trap

For the bottom-up proteomics approach, proteins were precipitated using Isopropanol (IPA) and loaded into the S-trap 96-well plates (Profiti, Huntington, NY, USA) where proteins were retained for subsequent trypsin digestion following S-trap manufacturer instructions (Zougman et al., 2014). Briefly, five microliters of plasma or serum were pipetted under a category 2 fume hoods into standard 96-well plates pre-filled with 200 µl of isopropanol following the 96-well plate layouts. Precipitation plates were kept at room temperature for 1h before transferring the isopropanol – plasma (serum) mixture into the 96-well S-trap plates. 200 µl of S-trap binding buffer (90% methanol, 100mM Triethylammonium bicarbonate (TEAB)) were added into each well of the precipitation plate to recover any protein precipitates and transferred into the corresponding 96-well S-trap plates. Plates were then spun at 1500g for 2 min. Protein colloidal precipitates were retained into the S-trap mesh. S-trap plates were then subjected to four consecutive washes with S-trap binding buffer (200 µl for the first wash and 350 µl the three remaining), followed by a 2 min spin at 1500g. 125 µl 50mM TEAB containing trypsin (Worthington) to a 1:25 wt:wt ratio was added into each well and incubated overnight at 37°C. Tryptic peptides were sequentially eluted from the S-trap into a clean 96-well plate with 80 µl of 50mM TEAB, 80 µl of 0.2 % formic acid (FA) in LC-MS water and 80 µl of 0.2% FA in 50% acetonitrile with 2min spins at 1500 x g after each solvent addition. Finally, plates containing tryptic digests were dried in the speed vac and reconstituted with 70 µl of 0.1 % formic acid (FA) for subsequent LC-MS/MS analysis after two serial dilutions (1/100 and 1/10) in 0.1 % FA for a final 1/1000 dilution plate to achieve suitable peptide concentration for LC-MS/MS loading.

##### Pools

For quality control (QC) purposes and for repeat injections, a pool for each clinical sample group was created and processed as described above. In addition to the sample group pools, an overarching master pool for each of the plasma and serum cohorts was also built from the individual non-diluted tryptic peptide sample group pool with master pools built to reflect the contribution of each individual group to the overall study.

#### High pH fractionation of Master pools to generate plasma/serum libraries

To increase the depth of the non-depleted plasma/serum proteome, master pools were subjected to a high pH fractionation using the SOLA HRP cartridges (Thermo Fisher) with the aim to create a protein library. In brief, an equivalent of 150µg of peptides from each master pool was fractionated into eight different fractions going from 0% to 70 % Acetonitrile in 10mM ammonium formate at pH 10. Prior sample loading, SOLA HRP columns were conditioned with 0.1% triflouroacetic acid (TFA) in 70% acetonitrile and washed with 0.1% TFA. Following the sample loading, columns were washed with 0.1% TFA before starting the stepwise high pH fractionation elution with 200µl of 0%, 10%, 15%, 20%, 25%, 30%, 40% and 70% ACN in 10mM ammonium formate pH 10. Eluted fractions were then dried in a speed vac and reconstituted with 40µl of 0.1 % FA prior to LC-MS analysis.

#### LC_MS/MS using the high-throughput Evosep One - Bruker TimsTOF Pro platform

C18 Evotips (Evosep one, Odense Denmark) were prepared following manufacturer’s instructions. Briefly, Evotips were conditioned with 1-propanol, washed with solvent B (0.1% FA in 100% acetonitrile) and equilibrated in solvent A (0.1% FA in LC-MS water) before loading them with a total volume of twenty microliters containing 10 µl of sample (peptides) from dilution 1/1000 plate (equivalent to an estimated 30 ng of peptides). Evotips were washed once with solvent A and stored in 100 µl solvent A till the sample injection.

Samples were analysed using a Evosep One LC system connected to the TimsTOF Pro mass spectrometer (Bruker Daltonics). Peptides were analysed using the pre-built 100 samples / per day method (Evosep) with an 11.5 min gradient (total cycle time of 14.4min) at a 1.2 µl/min flow rate (Bache et al., 2018). In brief, tryptic peptides were transferred from the pre-loaded C18 Evotips with a pre-built gradient to a sample loop and separated on an 8cm C18 analytical column (Evosep Pepsep, 3um beads, 100 um ID) with an overall gradient from 3 to 40% acetonitrile.

Mass spectrometry data were acquired in PASEF mode (oTOF control v6.0.0.12). The ion mobility window was set to 1/k0 start = 0.85 Vs/cm2 to 1/k0 end = 1.3 Vs/cm2, ramp time 100 ms with locked duty cycle, mass range 100 - 1700 m/z. MS/MS were acquired in 4 PASEF frames (3 cycles overlap). Target intensity was set to 6000 and threshold intensity 200. The data acquisition method has been deposited within the raw data in the ProteomeXchange data repository (PXD023175) (Perez-Riverol et al., 2019).

To assess the technical reproducibility, master pools were run before each sample group and sample group pools were run after the master pools, every twenty sample runs and after the last sample of the group. Blanks were injected in between groups to control for carry over.

#### Luminex assay

The concentrations of selected proteins in the plasma and serum were measured with Human Magnetic Luminex Kits (Bio-techne) with 3 panels containing total 51 analytes: C-C motif ligand (CCL)2/3/4/11/17/18/19/20, CD40 Ligand (CD40L), CD163, complement component 5a (C5a), C-X-C motif chemokine ligand (CXCL)1/5/10, epidermal growth factor (EGF), basic fibroblast growth factor (FGF2), granulocyte colony-stimulating factor (G-CSF), granulocyte-macrophage colony-stimulating factor (GM-CSF), granzyme B (GrB), interferon (IFN)a/b/g, interleukin (IL)- 1a/1b/2/3/5/6/8/10/12/13/15/17A/23/33, lactoferrin (LF), Lipocalin-2 (LCN2), Lymphotoxin-alpha (LT-a), macrophage colony-stimulating factor (M-CSF), Myeloperoxidase (MPO), beta-nerve growth factor (b-NGF), Oncostatin M (OSM), S100 calcium-binding protein A9 (S100A9), stem cell growth factor (SCGF), tissue factor (TF), tissue factor pathway inhibitor (TFPI), transforming growth factor alpha (TGF-a), Thrombopoietin (THPO), tumor necrosis factor (TNF) and triggering receptor expressed on myeloid cells 1 (TREM-1). The assays were conducted according to the manufacturer’s instruction. Results were obtained with a Bio-Rad Bio-Plex® 200 Systems. The data of fluorescence intensity (FI) from the assays were used for further analysis.

### QUANTIFICATION AND STATISTICAL ANALYSIS

#### Statistical and unsupervised analysis of clinical phenotyping using clustering

To analyse demographic and clinical features cohorts the following statistical tests were applied. For approximately gaussian distributed continuous data we used the anova test (using the function f_oneway from python’s scipy.stats package). For other distributions of continuous variables, we used the Kruskal-Wallis-test (using the function kruskal from python’s scipy.stats package). For categorical data we used the chi-squared test test (using the function chi2_contingency from python’s scipy.stats package).

Unsupervised and consensus k-means clustering were used to analyse the clinical data (Moniti et al., 2003; Seymour et al., 2019). We prepared the data by applying quantile-normalization to counts and shift+rescaling to zero mean and unit variance to all features (applying quantile normalization to all features, not just counts, further improves the correlation of the ordered consensus matrix with the who severity_at_sample classes). We then ran consensus k-means clustering to identify the best number of clusters. This means we sample from our clinical dataset 50% of the entries, run k-means clustering (where we vary k, i.e. the number of clusters), and record how often two elements were clustered together out of the time where they were in the 50% of samples data points in a so-called consensus matrix. We repeated this process 600 times for k=2,3,4,5,6 clusters. For each k, we computed the empirical cumulative distribution (CDF) and the change in the area under the CDF curves for different k. The optimal cluster number is obtained for the k after which the CDF curves (and hence the areas) do not change significantly anymore.

We then performed an unsupervised consensus k-means clustering analysis of (subsets) of the clinical variables (using python’s sklearn library). In order to mitigate the question of which features are independent, we performed this analysis on the principal components (using sklearn). After consensus clustering, we sorted the consensus matrix using hierarchical clustering. The methods were implemented in python and used the packages numpy, scipy and pandas. We also performed hierarchical clustering directly on the PCs, rather than on the consensus matrix (using AgglomerativeClustering and dendrogram from sklearn).

#### Mass cytometry data analysis

##### CyTOF data pre-processing

After acquisition, data was normalised and concatenated on CyTOF Software v7.0, compensated on CATALYST (version 1.5.3.23). Then, data was further processed for removal of beads and of DNA negative events and for Gaussian parameters using a manual gating strategy using FlowJo v10.6 as described in **Methods S1 Figure 1**. Then, samples were manually debarcoded (FlowJo v10.6) and CD45+ events (non-depleted samples), or CD66-Siglec8-CD45+ (granulocytes-depleted samples) events were selected for further processing. For each batch, two control samples were run together with the patients’ samples to allow for correction for batch effects. Batch correction was performed with CytoNorm software (version 0.0.5) separately for granulocyte depleted and non-depleted samples. For both analyses, the training of the algorithm was done with 20000 cells per sample and 5 clusters (parameter nClus) using one of the control samples and the results were assessed using the other control sample. Doublet cells were removed by manual gating on FlowJo v10.6 (**Methods S1 Figure 2**) and were deemed as Iridium^high^Ki67^-^ cells to avoid removing proliferating cells. Finally, to correct for potential biases due to highly varying cell numbers per sample, downsampling was performed to a maximum 75000 cells per sample for the granulocyte depleted samples and 40000 for the non-depleted samples.

#### Clustering

Clustering analysis and the identification of the different immune cell population was done using the analytical pipeline described in **Methods S1 Figure 3**. Clustering analysis was performed separately for granulocyte depleted and non-depleted samples using a self-organising map algorithm through an implementation of FlowSOM via the CATALYST R package (version 1.10.3). The initial clustering was performed using a resolution of 144 clusters (dimensions 12x12) and metacluster merging by consensus clustering was performed at a resolution of 25 clusters for both datasets. The clusters were then manually annotated to define the populations. Three major populations (T/NK, B/Plasmablasts and Myeloid) from the granulocyte depleted samples were further clustered to identify finer subpopulations using a resolution of 225 (dimensions 15x15) for T/NK and 144 (12x12) for the other two. The metacluster merging was performed at a resolution of 50 clusters for the T/NK population, 25 for the B/Plasmablast and 30 clusters for the Myeloid population. Clusters were manually annotated and further managed to provide cell type resolution at two different depths - one to define major cell types, and a finer one to define subpopulations. The cell type assigned to each cell ID in each of the three major population was then used to reconstitute the frequency of each population inside the granulocyte depleted sample. The finer annotation was used to evaluate the frequency of cell subtypes.

#### Differential abundance analysis

Differential abundance analysis was performed using code from the diffcyt package (version 1.8.8) with the option testDA_edgeR. The analysis was adjusted for the confounding variables batch, age and sex. For the analysis of the non-depleted samples the normalisation for composition bias was deemed necessary to account for the pronounced differences in neutrophil proportions. For all other analyses no normalisation was performed. The subpopulations of the granulocyte depleted samples were analysed independently of each other to avoid composition effects.

#### Manual gating and CD38 median intensity (MI)

Two subpopulations of cells were more precisely defined by manual gating rather than by clustering. To do that, the cluster of the parent population (cMono or MAIT) was exported from the R environment and gated in FlowJo. Also, the median intensity of CD38 expression were identified in different cell types within the granulocyte depleted samples.

#### Density plots

The density plots were made by downsampling to have the same number of events across all different conditions and using the geom_pointdensity function of the ggpointdensity package, with an adjust of 4, to generate the ggplot.

#### Integration of CyTOF and CITE-Seq data

The two datasets were aligned using Seurat (v3.9.9.9010) using only samples that had been analysed on both technologies (115 samples, after exclusion of one sample due to low number of cells in CITE-Seq). The integration was based on an overlapping set of 38 markers that were common between the CyTOF and ADT antibody sets. Both datasets were filtered to exclude cells with unclassified or uncertain annotations. The CITE-Seq dataset was further filtered to exclude potentially low-quality cells with i) number of genes, or i) number of features of ADT, or iii) number of total UMI (RNA) or iv) number of total UMI of ADT lower than 0.001 of their relevant distributions. The two datasets were then downsampled to 1000 cells per sample and the integration was performed on 230,000 cells in total.

The anchors between the query and reference datasets were found based on a CCA with 30 dimensions and the labels were transferred using a PCA for the weight reduction of the query dataset. The analysis was performed with either the CITE-Seq or the CyTOF as reference datasets in turn and all cells were given a predicted annotation from their counterpart dataset. Then the predicted annotation of the cells was compared to their original annotation and all the common major cell types were validated with very high accuracy (**Figures S1L, S1M**). Finally, the visualisation of the two datasets was performed with the CITE-Seq data as a reference. The anchors were used to impute the ADT markers for the CyTOF cells which were then merged with the CITE-Seq data and centred in order to run a UMAP analysis to visualise all the cells together (**Figures 1F,G**).

#### Flow cytometry data analysis

Data were analysed with Flowjo version 10. Cells were gated on live leukocytes and recompensated in Flowjo. Frequencies of individually gated populations were exported and plotted in Prism. Alternatively, 15000 CD3 cells per sample were concatenated and exported for subsequent analysis in R.

#### Whole blood total RNAseq analysis

##### RNA-sequencing data processing

We trimmed adaptor sequences using TrimGalore (v0.6.2, ref https://github.com/FelixKrueger/TrimGalore), and aligned reads to the reference genome (GRCh38.100) using multi-sample 2-pass mapping with STAR v2.7.3 (Dobin et al., 2013) (ENCODE Best Practices recommended parameters). We quantified gene expression using featureCounts (v1.6.4) (Liao et al., 2014) and annotations from Ensembl (v100). We calculated and checked QC metrics from FastQC,mapping metrics from STAR, duplication rates and other RNASeq metrics from Picard (v2.23) (http://broadinstitute.github.io/picard/) and RNASeQC (v2.3.5) (DeLuca et al., 2012) and checked for outliers in principal component analysis. We removed 1 sample with high proportions of duplicates and short reads, low exonic rate, number of mapped reads and number of genes identified, and which was also clearly outlying on PCA. We also confirmed that there were no sex mismatches that could indicate sample mix-ups based on sex chromosome gene expression using QTLTools (Delaneau et al., 2017). This resulted in a data set of 143 samples from 123 patients. We filtered out features that did not have at least 10 reads in at least 10 samples, retaining 23,063 features for downstream analysis. We then normalised the data using the trimmed mean of M-values method R package from edgeR; https://doi.org/10.1093/bioinformatics/btp616 and log2-transformation of counts-per-million. Code is available on GitHub (https://github.com/COMBATOxford/bulkrnaseq/mapping/).

#### Exploratory analysis

We carried out principal component analysis (PCA) on the normalised filtered data for 123 patients using prcomp (R v 3.6.2) with default parameters. We additionally performed PCA on just the samples from hospitalised COVID-19 patients (n=64). We selected informative PCs by taking the first n PCs that cumulatively described at least 80% of variance in the data. We explored the relationship between these PCs and the clinical variables available (transformed and filtered as described above) using Spearman correlation. Additionally, we investigated the biological relevance of the PCs through pathway enrichment analysis with the R package XGR (Fang et al., 2016b) and Reactome pathways. For each PC, we took the 500 genes with the highest absolute loading scores as input and compared to a background of all detected genes.

#### Differential expression

We performed differential expression analysis on the normalised data with one sample per patient using the limma R package (https://doi.org/10.1093/nar/gkv007). We included age, age^2^ and sex as fixed effects and compared each patient subgroup with all others, as well as investigating COVID-19 severity as a quantitative trait (mild=1, severe=2, critical=3). We defined significance for downstream analysis as FDR <0.05 and fold change >1.5. We investigated the impact of variation in cell proportions across samples calculated from hospital measurements for neutrophils, monocytes, and lymphocytes, by adding neutrophil and monocyte proportions to the model and comparing the estimated fold changes to the results to the more basic model. Pathway enrichment analysis was performed using Reactome pathways via the XGR R package (Fang et al., 2016b), with Fisher’s exact test and filtering of redundant terms by the xEnrichConciser function.

#### Weighted gene correlation network analysis

We applied weighted gene correlation network analysis (WGCNA) to describe modules of highly correlated genes within the whole blood total RNA-sequencing data (i.e. 143 samples from 123 patients) (Langfelder and Horvath, 2008). We utilised the “cornet” pipeline to wrap the WGCNA R package and perform gene set enrichment analyses (https://github.com/sansomlab/cornet.git). In brief, a stepwise approach of correlation network construction and module detection was adopted using log2-transformed counts-per-million RNA-sequencing data. Using the soft thresholding power of 4, a signed-hybrid network was built implementing the biweight midcorrelation as the adjacency function. The adjacency matrix was transformed into a topological overlap matrix in order to calculate the dissimilarity and a dissimilarity threshold of 0.3 was used to merge modules with very similar expression profiles. Module eigengene values (module first principal component) were used to summarise modules and perform module-trait correlation analyses (Pearson correlation). Gene set over representation analyses were performed using the default settings of the “cornet” pipeline (https://github.com/sansomlab/cornet.git).

#### CITE-seq: pre-processing and multi-modal annotation

##### Preprocessing of 10X Libraries (Gene expression, ADT, TCR, BCR)

As described above a total of n=140 PBMC samples from COVID-19, sepsis, influenza and healthy volunteers were mixed into n=10 pools. Each pool comprised of 14 samples (each from a different individual) and cells from each pool were captured using n=7 10x channels (**Methods S2 Figure 1**). From each channel, four libraries were generated, Gene expression (GEX), Surface proteome (ADT), TCR and BCR repertoires. The libraries were sequenced on an Illumina’s NovaSeq 6000 sequencer across nine S4 flowcells (4 lanes per flow cell). For each of the sample pools, n=7 GEX and ADT libraries were sequenced on 3 dedicated lanes (to eliminate index hopping between sample pools). The BCR/TCR libraries were sequenced on the remaining 6 lanes. Mapping indexes for the bulk and single-cell data were built using GRCh38 genome sequences and Ensembl version 100 annotations. FASTQ files were generated using Cellranger (v3.1.0) mkfastq and individually QC’ed using FastQC (Andrews, 2010). To mitigate index hopping between channel libraries within sample pool FASTQ files were filtered per pool using the 10x Genomics index-hopping-filter (https://support.10xgenomics.com/docs/index-hopping-filter) (v1.0.1). Per-channel (n=70) GEX and CITE-seq matrices were prepared using Cellranger (v3.1.0) count. Cell identification was performed on the GEX modality with both Cellranger (v3.1.0) and EmptyDroplets (Lun et al., 2019) (1.8.0) and the union of the calls from both algorithms taken forward as the set of identified cells.

#### Genetic demultiplexing of 10X GEX data

The GATK variant calling pipeline (https://github.com/gatk-workflows/gatk4-rnaseq-germline-snps-indels) (v4.1.7.0) was applied to the bulk RNA-seq data to identify sequence variants for the genetic demultiplexing of the single cell data. Minor modifications were made to the variant calling pipeline to allow execution from FASTQ files, incorporate metadata into the BAM alignment files, and produce a block gVCF file. We applied both Demuxlet (Kang et al., 2018) (v2; https://github.com/statgen/popscle/commit/038044660337b50ec89b0b355493ceb707f18ecd) and Vireo (Huang et al., 2019) (v0.4.0; https://github.com/single-cell-genetics/vireo/commit/4aecb54a4f2fa3a3413e2cd3e9f3515744385cba<u>), proceeding with the demultiplexing calls from Vireo as they were more robust to variation in read depth. The dataset was demultiplexed using full genotypes (per-pool gVCF files) from the bulk sequencing. For each of the n=70 10x channel sequence libraries, Vireo consistently demultiplexed 60 – 75% of cells. A total of n=884,587 singlet cells were demultiplexed.</u>

#### QC of 10X GEX data and preparation of GEX matrices

The pre-processing workflow was encapsulated in a set of CGAT python pipelines (Cribbs et al., 2019) and version controlled. Cell quality was assessed with a suite of metrics that included total UMI (GEX), number of genes detected, percent mitochondrial gene expression, percent ribosomal gene expression, percent IgG expression, scrublet doublet score (Wolock et al., 2019) and total UMI (ADT). Ambient RNA was assessed. Cell QC statistics, demultiplexing assignments and cell and patient metadata were centrally warehoused in an sqlite database. Following inspection of the QC metrics the dataset was filtered to retain cells with ngenes > 300 and pct_mitochondrial < 10%. In total, n=836,148 cells were selected for downstream analysis. Expression data for selected cells was extracted from the per-channel count matrices using R and combined into a single market matrix with AWK. RNA velocity matrices were computed for each of the channels with Velocyto (http://velocyto.org/) (La Manno et al., 2018) (v0.17.5). Velocity data for selected cells was extracted and combined into a single matrix using the Loompy Python library (http://loompy.org).

#### Multimodal cell annotation strategy overview

We used expert immunological knowledge to guide a curated integration of the data from the different modalities (GEX, ADT and VDJ) to identify and label the cell sub-populations present in the CITE-seq dataset (**Methods S2 Figure 2).** As detailed below, we first performed separate clustering of gene expression, clustering of surface protein expression and analyses of T and B cell receptor V(D)J sequences. Next, led by expert understanding of the three feature spaces we prioritised use of ADT surface phenotype for definition of major cell lineages and subsets where definitive marker expression was available. Cell types and subsets were further refined using information from the repertoire and GEX layers, or in the absence of definitive ADT information were identified by GEX cluster phenotype. Finally, the identified cell types and subsets were further divided by inferred functional state based on targeted assessment of information from all three modalities. For example, cell cycle phase was determined by GEX phenotype, T cell memory vs effector status was distinguished using information from both the GEX and ADT layers, while assignment of B cell maturation status involved use of information from all three modalities (including BCR mutational status). Information from all three modalities was used to identify and exclude doublets from downstream analysis.

#### Alignment and clustering of 10X GEX data

Prior to alignment data was normalised and log1p transformed using Scanpy (Wolf et al., 2018). The top n=3000 highly variable genes (HVG) were selected (Seurat flavour with n_top_genes set). The following MHC and variant immune receptor genes were excluded from HVG identification A/B/C/D/E/F/G]*, IGH[D/J/V]*, IGK[J/V]*, IGL[J/JCOR/L/ON/V]*, TRA[J/V]*, TRB[D/J/V/VA/VB]*,TRD[D/J]* and TRG[J/JP/V/VA/VB]*). Effects associated with total number of UMIs were regressed out and PCA components computed. We evaluated alignment with Harmony (Korsunsky et al., 2019), Scanorama (Hie et al., 2019) and BBKNN (Polanski et al., 2020) and chose to proceed with Harmony following inspection of UMAP projections of the alignment results (data not shown). Following inspection of the PCA scree (knee/elbow) plot, Harmony alignment of samples (n=140 levels) was performed in Python using the top n=65 PCs. Leiden clustering, marker discovery (wilcox tests) and visualisation (with violin plots, UMAPs, volcano plots, MA plots and heatmaps), automatic cell type identification (with singleR (Aran et al., 2019) and via over-representation analysis of xCell (Aran et al., 2017) genesets), basic composition analysis, geneset over-representation analysis (including of GO, KEGG and Biocarta genesets) and visualisation of the aligned datasets was performed using pipeline_scxl.py (https://github.com/sansomlab/tenx). Neighbor graphs were built using Euclidean distance and the “HNSW” algorithm (as implemented in ScVelo (Bergen et al., 2020), following the example of Pegasus (Li et al., 2020)). Clustering was performed using Scanpy, marker discovery with Seurat (Stuart et al., 2019) and pathway over-representation analysis with gsfisher (Croft et al., 2019) (https://github.com/sansomlab/gsfisher).

After alignment of the full manifold, we iteratively divided, re-aligned and re-clustered the dataset to achieve high-resolution clustering of different cell subsets. Variable gene identification was performed separately within each subset as described above. In the first step the major cell types – T/NK, myeloid and B/plasmablast cells were extracted into n=3 separate subsets. Each of these three subsets was then separately aligned and clustered (n=45 PCs for n=482k T/NK cells; n=45 PCs for n=279k myeloid cells; n=40 PCs for n=66k B/plasmablast cells). This process was then repeated with a final (third) round of alignment and clustering being performed separately on six subsets: (A) cells in the CD4 region of the manifold (with n=50PCs, n=302k cells), (B) cells in the CD8 region of the manifold (with n=45PCs, n=180k cells), (C) cells in the myeloid region of the manifold (with n=50PCs, n=261k cells), (D) cells in the B cell/Plasmablast cell region of the manifold (with n=40PCs, n=57k cells), (E) cells identified as doublets based on scrublet scores and marker gene expression (with n=40PCs, n=26k cells), (F) other cells types not falling into any of the first five categories (with n=50PCs, n=10k cells). The clustering strategy is shown in **Methods S2 Figure 3**. Choice of principle component number was guided by inspection of the scree plots.

Each of the cell six subsets was subject to Leiden clustering with a range of resolutions (1, 1.5, 2, 2.5, 3, 3.5, 4, 4.5, 6, 8) and cluster phenotypes assessed using pipeline_scxl.py (https://github.com/sansomlab/tenx) as described above, and ADT surface phenotype. For each subset a base resolution was selected and sub-populations further refined by manual splitting or merging of clusters based on alternate cluster resolutions guided by expert knowledge of cell identities and phenotypes to define a total of n=131 GEX clusters. These clusters were intersected with information from the ADT and repertoire modalities for the final multimodal annotation of the dataset as described below.

#### Pre-processing and analysis of the ADT data

For each channel the ADT signal was normalized independently to its background, using the DSB algorithm (Mulè et al., 2020) taking the set of cells identified from the GEX data as the foreground and unassigned GEMs with log10(GEX_nUMI +1) >=1.5 as the background. The workflow was written in R and Python, encapsulated in CGAT pipelines and version controlled. The normalised ADT data was subject to hierarchical stochastic neighbour embedding (hSNE) and unsupervised Gaussian mean shift (GMS) clustering as implemented in Cytosplore (van Unen et al., 2017) (www.cytosplore.org). This analysis, which was based on the expression of all n=192 ADT tags, partitioned the dataset into 66 discrete clusters. Based on expression of known lineage markers (CD3, CD4, CD8, CD56, CD19, CD20, CD27, CD38, CD14, CD123, CD1c, CD33, CD235, CD34, Vd2, Va72 and CD161) these clusters were apportioned to NK cells, B cells, Plasmablasts, Vd2+ T cells, CD14+ Monocytes, CD123+PDC, CD1c+ DC, red blood cells (RBC), CD34+ cells and distinct populations of CD3+ T cells (**Methods S2 Figure 4**). For identification of Va25+Ja18+ cells (iNKT cells), Vd2-Vg9+ cells, TCRgd+ cells, Va7.2+CD161+ (MAIT) cells and non MAIT T cells a sequential manual gating strategy was employed. After exclusion of likely contaminant CD33+ CD3+ and CD56+ cells subpopulations of B cells and plasmablasts were defined by re-clustering these cells based upon expression of CD10, CD24, CD25, CD19, CD20, CD22, HLADR, CD21, CD23, IgD, CD1c, IgM, CD38, CD29, CD71, CD39, CD27, HLA-ABC and IgA. In total, this analysis of the ADT data identified 12 T-cell subpopulations, 7 B cell/plasmablast subpopulations and five distinct sets of doublets (with markers of more than one major immune lineage). These clusters were intersected with information from the GEX and repertoire modalities for the final multimodal annotation of the dataset as described below.

#### Pre-processing and analysis of 10x V(D)J B and T cell repertoire data

T and B cell V(D)J gene usage and receptor sequences were quantified using Cellranger VDJ (v3.1.0) with reference sequences from the IMGT. For TCR chain usage we recorded additional information for each cell including (i) “clone proportion”, computed as the proportion of the entire T cell repertoire occupied by the clone in the sample, (ii) whether the cell was a doublet (defined as a singlet clone with TRA-TRA-TRB-TRB chains), (iii) whether the cell possessed an iNKT receptor phenotype (TRAV10 with TRAJ18), and (iv) whether the cell carried a MAIT receptor phenotype (TRAV1-2, with either TRAJ12/TRAJ20/TRAJ33). Immunoglobulin sequences were further analysed using IMGT High V-Quest. B cell/plasmablast doublets were identified as cells with multiple heavy chain (HC) contigs where the ratio of the number of UMIs for the top two ranked HC contigs was >0.125. Light chain (LC) information was not used for doublet identification as it is known that 1% of B cells may express dual light chains. The TCR and immunoglobulin chain usage information was intersected with information from the GEX and ADT modalities for the final annotation of the dataset as described below.

#### Multimodal cluster annotation

As described below, we performed curated intersections of the information from the different modalities to discern the cellular identities and functional phenotypes of the different PBMC populations. Information from all three modalities was used to screen and exclude doublets: in addition to inspection of scrublet scores (computed from the GEX layer), lineage according to surface phenotype, immune receptor sequence and gene expression profile was required to be congruent. In total, following curated intersection of GEX, ADT and repertoire information we obtained n=853 clusters. Of these we retained n=346 clusters (n=787,928 cells) for downstream analysis. We excluded n=92 clusters (n=20391) as likely cell-cell doublets/multiplets and n=270 clusters (n=19,801 cells) which appeared to be comprised of mixtures of cell types. A further n=128 clusters (n=7971 cells) of uncertain phenotype were also excluded. The retained cells comprised of n=343 clusters that were assessed to represent genuine singlet cell clusters based on expert knowledge. We assigned each of these clusters to a “minor subset”, “major subset” and “cell type” category in order to accommodate downstream analyses at different levels of granularity as summarised in **Methods S2 Figure 5**. In addition, we retained two clusters that appeared to comprise monocyte-platelet doublets (n=5562 cells) and a minor, lower confidence cluster of AXL+ DCs (n=121 cells) for inclusion in cluster-level analyses. Scripts were written in Python and R and version controlled.

Known T and NK cell subsets were identified by surface protein (ADT) and immune receptor (TCR) phenotype as recorded in **Methods S2 Table 1**. These identity assignments were then intersected with the gene expression clusters (GEX) to identify subsets of the different lineages that displayed distinct transcriptional phenotypes (e.g. naïve vs memory T cells). These sub-populations are shown for CD4 T-cells in **Methods S2 Figure 6**, for CD8 T-cells in **Methods S2 Figure 7**, for double positive (DP) T-cells in **Methods S2 Figure 8**, for double negative (DN) T-cells in **Methods S2 Figure 9**, for mucosal associated invariant T (MAIT) cells in **Methods S2 Figure 10**, for Vδ2+ gamma-delta (γδ) T cells in **Methods S2 Figure 11**, Vδ2-ve Vγ9-ve γδ T-cells in **Methods S2 Figure 12**, Vδ2-ve Vγ9+ve γδ T-cells in **Methods S2 Figure 13** and invariant natural killer T cells (iNKT) in **Methods S2 Figure 14**. The identified NK cell subsets are shown in **Methods S2 Figure 15**.

B cell and plasmablasts cell subsets were defined using information from the GEX, ADT and immunoglobin repertoire modalities as detailed in **Methods S2 Table 2** and shown in **Methods S2 Figure 16**. Sub populations of innate immune cells and other blood cells were delineated according to gene expression clusters and annotated based on their GEX and ADT phenotypes. Sub-populations of mononuclear phagocytes (MNPs) are shown in **Methods S2 Figure 17**, of haematopoietic stem (and progenitor) cells (HSCs) in **Methods S2 Figure 18** and of platelets and erythrocytes in **Methods S2 Figure 19**.

To assess the relative immune receptor diversity of the different T, B and plasmablast populations we computed repertoire Shannon and Gini indices (detailed in the ***Repertoire analysis*** section, and as shown in **Methods S2 Figures 6-14 and 16)**. Because of the large variance in cell number between clusters we estimated the indices by bootstrapping with a sample size of n=30, n=1000 times. Individual bootstrap samples were drawn without replacement to avoid introduction of artificial clonality. The indices were not estimated for clusters with fewer than n=30 cells.

#### CITE-seq: cell composition analysis

##### Overview

Three samples with <500 cells in total, and three samples with confirmed or suspected malignancy, were excluded from all composition analyses. For the hospitalised COVID-19 and sepsis clinical categories, only samples closest to maximum severity and only one sample per individual were included, such that for each category the following numbers of samples were analysed: healthy volunteers, *n*=10; COVID-19 acute in-patient mild (OUH), *n*=12; COVID-19 acute in-patient severe (OUH), *n*=20; COVID-19 acute in-patient critical (OUH), *n*=18; COVID-19 community COVID-19, *n*=12; influenza, acute in-patient, *n*=10; and sepsis acute in-patient, *n*=15. Scripts for all analysis were written in Python and R and version controlled.

Composition analysis was performed for the different levels of cellular granularity summarised in **Methods S2 Figure 5**, including for cell types (**Methods S2 Figures 20-22**); major cell subsets (**Methods S2 Figures 23-25**); minor cell subsets (**Methods S2 Figure 26-28**); T and natural killer (NK) cell clusters (**Methods S2 Figures 29-31**); B and plasmablast (PB) cell clusters (**Methods S2 Figure 33-34**); and mononuclear phagocyte (MNPs) clusters (**Methods S2 Figure 35-37**). To initially inspect cluster abundance across clinical categories, the percentage frequencies of cell subpopulations were quantified per sample and visualized as boxplots. For cell types (**Methods S2 Figure 20**), major cell subsets (**Methods S2 Figure 23**) and minor cell subsets (**Methods S2 Figure 26**), percentages were calculated out of total PBMCs. For higher resolution immune cell clusters, frequencies were calculated out of total T and NK cells (**Methods S2 Figure 29**), total B and PB cells (**Methods S2 Figure 32**), or total MNPs (**Methods S2 Figure 35**). Selected panels are reproduced in the main and supplementary figures.

#### Principal component analysis

Principal component analysis (PCA) was used for exploratory analysis, with pre-filtering to remove clusters with a count of *n*<10 cells in <10 individuals. Cluster counts were converted to centred log-ratios (CLRs) using the ALDEx2 R package, v1.18.0. Briefly, 1,000 Monte Carlo samples of the Dirichlet distribution were generated from the cluster counts for each sample and converted to CLRs. The median CLR for each cluster-sample combination was used for PCA which was performed using prcomp (R version 3.6.2) with default parameters. Association tests for clinical variables were performed using an omnibus analysis of variance (ANOVA) to test for association between the top 15 PCs (collectively explaining >80% of the total variance) and clinical category (source), age, sex, and sample pool, with the Benjamini-Hochberg procedure for multiple testing correction and a significance threshold of *P_c_*<0.05. For the hospitalised COVID-19 cases, association tests with additional variables that were related to acute disease or were secondary to infection were also performed. All variables had one or no missing values. These additional variables included: ethnicity; weight; days from symptom to sample and from symptom to admission; maximum temperature in the 24 hours preceding sampling; persistent fever in the 24 hours preceding sampling; fever, WHO ordinal, ventilation status, SaO_2_/FiO_2_ ratio, and SOFA oxygenation score at the time of sampling; quantile normalised white cell, neutrophil, lymphocyte, monocyte and platelet counts in the 24 hours before or after sampling; quantile normalised highest concentration of C-reactive protein in the 24 hours before or after sampling; clinical and/or radiological evidence of thromboembolism during hospitalisation; length of hospital stay; OpenSAFELY COVID-19 mortality propensity score; and death in the hospital.

The full results of the PCA and association test results are shown in **Methods S2 Figure 21** for the cell types; **Methods S2 Figure 24** for the major cell subsets; **Methods S2 Figure 27** for the minor cell subsets; **Methods S2 Figure 30** for the T and NK clusters; **Methods S2 Figure 33** for the B and PB clusters; and **Methods S2 Figure 36** for the MNP clusters with selected panels reproduced in the main and supplementary figures.

#### Differential abundance analysis

To compare cell subset/cluster abundance across clinical categories, the percentage frequencies of cells were quantified per sample and visualized as boxplots. For major and minor cell subsets, percentages were calculated out of total PBMCs. For higher resolution immune cell clusters, frequencies were calculated out of total myeloid cells, total B and plasmablast cells, or total T and natural killer (NK) cells. To determine the statistical significance of differences in cell subset/cluster abundance between groups, differential abundance analysis was performed using edgeR, v3.28.1 (Amezquita et al., 2020). Cell subset/cluster counts were modelled adjusting for age, sex and sample pool using a quasi-likelihood negative binomial generalized log-linear model (glmQLFit function), with pre-filtering to remove clusters with a count of *n*<10 cells in <10 individuals. Counts were normalised by the total number of cells in each sample for the major and minor cell subset analyses, or by the total myeloid, the total B and plasmablast, or the total T and NK cells for higher resolution immune cell cluster analyses. Differential abundance testing was performed using the quasi-likelihood F-test. The Benjamini-Hochberg procedure was implemented to correct for the number of clusters and the number of pairwise clinical category comparisons, and a significance threshold of *P_c_*<0.05 was used. Testing for composition effects did not provide evidence for biases in cluster abundance. In addition, for the hospitalised COVID-19 patients, edgeR analysis was performed using ANOVA to identify statistically significant associations between cluster abundance and patient characteristics/clinical variables (as detailed for the PCA association tests).

The full results of the differential abundance and covariate analysis results are shown in **Methods S2 Figure 22** for the cell types; **Methods S2 Figure 25** for the major cell subsets; **Methods S2 Figure 28** for the minor cell subsets; **Methods S2 Figure 31** for the T and NK clusters; **Methods S2 Figure 34** for the B and PB clusters; and **Methods S2 Figure 37** for the MNP clusters.

#### CITE-seq: GEX PCA, differential expression and pathway analysis

##### Data pre-processing

Pseudobulk counts were generated for each combination of gene and sample at minor subset, major subset and cell type level by summing together the within-group gene counts. These were then converted to reads-per-million (RPM) by normalizing by the total count across all genes for each combination of sample and cell type. Finally, residuals were calculated by taking the log(1 + count) and subtracting the predicted value from a linear model with pool as the independent variable. The six poorly performing samples mentioned in the Composition section above were removed from all further processing.

##### PCA analysis

PCA was carried out on the residuals using the prcomp function in R. We generated PCs separately at minor subset, major subset and cell type level using residual values from genes with a mean RPM > 1 in among all sources, and with six poorly performing samples removed. Outlying samples (more than 5 SDs away from the mean across the top 10 PCs) were then removed and PCs recalculated. Plots of the top two principal components for each major subset is shown in **Methods S2 Figure 38**. To test differences in major subset PCs across categories, we used an omnibus ANOVA (i.e. a test of a linear model including all diagnoses/severity categories as dummy variables, against a model with no difference between these groups) to test for association between the first 10 PCs and diagnosis/severity (across all samples) and disease severity (measured by WHO criteria, within COVID-19 samples) (**Methods S2 Figure 39**). The linear model also includes terms for age and sex. Omnibus p-values were corrected for multiple testing using the Benjamini-Hochberg procedure. We also repeated the principal component analysis only within hospitalized COVID-19 samples and tested their association with clinical covariates. Only two major subsets showed adjusted p-values < 0.01, classical monocytes and DCs, with association patterns shown in **Methods S2 Figure 40**.

We generated UMAPs from the top n=10 PCs for each level of cell clustering (using the R package “umap”). For this analysis we excluded clusters with n=0 cells for ≥10 samples and excluded samples with n=0 cells in remaining clusters. The resulting plots for minor subsets, major subsets and cell types are shown in **Methods S2 Figure 41**.

##### Differential Expression Analysis

We carried out differential expression tests for a range of contrasts (including subgroups of COVID-19 against healthy volunteers, sepsis and influenza vs healthy volunteers, COVID-19 vs sepsis, COVID-19 vs influenza and COVID-19 subgroups against one another. These tests were performed using the R edgeR library (McCarthy et al., 2012). We included age, sex and pool as covariates, and filtered out samples with <5 cells and gene with mean RPM < 1, with significant genes selected based on log fold change (logFC) > 2 and false discovery rate (FDR) < 0.01 (calculated by Benjamini-Hochberg). To test for a set of genes that differed across COVID-19 severity categories, we also carried out a “COVID-19 omnibus” likelihood ratio test, with dummy variables for WHO severity and a separate variable for mild recording healthcare workers (in this case, genes were selected if they had logFC > 2 between any pair of categories).

The number of differentially expressed genes (FDR < 0.01, absolute log-fold change > 2) in each cell cluster for each contrast is shown in **Methods S2 Figure 42**, volcano plots of differentially expressed genes for critical COVID-19 patients vs healthy volunteers for each major subset are shown in **Methods S2 Figure 43**, and the top 10 most differentially expressed genes for each contrast (across all cell clusters) are shown in **Methods S2 Table 3**. We also investigated the relationship between the frequency of a cell subset and the number of differentially expressed genes (**Methods S2 Figure 44**), and the relationship between differential composition and number of differentially expressed (**Methods S2 Figure 45**).

##### Pathway Analysis

Gene-set enrichment analysis GSEA of differentially expressed genes were performed using the FGSEA algorithm (Korotkevich et al., 2021). We performed GSEA for biological pathways from the MSigDB database (include for KEGG, GO, canonical pathways, regulatory sets, immune signatures and Hallmark genesets). Enrichment analysis was carried out separately for each pair of cell cluster and contrast, with genes ranked by p-value. For the Hallmark genesets, we also carried out a signed enrichment test (ranking by log(pvalue) x sign(logFC)), shown as a heatmap for major subsets and a range of contrasts in **Methods S2 Figure 46.** The top 10 pairs of cell cluster and GO term, canonical pathway and immunologic signature for critical COVID-19 vs healthy volunteers are shown in **Methods S2 Table 4**.

In addition, we applied GSEA to a set of experimentally derived interferon-stimulated genes (ISGs) (Schoggins et al., 2011). Genes within this gene set were differentially expressed across a wide range of minor subsets and contrasts (**Methods S2 Figure 47**), with the most significant enrichment being seen in HSCs in critical COVID-19 patients (**Methods S2 Figure 48**). We focused in on classical dendritic cells, as this minor subset showed enrichment of differential expression in ISGs (p < 1e-5) in 9 out of 13 contrasts tested. **Methods S2 Figure 49** shows the mean expression in DCs across the different disease groups of a subset of ISGs that were identified both as driving the ISG enrichment signature (leading edge genes) and showing individually strong differential expression (FDR < 0.001, absolute fold change > 3).

#### CITE-seq: WGCNA analysis

We performed separate WGCNA (Langfelder and Horvath, 2008) analyses of n=7 selected “major cell types” consisting of cell populations that were annotated as “cell types” (B cells, Plasmablasts, NK cells) or “major subsets” (cMono, ncMono, CD4 T cells, CD8 T cells). For this analysis per-patient pseudobulk-summarized RPM-normalized counts from the prioritised sample set were used as input. Patients with low total cell numbers (<500 cells; n=3), with confirmed or suspected malignancies (n=3), or from the London COVID-19 cohort (n=2) were excluded from the analysis.

Input gene expression matrices were filtered to retain genes with expression above a certain threshold in a given minimum number of samples. Based on inspection of expression distributions RPM thresholds of 1 (cMono, NK cells, CD4 T cells and CD8 T) and 3 (ncMono, PB and B cells) were chosen for the indicated “major cell types”. The number samples that constituted the smallest patient group was used as the minimum number of samples. After filtering we retained 9000-12000 genes per “major cell type”. Log2 RPM+1 values were batch corrected using the ComBat algorithm (sva R package v 3.36.0) (Johnson et al., 2007; Leek et al., 2012) specifying the multiplexing sample pool as the adjustment variable (together with an intercept term).

The WGCNA analysis was performed using pipeline_wgcna.py from the https://github.com/sansomlab/cornet repository, and geneset over-representation analysis was carried out using the gsfisher R library https://github.com/sansomlab/gsfisher. The parameters used for WGCNA runs are given in **Methods S2 Table 5**. We excluded 3 “ambient_rna” modules whose eigengene gene loadings correlated strongly with ambient RNA species abundance (as quantitated in “empty” droplets) (Spearman’s. rho ≥0.83, p < 2.2 x 10^-16^; data not shown) and unassigned grey modules from downstream analysis.

Modules were characterised and named by inspection of their gene membership, over-representation of biological pathways (GP Biological Process, Go Cellular Component, KEGG, MsigDB REACTOME and MSigDB transcription factor motifs) and correlation with AUCell (Aibar et al., 2017) (v 1.12.0) expression scores for specific sets of genes. The “curated type I IFN response” geneset comprised of GBP2, IFI27, IFI44L, IFIT1, IFITM1, IFITM3, IFNA1, IFNA10, IFNA13, IFNA14, IFNA16, IFNA17, IFNA2, IFNA21, IFNA4, IFNA5, IFNA6, IFNA7, IFNA8, IFNB1, IRF1, IRF3, IRF7, IRF9, ISG15, MX1 MX2, OASL, RSAD2, SIGLEC1, TRIM56, USP18 and XAF1.The “curated AP1 TF family” geneset comprised of the genes shown in **Figure 2M**. The custom “zf-C2H2 (PF00096) domain” geneset comprised of genes containing the PF00096 domain (Pfam database). For geneset over-representation analysis only genesets with a BH corrected p-value < 0.05 are reported (separate corrections performed within major cell type and ontology).

Inter-module eigengene expression correlation (**Figure S2O**) was computed for the subset of n=77 COVID-19 hospitalized patients that commonly passed filters for inclusion in the analyses for all “major cell types”. Module eigengene correlations with disease were computed by numerically coding the given x_vs_y disease groups as 1 and 0 respectively (**Figure 2K**). Module eigengene associations with variance between patient groups were assessed by ANOVA (COVID+HV_ANOVA, All_Groups_ANOVOA, **Figure 2K**). Module eigengene correlations with clinical variables were computed using pairwise-complete observations. Where appropriate clinical variables were quantile normalised as described for analysis of clinical phenotype, as described below. Stars in **Figure 2K** indicate a significant association between the contrast, clinical variable or geneset score and the module eigengene (BH corrected p-value < 0.05, p-value correction performed separately for each contrast, variable or score).

#### Repertoire analysis

##### Strategic approach

Here, we used both single cell (sc) and bulk cell VDJ sequencing to decipher the immune responses in COVID-19 patients. The benefits of scVDJ-seq is the joint analysis of the VDJ BCR or TCR sequence along with the gene expression and CITE-seq for each individual cell, allowing for a detailed linking of antigen receptor function or type with cellular phenotype or function. However, this is limited by the number of cells that can be captured in a single experiment, usually between a few 10’s to 1000’s. Therefore, bulk VDJ-seq can also be used to capture a more holistic view of the immune cell repertoire. While this does not capture VDJ sequences on a single cell level, many aspects of the repertoire may be investigated, such as changes in clonality, dynamics, selection and tolerance. Because bulk VDJ-seq is able to capture the immune receptors of 1000’s to 100,000’s of cells, this means that the resulting repertoire is more representative of the individual that single cell VDJ-seq, and has greater power to detect low frequency clones or repertoire features. Finally, through capturing a higher magnitude of BCR/TCR sequencing, imputing the germline IGHV/J germline alleles becomes possible, and therefore allowing for analyses of differential genetics between groups.

##### Additional pre-processing of scBCR-seq data for repertoire analysis

The BCR outputs from CellRanger were run through IMGT V-QUEST (Giudicelli et al., 2004) to determine the TCR chain types and annotation. In addition, only BCRs from droplets that were confidently annotated as (a) singlets, (b) within the B cell annotated clusters in the GEX data, and (c) were confidently genetically demultiplexed were included. Processed FASTA sequences, corresponding to high confidence VDJ contigs from 10X Genomic’s Cell Ranger v4.0.0 pipeline, were annotated using IMGT HIGHV-QUEST. To ensure high quality contigs, non-productive rearranged sequences were removed and only sequences corresponding to cell barcodes that past QC cut-offs for gene expression data were analysed. For B cell VDJ sequences where multi-chains were detected, e.g. two or more heavy or light chain sequences, only contigs ranked above the first derivative of log10 ranked contig ratios were selected for downstream analysis. Contig ratios were defined by:

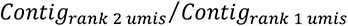

Where multi-chains passed this quality threshold, the contig corresponding to the lowest UMIs detected was considered ambient RNA contamination and removed. Where multi-chains did not pass this threshold, we considered these to be low confidence of being true single cell droplets and likely to represent homotypic doublets; thus, they were removed from further analysis.

Cells with filtered BCRs were concentrated in the regions annotated as B cells or plasmablasts/plasma cells through gene expression analysis (**Figure S4E**). Plasmablasts/plasma cells exhibited higher BCR IGH/ IGK/L expression compared to non-plasmablasts/plasma cell populations.

##### Additional pre-processing of scTCR-seq data for repertoire analysis

The TCR outputs from CellRanger were filtered based on called productivity, and chain identity (TRA or TRB). Only T cells that contained either (a) 1 beta chain, (b) 1 alpha and 1 beta, (c) 2 alpha chains and 1 beta were retained. Given that bona fide T cells with UMI counts of 1 have been validated in previous datasets (data not shown), the minimum number of UMIs required per cell to accept was 1. In addition, only TCRs from droplets that were confidently annotated as (a) singlets, (b) within the T cell annotated clusters in the GEX data, and (c) were confidently genetically demultiplexed were included.

The IMGT annotation of cell types demonstrated that the majority of droplets for which TCRs were captured were able to pass the chain type filters (**Methods S2 Table 1**). After removal of low-quality droplets, there remain 94 samples corresponding to the maximal disease severity per patient. Four samples were excluded due to low T cell capture (<200 cells). Age was used as a co-variate in downstream analyses.

##### scBCR clonality measurements

Clonal assignment was performed using a custom procedure. First, concatenated VH/VJ nucleotide sequences were clustered using an adapted R implementation of CD-HIT (https://github.com/thomasp85/FindMyFriends). Subsequently, within-cluster CDR3 normalised levenshtein distances were generated using the “stringdist” R package. The clonal threshold was set as the local minimum of the CDR3 distance distribution. Convergent clones were assigned using the same procedure without constraints accounting for biological replicates. The presence of published SARS-CoV-2 binding antibody sequences from CoV-AbDab (Raybould et al., 2020) within the dataset was identified by first requiring identical VH and VJ gene pairings, followed by CDR3 distance thresholding (as described above).

The clonal expansion index (CEI) was calculated as the Gini index (unevenness) of the number of total BCRs per clone. The clonal diversification index (CDI) was calculated as the GINI index of the number of unique BCRs per clone. This is a measure of unevenness based on how many non-identical members of each clone are diversified from their inferred germline ancestor.

##### scTCR clonality measurements

A clone ID was defined by a concatenation of amino acid CDR3 chains present, with cells sharing identical clone IDs classed as members of the same clonotype. Clonal proportions were calculated by dividing the number of cells in each clone per sample by the total of number of cells per sample. Shannon diversity was calculated from count data of clones per sample using the R package entropy (Hausser, 2009). Statistical analysis was performed using a linear model with covariates for age and sample size.

For the scTCR analysis the cluster annotations were merged to create simplified clusters for analysis. The following reannotated clusters were used in TCR clonality analysis, with constituent “minor subsets” shown in brackets: CD8 T effector memory (CD8.TEMRA + CD8.TEM); CD8 T central memory (CD8.TCM + CD8.TCM.CCL5); CD8 T effector (CD8.TEFF + CD8.TEFF.prolif); CD8 naïve (CD8.NAIVE); CD4 T effector (CD4.TEFF + CD4.TEFF.prolif); MAIT (MAIT). Where comparisons of clone size across samples has been performed in terms of absolute number of member cells (e.g. **Figure 5J, S5F,G**) each sample was randomly down-sampled without replacement (n=1250 for CD4, n=250 for CD8) to generate repertoires of identical size across all samples. The mean size of clone was then calculated from the down-sampled repertoires. This process was bootstrapped 100 times and the mean size from these iterations is presented. This approach allows for accurate comparisons between samples containing different cell numbers and controls for sample size.

##### scTCR cytotoxicity and kmer analysis

Cytotoxicity score was calculated using the AddModuleScore function in Seurat (Butler et al., 2018) with a gene-set of the top 50 genes that significantly correlated with IFNG expression in an independent single cell dataset (Watson et al., 2020b) and were identified as variable features in Seurat. The gene-set was then used as an input to generate phenotype scores.

CDR3b sequences of T cells identified as CD4+ or CD8+ T cells were broken down into 4-amino acid length sequences (kmer). A Fisher’s exact test was performed per kmer across each group of patients (HV, COVID-19 or sepsis patients) and p values adjusted using a Bonferroni’s correction, with a leave-one-out re-sampling of individuals employed to ensure inter-individual reliability. Kmer sequences significantly enriched in COVID-19 patients over HV or sepsis patients across more than 95% of the re-samples were identified as COVID-enriched kmers. The proportion and cellular phenotype (i.e. subset and cytotoxicity) of T cells with a CDR3b sequence containing a COVID-enriched kmer was subsequently compared between COMBAT clinical groups.

##### Identification of SARS-CoV2 specific T cell clones using public CDR3 databases

Putative viral antigen specificity of clonotypes within COMBAT cohort was determined using publicly available databases of viral antigen specific CDR3 sequences. At the time, two databases containing TCR sequences with reported binding to SARS-CoV-2 epitopes were available: VDJdb (Bagaev et al., 2019) and ImmuneCODE™ (Nolan et al., 2020). From VDJdb, all unique CDR3 amino acid sequences (alpha as well as beta chain of human TCR) of all lengths and MHC-restrictions with reported binding to known SARS-CoV2 associated antigens were selected. Sequences with low confidence scores assigned to TCR:peptide:MHC complexes or missing sequencing/ specificity validation data were then filtered out. ImmuneCODE™ database includes deeply sampled TCRb repertoires from over 1,400 subjects exposed to or infected with the SARS-CoV-2 virus. Within these, over 135,000 TCRs were deemed to be SARS-CoV-2-specific with high-confidence using Multiplex Identification of Antigen-Specific T-Cell Receptors Assay (MIRA) and were included(Klinger et al., 2015). Unique CDR3 amino acid sequences thus identified were collated and interrogated against TCR sequences within COMBAT cohort for matches within CDR3a, CDR3a2 and CDR3b regions. A similar strategy was used to identify CMV, EBV and Influenza specific clones. Finally, proportions of individual repertoires occupied by viral clones were compared between COMBAT clinical groups.

##### Bulk BCR and TCR quality control and filtering

Raw sequencing reads were filtered for base quality (median Phred score >32) using QUASR (Watson et al., 2013). Forward and reverse reads were merged if they contained identical overlapping region of >50bp, or otherwise discarded. Universal barcoded regions were identified in reads and orientated to read from V-primer to constant region primer. The barcoded region within each primer was identified and checked for conserved bases. Primers and constant regions were trimmed from each sequence, and sequences were retained only if there was >80% per base sequence similarity between all sequences obtained with the same barcode, otherwise discarded. The constant region allele with highest sequence similarity was identified by 10-mer matching to the reference constant region genes from the IMGT database (Lefranc, 2011), and sequences were trimmed to give only the region of the sequence corresponding to the variable (VDJ) regions. Isotype usage information for each BCR was retained throughout the analysis hereafter. Sequences without complete reading frames and non-immunoglobulin/TCR sequences were removed and only reads with significant similarity to reference IGHTCR V and J genes from the IMGT database using BLAST (Altschul et al., 1990) were retained. Ig/TCR gene usages and sequence annotation were performed in IMGT V-QUEST, where repertoire differences were performed by custom scripts in Python.

Bulk BCR sequencing resulted in 2,356,813 BCRs consisting of 1,905,867 unique BCR sequences, resulting in 79 samples passing all QC measures and with the minimum number of BCR sequences per sample at 1000. Bulk TCR sequencing resulted in 1,200,656 TCRs consisting of 1,159,363 unique TCR sequences, resulting in 77 samples passing all QC measures and with the minimum number of TCR sequences per sample at 1000.

##### Isotype frequencies, somatic hypermutation, CDR3 lengths and IGHV gene usages

Analysis methods are based on (Bashford-Rogers et al., 2019). To account for the greater numbers of BCR RNA molecules per plasmablast compared to other B cell subsets, the normalised isotype usages, defined as the percentage unique VDJ sequences per isotype, thus controlling for differential RNA per cell and reducing potential biases from differential RNA per cell. Similarly, mean somatic hypermutation levels and CDR3 lengths were calculated per unique VDJ region sequence to reduce potential biases from differential RNA per cell. IGHV gene usages were determined using IMGT, and proportions were calculated per unique VDJ region sequence. The representation of IGHV genes in the BCR repertoire reflects their presence in the germline, the naïve repertoire and their expansion after antigenic exposure. We therefore compared the frequency of IGHV gene use in PBMC-derived BCRs identified by sequence as being enriched for naive (IgM+D+SHM-: >78% naïve B cells by flow cytometry) and antigen-experienced B cells (including both unswitched (IgM+D+SHM+) and class-switched memory (IgA+/G+/E+) subsets) as shown in (Bashford-Rogers et al., 2019).

##### Bulk BCR class-switching event analyses

Relative class-switch event frequency was the frequency of unique VDJ regions expressed as two isotypes (i.e. from more than one B cell, where one has undergone class-switch recombination). This was determined as proportion of unique BCRs present as both isotypes IgX and IgY within a random subsample of 8000 BCRs, where the mean of 1000 repeats was generated. This provides information on the frequency of BCRs observed associated with any two isotypes (class-switching events) while accounting for total read depth, but not accounting for differences in the relative frequencies of BCRs per isotype.

The per-isotype normalized class-switch event frequencies determines frequency of unique VDJ regions expressed as two isotypes whilst normalizing for differences in isotype frequencies. To account for differences in isotype proportions, BCRs from each isotype were randomly subsampled to a fixed depth of 100 BCRs, and the proportion of unique VDJ sequences present between each pair of isotypes was counted. The mean of 1000 repeats was generated.

##### RNA-velocity B cell analysis

The single-cell RNA-seq data sets were subjected to the standard RNA-velocity pipeline, and the trailing analyses were performed using the scVelo package (v0.1.24). The scVelo package was used to normalize the counts and select highly variable genes based on spliced counts. Following this, the dynamical model implemented in scVelo was used to estimate the RNA velocity for the cells. The estimated velocities were then visualized using Partition-based graph abstraction (PAGA) plots using the previously computed UMAP embeddings.

#### scATACseq data analysis

Raw data pre-processing was performed with Cell Ranger ATAC (10X Genomics). ‘cellranger-atac count’ pipeline was used to align reads and generate single-cell accessibility counts for the cells. The reference genome assembly file was Ensembl GRCh38 v100 Primary Assembly corresponding to hg38, downloaded from http://ftp.ensembl.org/pub/release-100/fasta/homo_sapiens/dna/Homo_sapiens.GRCh38.dna.primary_assembly.fa.gz. This file was used as reference genome file for alignment and generation of single-cell accessibility counts. Annotations were from Ensembl GRCh38 v100 Gene Annotation downloaded from http://ftp.ensembl.org/pub/release-100/gtf/homo_sapiens/Homo_sapiens.GRCh38.100.gtf.gz.

VIREO (Huang et al., 2019) and demuxlet (Kang et al., 2018) packages were used to demultiplex patient samples within a channel and identify inter-sample nuclei doublets. Concordantly filtered barcodes from individualised samples were then used to create individual fragment files for each patient using HTSlib –c function. Following the creation of fragment files, downstream analysis of the scATACseq data was performed using the ArchR v0.9.3 R package (Granja et al., 2021).

Fragment files were first checked using the reformatFragmentFiles function. Arrow files were then generated by reading each sample’s specific fragment files and a tile matrix was created using 500-bp bins. Cells with a transcription start site enrichment score < 4, with fewer than 1000 detected fragments or containing intra-sample nuclei doublets were removed, resulting in ∼46000 cells, which were subjected to dimensionality reduction with iterative Latent Semantic Indexing (LSI) and Singular Value Decomposition (SVD), followed by Uniform Manifold Approximation and Projection (UMAP) embedding calculation to visualize the data structure in the two-dimensions. ∼4000 cells were manually removed at this stage, as we identified putative batch effects. The resulting ∼42000 cells were subjected to UMAP and were clustered using the implementation from Seurat R package (Stuart et al., 2019). Cluster-specific gene activity scores were identified based on the local chromatin state, and marker genes were identified (FDR ≤ 0.01 & Log2FC ≥ 1.25). Unconstrained integration with cognate scRNA-seq profiles was performed using the addGeneIntegrationMatrix (ArchR) method and scRNA-seq cell type annotations were used to label scATAC-seq clusters. We performed peak calling using MACS2 with the addReproduciblePeakSet (ArchR) function using pseudo-bulk replicates. Such replicates were grouped with different variables, such as cell-type, condition and patient, as well as a combination of these variables. Differentially accessible peaks (FDR <= 0.1 & Log2FC >= 0.5) were identified between pairwise comparisons, and peak-to-gene linkages were calculated using the addPeak2GeneLinks (ArchR) method using a correlation cut-off of 0.45 and resolution = 1000. We then used the ‘cisbp’ motif set to annotate motifs in accessible peaks using the addMotifAnnotations (ArchR) function. Motif enrichments in differentially accessible peaks were calculated using the peakAnnoEnrichment (ArchR) method. Finally, motif footprinting was performed by measuring Tn5 insertions in genome-wide motifs and normalized by subtracting the Tn5 bias from the footprinting signal.

#### Luminex data analysis

##### Cytokine enrichment profile analysis

The concentrations of 51 circulating proteins in plasma were presented as mean FI ± SEM and compared between HV and each disease severity group of COVID-19 using unpaired student’s t-test, and then depicted with Prism.

##### Principal Component Analysis (PCA)

PCA on 171 plasma samples was conducted with the Scater package in R Programme. The values of FI were normalised with logNormCounts function, and then calculated with the RunPCA function. The loadings were generated by the value of first two components of eigenvectors multiplied by 10.

##### Heatmap

The heatmap was coloured by the log10 of the fold-change in the natural log of the fluorescence intensity (FI), normalised against the mean value of HV for the plasma cohort and HS for the serum cohort. When comparing mortality in severe and critical COVID-19 patients, the data is normalised to the survivor group. The colour-scale is bounded at ±5 fold change (0.7 in log10), with an increased FI shaded red; decreased FI shaded green; unchanged FI shaded yellow.

##### Volcano plots

The p-values in the Volcano plots are calculated using a two-tailed two-sample unpaired T-test (ttest2, MATLAB). The T-test was taken for the natural log of the FI of the test and control conditions. The p-values are plotted against the log2 of the fold-change in the natural log of the fluorescence intensity (FI) between the test and control conditions.

##### Uniform manifold approximation (UMAP)

The UMAPs [L. McInnes, J. Healy, J. Melville, (2018) UMAP: Uniform Manifold Approximation and Projection for dimension reduction, arXiv:1802.03426v2] were calculated using the MATLAB package of Meehan, Meehan and Moore [Connor Meehan, Stephen Meehan, and Wayne Moore (2020). Uniform Manifold Approximation and Projection (UMAP) (https://www.mathworks.com/matlabcentral/fileexchange/71902), MATLAB Central File Exchange.] The UMAP is partially supervised, with 2/3rds of the patients in each condition randomly chosen to train the network. The UMAPs are set to have 45 nearest neighbours, with a minimum distance of 0.3 with a correlation metric. The UMAP was reduced over 2000 epochs.

##### Linear correlation between analytes / Linear correlation between clinical traits

The correlation coefficient, r, was calculated using a Pearson correlation coefficient (corcoeff, MATLAB). All quoted r values have an associated p-value (also computed by corcoeff) of less than 0.05.

##### Correlation network analysis

All values used for the correlation network analysis were the FI of Luminex results and/or the clinical readout. Severities were scored as HV=0, CH=1, CM=2, CS=3 and CC=4. Quality control (QC) was conducted on the matrix of expression values. Seven out of total 51 proteins were removed from the correlation analysis due to their low FI (<10 after subtracting FI in blank) and small SD (<10). Pearson correlation coefficient was performed using pairwise-complete correlation. Correlation matrix plots were generated using a modified package corrplot. Correlation matrix summary plots were made manually by R. Network plots were created by ggraph.

#### Tims-TOF mass spectrometry analysis

##### Primary data analysis (Identification and Quantitation)

Data was analysed by the Fragpipe pipeline consisting of Fragpipe 13.0 (Kong et al., 2017), MSFragger 3.0, Philosopher 3.2.9 (da Veiga Leprevost et al., 2020) and Python 3.8.2. Blank runs were excluded and each file defined as experiment to facilitate LFQ. Data were searched against a fused target/decoy database generated by Philosopher and consisting of human UniProt SwissProt sequences and UniProt SARS-nCov02 (retrieved 17/07/2020), plus common contaminants. The database had 40860 entries (including 50% decoy entries). MSFragger parameters were set to allow a precursor mass tolerance of plus/minus 10ppm and a fragment tolerance of 20ppm. Isotope error was left at 0/1/2 and masses were set to re-calibrate. Protein digestion was set to semi specific trypsin with up to 2 allowed missed cleavage sites, allowing peptides between 7 and 50 residues and mass range 500 to 5000 Da. N-terminal protein acetylation and Methionine oxidation were set as variable modifications. ID validation was done with PeptideProphet and ProteinProphet (Nesvizhskii et al., 2003) with default settings.

Label free quantitation was conducted with IonQuant (Yu et al., 2020) and Match-Between-Runs enabled (with default parameters) and using Top-3 quantitation. Feature detection tolerance was set to 10ppm and RT Window to 0.6 minutes with an IM Window of 0.05 1/k0. For matching, ion, peptide and protein FDRs were relaxed to 0.1 and min correlation set to 0 in order to allow pre-fractionated library samples to be included. MBR top runs was set to 600.

##### Data Handling

The proteomics dataset was processed as follows: (1) Protein filtering such that proteins with at least 50% of valid values in one group were kept; (2) Sample filtering such that samples with more than 50% of missing values were removed from the dataset; (3) Data normalization with log_2_ transformation and median-centring of the dataset. Imputation of missing values was performed using a mixed model that combines a K-Nearest Neighbour approach (KNN) when at least 60% of valid values are present, otherwise a Minimum probability approach is used where missing values are randomly drawn from a Gaussian distribution (shift=1.8, nstd=0.3). The resulting data matrix contains 353 samples and 105 proteins. Thirteen samples were further excluded from analysis for malignancy, immunosuppression, or being alternative samples.

##### Statistical analysis

Unsupervised hierarchical clustering was performed based on Euclidean distance and Ward’s method for calculating linkage. Differential abundance analysis was performed by fitting protein abundance in linear models with the limma package, using only one sample at the maximal severity of the patient and including age and sex as covariates. The Benjamini-Hochberg procedure was applied to correct for multiple comparisons. FDR < 0.05 and fold change > 1.5 was taken as statistical significance. Pathway enrichment analysis was performed using the XGR package with annotations either from Gene Ontology Biological Process or the Reactome pathway database. Significantly enriched terms were defined by FDR < 0.05 in hypergeometric tests with all proteins detected in plasma (including in library samples) as the background. Statistical analysis was performed in R.

##### Protein-protein interaction network

Protein-protein interaction data was retrieved from STRING v11 database with a confidence score cut-off of 0.7 and zero additional interactors. The network was visualised through Cytoscape v3.8.0 platform (Shannon et al., 2003) using perfuse force directed layout, and divided into clusters with the Markov cluster algorithm applied in the “clusterMaker” plugin. Node colour was mapped to Pearson’s correlation coefficients of PC1 scores and the protein level across samples, as lower PC1 score was shown to correlate with higher disease severity.

##### Clinical knowledge graph

The MS-based proteomics data were also analysed using the Clinical Knowledge Graph (CKG) (Santos et al., 2020). CKG provides a Python framework for downstream analysis and visualization of proteomics data: protein ranking, dimensionality reduction, functional Principal Component Analysis (PCA), Analysis of Variance (ANOVA), protein-clinical variable correlation analysis and network summarization.

CKG runs 3 feature reduction algorithms: Principal Component Analysis (PCA), Uniform Manifold Approximation and Projection (UMAP) and functional PCA. The functional PCA is based on the results of the method single-sample Gene Set Enrichment Analysis (Subramanian et al., 2005), which identifies enrichment of Biological processes (Gene Ontology) (GOBP) in single samples derived from the ranked intensities of the identified proteins. This method generates a vector of biological processes enrichment scores for each sample. Loadings of the top 15 proteins and GOBP driving the separation of the conditions studied are included in the PCA and functional PCA respectively. In this analysis, the drivers are biological processes such as acute-phase response and inflammation and retinoid and lipoprotein metabolic processes and cholesterol transport.

We performed ANOVA analysis to identify differentially regulated proteins across conditions. Further, we run posthoc analysis (pairwise t-test) to show specific differences when comparing disease conditions to healthy volunteers or community COVID-19 and also between severity levels.

We performed functional enrichment analysis (Fisher’s exact test) to identify enriched GO among the up- and down-regulated proteins in each pairwise comparison. Enrichments are plotted as scatterplots showing up- or down-regulated enriched GOBP and their adjusted p-values (Benjamini-Hochberg FDR: cutoff 0.05).

CKG performed a Spearman correlation analysis using the clinical metadata and the proteomics dataset. This Protein-clinical correlation is shown in a network where nodes are either clinical variables (diamond shape) or proteins (circle shape), and edges represent correlation (red=positive correlation, blue=negative correlation; Spearman correlation cutoff>=0.5; Benjamini-Hochberg FDR: cutoff 0.05). CKG applied a clustering algorithm (Louvain community detection) to identify clusters of highly connected nodes (nodes coloured by cluster).

The results from all CKG analyses are summarized in a single visualization all the findings in the different analyses and all relevant biomedical context associated (diseases, drugs, biological processes, pathways, protein complexes, publications). The summarization algorithm prioritizes what nodes are shown in the network based on betweenness centrality. The top 15 central nodes are shown for each node type.

#### Similarity network fusion analysis

The similarity network fusion (SNF) analysis was performed using the function in CKG that makes use of the python library pySNF. The method is used to analysed the proteomics datasets (MS-based proteomics and luminex) in combination to identify in an unsupervised manner clusters of similar patients. The number of clusters was not defined initially and optimized using the eigengap method and the clusters identified using Spectral clustering. The SNF analysis used Euclidean distance to calculate similarity (k_affinity=20, mu_affinity=0.6). The function returns the clusters and a mutual information score for each feature included in the analysis (MIscore). We used this score to prioritize a reduced number of features mainly driving the separation of the clusters (11 features) (MIscore>=0.15). The clusters are visualized using PCA plots.

##### SNF Cluster validation

In order to validate the identified clusters using SNF on the proteomics data, we used an independent cohort studied using a different technology, namely targeted proteomics by Olink (Filbin et al., 2020). Access to the dataset was granted by Olink https://info.olink.com/mgh-covid-study-overview-page. The processing of the data was done using CKG analytics core functions to map protein identifiers to names, transform the data into wide format and impute missing values using a mixed model as previously described. The clustering on these data followed a similar approach to how the SNF clusters were calculated (optimal clusters and Spectral clustering) but using only the selected features in the SNF analysis (7/11 features).

##### SNF clusters survival analysis

To evaluate the clinical relevance of the identified clusters in COMBAT, we performed a survival analysis and plotted the Kaplan-Meier curve using R packages survival (Therneau and Grambsch, 2000) and survminer. The input data is a data frame specifying the time to event, the event (death or end of observation) and the groups (SNF clusters). The comparison of the survival distributions between clusters was performed and the p value given using log-rank test. The hazard ratio was calculated using Cox proportional hazard model. In the Olink dataset survival status is only available at 4 timepoints: 0, 3, 7 and 28 days. Deaths at these time points were collected according to WHO category 1, defined as death, in these timepoints (WHO 0, WHO 3, WHO 7 and WHO 28). We compared the 28-day mortality between the two SNF clusters by chi-squared test.

##### Olink and COMBAT correlation analysis

To evaluate the robustness of the identified clusters between technologies/studies and to eliminate that only the 7 selected features used for the clustering of the Olink data were similar, we calculated the correlation (Pearson correlation) of fold-changes between clusters for all the common proteins in these studies (n=43).

#### Topological data analysis

Mapper is an algorithm in Topological Data Analysis for multiscale clustering and attempts to capture the topology of complex high-dimensional data (Singh et al., 2007; van Veen et al., 2019). We analysed the whole blood total RNAseq with a filter function (L2 norm) of principal component 1 to produce a Mapper graph. We used 25 evenly spaced bins with 50% overlap of the filter functions image. We used DBSCAN with eps=70, minpts=1 (equivalent to single-linkage with threshold 70). The clustering algorithm is applied separately to each preimage of each bin under the filter function. These clusters are the vertices of the final graph. As the bins are overlapping, the clusters between the preimage of different bins may share points. When this happens, an edge is drawn between the clusters that share a point. The resulting graph is the Mapper output with the vertices of this graph coloured by the average filter function value on that cluster.

#### Tensor and matrix decomposition

A tensor and matrix decomposition method, Sparse Decomposition of Arrays (SDA), as defined in (Hore et al., 2016), was used to integrate 152 samples from the different ‘omics datasets defined above, allowing that some samples were missing certain ‘omics data.

The whole blood total RNAseq (9 missing samples) and the pseudobulk from 10x CITEseq scRNAseq (22 missing samples) were combined into a three dimensional tensor consisting of 152 samples by 9 tissue types by 14,989 genes (which passed QC in both datasets). This expression was normalised by sample in each tissue type by log2-transformation of counts-per-million + 1.

The number of cells per cell types as defined by 10x CITE-seq (a two-dimensional matrix of 152 samples by 97 cell types, with 22 samples missing) and mass cytometry (CyTOF) (One two-dimensional matrix of 152 samples by 10 cell types for the all cells dataset, with 21 samples missing. Another two-dimensional matrices of 152 samples by 51 cell types for the granulocytes-depleted cells dataset, with 20 samples missing). We filtered out any samples with fewer than 500 cells in any matrix. The data in each matrix was normalised by a log2 transformation of counts-per-million + 1.

The proteomics data from Luminex (in a two-dimensional matrix of 152 samples by 51 proteins, with 20 samples missing) and mass spectrometry (Tims-TOF) (in a two-dimensional matrix of 152 samples by 105 proteins, with 17 samples missing) were used, with the data normalised as described in the Luminex and Tims-TOF sections.

As described in (Hore et al., 2016), to find robust components we ran the tensor and matrix decomposition ten times for 1000 components. Each time around 290 components were estimated to be zero. Once again, similar to the (Hore et al., 2016) method the absolute correlation (r) was calculated for the sample scores for each pair of components, clustered using Hierarchical clustering on 1-r (dissimilarity measure) and formed flat clusters in which the components in each flat cluster have no greater a cophenetic distance than 0.4. We chose the flat clusters that had components from at least 5 of the 10 runs. The final sample, tissue and gene or protein or cell score was the mean of all the components within the chosen clusters. This resulted in 381 clusters.

Components were identified as associated with COVID-19 if they (1) showed significant variation (BH adjusted p < 0.01) in an analysis of variance between the COVID-19 categories and healthy volunteers and (2) showed a significant |spearman’s rho| >= 0.5 (and Benjamini/Hochberg adjusted p < 0.01) in at least one of the contrasts between the COVID-19 groups vs healthy volunteers (in total we found n=130 such components; **Figure 7C**). To identify COVID-19 specific components the median loadings of the components for the different comparator (source) groups (i.e. including influenza and all-cause sepsis) were clustered (k-means). Individual component associations, for example with comparator group, severity or clinical features were assessed with Spearman correlation (between component loadings and numerical variables) or ANOVA (between component loadings and categorical variables). The overview heatmap (**Figure S7G**) was generated by combining the top significant (Benjamini/Hochberg adjusted p < 0.05, abs(r) > 0.5, max 10) components from each of the individual analyses. Pathway enrichment of the genes expression highlighted (those with posterior inclusion probability > 0.5, weighted by their ranking in loading score magnitude) in components was done using gene set enrichment analysis as implemented in Pi’s xPierGSEA (Fang et al., 2019).

#### Integrated data analysis of multi-omics data using machine learning feature selection to distinguish COVID-19 severity groups

We used supervised machine learning (from sklearn) to classify samples according to their WHO severity based on PCs obtained from data across the different modalities (timsTOF, luminex, and total RNAseq) (**Figure S7A**). We performed permutation feature scoring to find the most important PCs to predict severity. After that, we extract the most important features of the most important PCs and rerun the algorithm directly on these features, again ranking them according to their importance.

#### Machine learning using SIMON to distinguish COVID-19 and sepsis

##### Generation of an integrated COMBAT dataset

The pre-processed data from individual COMBAT assay datasets were automatically processed using the standard extract-transform-load (ETL) procedure to generate an integrated dataset. Datasets were merged using shared variable, the COMBAT sample ID and assay-specific sample IDs. Next, we generated novel features, such as name of specific assays to indicate if a sample was analysed in a specific assay (yes/no), ordering of samples shared between assays (*first_sample_across_assays*), day when sample was obtained from the maximum disease (*day_sampling_from_max_disease*), state of the disease when sample was acquired (recovered/ongoing), *hospitalized, ventilation, oxygenation, sampling* (<=10 days as early or >10 days as late phase of the disease), samples acquired before or after maximum disease (*sampling_from_max_disease) and disease progress for longitudinal samples (deterioration or recovery).* Donors that were excluded or frail were not included in the integrated dataset. We standardized names of the immune cells subsets and analytes to reflect the measurement, such as frequency (*freq*_cell subsets) or intensity (Luminex parameter_*intens*). In total, the integrated COMBAT dataset contained information on 428 samples from 268 donors on more than a million parameters.

##### Data pre-processing

For the multi-omics data integration, after filtering for samples not analysed across all assay modalities and restricting to the first available sample after admission from sepsis and hospitalized COVID-19 patients, the final dataset included 15 sepsis patients and 53 hospitalized COVID-19 patients with 184 features analysed using CyTOF, 79 using FACS, 8 using GSA, 105 mass spectrometry, 102 Luminex and 23,063 features from whole blood total RNAseq. The data for each assay was cantered and scaled, missing values were imputed, features with zero-variance and near-zero-variance were removed and finally, highly correlated features with cut-off 0.85 were also removed.

##### Feature selection process

To avoid ‘curse of dimensionality’, reduce overfitting and improve accuracy, we implemented a wrapper approach as described (Kohavi and John, 1997). Briefly, the initial dataset containing all 68 donors was partitioned into training (52 donors) and test set (16 donors) with balanced class distribution of sepsis and COVID-19 patients using the data partitioning function (Kuhn, 2008) as described previously (Tomic et al., 2019). The same training/testing dataset split was used for each assay. The reduction of features using the recursive feature elimination (RFE) algorithm was performed on the training set and the model was evaluated using the 10-fold cross-validation repeated 5 times. For whole blood total RNAseq, prior to training the model using RFE algorithm, we have performed analysis of differentially expressed genes between sepsis and hospitalized COVID-19 patients, reducing the number of genes to 2,989 based on the FDR <0.05 and fold change >1.5, and accounted for age and sex in the model. The RFE method removes the features that do not contribute to the final model, while it keeps the features that contribute the most to the final model as evaluated using the variable importance score (Kuhn and Johnson, 2019). Finally, after the RFE analysis, the selected features from each assay (36 features from CyTOF, 20 from FACS, 20 from Luminex, 32 from mass spectroscopy, 28 from whole blood total RNAseq and 1 from GSA dataset) were merged in the final training dataset containing 52 donors and 137 features.

##### Machine learning using SIMON

To identify immunological and molecular signature that can discriminate between sepsis and COVID-19 patients, we used SIMON (Sequential Iterative Modeling “Over Night”) (Tomic et al., 2019). SIMON is a free and open-source software that provides a standardized ML method for data pre-processing, data partitioning, building predictive models, evaluation of model performance and selection of features. For the analysis, we applied four machine learning algorithms, naïve Bayes, Shrinkage discriminant analysis (*sda*), Support vector machine with linear kernel (*‘svmLinear2’*) and C5.0 decision tree. Since the entire ML process in SIMON is unified, resulting models built with different algorithms can be compared and the best performing models can be selected. First, models are built on training set and the performance is evaluated using a 10-fold cross-validation repeated five times and cumulative error rate is calculated. To prevent overfitting, in the second step, each model is evaluated on the withheld test set. The performance of classification models was determined by calculating the area under the receiver operating characteristic curve (AUROC) for test set (test AUROC). The best performing model was built using the naïve Bayes (testing AUROC 0.85, 95% CI 0.59-1.00). In the final step, SIMON calculated the contribution of each feature to the model as variable importance score (scaled to maximum value of 100).

#### Data management

To support consistent and coherent communication of data and metadata within the project, a unified identifier system for all samples was implemented. The COMBAT sample identifier system encodes information regarding the sample providence in terms of cohort, de-identified patient ID, location where the sample was taken, the time point of the sample relative to the initial collection and details regarding the processing of the sample itself.

Datasets from each modality were stored within the consortium via the COMBAT datawarehouse, consisting of over 100TB of fast storage connected to a research computing cluster. This enabled data processing to occur within the datawarehouse, reducing the risk of duplication of datasets and the possibility of uncontrolled changes. Once datasets were ready to be shared within the consortium, to support (for example) data integration work, they were formally given a unique identifier and placed in a dedicated dataset directory. The existence of this deposition and its associated metadata, including information regarding associated samples and status was then made available via a web application which captures this information in a back-end database. This application allows consortium members to search by modality and status, providing information about the purpose of each dataset and its location in the datawarehouse.

The governance of data management was supported by the existence of a short but well-defined Data Access Agreement, which all consortium members were required to sign before gaining access to the datawarehouse. Furthermore, granular permissions within the datawarehouse enabled careful access controls to be applied to particularly sensitive data (such as rich clinical data). Web applications supporting the consortium are all protected by a federated Shibboleth-based authentication approach, allowing collaborators from outside of Oxford to gain access as required.

#### Data visualisation

##### Multi-locus viewer

In order to visualise the data from the different modalities (experiments), a module, Multi Experiment Viewer (MEV https://github.com/Hughes-Genome-Group/MEV) was built for Multi Locus View (MLV DOI 10.1101/2020.06.15.151837) such that the data was pivoted on sample rather than genome location. The data was loaded in from each modality and for the CITEseq data, psuedobulk values (on a sample/cell type basis) were used. In order to compare radically different datatypes (read counts, fluorescence, % cell type etc.), percentiles for each observation were calculated from the 10th to the 90th, in steps of 10. Values were then placed into 10 bins, e.g., if a value was <=10th percentile it would be given a value of 1 and if >2nd and <=3rd, it would get a value of 2 etc. This was used as the default value, although depending on the modality other values e.g, raw counts, log transformed values were also added and can be selected by the user. Users can create their own views by searching for genes or loading in specific data sets and then combine them with a limited set of clinical data. Charts such as histograms, heatmaps and scatter plots can then be added and cross-filtered to identify samples or features of interest. An instance can be found at https://mlv.combat.ox.ac.uk/, with links to predefined views of the data.

##### Shiny apps

Shiny apps (https://shiny.combat.ox.ac.uk) were developed using the R package shiny to display the results of the whole blood total RNAseq differential expression analysis and the principal component analysis. In each, derived data are loaded (limma fitted models and pre-calculated principal components respectively) together with limited metadata, and plots are generated within the app using ggplot2.

## ADDITIONAL RESOURCES

### KEY RESOURCES TABLE

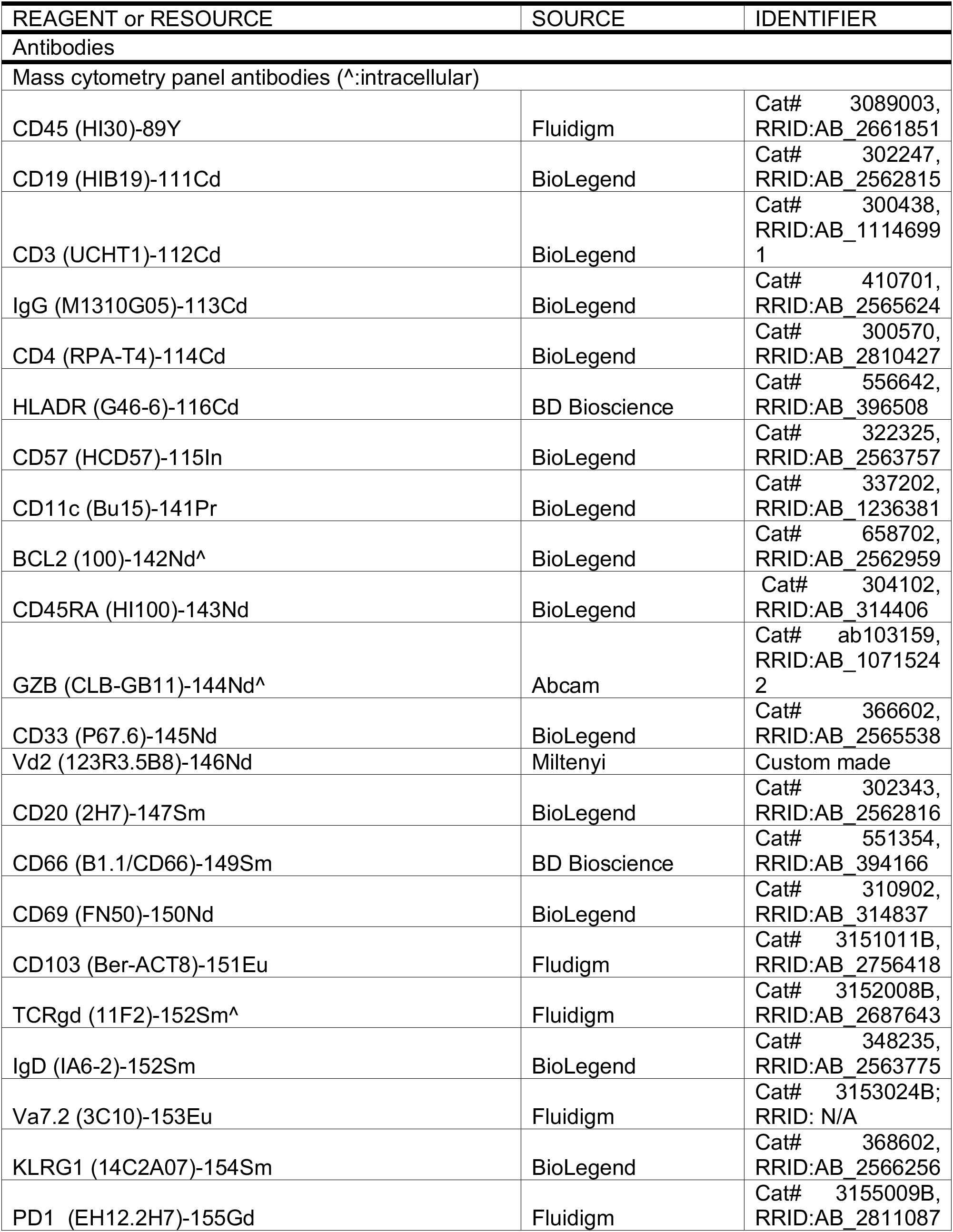
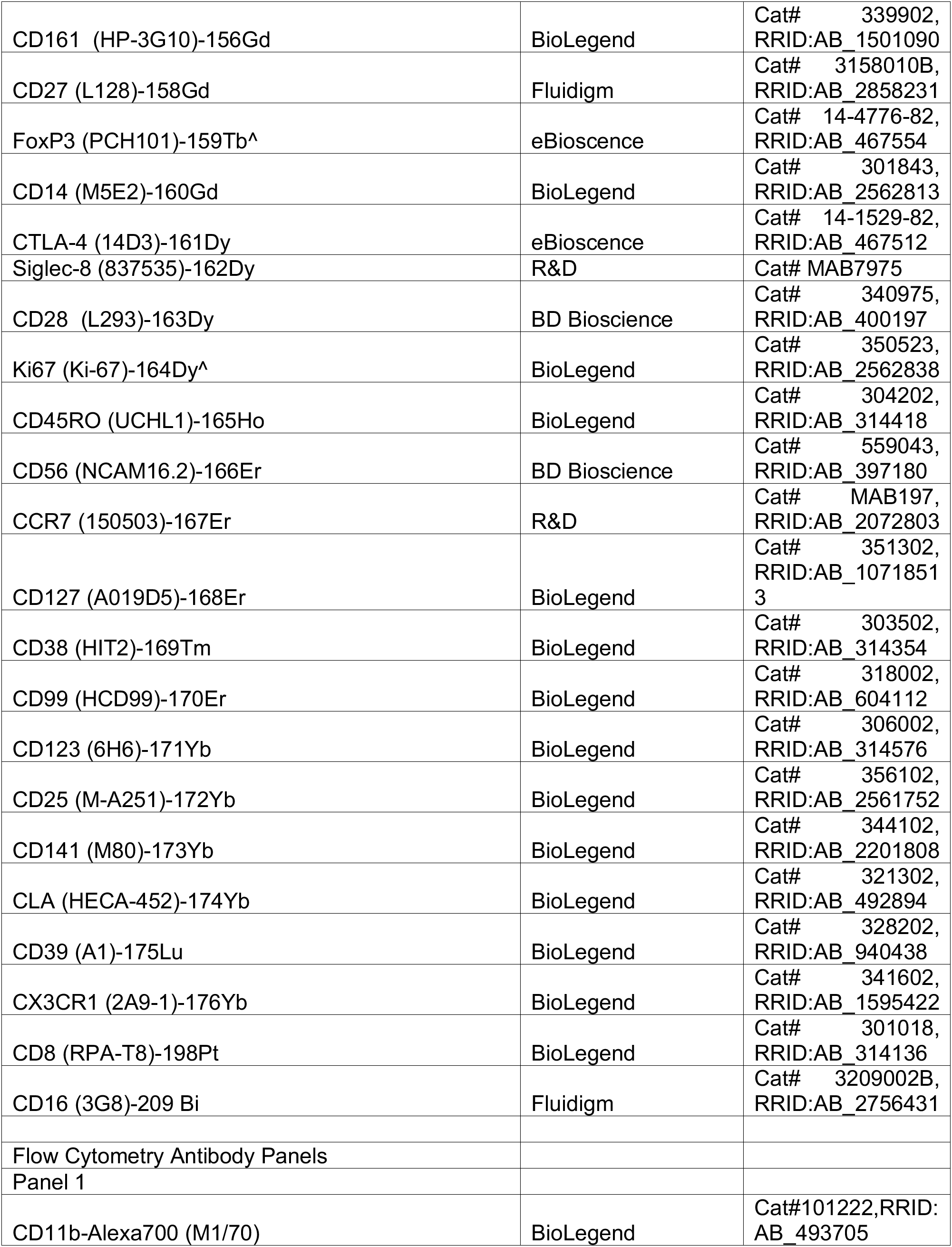
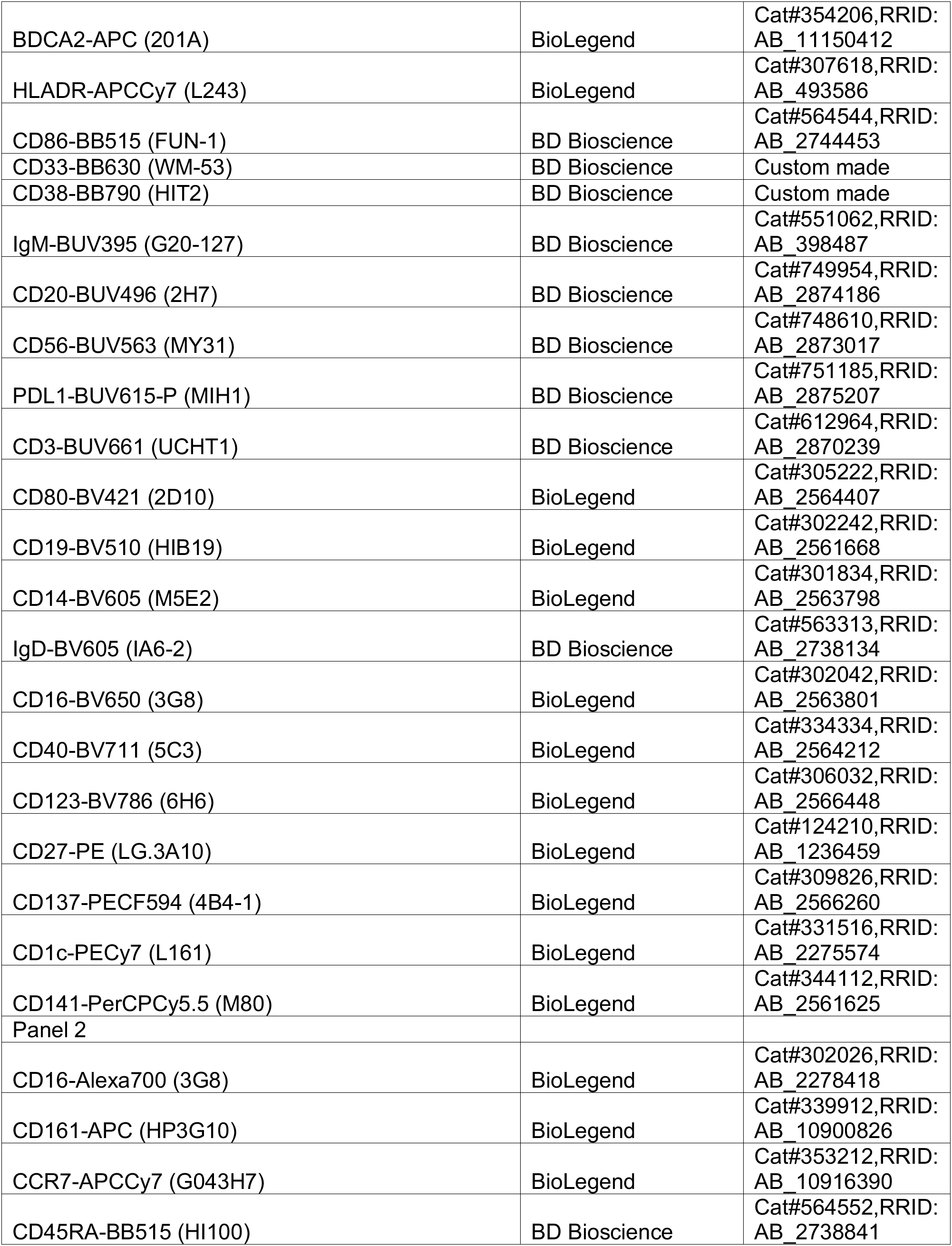
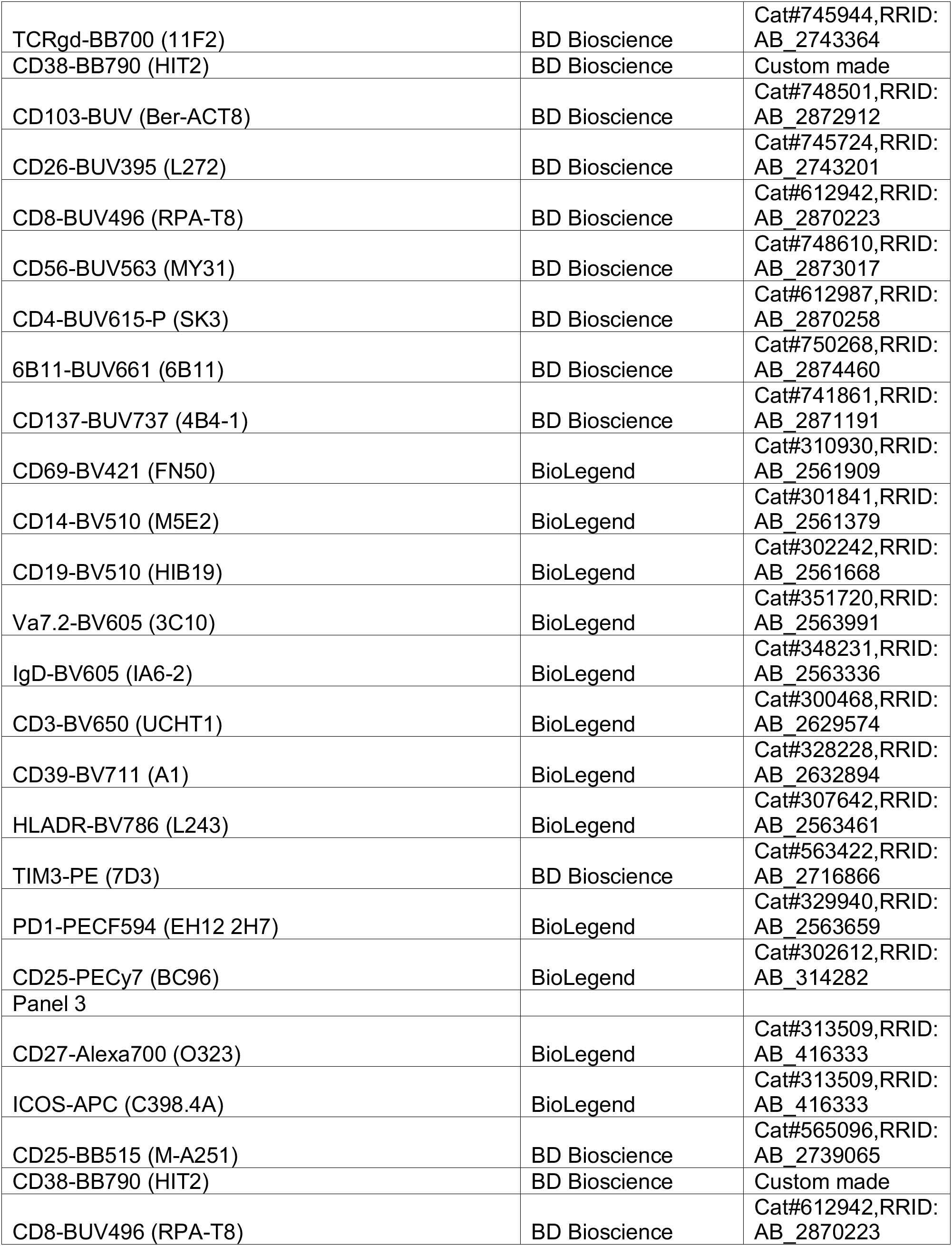
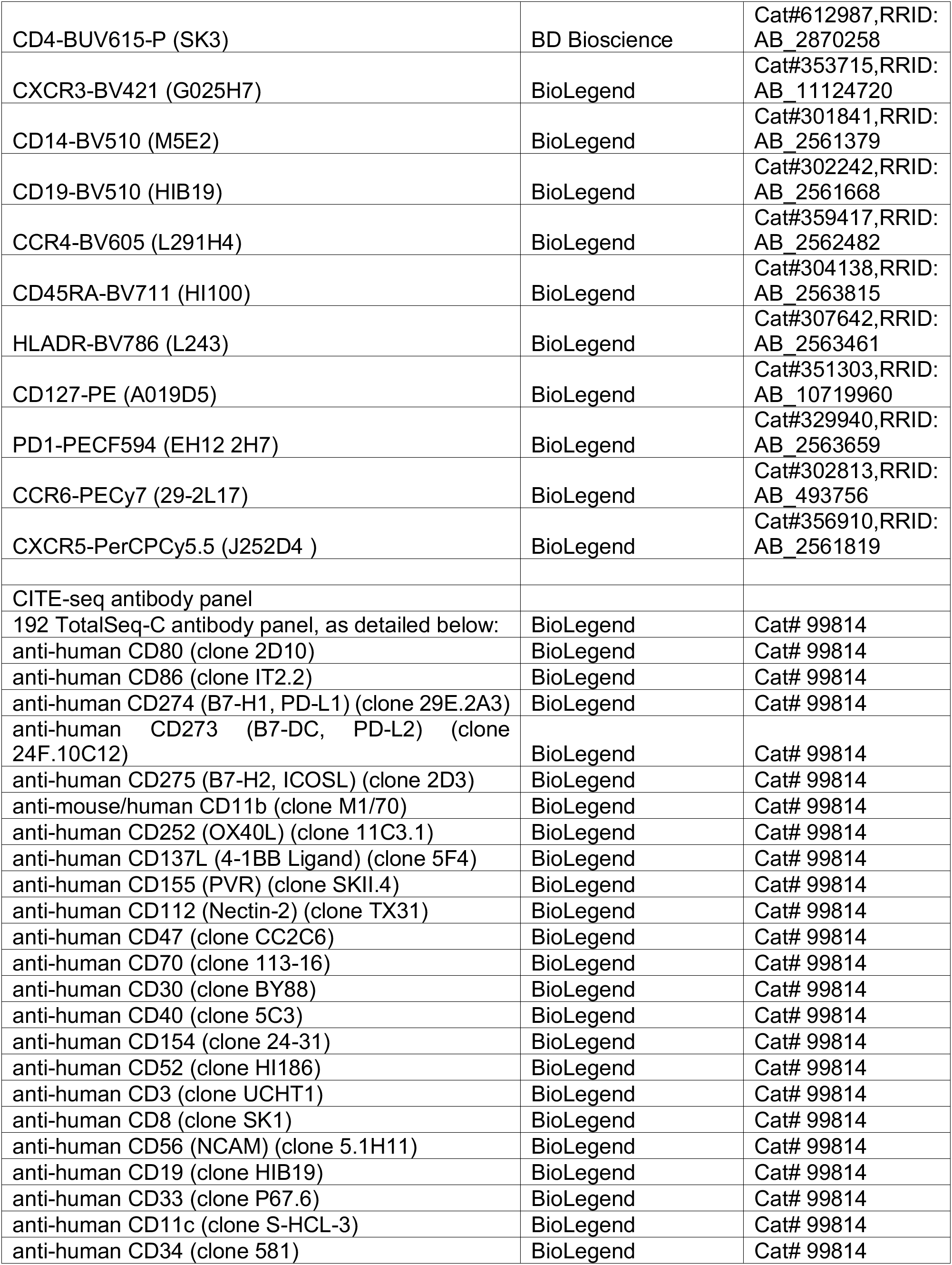
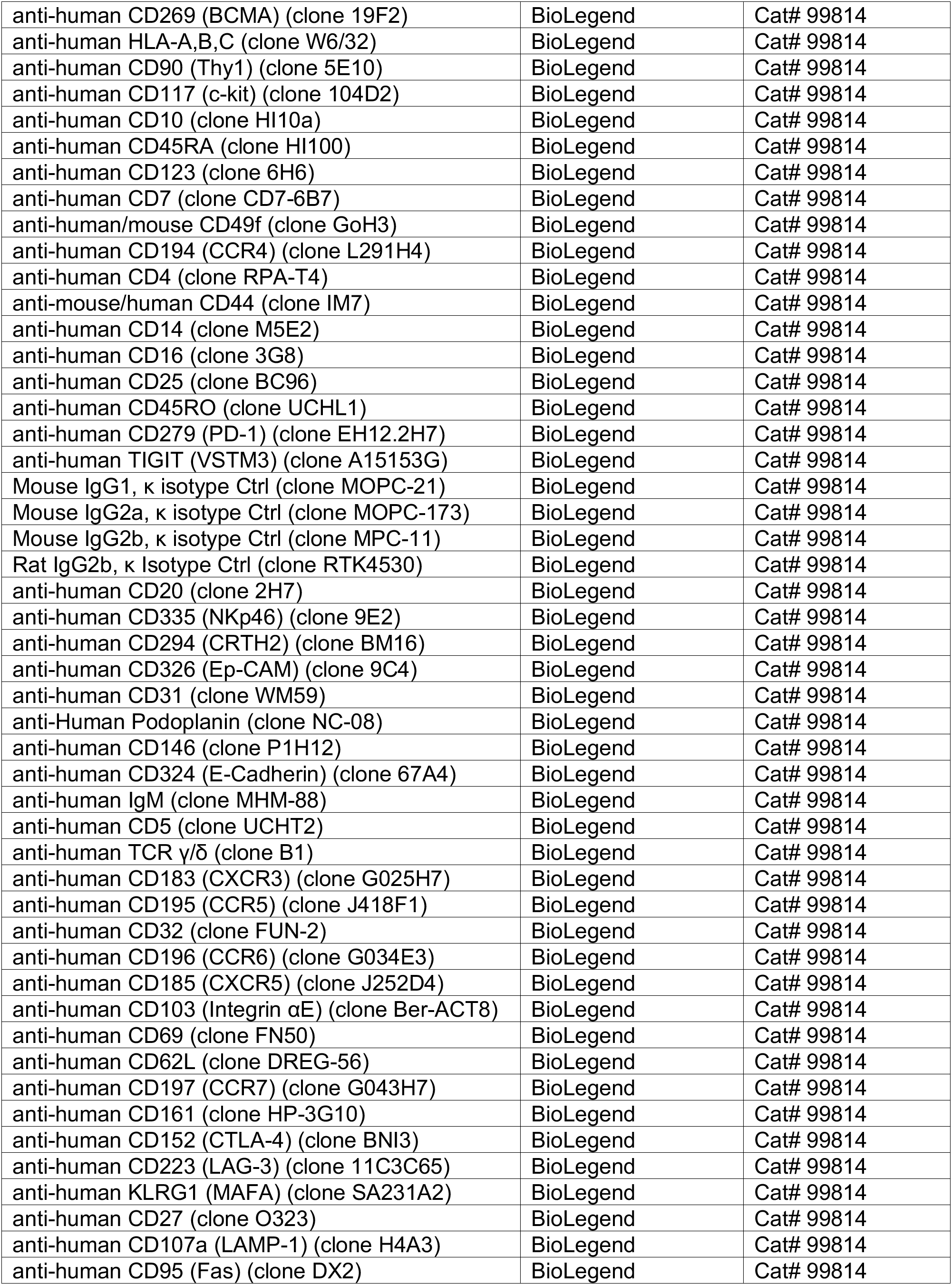
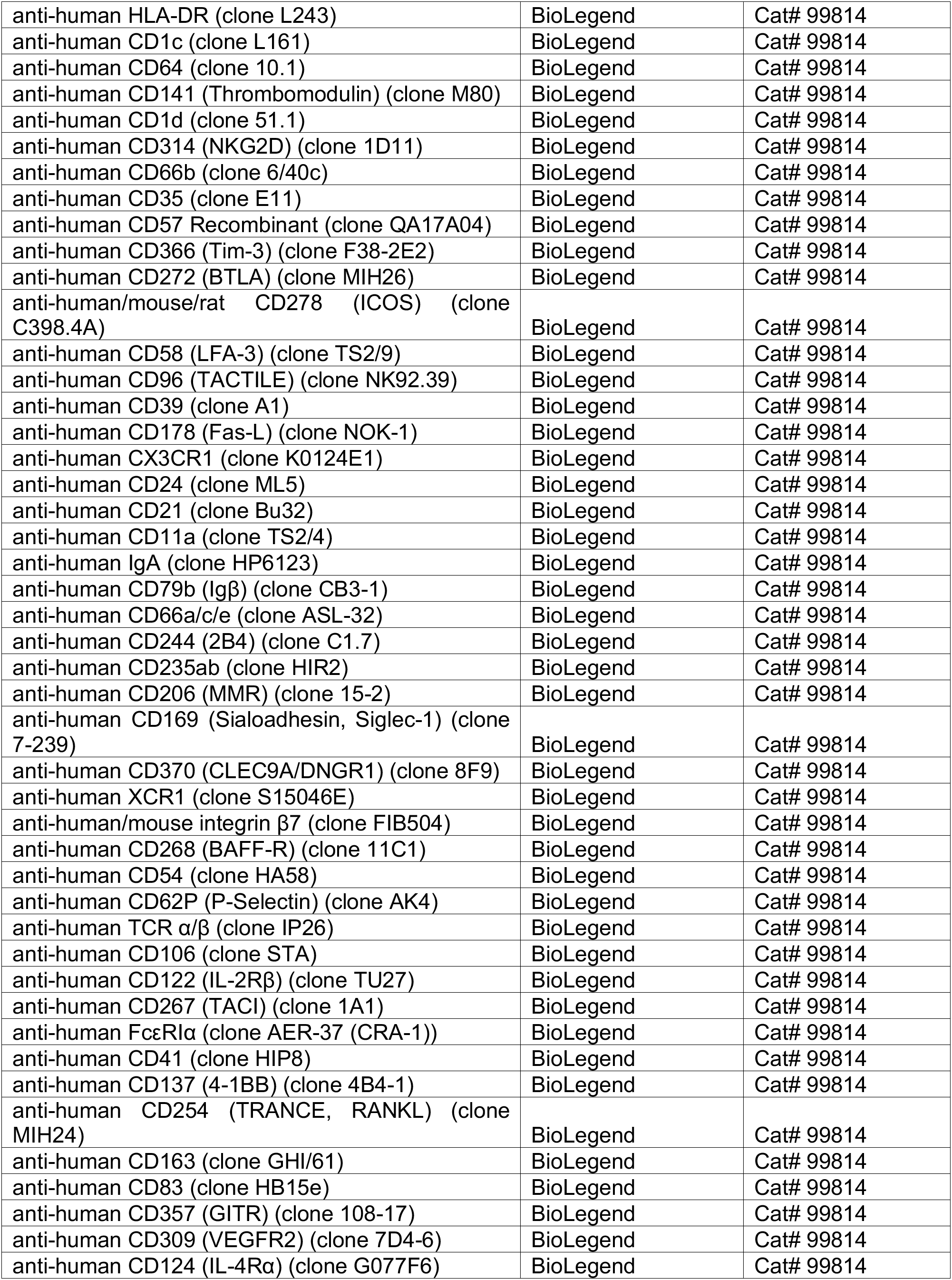
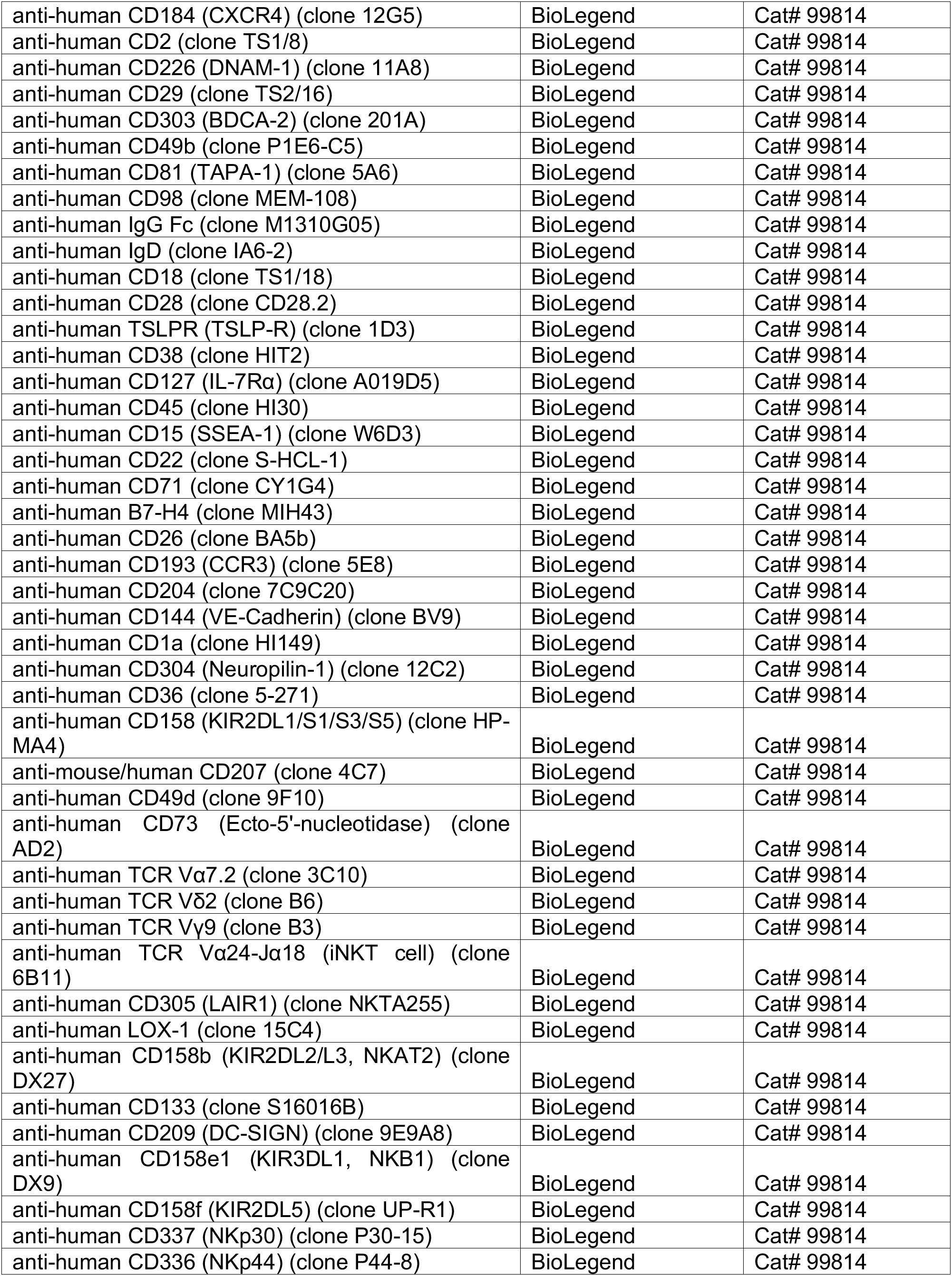
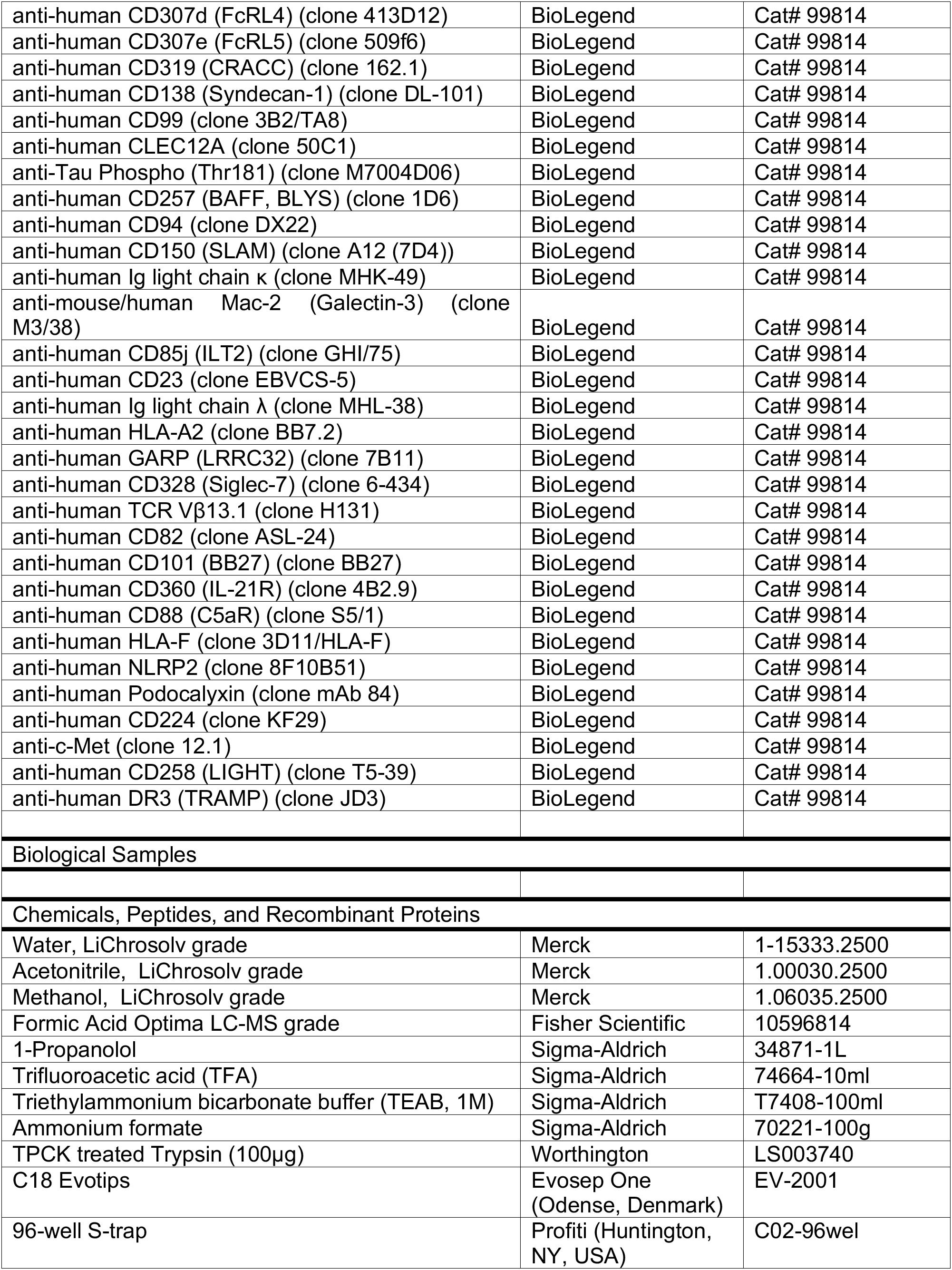
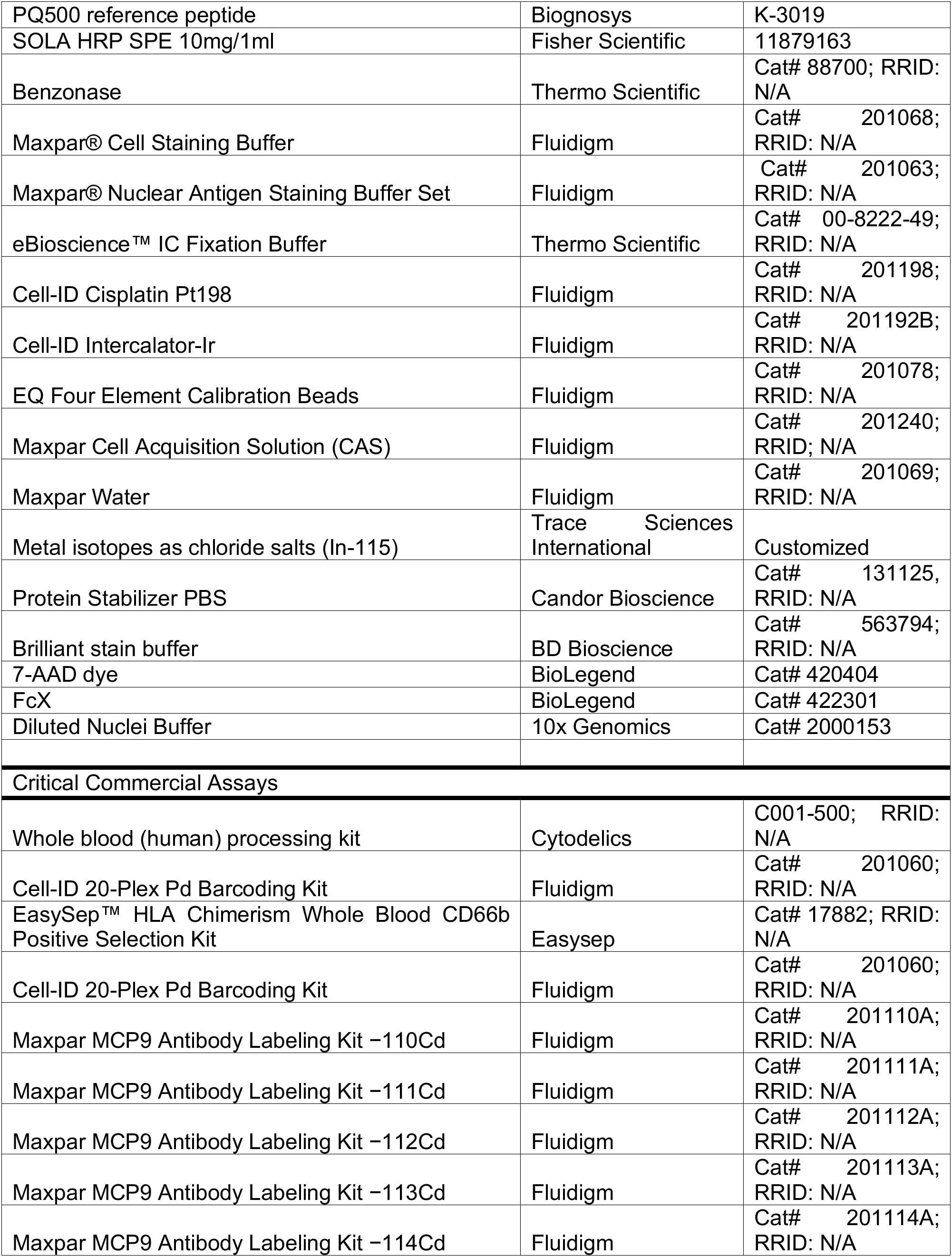
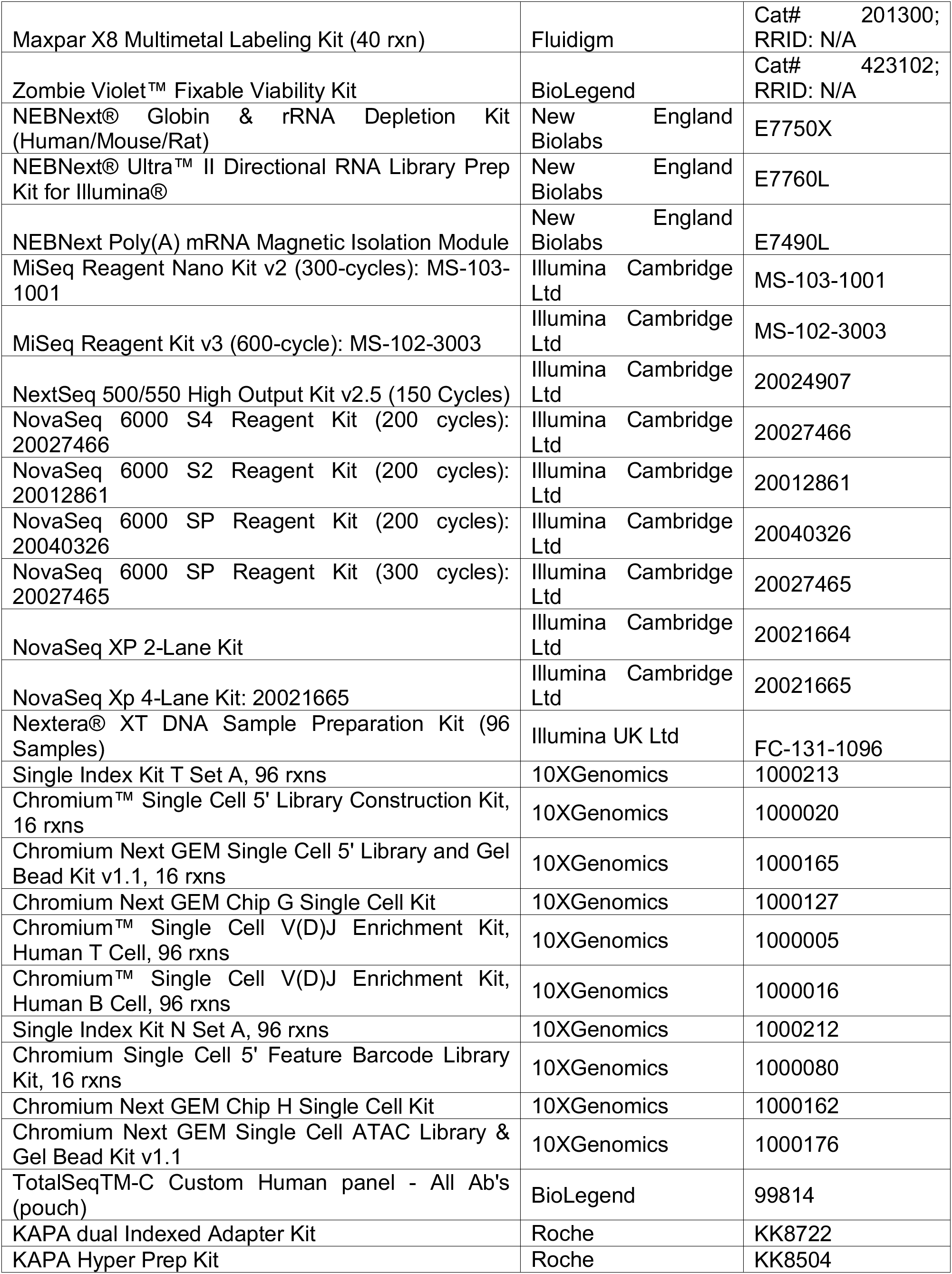
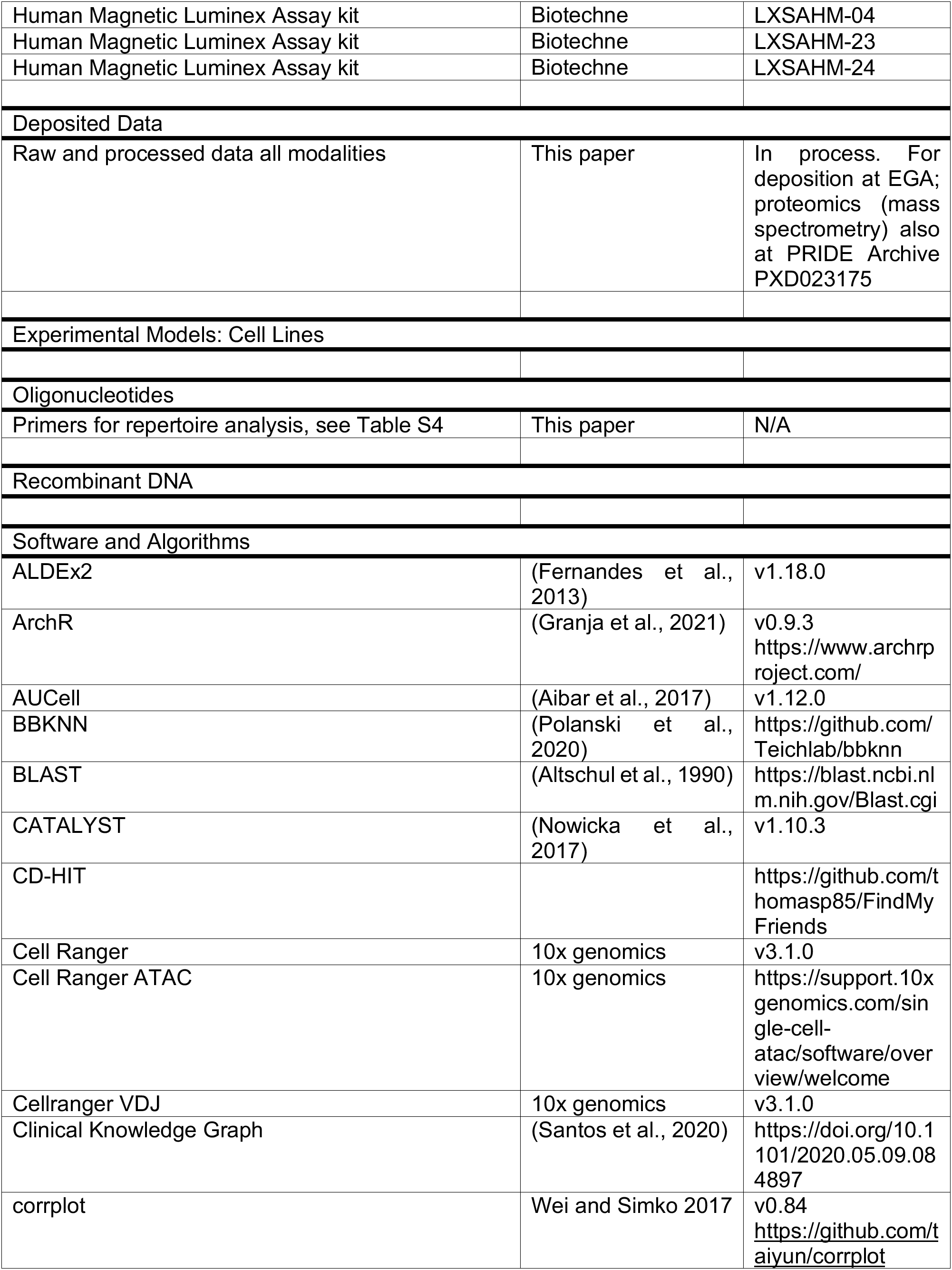
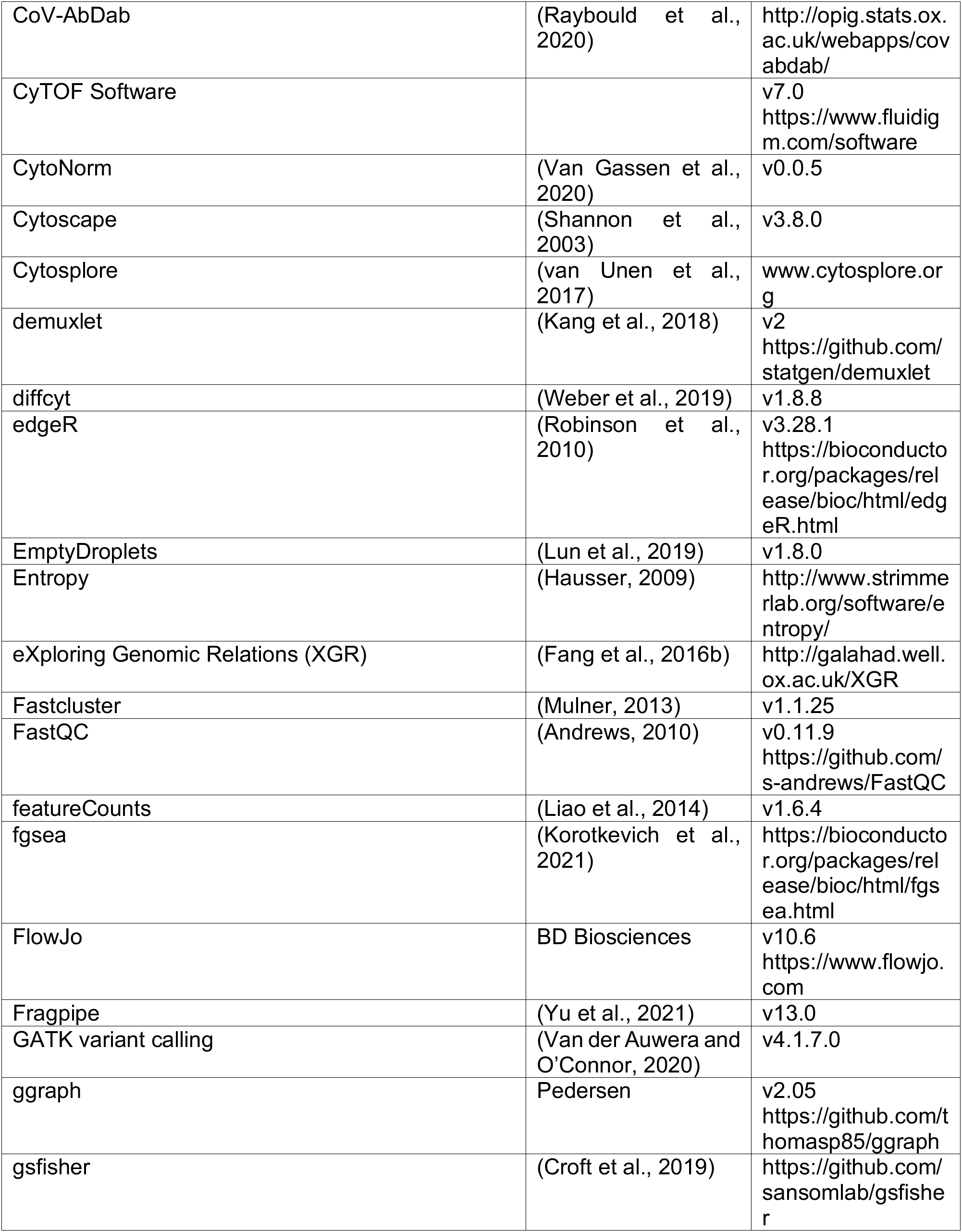
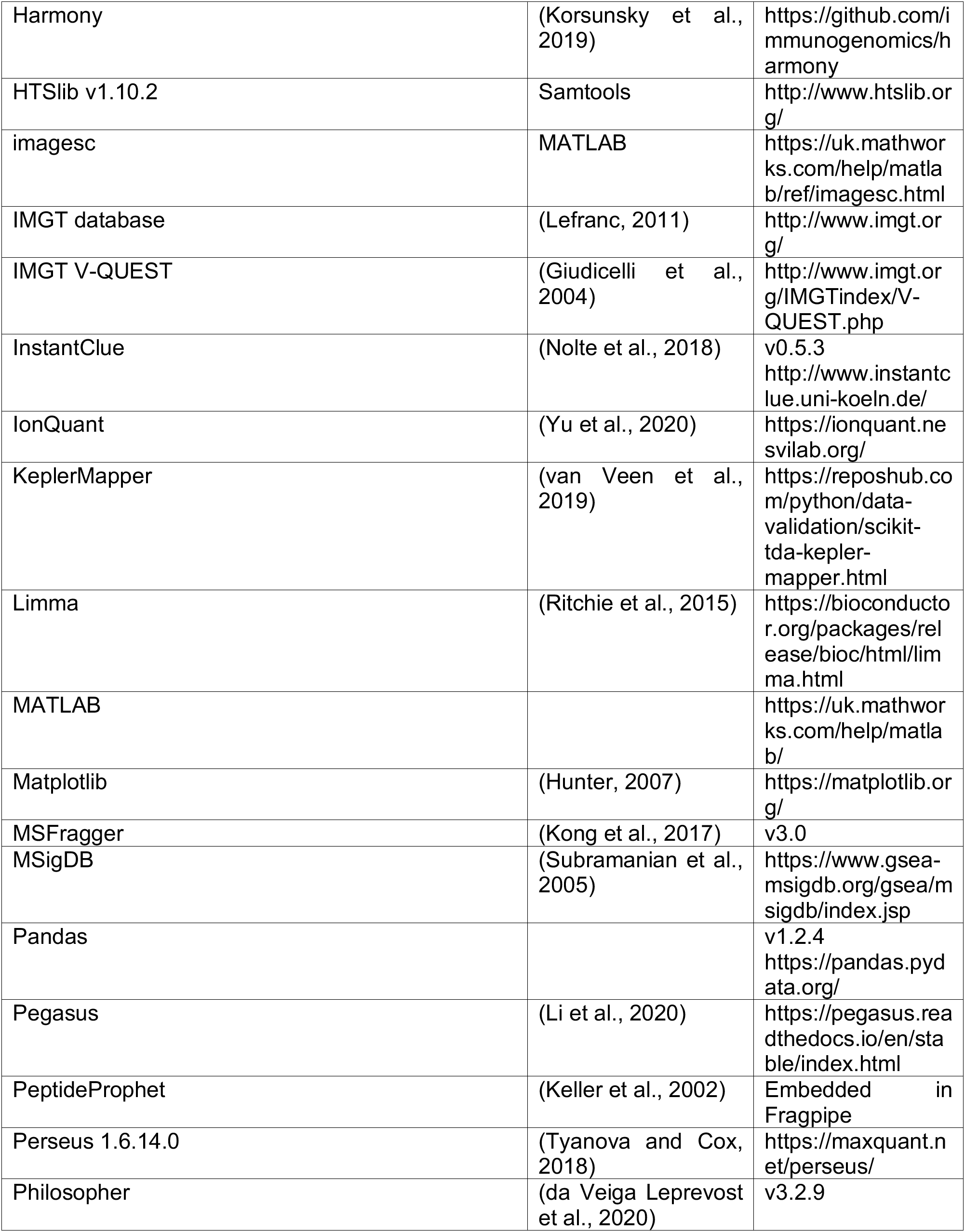
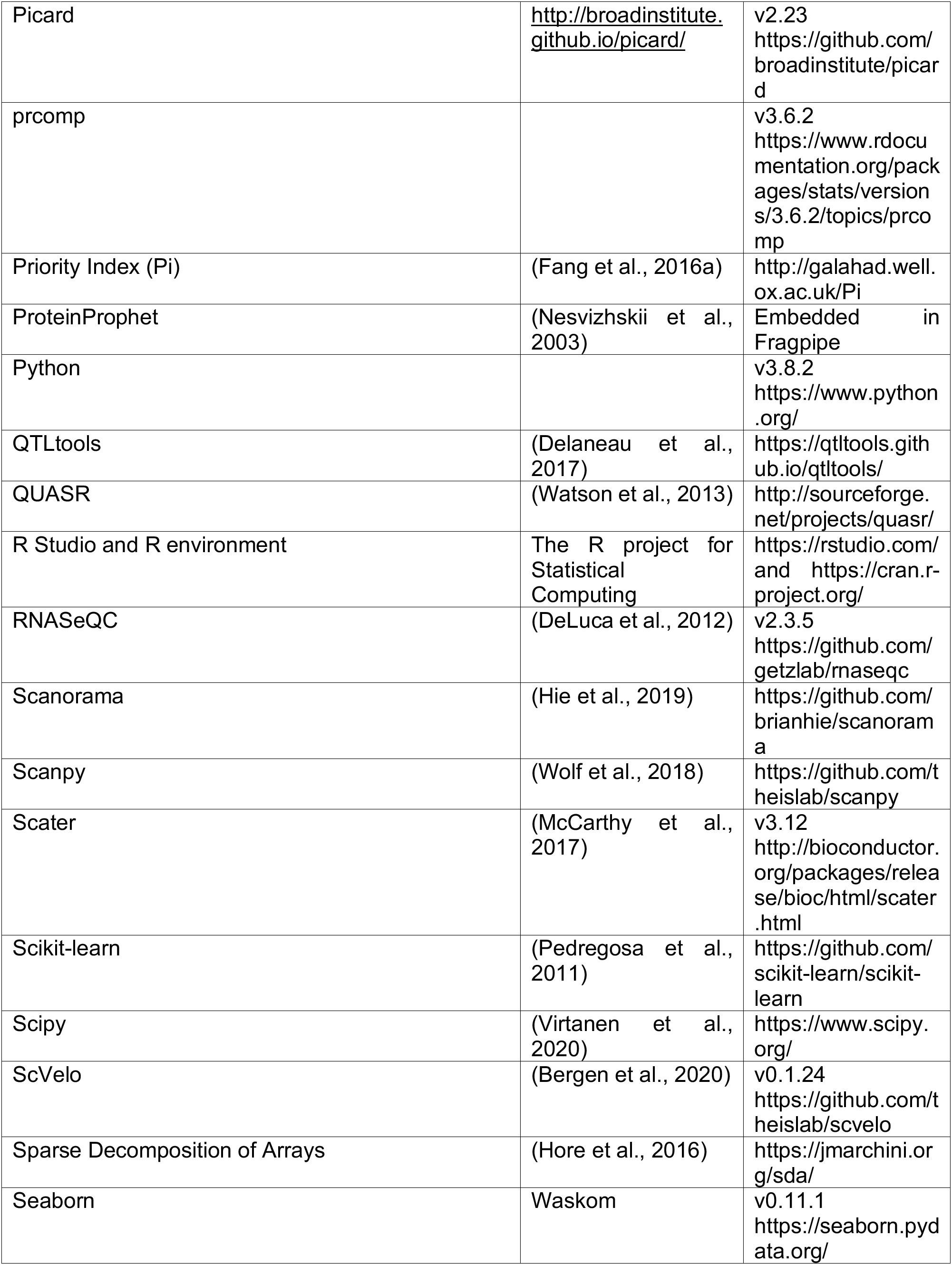
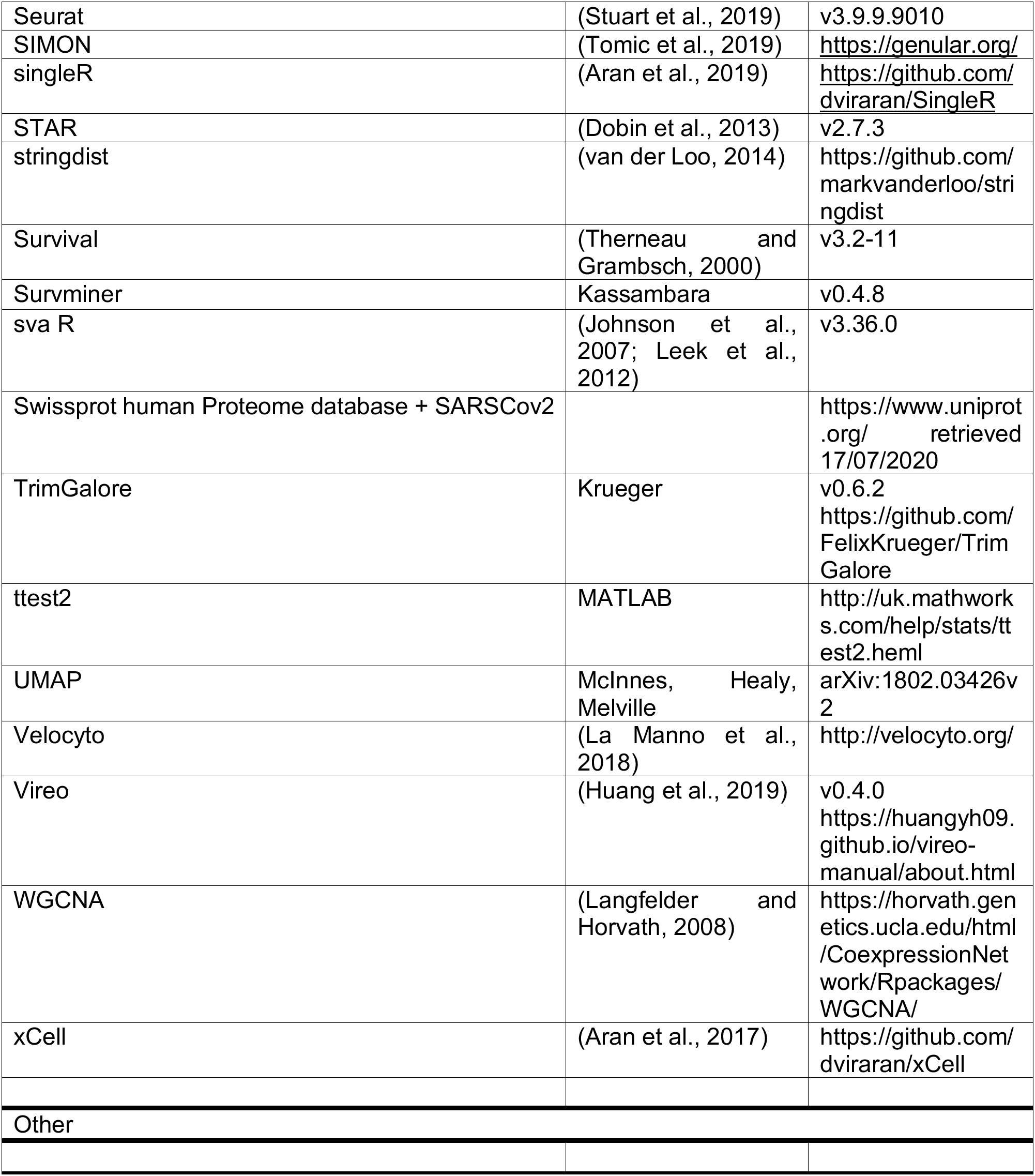

### SUPPLEMENTAL INFORMATION

**Table S1. COVID-19 Multi-omic Blood Atlas (COMBAT) Consortium author details**

**Table S2. Demographics and clinical features for overall cohorts and hospitalised COVID-19 patients**

**Table S3. Tensor and matrix decomposition analysis.** Data for 152 samples across modality types and all patient groups, including significant associations of components with group memberships, gender, detail of sample loading scores and pathway enrichment.

**Table S4. Primers used for BCR and TCR sequencing**

**Methods S1. Supporting figures for mass cytometry, related to STAR Methods.** Data analysis supplement including gating strategies, analysis pipeline and phenotyping.

**Methods S2. Supporting figures and tables for CITE-seq, related to STAR Methods.** Experimental design and cell capture, multimodal annotation of PBMC sub-populations, compositional analysis, PCA, differential expression analysis and pathway analysis.

**Methods S3. COVID-19 Multi-omic Blood Atlas (COMBAT) Consortium author contributions**

