## Supplementary material for "A blood atlas of COVID-19 defines hallmarks of disease severity and specificity": Methods S1

### Methods S1 - Supporting figures for mass cytometry, related to STAR Methods

**Figure 1-2:** Gating strategies for clean-up and doublets exclusion

**Figure 3:** Analysis pipeline

**Figure 4-6:** Heatmaps showing the phenotype of populations described in Figures 1D, 3C, 5A.

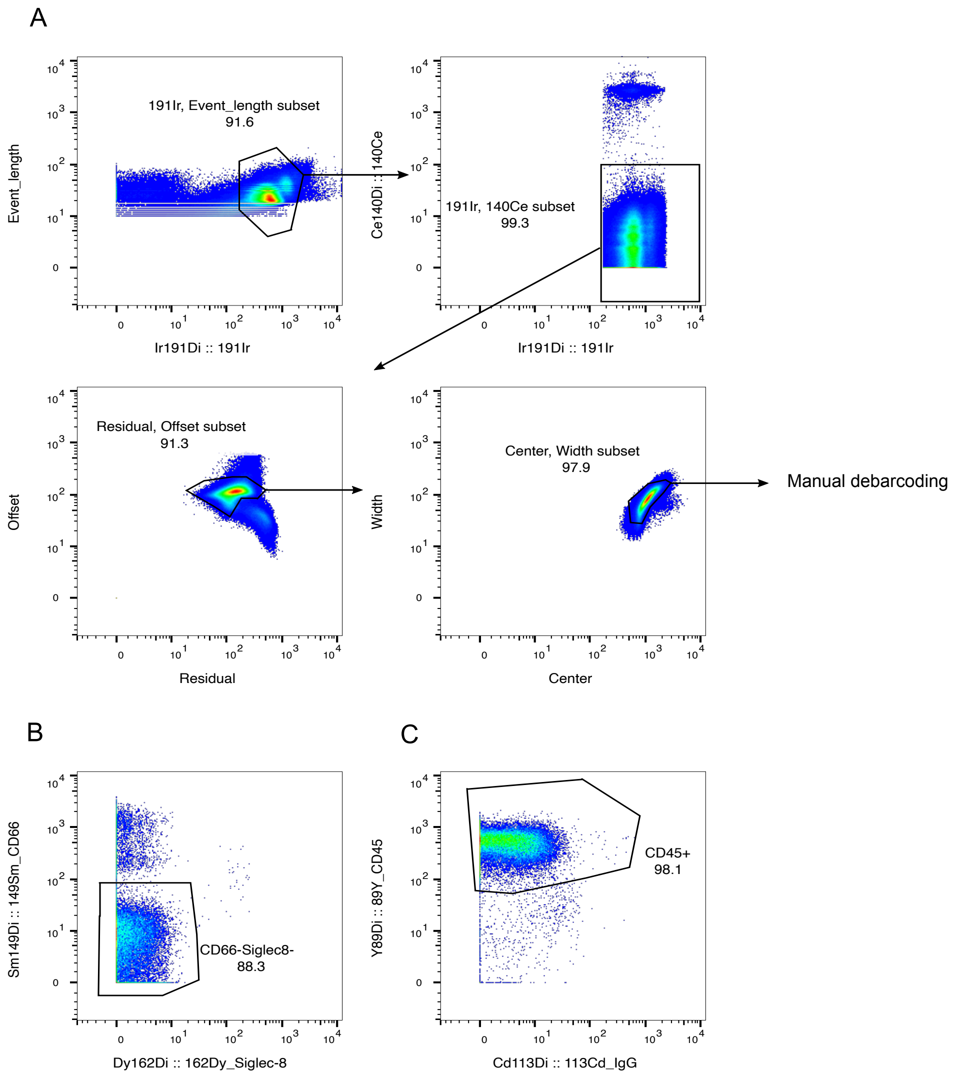


**Figure 1: A** Gating strategy for removal of beads, DNA negative events and for Gaussian parameters clean-up prior to Manual Debarcoding. **B** Clean up step to remove residual CD66+ granulocytes and Siglec-8 Eosinophils, in granulocytes depleted samples. This step was followed by gating of CD45+ cells. **C**. Gating of CD45+ cell for subsequent analysis. This step was performed for both depleted and non-depleted (original) samples.

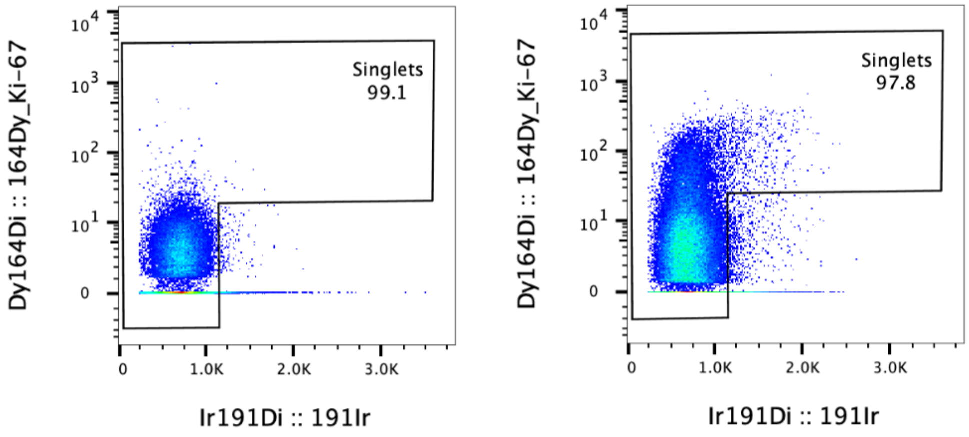


**Figure 2:** Strategy for doublets exclusion. Representative scatter plots showing DNA content on x-axis (191Ir - Iridium) versus the proliferation marker Ki67 on y-axis. The sample on the right shows a population of proliferating Ki67$^{+}$ cells absent on the sample on the left. To not exclude proliferating cells from the analysis, the gating was made excluding only Iridium$^{high}$Ki67$^{-}$ cells.

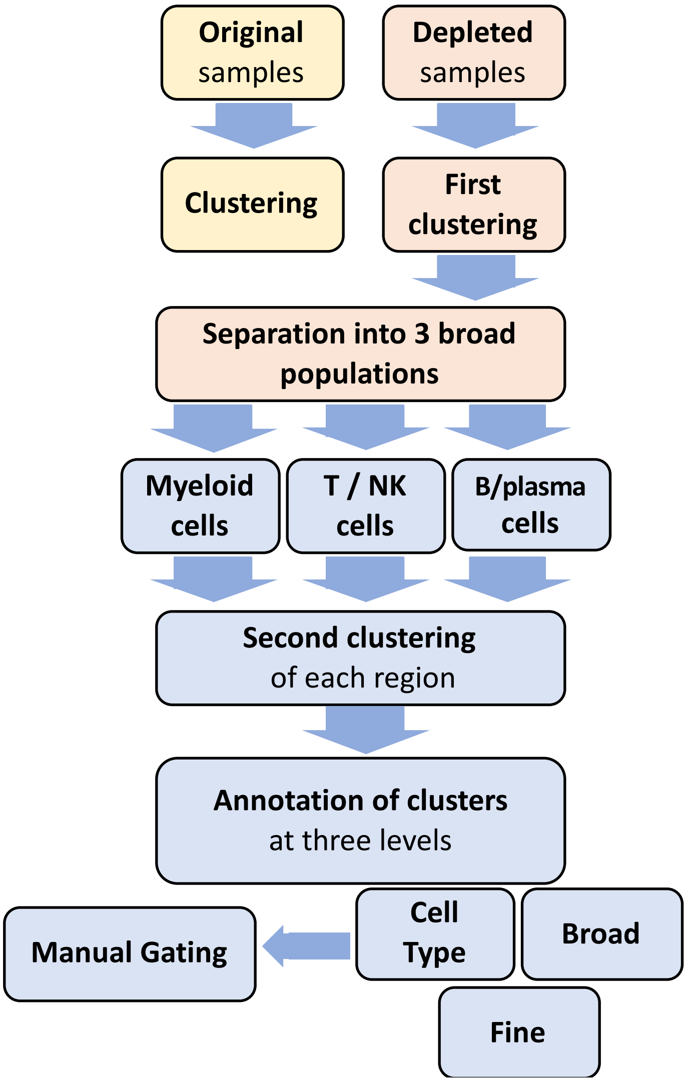


**Figure 3:** Flow-chart describing the analytical pipeline for the analysis of the Mass Cytometry data.

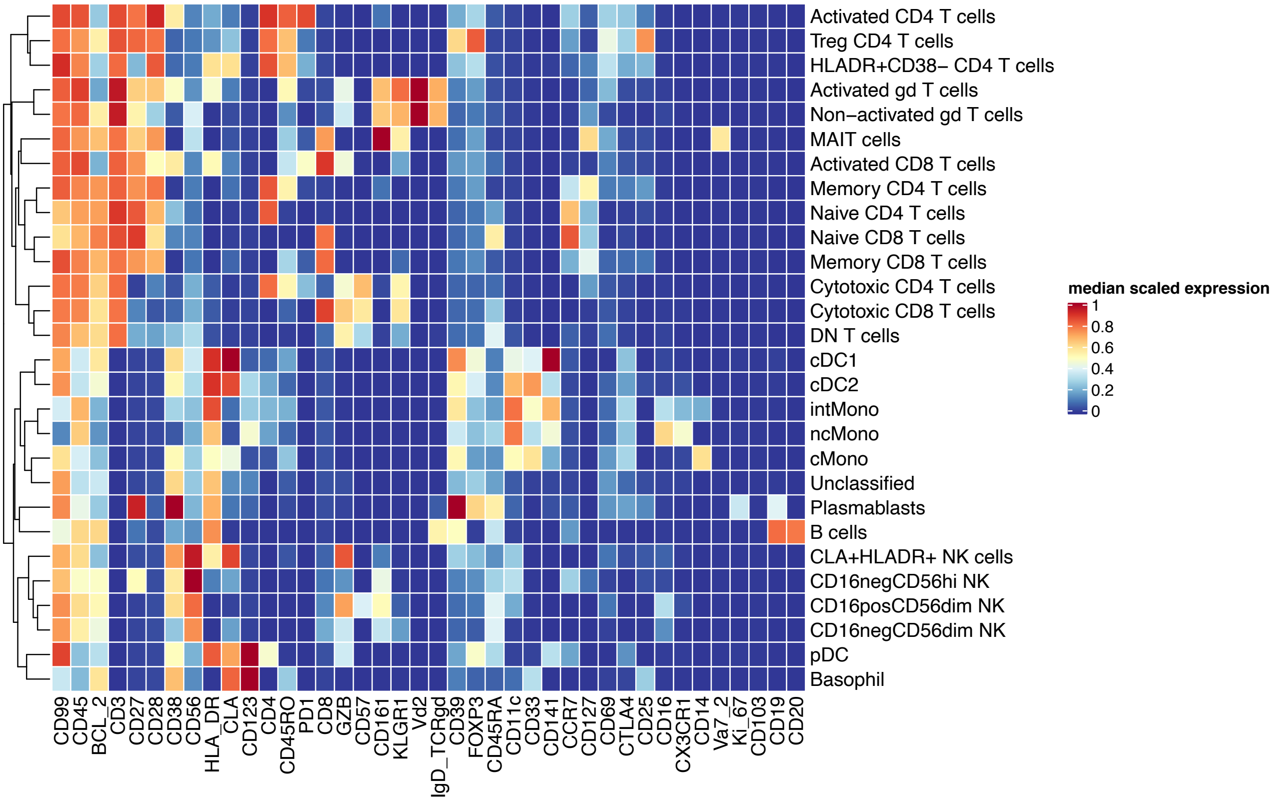


**Figure 4:** Heatmap showing the phenotype of broad cell populations described in Figure 1D.

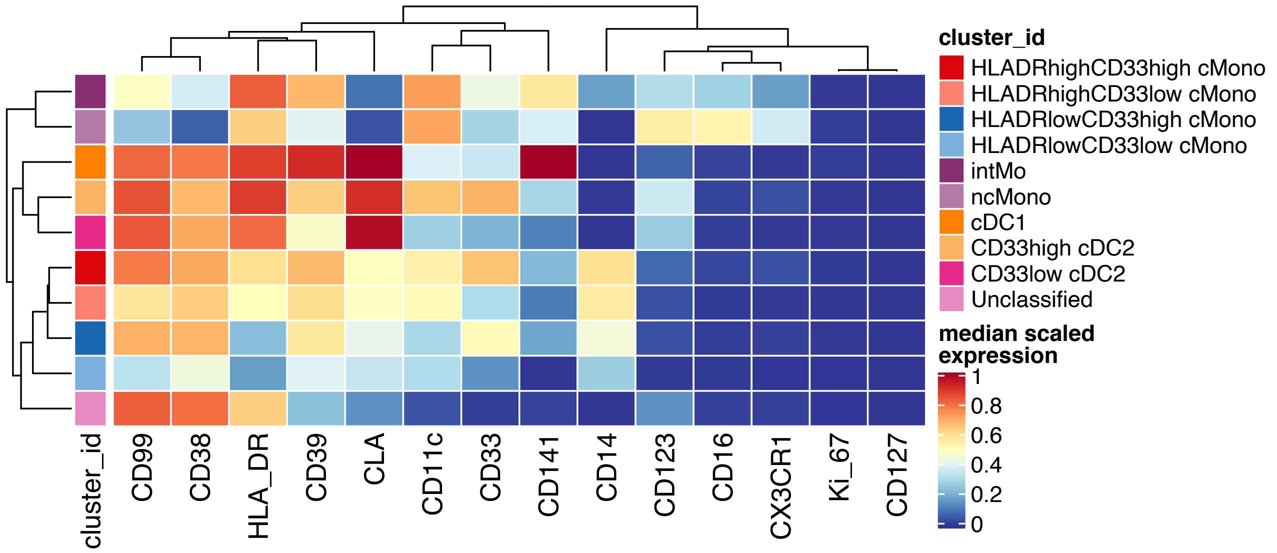


**Figure 5:** Heatmap showing the phenotype of the fine myeloid cell populations described in Figure 3C.

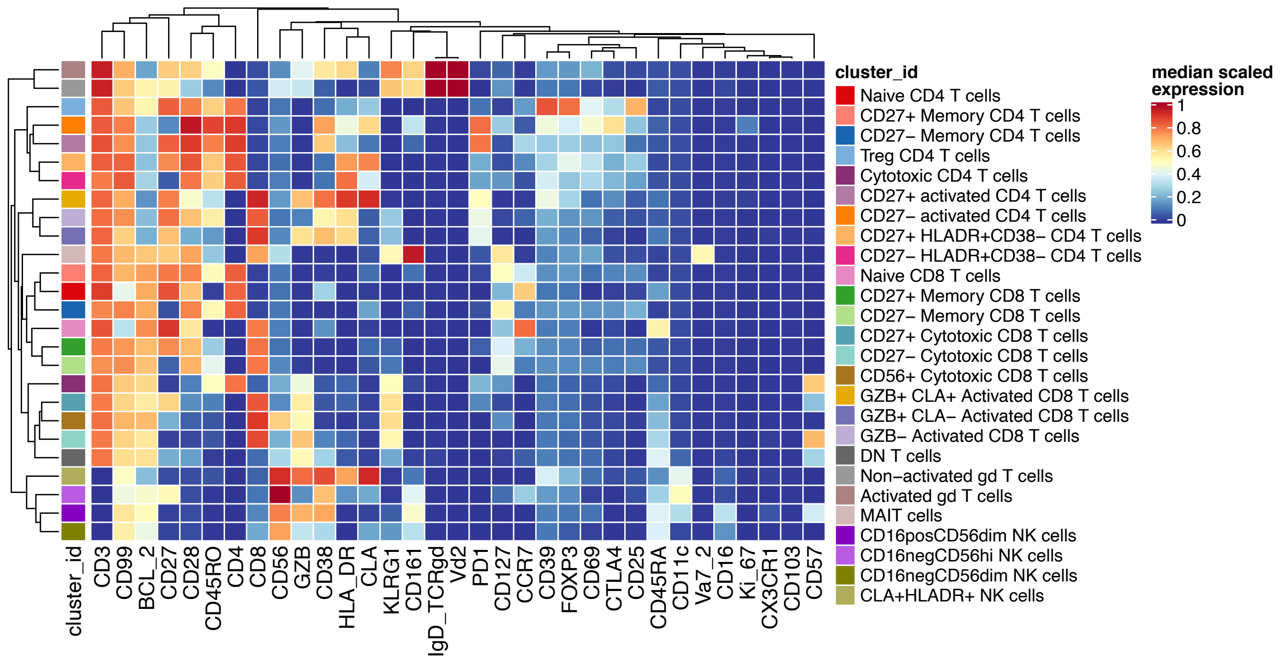


**Figure 6:** Heatmap showing the phenotype of the T/NK cell populations described in Figure 5A.
