## Supplementary material for "A blood atlas of COVID-19 defines hallmarks of disease severity and specificity": Methods S3

#### 2. Patient recruitment and cohorts

Alexander Mentzer, Mark Ainsworth, Carolina V Arancibia-Cárcamo, J. Kenneth Baillie, Archana Bashyal, Sally Beer, Tihana Bicanic, Andrew Brent, Neil Davidson, Julie Dequaire, Susanna J Dunachie, Alexis Espinosa, Rory Fairhead, Shayan Fassih, John Frater, Maria Fernandez Mendoza, Thomas Foord, Anastasia Fries, Veronica Gallardo Sanchez, Dominique Georgiou, Charles Hinds, Clare Hird, Paula Hutton, Katie Jeffery, David Kim, Paul Klenerman, Julian Knight, Andrew Kwok, Teresa Lockett, Jose Martinez, Philippa Matthews, Stuart McKechnie, Denise O'Donnell, Alex Novak, Ian Pavord, Elena Perez, Thomas Ritter, Matthew Rowland, Malcolm G Semple, Donal Skelly, Alberto Sobrinodiaz, Lizzie Stafford,

Adan Taylor, Hannah Thraves, Lance Turtle, Holm Uhlig, Rebecca K. Young, Jonathan Youngs

COVID-19 patients (Oxford)
Patient recruitment, consent and phenotyping by Alexander Mentzer, Mark Ainsworth, Archana Bashyal, Sally Beer, Neil Davidson, Julie Dequaire, Alexis Espinosa, Rory Fairhead, Shayan Fassih, Maria Fernandez Mendoza, Thomas Foord , Anastasia Fries, Veronica Gallardo Sanchez, Dominique Georgiou, Clare Hird, Paula Hutton, David Kim, Andrew Kwok, Teresa Lockett, Jose Martinez, Elena Perez, Thomas Ritter, Alberto Sobrinodiaz, Adan Taylor, Hannah Thraves, Rebecca K Young. Patients were recruited as part of Sepsis Immunomics (Julian Knight, Chief Investigator for Sepsis Immunomics; ISARIC/WHO Clinical Characterisation Protocol for Severe Emerging Infections (J. Kenneth Baillie, Paul Klenerman, Malcolm G Semple, Lance Turtle); OPTIC study (Carolina V Arancibia-Cárcamo, Christopher Conlon, Susanna J Dunachie, John Frater, Katie Jeffery, Philippa Matthews, Denise O'Donnell, Donal Skelly, Lizzie Stafford,).

Influenza patients and COVID-19 patients (non-Oxford)
Patients were recruited by Jonathan Youngs, members of the AspiFlu team and clinical research nurses. Supervised by Tihana Bicanic (Chief Investigator for AspiFlu).

Sepsis patients
Sepsis patients were recruited by Andrew Kwok, Sally Beer and the clinical research nurse team at Oxford University Hospitals Trust, with supervision and input by Julian Knight (Chief Investigator for Sepsis Immunomics), Stuart McKechnie (local PI), Charles Hinds, Alexander Mentzer, Andrew Brent, Alex Novak and Matthew Rowland.

Healthy volunteers
Healthy volunteers were recruited and sampled by Andrew Kwok with support from Alexander Mentzer, Andrew Brown and Alice Allcock. Supervised by Julian Knight as Chief Investigator (healthy volunteer study).

Supervision of recruitment and cohorts
Overall supervision and direction by Alexander Mentzer, with input from Julian Knight and Andrew Kwok.

#### 3. Sample processing and extraction

Alexander Mentzer, Alice Allcock, Chris Allan, Amy Beveridge, Sagida Bibi, Tihana Bicanic, Luke Blackwell, Andrew Brown, David Buck, Susana Camara, Elizabeth Clutterbuck, Fabiola Curion, Christina Dold, Tao Dong, Sally Felle, Angie Green, Jennifer Hill, Elizabeth Jones, Julian Knight, Andrew Kwok, Angela Lee, Aline Linder, Lorne Lonie, Maria Lopopolo, Spyridoula Marinou, Adam Mead, Yanchun Peng, Andrew J Pollard, Laura Silva-Reyes, Christine Rollier, Giuseppe Scozzafava, Hubert Slawinski, Lisa Stockdale, Marije Verheul, Lorna Witty Katherine Wray, Zixi Yin, Jonathan Youngs

Sample processing
Preparation of samples, separation and cryopreservation of peripheral blood mononuclear cells Chris Allan, Amy Beveridge, Sagida Bibi, Luke Blackwell, Susana Camara, Elizabeth Clutterbuck, Christina Dold, Sally Felle, Angie Green, Jennifer Hill, Elizabeth Jones, Andrew Kwok, Aline Linder, Spyridoula Marinou, Yanchun Peng, Laura Silva-Reyes, Christina Simoglou Karali, Christine Rollier, Marie Strickland, Lisa Stockdale, Marije Verheul, Katherine Wray, Zixi Yin, Jonathan Youngs

Purification of genomic DNA and RNA
Alice Allcock, Andrew Brown, Andrew Kwok, Giuseppe Scozzafava.

Sample management, curation and cataloguing
Andrew Kwok, Alexander Mentzer, Giuseppe Scozzafava.

Oxford Genomics Centre (OGC) core sequencing facility
Fabiola Curion, Angela Lee, Maria Lopopolo, Lorne Lonie, Hubert Slawinski, Lorna Witty

Supervision
Supervised in specific labs by Tihana Bicanic (London), David Buck (OGC), Tao Dong (WIMM), Julian Knight (WHG), Andrew J Pollard (CCVTM); and with overall direction by Alexander Mentzer and Julian Knight.

### 4. Clinical phenotyping

Alexander Mentzer, J. Kenneth Baillie, Eleanor Barnes, Tihana Bicanic, Calliope Dendrou, Lucy Garner, Ling-Pei Ho, Julian Knight, Andrew Kwok, Luke Jostins, T. Phuong Quan, Fabian Ruehle, Stephen Sansom, Malcolm G Semple, Alberto Santos Delgado, Lance Turtle, Jonathan Youngs

### 5. Flow and mass cytometry

Giorgio Napolitani, Mariolina Salio, Claudia Monaco, Irina Udalova, David J Ahern, Yasemin-Xiomara Zurke, Lea Dib, Rachel Etherington, Julian Knight, Prathiba Kurupati, Andrew Kwok, Michalina Mazurczyk, Graham Ogg, Inhye Park, Mariana Periera Pinho, Emmanouela Repapi, Lihui Wang

#### *Mass Cytometry Whole Blood (figs 1, 3, 4, 5)*

Experimental design and data generation
Andrew Kwok developed the antibody panel with help from Rachel Etherington and Michalina Mazurczyk and input from Giorgio Napolitani. Andrew Kwok processed the samples and performed the staining with help from Giorgio Napolitani. Rachel Etherington and Giorgio Napolitani acquired the CyTOF samples on the Helios Mass Cytometer.

Data analysis
Giorgio Napolitani performed manual de-barcoding and initial processing. Emmanouela Repapi performed processing and normalization of the data. Mariana Pereira Pino, and Emmanouela Repapi performed overall analysis with inputs from Giorgio Napolitani and Andrew Kwok. Mariana Pereira Pino performed cluster annotation and manual gating analysis with inputs from Giorgio Napolitani. Emmanouela Repapi performed the statistical analysis.

Data integration:
Emmanouela Repapi performed the CITE-Seq Mass Cytometry data integration.

Supervision
Giorgio Napolitani supervised the experimental design, data generation and data analysis with inputs from Julian Knight.

#### *Mass Cytometry- Myeloid panel (fig 2)*

Experimental design and data generation
David J Ahern developed the myeloid CyTOF panel with input from Claudia Monaco, Irina Udalova, Lea Dib, Nikolaos Sousos and Lihui Wang. David J Ahern, Lea Dib  and Inhye Park performed the staining.

Data analysis
Yasemin-Xiomara Zurke and David J Ahern performed the CyTOF analysis and clustering.

Supervision
Claudia Monaco and Irina Udalova supervised the data analysis and performed the interpretation of the data.

#### *Multi Colour Flow Cytometry*

Data generation
Mariolina Salio developed and tested the flow cytometry panels. Mariolina Salio processed, stained and acquired the samples for Multicolour Flow Cytometry and did the data acquisition with help from Giorgio Napolitani.

Flow Cytometry Data Analysis
Mariana Pereira Pino and Mariolina Salio performed data analysis with inputs from Giorgio Napolitani.

Supervision
Mariolina Salio supervised the experimental design, data generation and data analysis.

### 6. Whole blood total RNAseq

Alexander Mentzer, Moustafa Attar, Katie L Burnham, Emma E Davenport, James Docker, Clementine Geeves, Charles Hinds, Julian Knight, Andrew Kwok, Angela Lee, Daniel O'Connor, Santiago Revale, Justin Whalley

Data generation
Data was generated by Andrew Kwok, Moustafa Attar, James Docker, Clementine Geeves.

Data analysis
Data analysis was performed by Katie L Burnham, Andrew Kwok, Daniel O'Connor (WGCNA), Santiago Revale, Justin Whalley.

Supervision
Supervised by Alexander Mentzer and Julian Knight with Emma E Davenport and Charles Hinds.

### 7. 10X CITEseq and repertoire

Stephen Sansom, Calliope Dendrou, Luke Jostins, Julian Knight, Rachael Bashford-Rogers, Benjamin Fairfax, Dominik Aschenbrenner, Moustafa Attar, Paul Bowness, Adam P Cribbs, Fabiola Curion, Tao Dong, Ricardo Ferreira, Lucy Garner, Maria Gomez Vazquez, Anna James-Bott, Ashwin Jainarayanan, Kathrin Jansen, Paul Klenerman, Piyush Kumar Sharma, Andrew Kwok, Angela Lee, Adam Mead, Alexander Mentzer, Ruddy Montadon, Giorgio Napolitani, Isar Nassiri, Lauren Overend, Yanchun Peng, Frank Penkava, Bethan Psaila, Emmanouela Repapi, Santiago Revale, Jean-Baptiste Richard, Charlotte Rich-Griffin, Hubert Slawinski, Bo Sun, Chelsea Taylor, Supat Thongjuea, Orion Tong, Felicia Anna Tucci, Alexandru Voda, Guanlin Wang, Robert Watson, Hing Yuen Yeung, Yasemin-Xiomara Zurke

Experimental design and data generation

Fabiola Curion, Julian Knight, Andrew Kwok, Luke Jostins, Alexander Mentzer, Rachael Bashford-Rogers and Stephen Sansom contributed to sample selection and experimental design. Sample preparation, cell QC and single-cell capture was performed by Andrew Kwok, Hubert Slawinski and Moustafa Attar.

Analysis of the gene expression (GEX) data

Quantitation and genetic demultiplexing was performed by Fabiola Curion and Santiago Revale. Kathrin Jansen computed velocity matrices. Pre-processing and QC assessment of the GEX data was performed by Bo Sun, Fabiola Curion, Kathrin Jansen and Maria Gomez Vasquez with input from and Lauren Overend. Databasing of cell metrics was performed by Adam P Cribbs with help from Stephen Sansom. Extraction of matrices for downstream analysis was performed by Stephen Sansom. Alignment of the data was performed by Kathrin Jansen with advice from Supat Thongjuea and Stephen Sansom. Clustering, marker and pathway analysis was performed by Stephen Sansom. Initial clustering results were independently confirmed by Charlotte Rich-Griffin and Calliope Dendrou. Analysis of viral transcripts was performed by Adam P Cribbs, Anna James-Bott and Lauren Overend.

Analysis of the antibody-derived tag (ADT) data

Analysis of the ADT data was performed by Fabiola Curion and Giorgio Napolitani with help from Emmanouela Repapi and input from Calliope Dendrou and Stephen Sansom.

Analysis of the CITE-seq and bulk BCR and TCR data

Santiago Revale performed CITE-seq VDJ pre-processing. Bo Sun performed single cell BCR repertoire analysis. Single cell TCR repertoire analysis with exploration of clonal size, integration with gene expression and Kmer usage was performed by Rob Watson, Orion Tong, Chelsea Taylor and Piyush Kumar Sharma. Felicia Anna Tucci performed the bulk BCR and TCR repertoire amplification, library preparation and sample handling. Rachael Bashford-Rogers performed the bulk BCR and TCR repertoire analysis. Repertoire team members who have provided guidance and feedback to the analyses and manuscript were Lauren Overend, Tao Dong, Charlotte Rich-Griffin, Yanchun Peng, Isar Nassiri, Paul Bowness, and Frank Penkava.

Multi-modal annotation of the CITE-seq dataset

Annotation of B-cell subpopulations was performed by Bo Sun, Calliope Dendrou and Charlotte Rich-Griffin with input from Felicia Anna Tucci, Hing Yuen Yeung and Rachael Bashford-Rogers. Annotation of T/NK subpopulations was performed by Calliope Dendrou, Lucy Garner, Ricardo Ferreira, Giorgio Napolitani and Stephen Sansom with input from Paul Klenerman, Andrew Kwok and Hing Yuen Yeung. Annotation of the myeloid subsets was performed by Andrew Kwok and Stephen Sansom. Annotation of HSC and platelets was performed by Stephen Sansom and Calliope Dendrou with input from Bethan Psaila. Annotation of doublet populations was performed by Fabiola Curion and Kathrin Jansen. Initial marker visualisation was performed by Charlotte Rich-Griffin and Fabiola Curion. Curation of annotations was performed by Stephen Sansom with review from Andrew Kwok, Lucy Garner and Calliope Dendrou. Manuscript figures were prepared by Stephen Sansom.

Composition analysis

Composition PCA and association testing was performed by Lucy Garner and Charlotte Rich-Griffin with input from Calliope Dendrou. Comparison of multiple differential abundance analysis methods and final analyses were performed by Charlotte Rich-Griffin and Lucy Garner with input from Calliope Dendrou. Clinical variable selection and covariate analysis were performed by Lucy Garner, Charlotte Rich-Griffin and Calliope Dendrou. Calliope

Dendrou interpreted the results and prepared final manuscript figures with input from Charlotte Rich-Griffin, Julian Knight and Lucy Garner.

Pseudobulk analysis

Initial pseudobulk counts were generated by Stephen Sansom and assessed by Fabiola Curion and Luke Jostins. Normalized pseudobulk counts, principal components and UMAP co-ordinates were generated by Luke Jostins with advice from Stephen Sansom and Fabiola Curion. Alexandru Voda performed differential expression analyses with input from Luke Jostins and Stephen Sansom. Luke Jostins collected pathway gene lists with input from Lucy Garner, Adam P Cribbs, Andrew Kwok, Jean-Baptiste Richard and Dominik Aschenbrenner, and carried out gene set enrichment analyses. Clinical covariate analysis was carried out by Luke Jostins with input from Lucy Garner, Charlotte Rich-Griffin and Calliope Dendrou. Luke Jostins interpreted results and generated manuscript figures with input from Stephen Sansom, Calliope Dendrou and Julian Knight.

WGCNA and network analysis

Lucy Garner performed initial exploratory analyses. WGCNA network analysis was performed by Charlotte Rich-Griffin, Fabiola Curion and Stephen Sansom. Charlotte Rich-Griffin and Fabiola Curion identified sets of modules common to multiple cell types and characterised their relationship with the type I IFN signalling response. Stephen Sansom tested module associations, performed additional analyses and interpreted the results with input from Calliope Dendrou, Charlotte Rich-Griffin, Fabiola Curion, Andrew Kwok and Julian Knight. Charlotte Rich-Griffin, Fabiola Curion and Stephen Sansom prepared the figures.

Cell-cell interaction analysis

Cell-cell interaction was performed by Guanlin Wang with input from Supat Thongjuea.

Supervision

Luke Jostins, Julian Knight, Rachael Bashford-Rogers and Stephen Sansom supervised the experimental design. Rachael Bashford-Rogers supervised the BCR and repertoire analysis. Benjamin Fairfax supervised the TCR analysis. Luke Jostins supervised the pseudobulk analysis and provided expert statistical guidance. Calliope Dendrou supervised the composition analysis and co-supervised the multimodal annotation. Supat Thongjuea co-supervised the alignment work. Supat Thongjuea and Adam Mead supervised the cell-cell interaction analysis. Stephen Sansom supervised the GEX pre-processing, alignment and clustering, the multimodal annotation and the WGCNA network analysis. Overall supervision and facilitation of work by Stephen Sansom with Calliope Dendrou (CITEseq) and Rachael Bashford-Rogers (repertoire).

### 8. ATACseq

Tatjana Sauka-Spengler, Ivan Candido Ferreira, Martyna Lukoseviciute, Moustafa Attar, Santiago Revale, Julian Knight, Andrew Kwok, Angela Lee, Hubert Slawinski, Michael Weinberger

Data generation
Data was generated by Andrew Kwok, Moustafa Attar and Hubert Slawinski.

Data analysis
Data analysis was performed by Ivan Candido Ferreira, Martyna Lukoseviciute, Santiago Revale, Andrew Kwok, Tatjana Sauka-Spengler and Michael Weinberger.

Supervision
Tatjana Sauka-Spengler and Julian Knight

### 9. Genetics

Luke Jostins, Amanda Chong, Damien Downes, Chris Eijsbouts, Ben Fairfax, Jim Hughes, Julian Knight, Alexander Mentzer, Yuxin Mi, Isar Nassiri, Ron Schwessinger, Ping Zhang

Alexander Mentzer and Andrew Kwok prepared samples for genotyping, Ben Hollis and Luke Jostins carried out quality control, principal component analysis and imputation. Amanda Chong carried out association analysis. Damien Downes, Chris Eijsbouts, LJ, Yuxin Mi, Isar Nassiri, Ron Schwessinger and Ping Zhang carried out integration with other data modalities. Calliope Dendrou, Ben Fairfax, Jim Hughes, Luke Jostins, Julian Knight, Alexander Mentzer and Stephen Sansom supervised specific analyses, and Luke Jostins provided overall supervision.

### 10. Proteomics (mass spectrometry)

Roman Fischer, Alberto Santos Delgado, Georgina Berridge, Philip Charles, Simon Davis, Raphael Heilig, Svenja Hester, Julian Knight, Yuxin Mi, Darragh O'Brien, Iolanda Vendrell

Experimental design
Iolanda Vendrell, Georgina Berridge and Roman Fischer developed the experimental design with inputs from Philip Charles, Simon Davis, Raphael Heilig.

Data generation
Georgina Berridge, Iolanda Vendrell, Svenja Hester, Darragh O'Brien and Roman Fischer processed samples. Data was generated by Georgina Berridge, Iolanda Vendrell, Simon Davis and Raphael Heilig.

Data analysis
Roman Fischer and Philip Charles performed identification and quantitation. Yuxin Mi and Alberto Santos Delgado performed downstream analysis and interpretation.

Supervision
Iolanda Vendrell supervised the sample processing, Roman Fischer supervised the team.

Acknowledgements
The Proteomics team is grateful for the help of Zuzana Bencokova, Darren Blase and Benedikt Kessler (all TDI) for providing the environment for safe sample handling and analysis. We also thank Alexey Nesvizhskii and Fengchao Yu (both Department of Pathology. University of Michigan Medical School) for providing help and advice for data processing in Fragpipe.

### 11. Proteomics (Luminex)

Luzheng Xue, Wentao Chen, Yi-Ling Chen, David A. Duncan, Paul Klenerman, Jian Luo, Claudia Monaco, Graham Ogg, Ian Pavord, Irina Udalova

Experimental design
Wentao Chen and Luzheng Xue designed the Luminex assay panels with inputs from Claudia Monaco, Irina Udalova, Ian Pavord, Graham Ogg and Paul Klenerman.

Data generation

Yi-Ling Chen and Jian Luo processed the samples, performed the assays and generated the data.

Data analysis
Yi-Ling Chen, Jian Luo, Wentao Chen and David A. Duncan performed data analysis.

Supervision
Luzheng Xue supervised the experimental design, data generation and data analysis.

### 12. Data Integration

Julian Knight, Rachael Bashford-Rogers, Katie L Burnham, Helen Byrne, Mark Coles, Fabiola Curion, Calli Dendrou, Heather Harrington, Ling-Pei Ho, Renee Hoekzema, Jim Hughes, Matthew Jackson, Luke Jostins, Simon McGowan, Alexander Mentzer, Yuxin Mi, Giorgio Napolitani, Emmanouela Repapi, Fabian Ruehle, Alberto Santos Delgado, Stephen Sansom, Martin Sergeant, Anna Seigal, David Sims, Otto Sumray, Stephen Taylor, Adriana Tomic, Justin Whalley

Clinical phenotyping
Statistical and machine learning analysis by Fabian Ruehle with input and interpretation from Ling-Pei Ho, Julian Knight, Alexander Mentzer and Stephen Sansom.

Integrative analysis of mass cytometery and CITEseq cell composition
Analysis by Emmanouela Repapi and Yasemin-Xiomara Zurke with input from Fabiola Curion, Calli Dendrou, Giorgio Napolitani and Stephen Sansom.

Similarity network fusion and patient subphenotyping
Analysis by Alberto Santos Delgado with input and interpretation from Katie L Burnham, Roman Fischer, Yuxin Mi, Julian Knight.

Machine learning
Fabian Ruehle and Matthew Jackson undertook machine learning analysis of phenotyping and experimental datasets with input from Ling-Pei Ho, Stephen Sansom, Luke Jostins and Julian Knight. Adriana Tomic performed analysis using Sequential Iterative Modeling “Over Night” (SIMON).

Topological data analysis
Otto Sumray and Renee Hoekzema performed the analysis with supervision by Heather Harrington and input from Ling-Pei Ho and Stephen Sansom.

Tensor and matrix decomposition
Data was analysed and interpreted by Justin Whalley, Heather Harrington, Julian Knight, Anna Seigal and Stephen Sansom.

Data mining
Datasets identified and curated by Calli Dendrou with input from Guanlin Wang and Charlotte Rich-Griffin.

Data visualisation
Martin Sergeant, Simon McGowan and Stephen Taylor developed software for data visualisation and hosting through Multi Locus View (MLV), with input from Jim Hughes and Stephen Sansom. Hallmark collation Katie L Burnham, Julian Knight and Yuxin Mi with graphical visualisation (Julian Knight) and interactive visualisation in MLV (Martin Sergeant, Stephen Taylor) with input from Stephen Sansom. COMBAT logo and manuscript artwork Julian Knight.

Supervision
Julian Knight led and facilitated work by the group; Stephen Sansom helped supervise the work.

### 13. Data management

Brian Marsden, Adam P Cribbs, Robert Esnouf, Hai Fang, Hong Harper, Luke Jostins, Julian Knight, Georgina Kerr, Vinod Kumar, Alexander Mentzer, Stephen Sansom, David Sims, Dapeng Wang

Data Management
Hai Fang and Dapeng Wang managed data set registration and curation. Brian Marsden, Vinod Kumar and Hong Harper developed and implemented data management solutions. Alexander Mentzer, Brian Marsden, Julian Knight, Vinod Kumar, Georgina Kerr and Giuseppe Scozzafava implemented and managed processes for sample tracking and management. Brian Marsden and Luke Jostins designed the protocol for capture of project dataset metadata.

Digital Communications
Brian Marsden, Julian Knight, Georgina Kerr, Stephen Sansom and Dapeng Wang set up and managed communication platforms, website and channels for the consortium.

High-performance computing environment and Code management
A bespoke high-performance computational platform was provided by the University of Oxford's Biomedical Research Computing facility under the guidance of Robert Esnouf. Stephen Sansom, Brian Marsden and Hai Fang helped configure and manage use of the platform with input from Adam P Cribbs. Stephen Sansom documented use of the platform, configured the COMBATOxford Team Github and defined coding practice and management policies with input from Adam P Cribbs and David Sims.

Data management operational group
Brian Marsden, Hai Fang, Hong Harper, Julian Knight, Vinod Kumar, Alexander Mentzer, Dapeng Wang

Data Access Committee
Julian Knight, Paul Klenerman, Brian Marsden, Alexander Mentzer, Fiona Powrie, John A Todd

Supervision
Brian Marsden supervised the overall data management strategy and implementation with Julian Knight. Brian Marsden, Stephen Sansom and Georgina Kerr supervised development and use of the computational platform. Stephen Sansom and Adam P Cribbs supervised management of code. Alexander Mentzer supervised sample tracking and management.

### 14. Oversight and Writing

Julian Knight, Rachael Bashford-Rogers, Katie L Burnham, Richard Cornall, Fabiola Curion, Calliope Dendrou, Benjamin Fairfax, Hai Fang, Roman Fischer, Lucy Garner, Heather Harrington, Charles Hinds, Luke Jostins, Paul Klenerman, Andrew Kwok, Brian Marsden, Helen McShane, Alexander Mentzer, Yuxin Mi, Giorgio Napolitani, Graham Ogg, Andrew J Pollard, Fiona Powrie, Charlotte Rich-Griffin, Fabian Ruehle, Stephen Sansom, Alberto Santos-Delgado, Tatjana Sauka-Spengler, Gavin Screaton, Anna Seigal, Martin Sergeant, Stephen Taylor, Bo Sun, Felicia Tucci, John A Todd, Adriana Tomic, Justin Whalley, Luzheng Xue

Writing group
Julian Knight, Rachael Bashford-Rogers, Calliope Dendrou, Ben Fairfax, Roman Fischer, Luke Jostins, Paul Klenerman, Alexander Mentzer, Giorgio Napolitani, Stephen Sansom, Tatjana Sauka-Spengler, John A Todd, Luzheng Xue

Living reports contributing to manuscript
Living reports on individual assay modalities writing leads Rachael Bashford-Rogers and Ben Fairfax (B and T cell repertoire); Katie L Burnham and Alexander Mentzer (whole blood total RNAseq); Fabiola Curion, Charlotte Rich-Griffin, Stephen Sansom (WGCNA scRNAseq); Calliope Dendrou (10X CITEseq composition), Roman Fischer (proteomics mass spectrometry); Heather Harrington (topological data analysis); Luke Jostins (genetics); Alexander Mentzer (clinical phenotyping); Giorgio Napolitani (flow and mass cytometry); Fabian Ruehle (statistical analysis clinical data, machine learning); Martin Sergeant and Stephen Taylor (Multi Experiment Viewer); Stephen Sansom (10X CITEseq pre-processing, alignment and clustering); Adriana Tomic (Sequential Iterative Modeling “Over Night” SIMON); Justin Whalley and Stephen Sansom (Tensor analysis); Luzheng Xue (proteomics luminex). Contributions to reports by team members detailed within teams sections.

Manuscript writing, review and editing
Original draft written by Julian Knight. Review and editing by Rachael Bashford-Rogers, Katie Burnham, Richard Cornall, Calliope Dendrou, Hai Fang, Ben Fairfax, Roman Fischer, Lucy Garner, Heather Harrington, Charles Hinds, Luke Jostins, Andrew Kwok, Paul Klenerman, Alexander Mentzer, Yuxin Mi, Giorgio Napolitani, Fiona Powrie, Stephen Sansom, Alberto Santos Delgado, Anna Seigal, Bo Sun, Felicia Tucci, John A Todd, Luzheng Xue. All other Consortium authors saw, had the opportunity to comment on, and approved the final draft.

Leadership and oversight
Julian Knight led the COMBAT consortium including the work described in this paper. Paul Klenerman led the wider Oxford COVID-19 Immunology Research programme. Stephen Sansom helped lead data analysis and interpretation. COMBAT team leadership and oversight Rachael Bashford-Rogers, Calliope Dendrou, Ben Fairfax, Roman Fischer, Luke Jostins, Julian Knight, Paul Klenerman, Brian Marsden, Alexander Mentzer, Giorgio Napolitani, Stephen Sansom, John A Todd, Tatjana Sauka-Spengler, Luzheng Xue. Georgina Kerr provided project management.

Steering committee
Richard Cornall, Fiona Powrie, Julian Knight, Paul Klenerman, Alexander Mentzer, Helen McShane, Graham Ogg, Andrew J Pollard, Gavin Screaton, John A Todd
