## Supplementary material for "A blood atlas of COVID-19 defines hallmarks of disease severity and specificity": Methods S2

### Methods S2 - Supporting figures and tables for CITE-seq, related to STAR Methods

#### Introduction

This document contains additional figures and tables (as referenced in the STAR methods) that describe the computational analysis and interpretation of the CITE-seq dataset.

#### Experimental design and cell capture

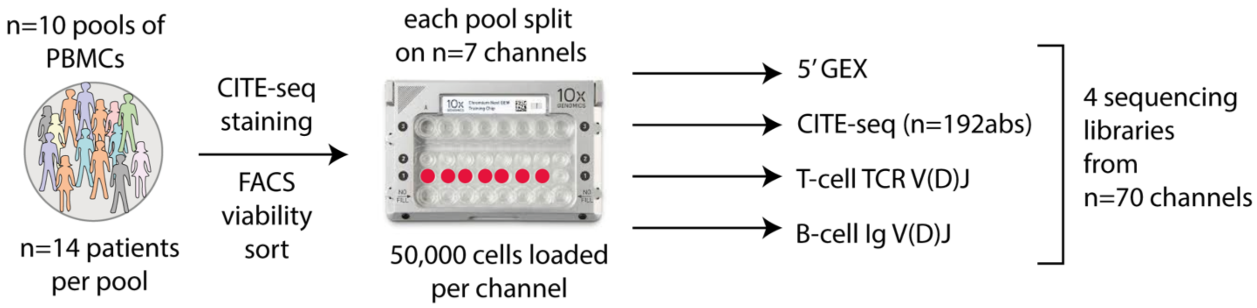

Figure 1: Sample pooling and cell capture strategy

### Multimodal cell identification and annotation

#### Strategy

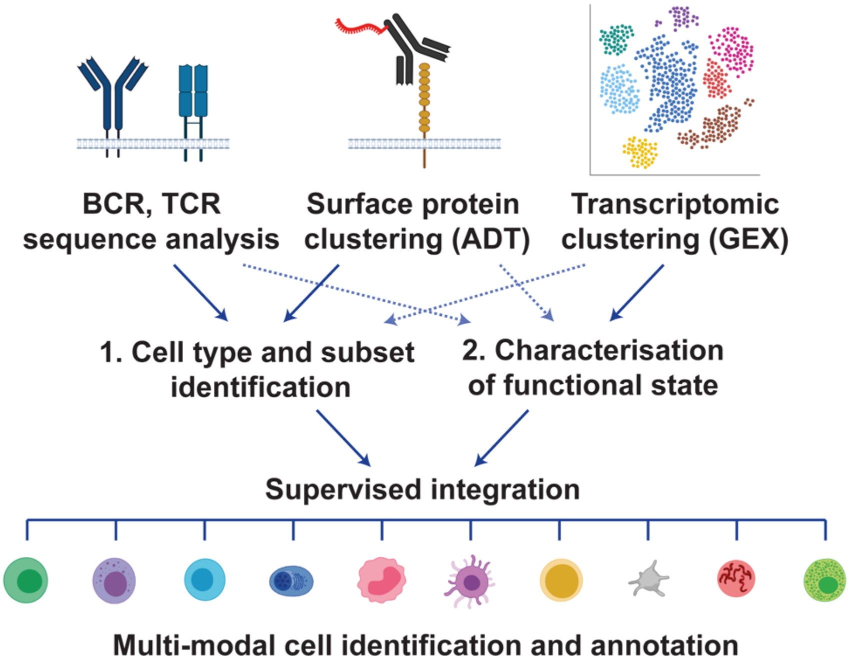

Figure 2: Overview of multi-modal annotation strategy. We first performed separate clustering of surface protein expression, clustering of gene expression and analyses of T and B cell receptor V(D)J sequences. Next, led by expert knowledge of the three feature spaces we prioritised use of ADT surface phenotype for definition of major cell lineages and subsets where definitive marker expression was available. Cell types and subsets were further refined using information from the repertoire and GEX layers, or in the absence of definitive ADT information were identified by GEX cluster phenotype. Finally, the identified cell types and subsets were further divided by inferred functional state based on targeted assessment of information from all three modalities. For example, cell cycle phase was determined by GEX phenotype, T cell memory vs effector status was distinguished using information from both the GEX and ADT layers, while assignment of B cell maturation status involved use of information from all three modalities (including BCR mutational status). Information from all three modalities was used to identify and exclude doublets from downstream analysis.

#### Clustering of gene expression (GEX) data

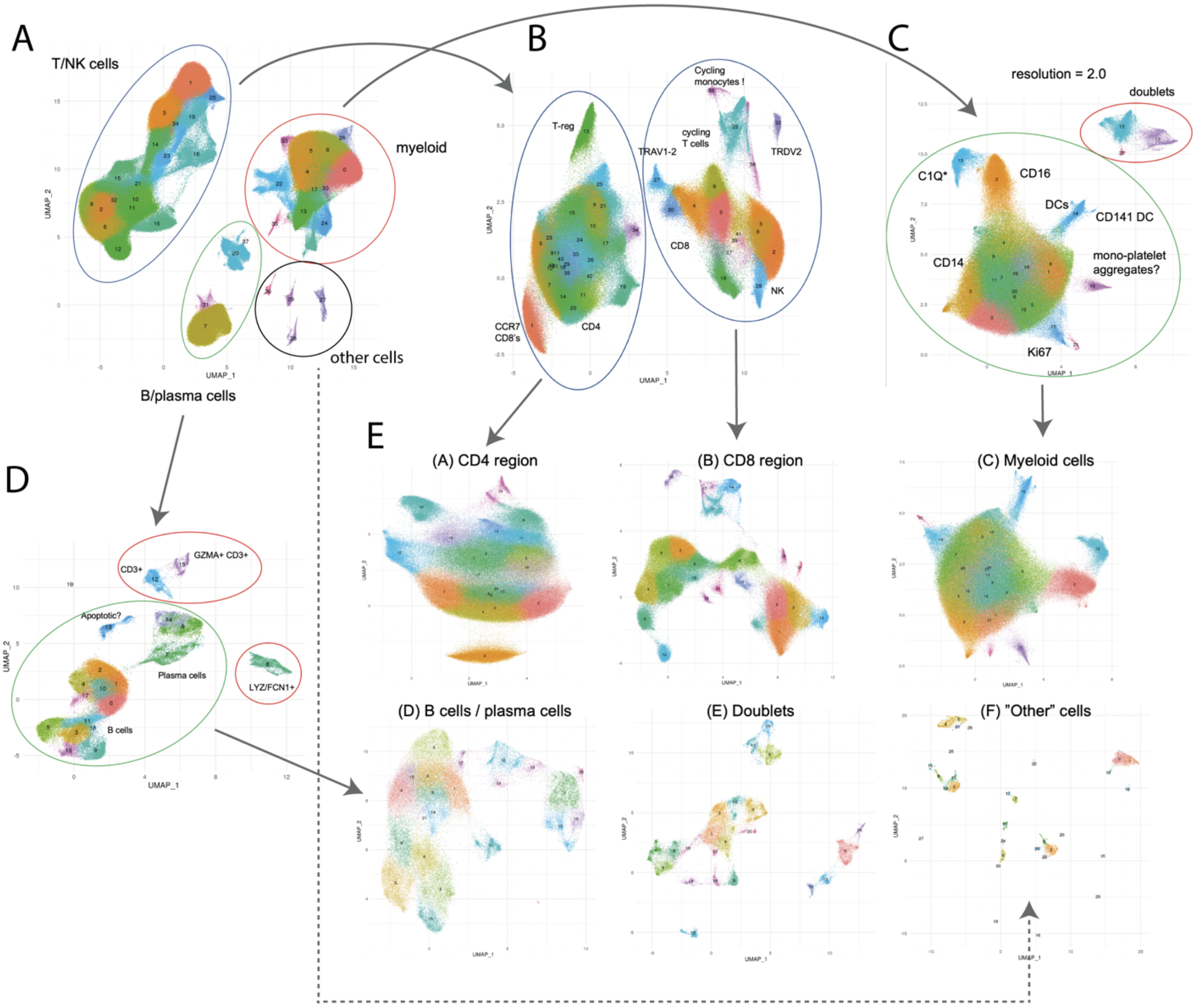

Figure 3: (A) Alignment and clustering of the full dataset (n=836k cells). (B) T/NK cells were extracted (A) and aligned and clustered separately. (C) The myeloid cells were extracted (A) and aligned and clustered separately. (D) The B and plasma cells were extracted (A) and aligned and clustered separately. (E) The final six subsets that were used for annotation: A) CD4-region cells from (B), B) CD8/NK-region cells from (B), C) Myeloid cells (minus doublets) from (C), D) B/plasma cells (minus doublets) from (D), E) doublets from (C) and (E), F) other cells from (A). Cells for each of the final six subsets were aligned and clustered separately.

#### Clustering of surface protein (ADT) data

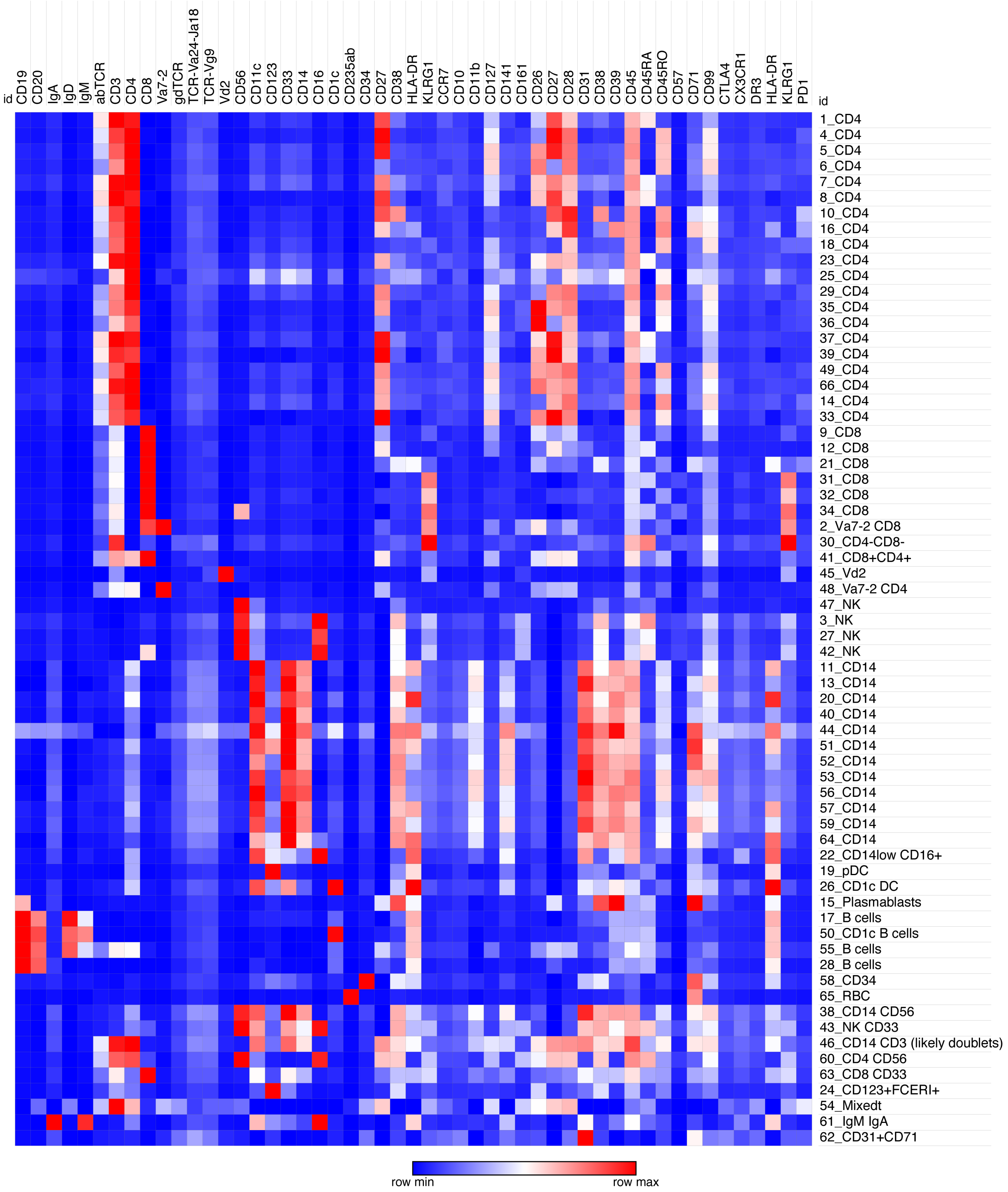

Figure 4: Initial cytosplore clustering of the cells by ADT surface phenotype based on n=192 ADT tags. Columns: selected surface proteins, rows: cluster assignments.

#### Summary of identified PBMC sub-populations

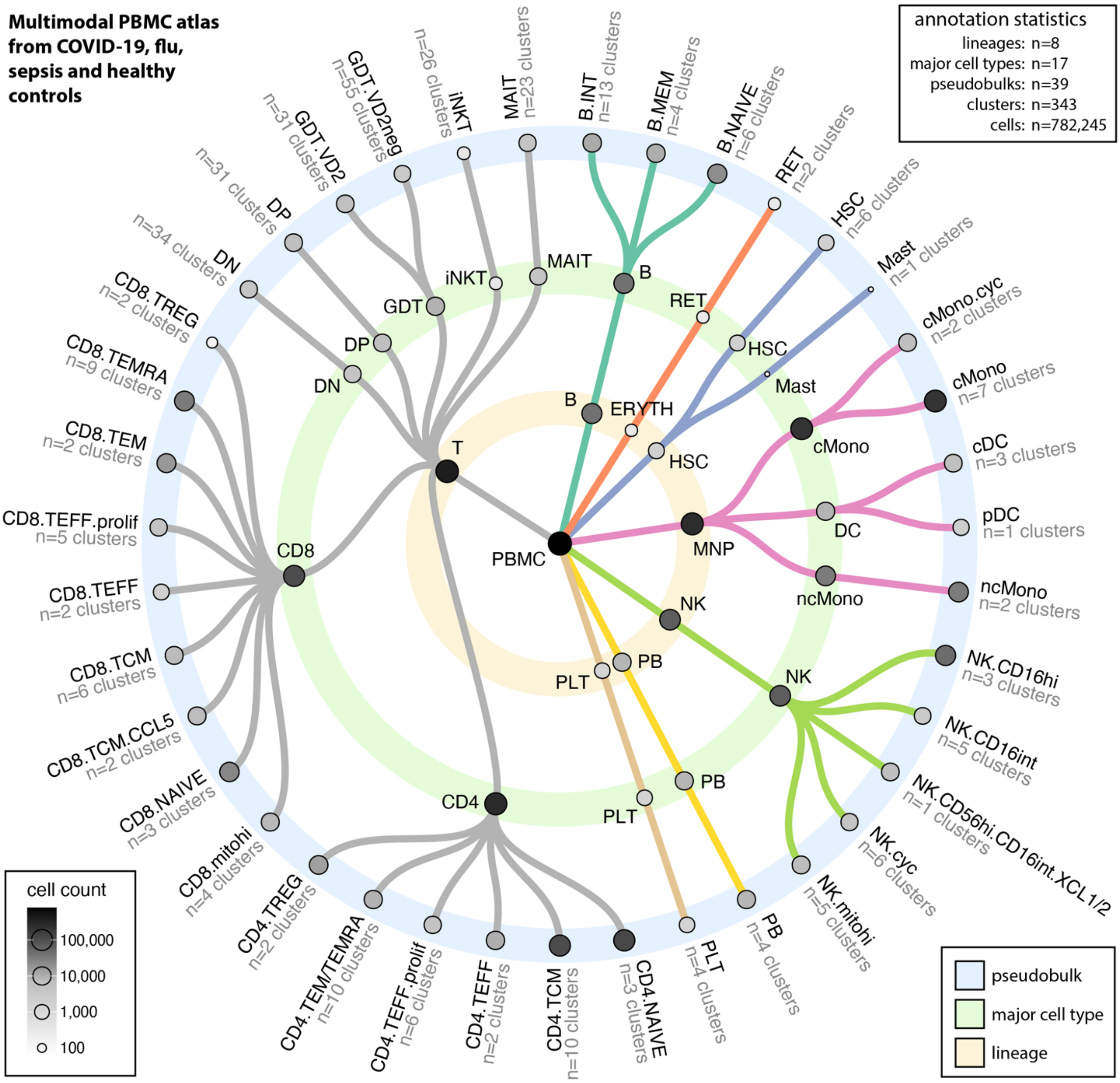

**Abbreviations** B: B cell; cDC: classical dendritic cell; cMono: classical monocytes; cyc: cycling; DC: dendritic cell; DN: CD4/CD8 double negative; DP: CD4/CD8: double positive; ERYTH: erythrocyte; GDT: gamma delta T; hi: high; HSC: haematopoietic stem (and progenitor) cells; iNKT: innate natural killer T; INT: intermediate; MAIT: Mucosal associated invariant T; MEM: memory; mito: mitochondrial; MNP: mononuclear phagocyte; ncMono: non-classical monocyte; NK: natural killer cell; PB: plasmablast; PBMC: peripheral blood mononuclear cell; pDC: plasmacytoid dendritic cell; PLT: platelet; RET: reticulocyte; T: T cell; TCM:T central memory; TEFF: T effector; prolif: proliferating; TEM(RA): T effector memory (CD45RA re-expressing); TREG: T regulatory cell

Figure 5: Summary of cell populations defined following expert curation of the dataset using information from the gene expression (GEX), surface protein (ADT) and repertoire (TCR/immunoglobulin) modalities.

#### Table of T and NK sub-population phenotypes

| Population | Subset | ADT phenotype | TCR phenotype | BCR phenotype | GEX phenotype | Cell count |
|---|---|---|---|---|---|---|
| **CD4** | | singlet, CD3+ CD4+ | singlet | <1% IgG, <20 HC umi | CD4+ | n = 270,003 |
| **CD8** | | singlet, CD3+ CD8+ | singlet | <1% IgG, <20 HC umi | CD8A+ | n = 106,245 |
| **DP** | | singlet, CD3+ CD4+ CD8+ | singlet | <1% IgG, <20 HC umi | | n = 7,102 |
| **DN** | | singlet, CD3+ CD4- CD8- | singlet | <1% IgG, <20 HC umi | | n = 4,644 |
| **MAIT** | **CD8+** | singlet, CD3+ CD8+ Vα7.2+ CD161+ | singlet, TRAV1-2, TRAJ33 , TRAJ12 , TRAJ20 | <1% IgG, <20 HC umi | Various | } n = 3,935 |
| | | singlet, CD3+ CD8+ Vα7.2+ CD161+ | singlet, TCR chains include Mait sequences | <1% IgG, <20 HC umi | MAIT | |
| | **DN** | singlet, CD3+ Vα7.2+ CD161+ | singlet, TRAV1-2, TRAJ33 , TRAJ12 , TRAJ20 | <1% IgG, <20 HC umi | Various- | } n = 639 |
| | | singlet, CD3+ Vα7.2+ CD161+ | singlet, TCR chains include Mait sequences | <1% IgG, <20 HC umi | MAIT | |
| **γδ T** | **Vδ2+ve** | singlet, CD3$^{+ve}$ Vδ2+ Vγ9+ | singlet | <1% IgG, <20 HC umi | TRDV2+ | n = 6,239 |
| | **Vδ2-ve Vγ9-ve** | singlet, CD3+ TCRγδ+ | singlet | <1% IgG, <20 HC umi | TRDV1+ (TRDV3+) | n = 2,238 |
| | **Vδ2-ve Vγ9+ve** | singlet, CD3+ Vδ2- Vγ9+ | singlet | <1% IgG, <20 HC umi | TRDV1+ (TRDV3+) | n = 1,227 |
| **iNKT** | **true** | singlet, Vα24Jα18+ | singlet, TRAV10, TRAJ18 | <1% IgG, <20 HC umi | | n = 293 |
| | **ADT** | singlet, Vα24Jα18+ | singlet, TRAV10, TRAJ18 + additional V/J chains | <1% IgG, <20 HC umi | | n = 93 |
| **NK** | | singlet, NCAM1+ | <50 TCRA & TCRB reads | <1% IgG, <20 HC umi | NCAM1+ | n = 69,997 |

Table 1: Definition of T and NK cell identities. red: identifying features, green: additional criteria, blue: confirmatory observations

#### CD4 and CD8 T cell sub-popuations

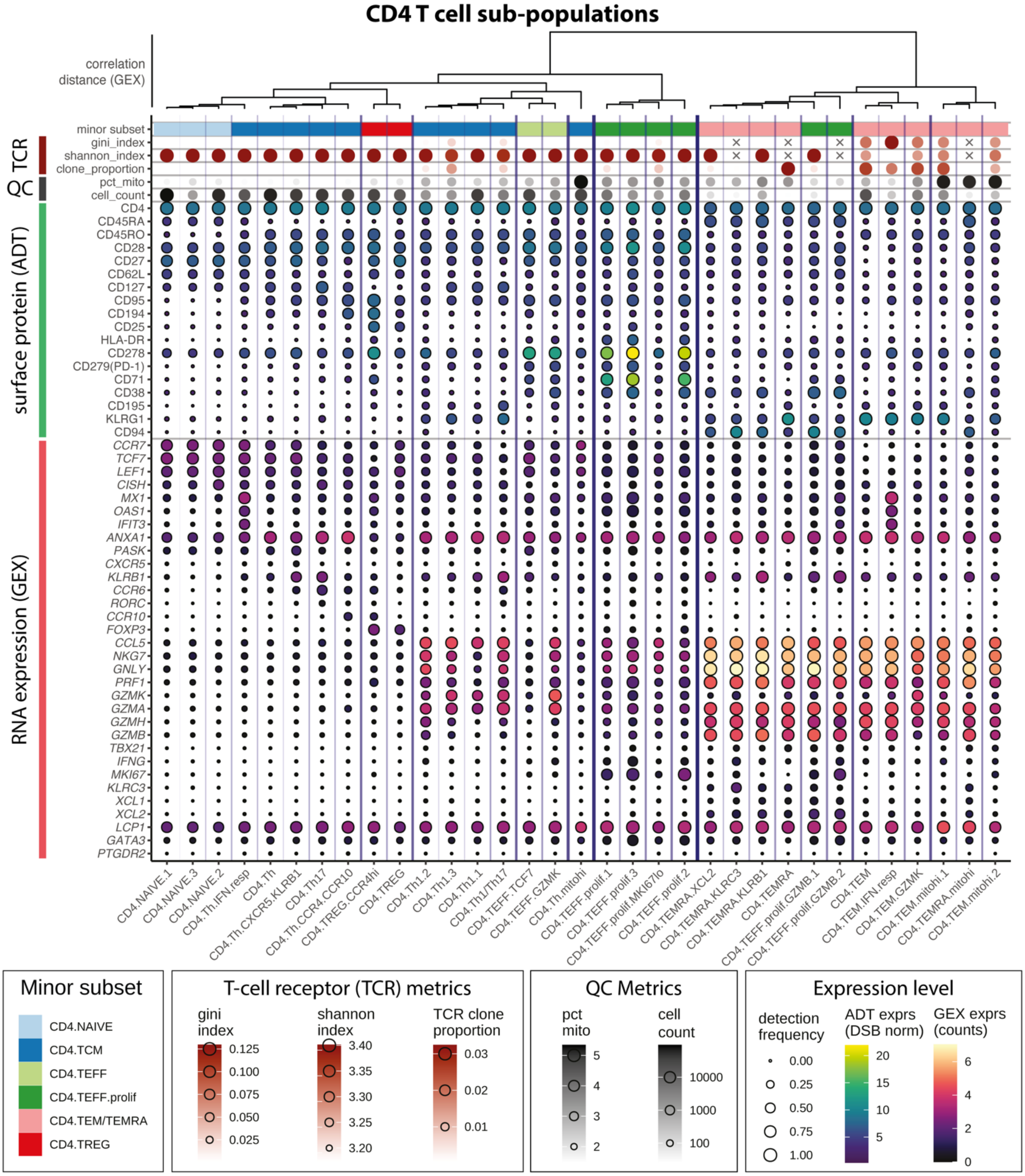

**Abbreviations** hi: high; IFN: interferon; lo: low; mito: mitochondrial; prolif: proliferating; resp: responsive; TEFF: T effector; TEM: T effector memory; TEMRA: CD45RA+ T effector memory; Th: T-helper; TREG: regulatory T cell. **Symbols** x: NA.

Figure 6: CD4 T-cell sub-populations

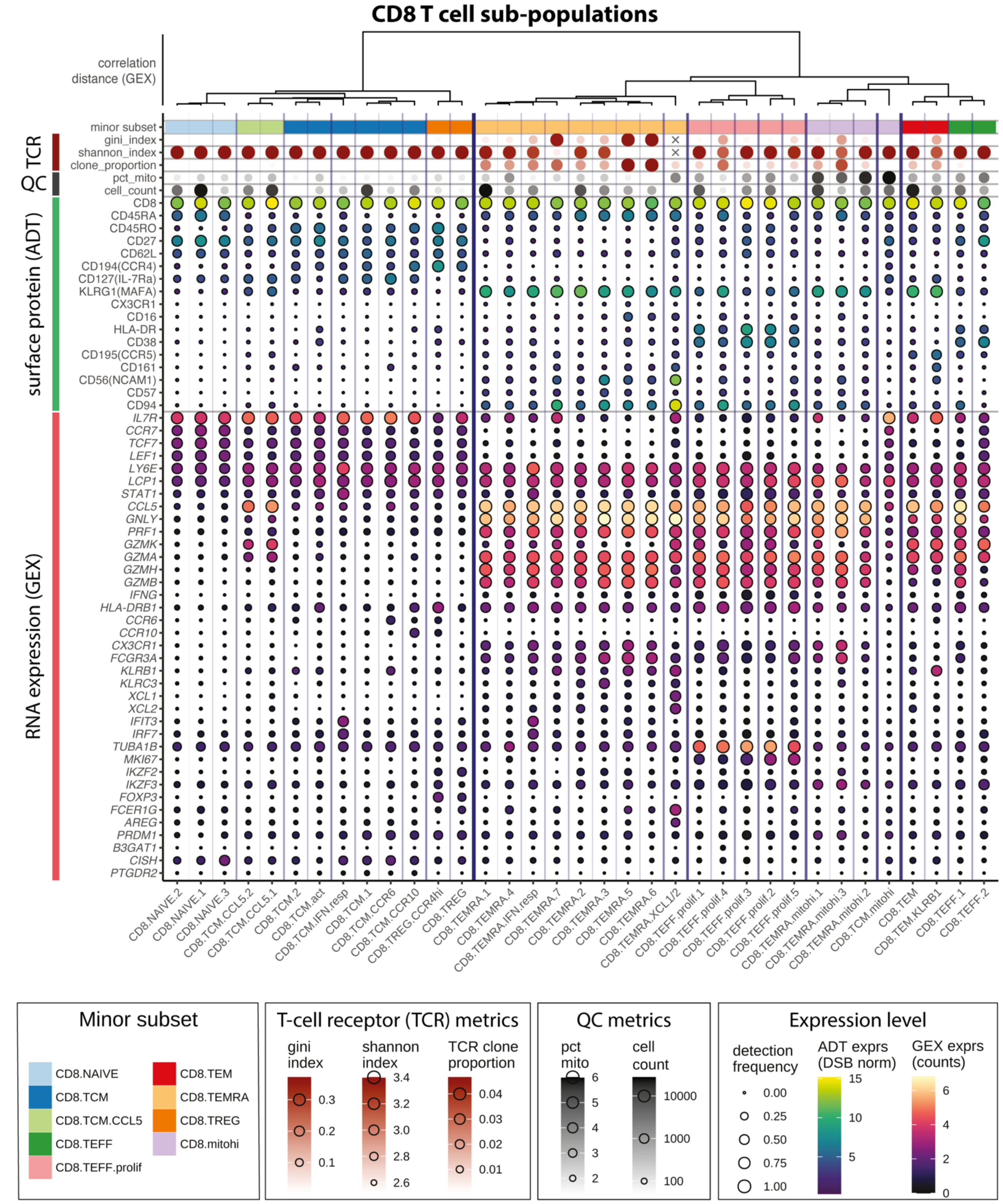

**Abbreviations** act: activated; hi: high; IFN: interferon; lo: low; mito: mitochondrial; prolif: proliferating; resp: responsive; TCM: T central memory; TEFF: T effector; TEM: T effector memory; TEMRA: CD45RA+ T effector memory; Th: T-helper; TREG: regulatory T cell. **Symbols**: x: NA.

Figure 7: CD8 T-cell populations

#### Double positive and negative T cell sub-populations

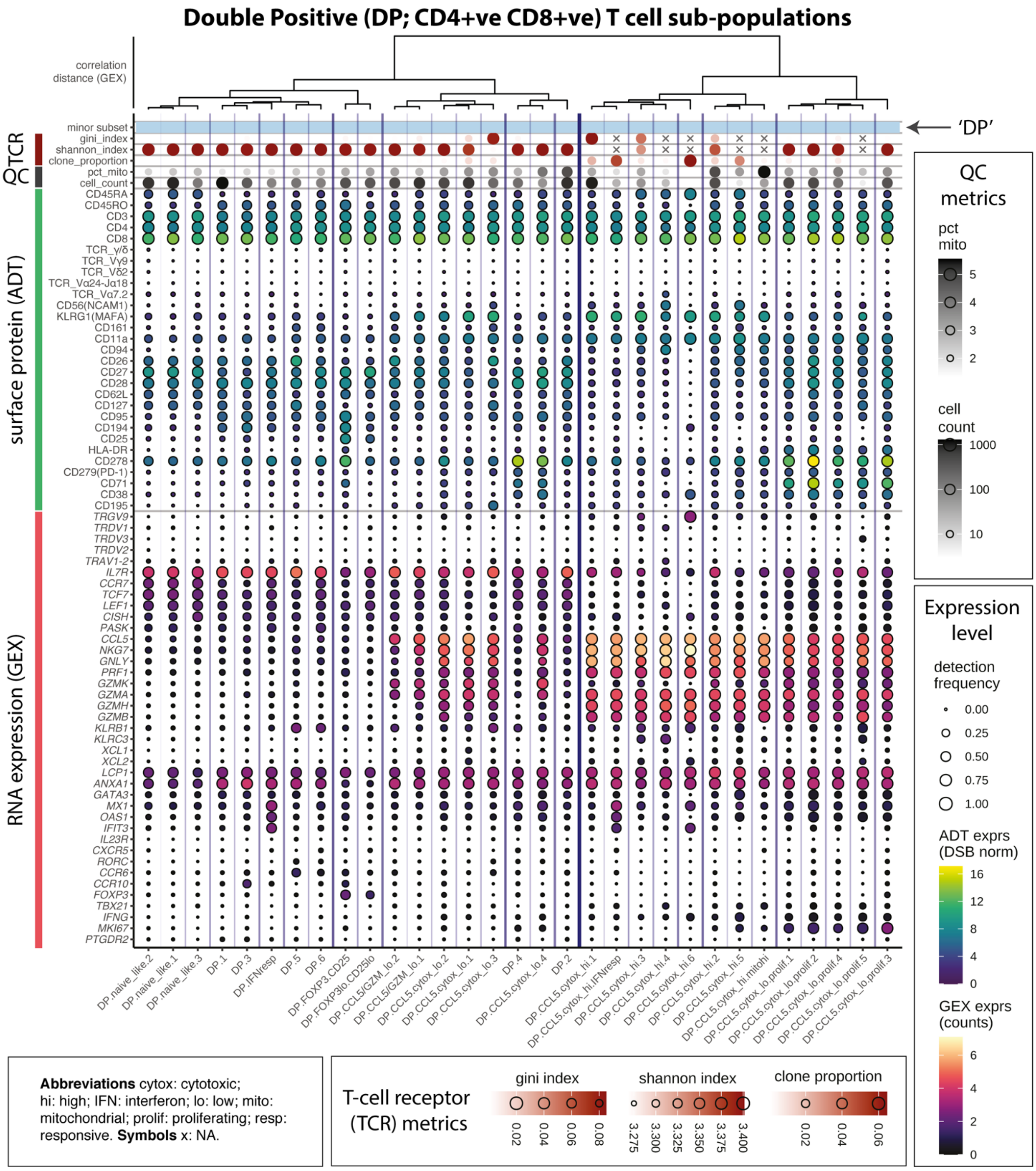

Figure 8: Double positive (DP) T-cell populations

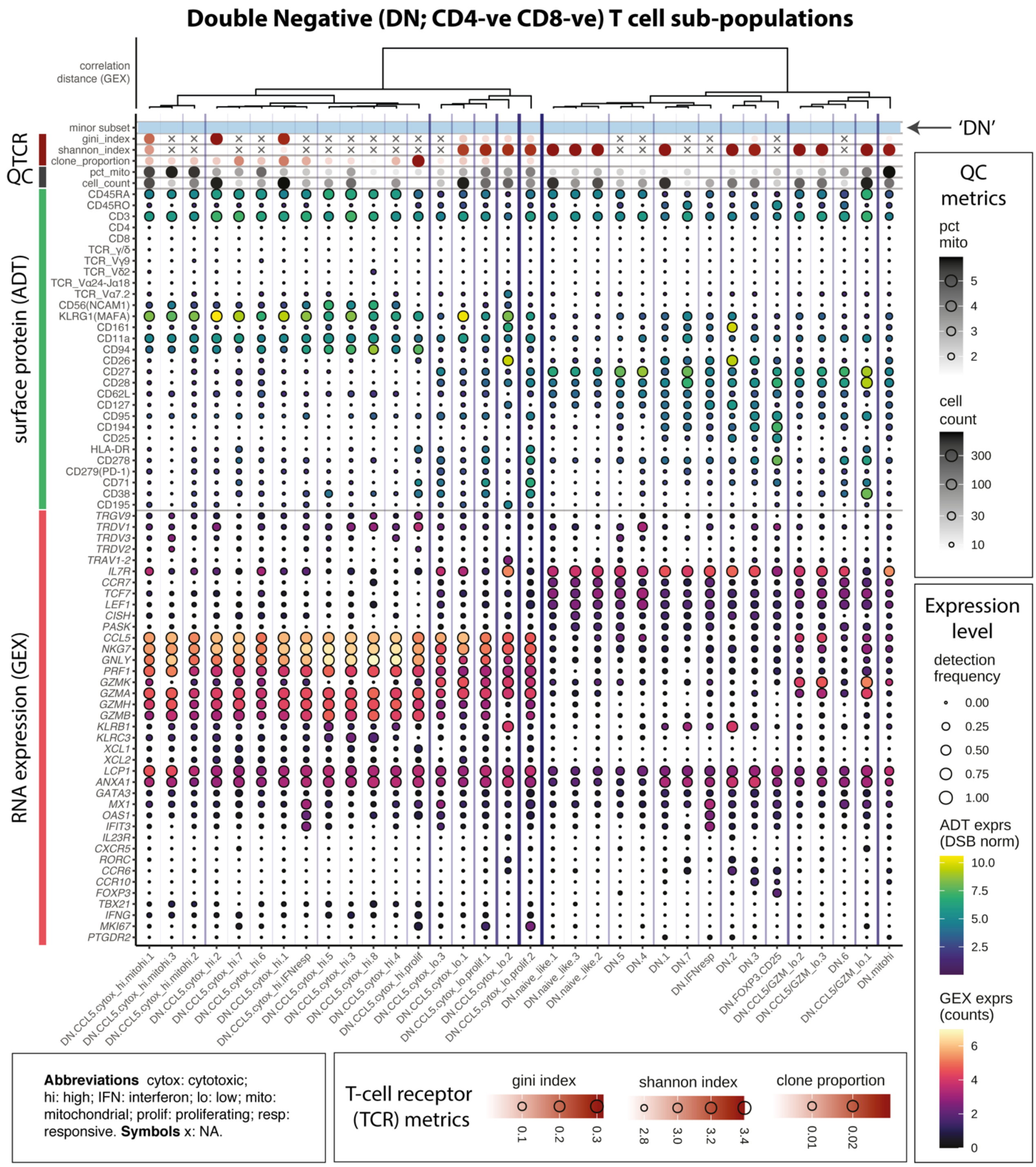

Figure 9: Double negative (DN) T-cell populations

#### MAIT cell populations

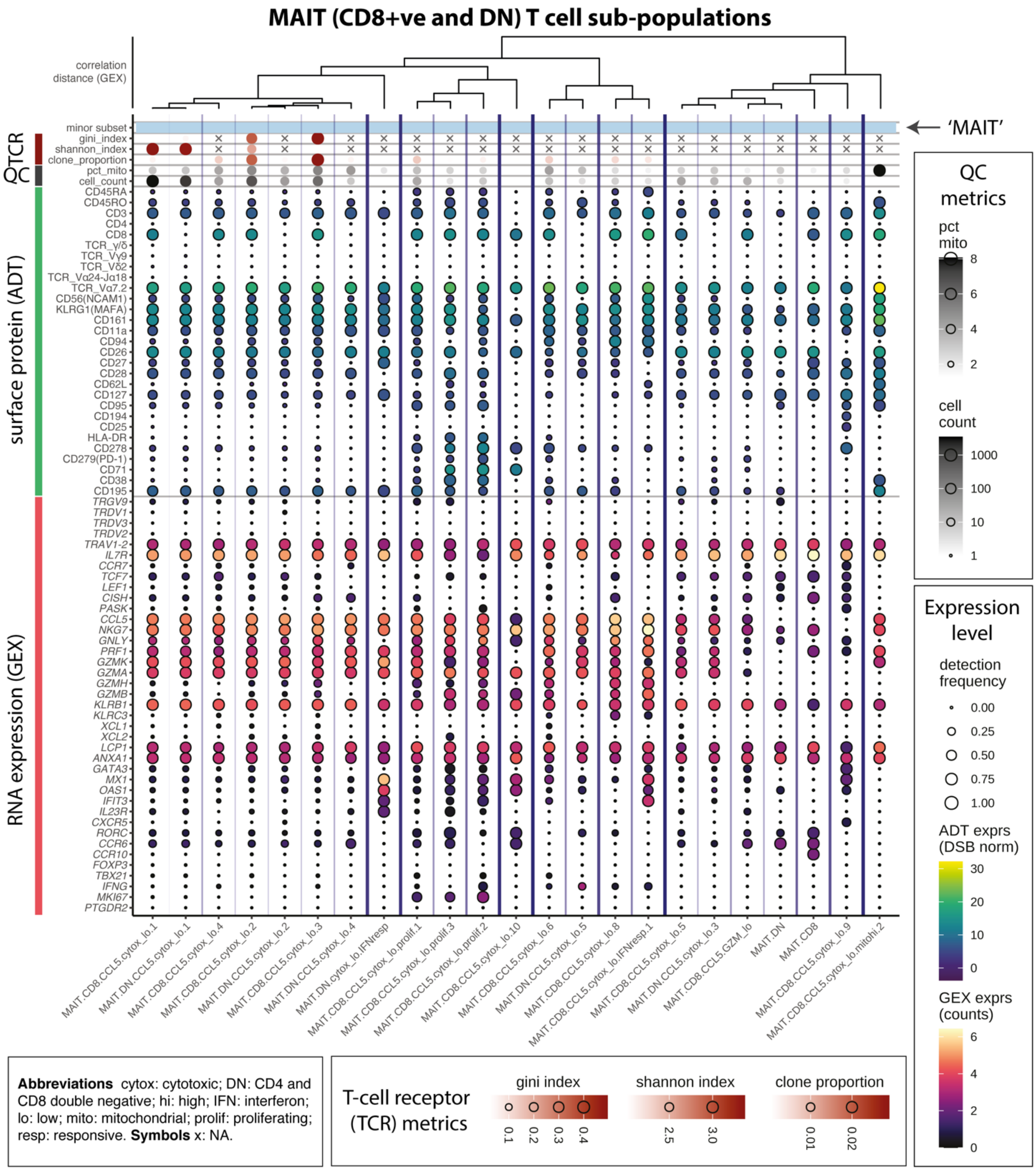

Figure 10: Mucosal associated invariant T (MAIT) cell subpopulations

#### Gamma delta T cell sub populations

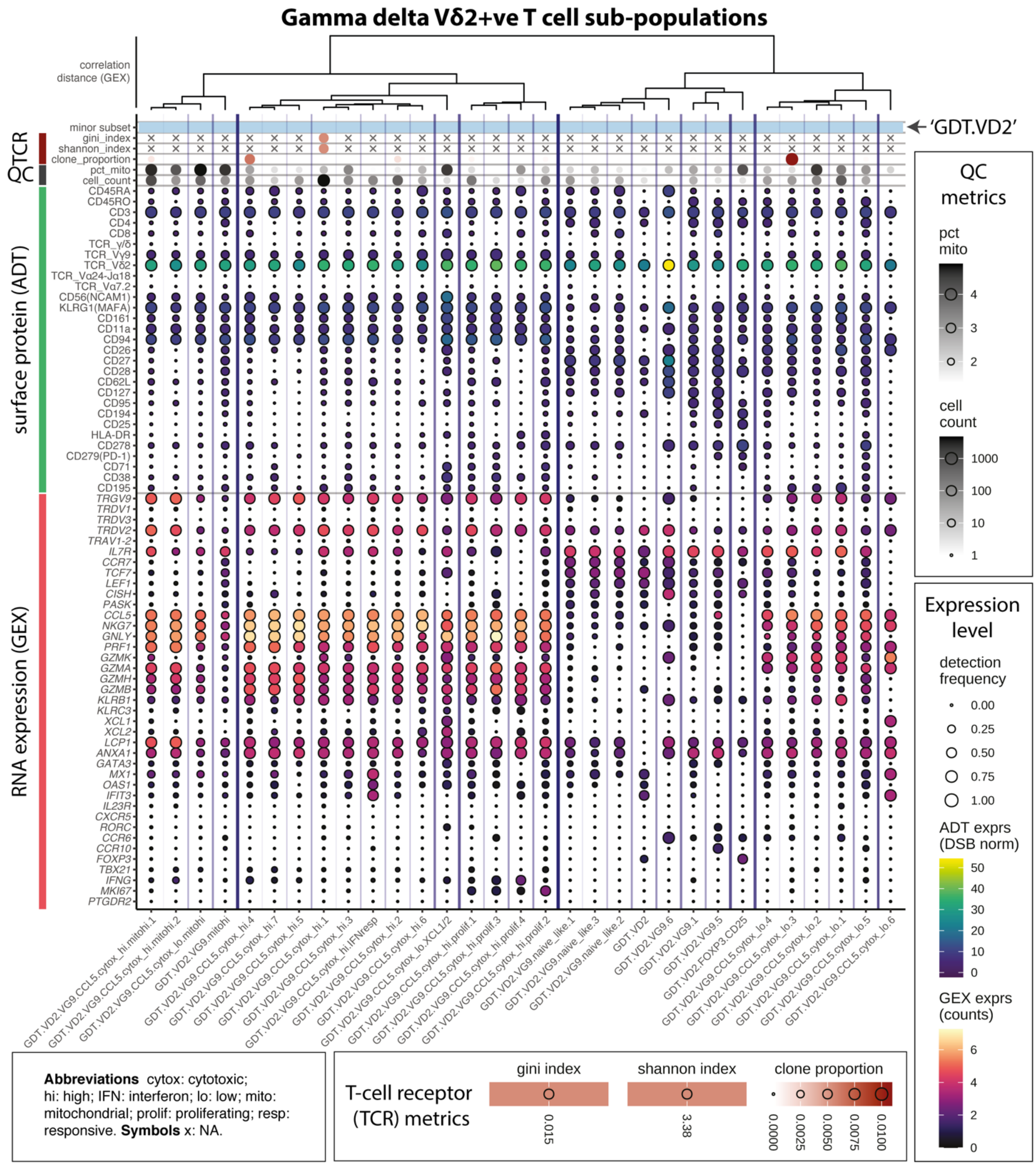

Figure 11: Vδ2+ve γδ-T cell subpopulations

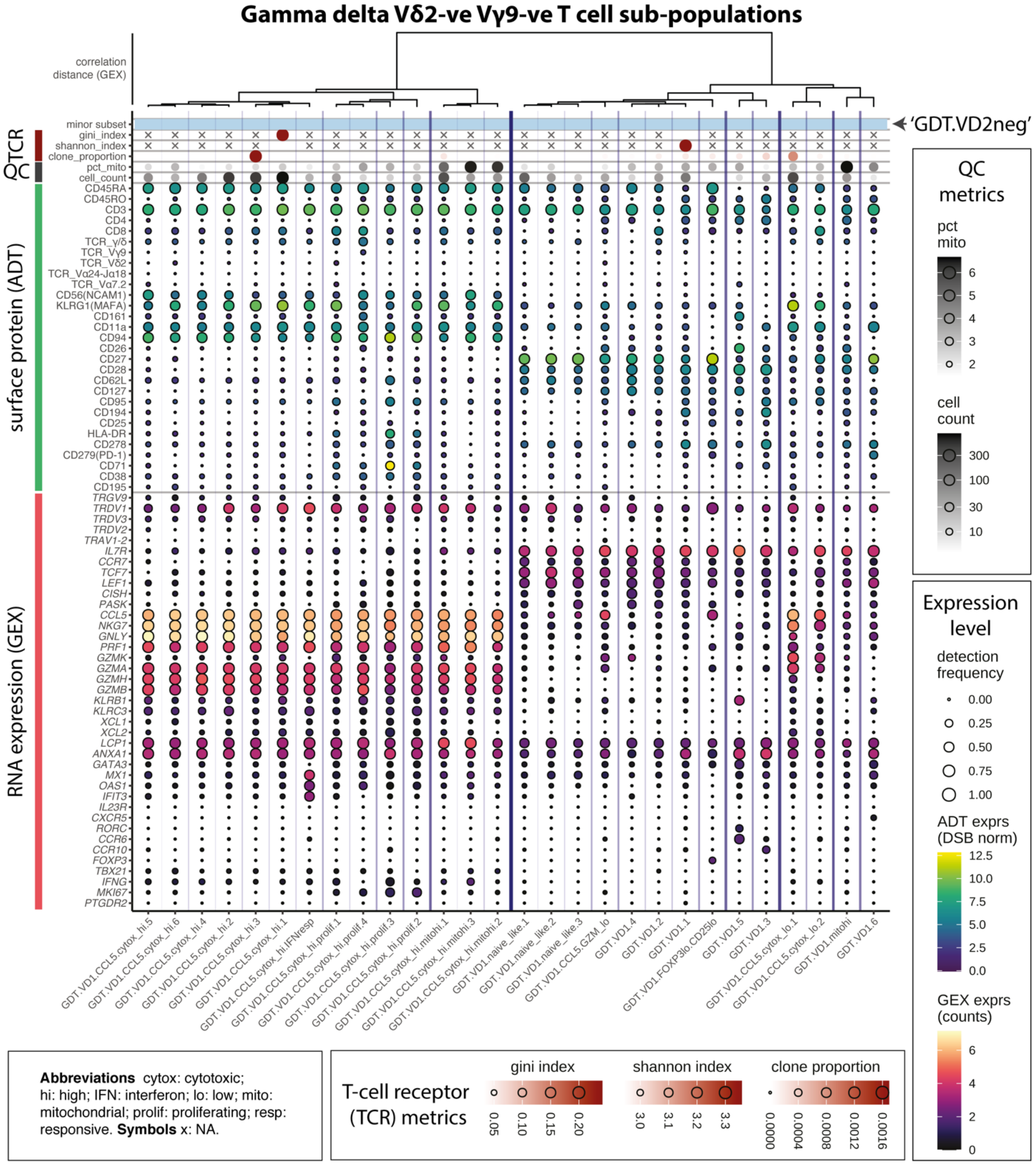

Figure 12: Vδ2-ve Vγ9-ve γδ-T cell subpopulations

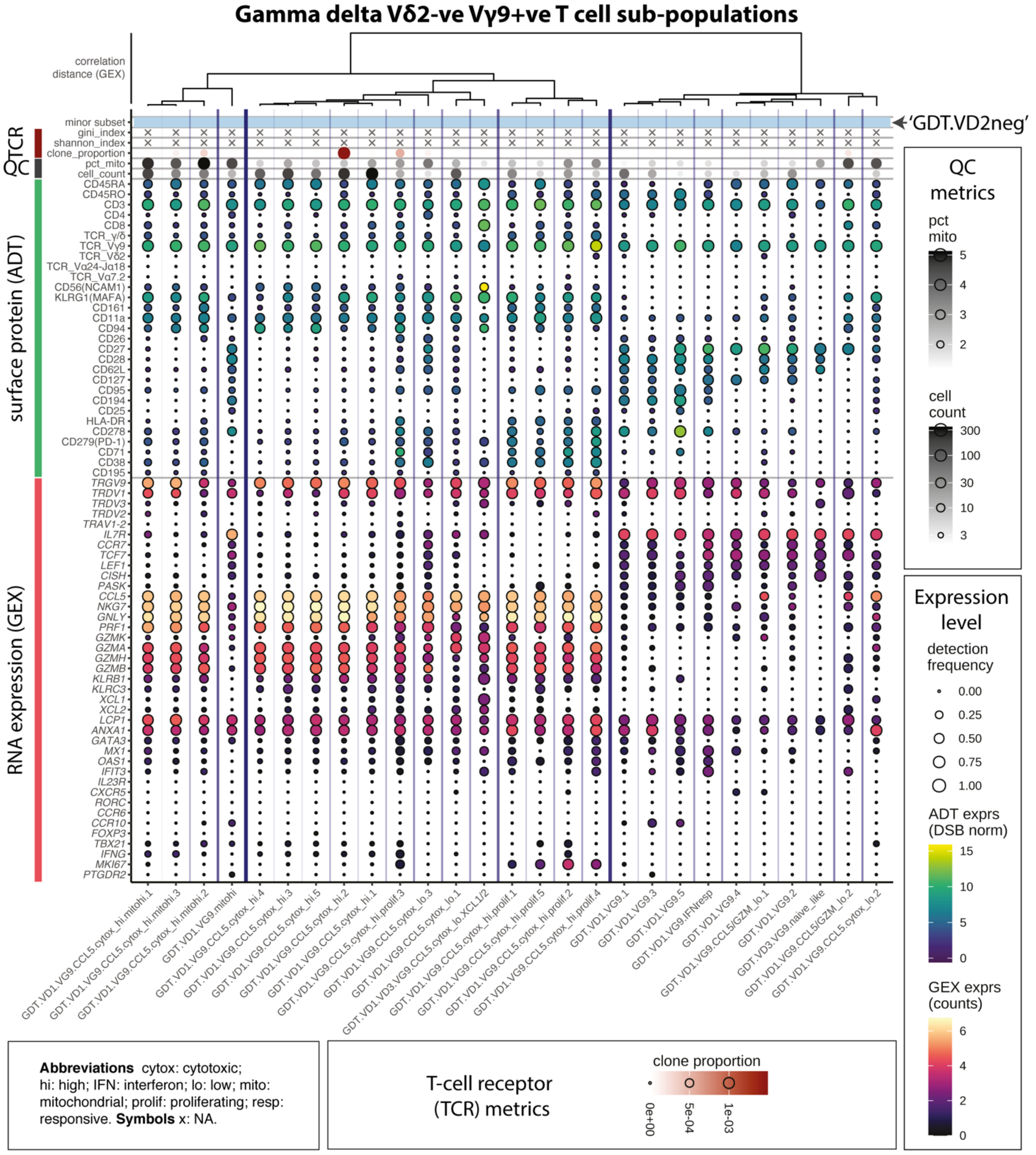

Figure 13: Vδ2-ve Vγ9+ve γδ-T cell subpopulations

#### iNKT sub-populations

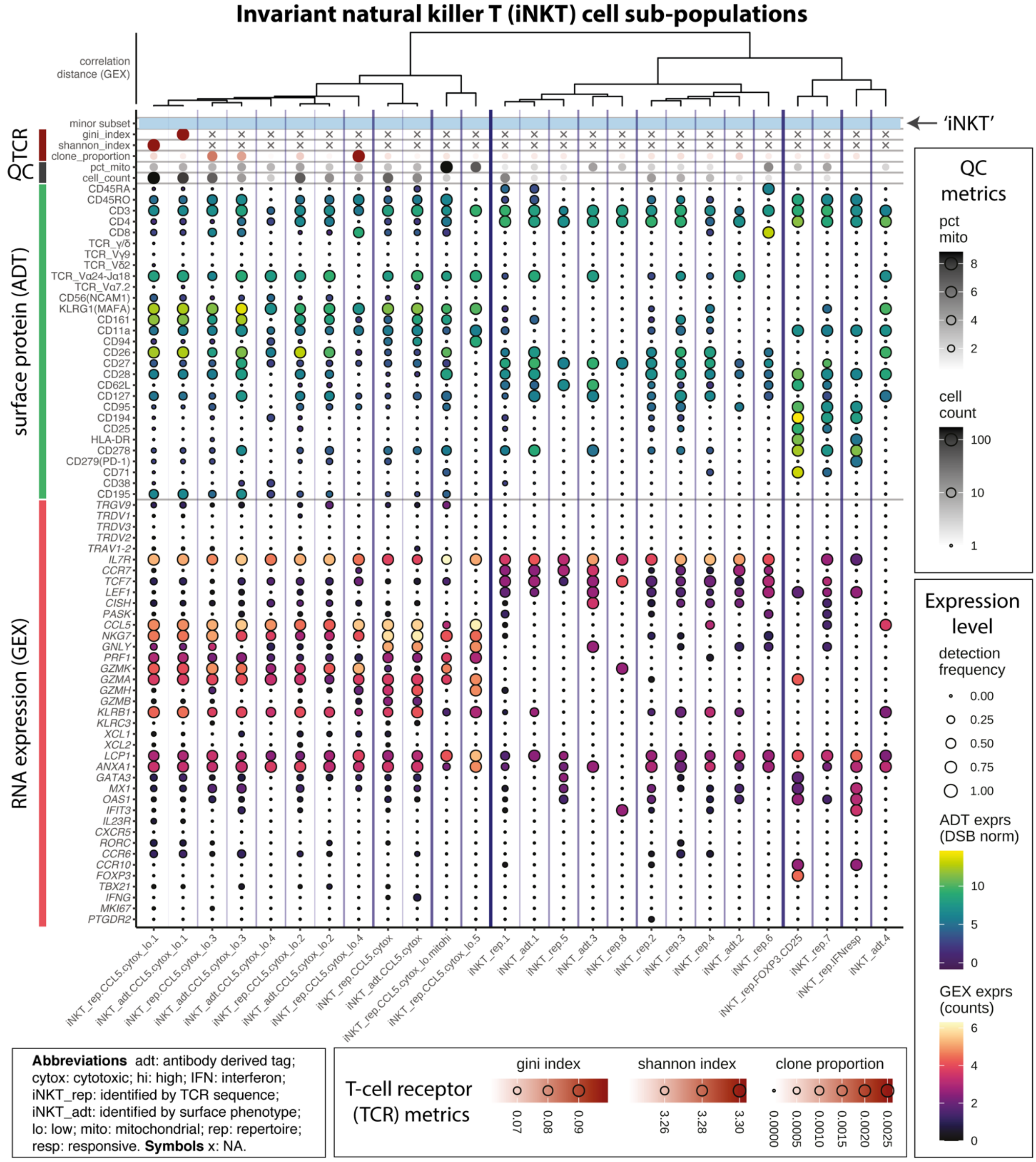

Figure 14: Invariant natural killer T (iNKT) cells

#### NK cell sub-populations

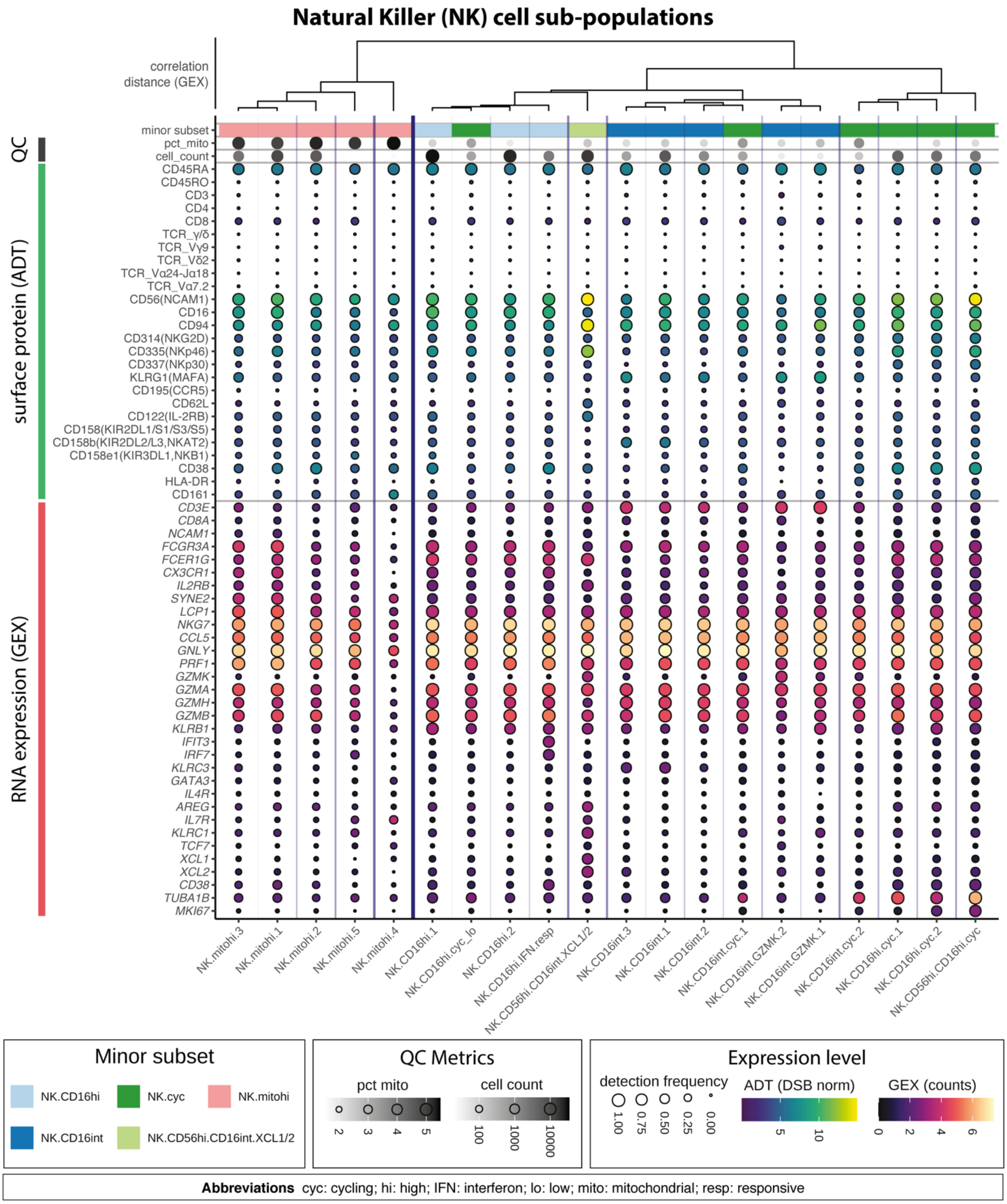

Figure 15: Natural Killer (NK) cell subpopulations

#### Table of B and plasma cell sub-population phenotypes

| Population | Subset | ADT phenotype | TCR phenotype | BCR phenotype | GEX phenotype | Cell count |
|---|---|---|---|---|---|---|
| **B** | | singlet, CD19+ | <50 TCRA & TCRB reads | singlet | CD19+ | n = 43,043 |
| | B.NAIVE.1 | IgD+ IgM+ CD27- | | IGHD, IGHM (<2 mutations) | IGHD+ IGHM+ TCL1A+ | n = 16,032 |
| | B.NAIVE.2 | IgD+ IgM+ CD27- | | | IGHD+ IGHM+ TCL1A+ | n = 2,547 |
| | B.NAIVE.3 | IgDint IgM+ CD27- | | IGHD, IGHM (<2 mutations) | IGHD+ IGHM+ TCL1A+ | n = 531 |
| | B.NAIVE.IgDlo | IgDlow IgMlow CD27- | | IGHD, IGHM (<2 mutations) | IGHD+ IGHM+ TCL1A+ | n = 71 |
| | B.NAIVE.IFN.resp | IgD+ IgM+ CD27- | | IGHD, IGHM (<2 mutations) | IGHD+ IGHM+ TCL1A+; IFN resp gene +ve (e.g. MX1+) | n = 1,176 |
| | B.NAIVE.CD1c | IgD+ IgM+ CD27- CD1c+ | | IGHD, IGHM (<2 mutations), or none | IGHD+ IGHM+ TCL1A+ | n = 922 |
| | B.int.1.IFN.resp | IgD+ IgM+ CD27int | | | IFN resp gene +ve (e.g. MX1+) | n = 199 |
| | B.int.1.early.act | IgD+ IgM+ CD27int | | IGHD, IGHM (≥2 mutations) | IGHD+ IGHM+ TCL1Aint | n = 388 |
| | B.int.1.early.act/sw.1 | IgDlow IgM+ CD27int | | IGHD, IGHM (≥2 mutations), or IGHA, IGHE, IGHG | IGHDlow IGHM+ TCL1Aint | n = 433 |
| | B.int.1.early.act/sw.2 | IgD- IgM-CD27int | | IGHD, IGHM (≥2 mutations), or IGHA, IGHE, IGHG | IGHD- IGHMlow TCL1A- | n = 168 |
| | B.int.2.unsw | IgD+ IgM+ CD27int | | IGHD, IGHM (<2 mutations) | IGHD+ IGHM+ TCL1Aint | n = 835 |
| | B.int.2.IFN.resp | IgDlow IgM+ CD27int | | | IGHDlow IGHM+ TCL1Aint; IFN resp gene +ve (e.g. MX1+) | n = 152 |
| | B.int.2.early.act.IFN.resp | IgD+ IgM+ CD27int | | IGHD, IGHM (≥2 mutations) | IGHD+ IGHM+ TCL1Aint; IFN resp gene +ve (e.g. MX1+) | n = 68 |
| | B.int.2.early.act/sw.1 | IgDlow IgM+ CD27int | | IGHD, IGHM (≥2 mutations), or IGHA, IGHE, IGHG | IGHDlow IGHM+ TCL1Aint | n = 3,978 |
| | B.int.2.early.act/sw.2 | IgD+ IgM+ CD27int | | IGHD, IGHM (≥2 mutations), or IGHA, IGHE, IGHG | IGHDlow IGHM+ TCL1Aint | n = 1,016 |
| | B.TRANSIT.CD10 | IgD+ IgM+ CD27- CD10+ | | | IGHD+ IGHM+ TCL1A+ | n = 1,116 |
| | B.mitohi.1 | | | | >4% mito | n = 1,632 |
| | B.mitohi.2 | | | | >4% mito | n = 470 |
| | B.cyc | | | | MKI67+ | n = 744 |
| | B.UNSW.MEM | IgD+ IgM+ CD27+ | | IGHD, IGHM | IGHD+ IGHM+ TCL1Aint | n = 2,597 |
| | B.SW.MEM.1 | IgD- IgM- CD27+ IgA+ or IgG+ | | IGHA, IGHE, IGHG | IGHD- IGHMlow TCL1A- | n = 5,855 |
| | B.SW.MEM.2 | IgD- IgM- CD27+ IgA+ or IgG+ | | | IGHD- IGHMlow TCL1A- | n = 1,970 |
| | B.SW.MEM.IFN.resp | IgD- IgM- CD27+ IgA+ or IgG+ | | IGHA, IGHE, IGHG or none | IGHD- IGHMlow TCL1A-; IFN resp gene +ve (e.g. MX1+) | n = 93 |
| **Plasma-blasts** | | singlet, CD19low CD20- CD38hi CD39hi | <50 TCRA & TCRB reads | singlet | XBP1+ SLAMF7+ PRDM1+ | n = 8,596 |
| | PB | | | >100 HC UMI | | n = 5,948 |
| | PB.IFN.resp | | | >100 HC UMI | IFN resp gene +ve (e.g. MX1hi) | n = 38 |
| | PB.mitohi | | | >100 HC UMI | >4% mito | n = 36 |
| | PB.cyc | | | >100 HC UMI | MKI67hi | n = 2,574 |

Table 2: Definition of B and plasmablast subsets. red: identifying features, green: additional criteria, blue: confirmatory observations (thresholds checked but not applied)

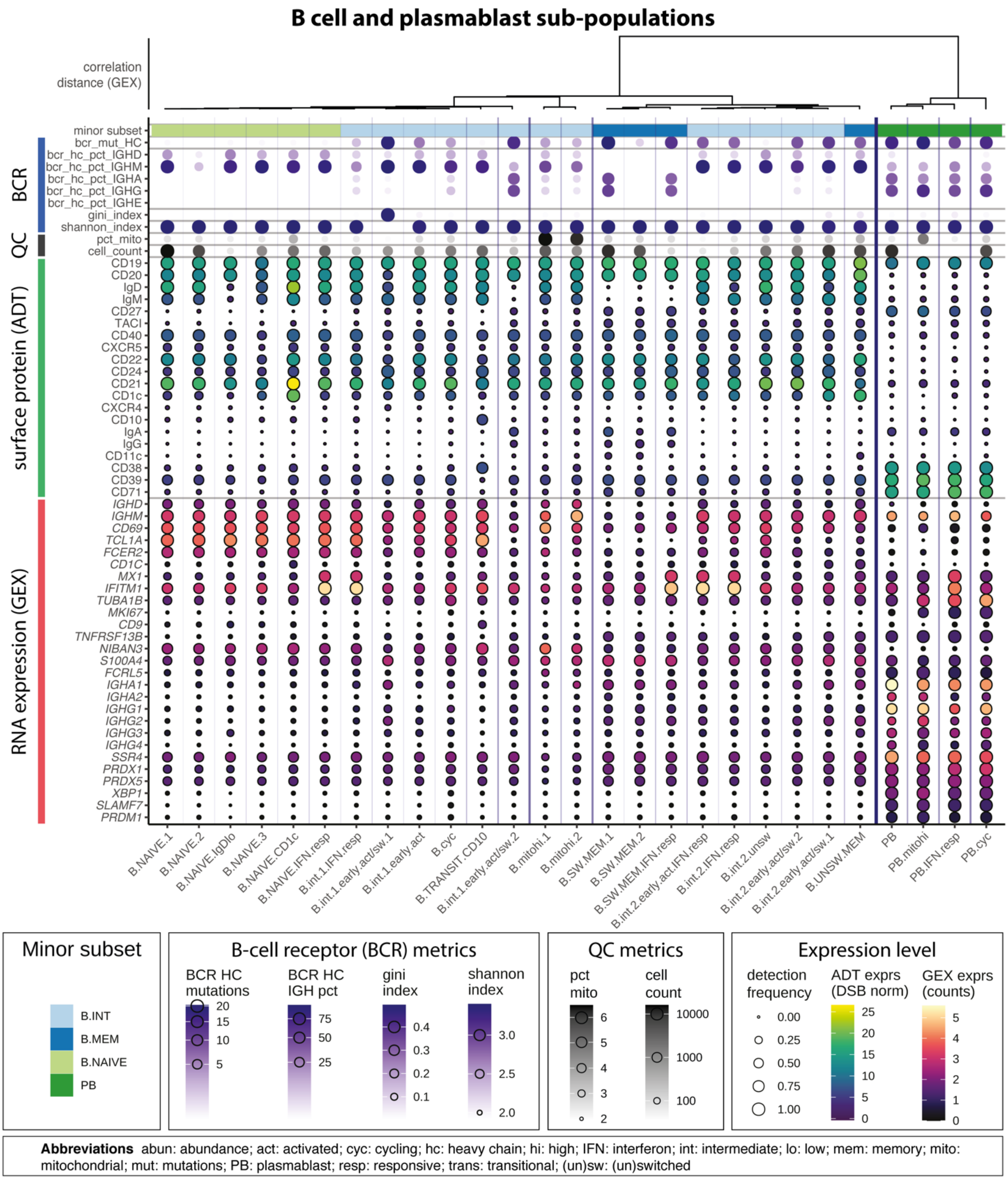

Figure 16: B cell and plasmablast sub-populations.

#### Mononuclear phagocyte sub-populations

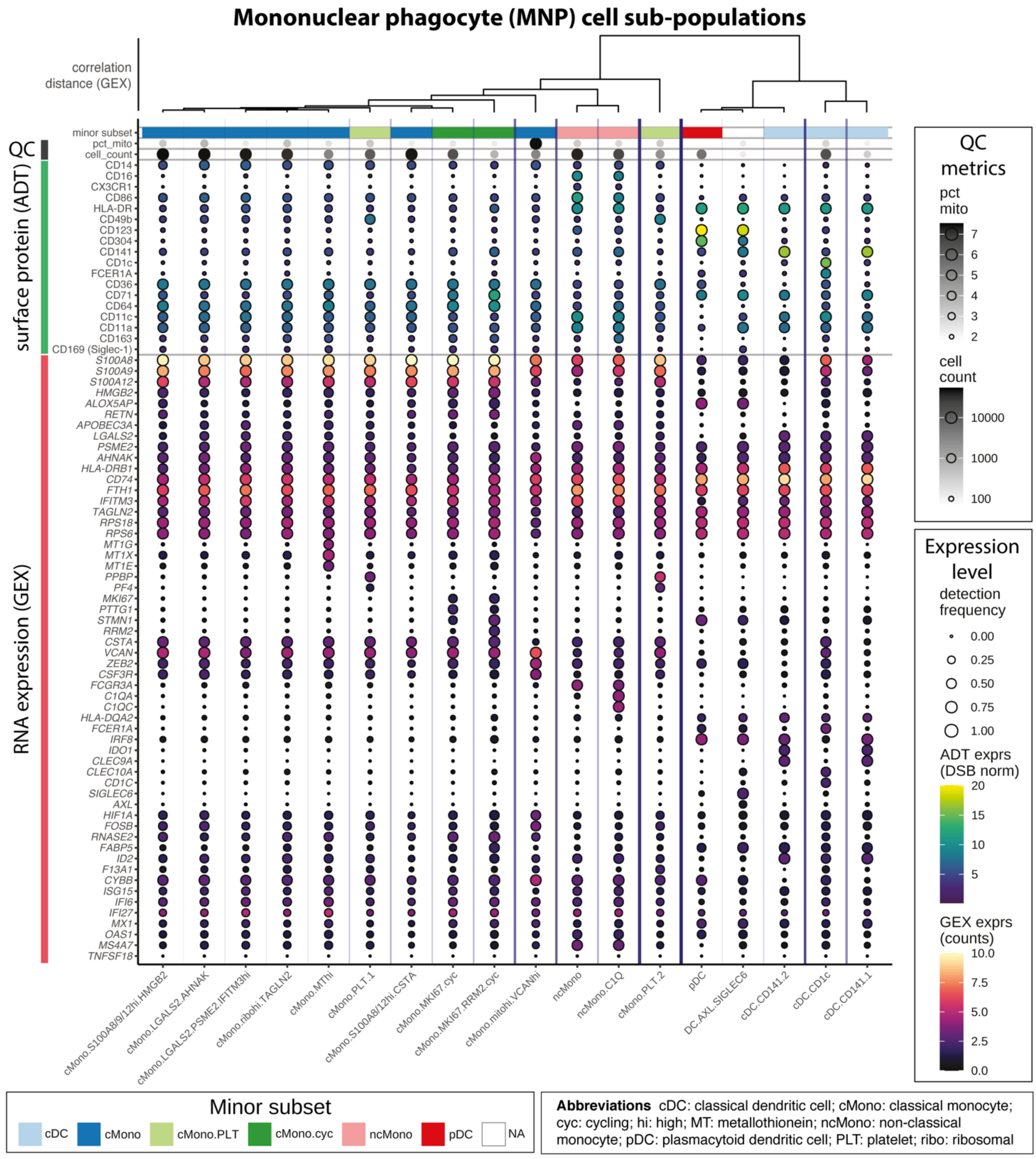

Figure 17: Mononuclear phagocyte (MNP) cell sub-populations

#### Haematopoietic stem (and progenitor) cell sub-populations

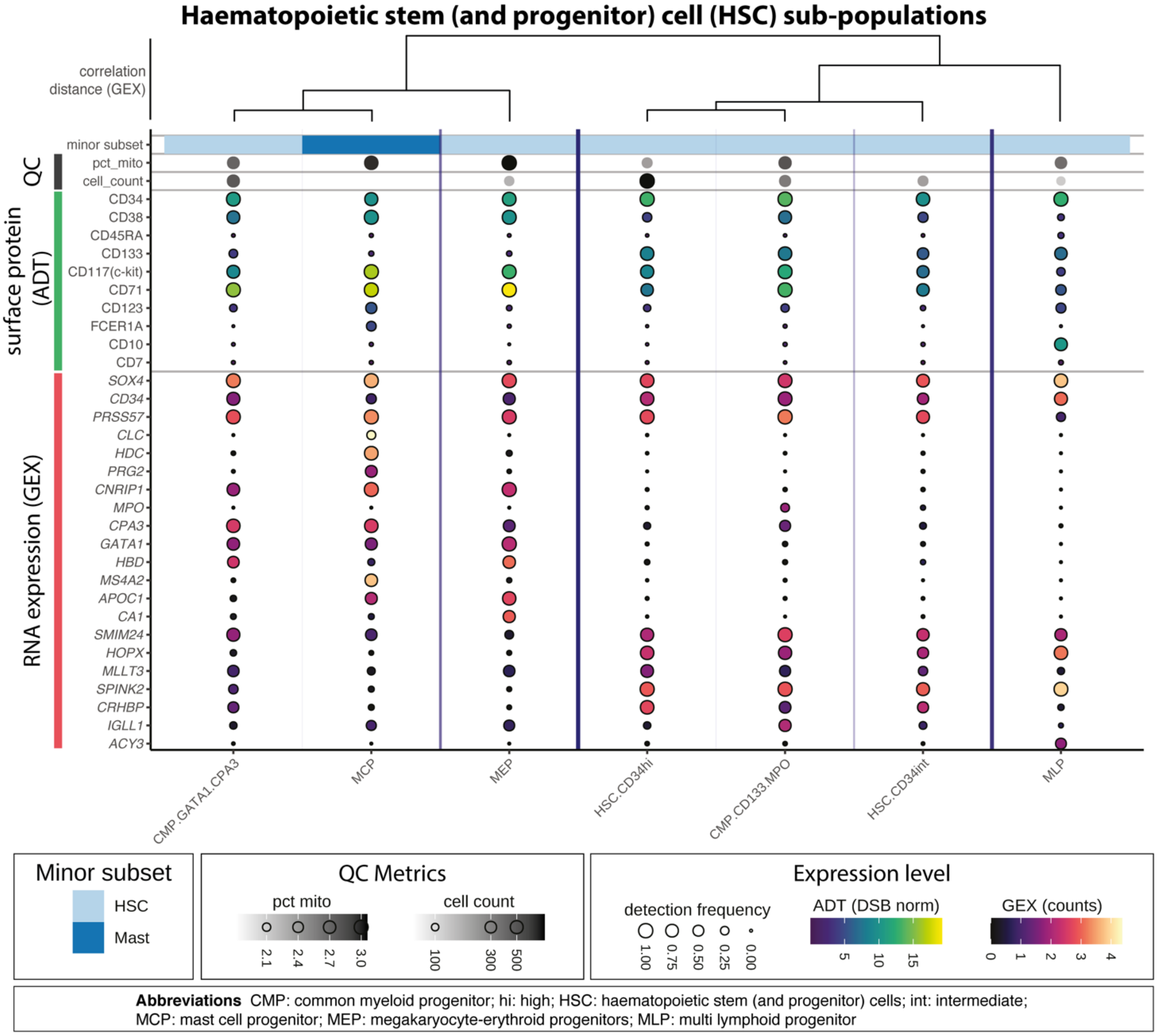

Figure 18: Haematopoietic stem (and progenitor) cell (HSC) sub-populations

#### Platelet and erythrocyte sub-populations

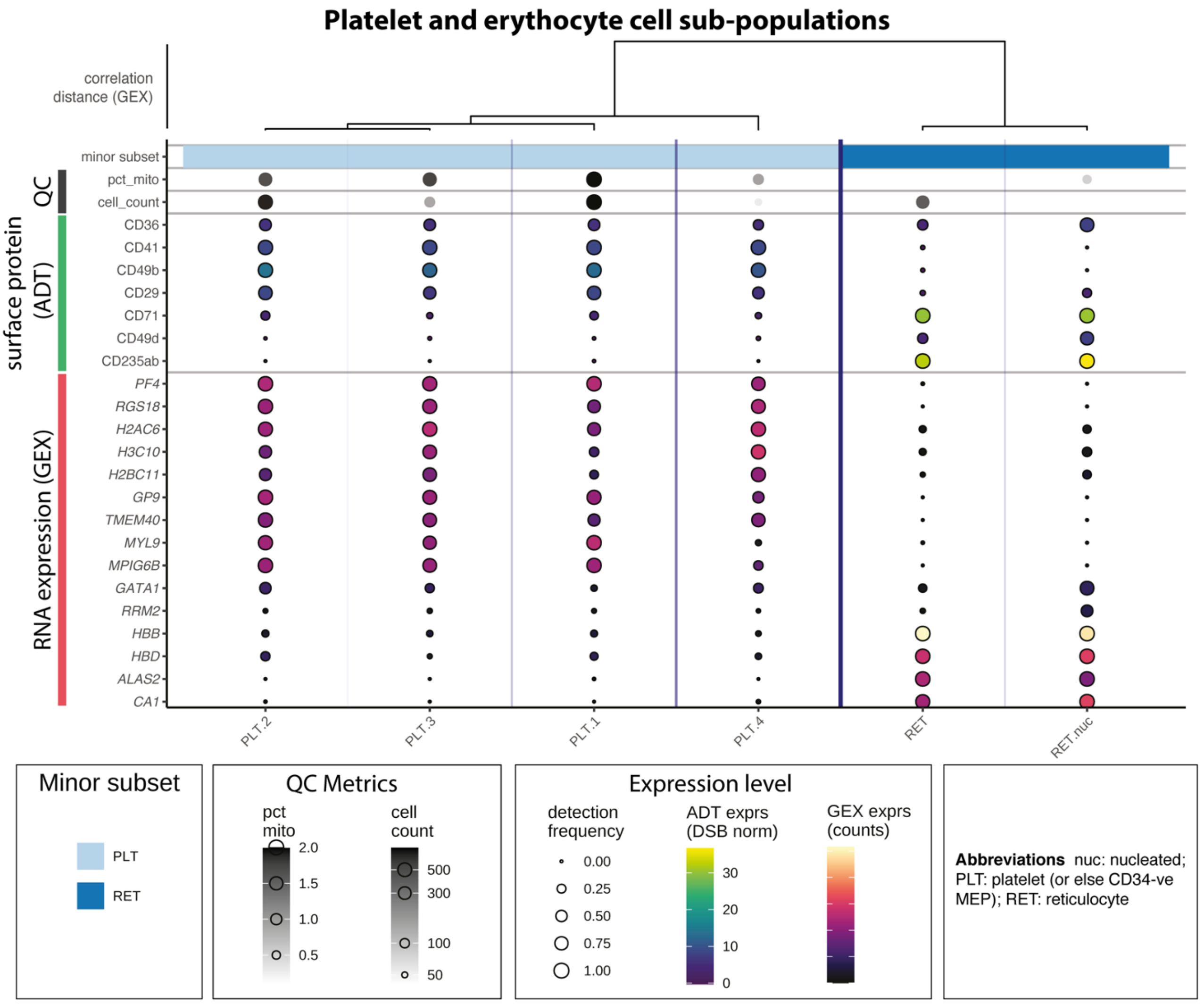

Figure 19: Platelet and erythrocyte cell sub-populations

### Composition analysis

#### Composition analysis of cell types

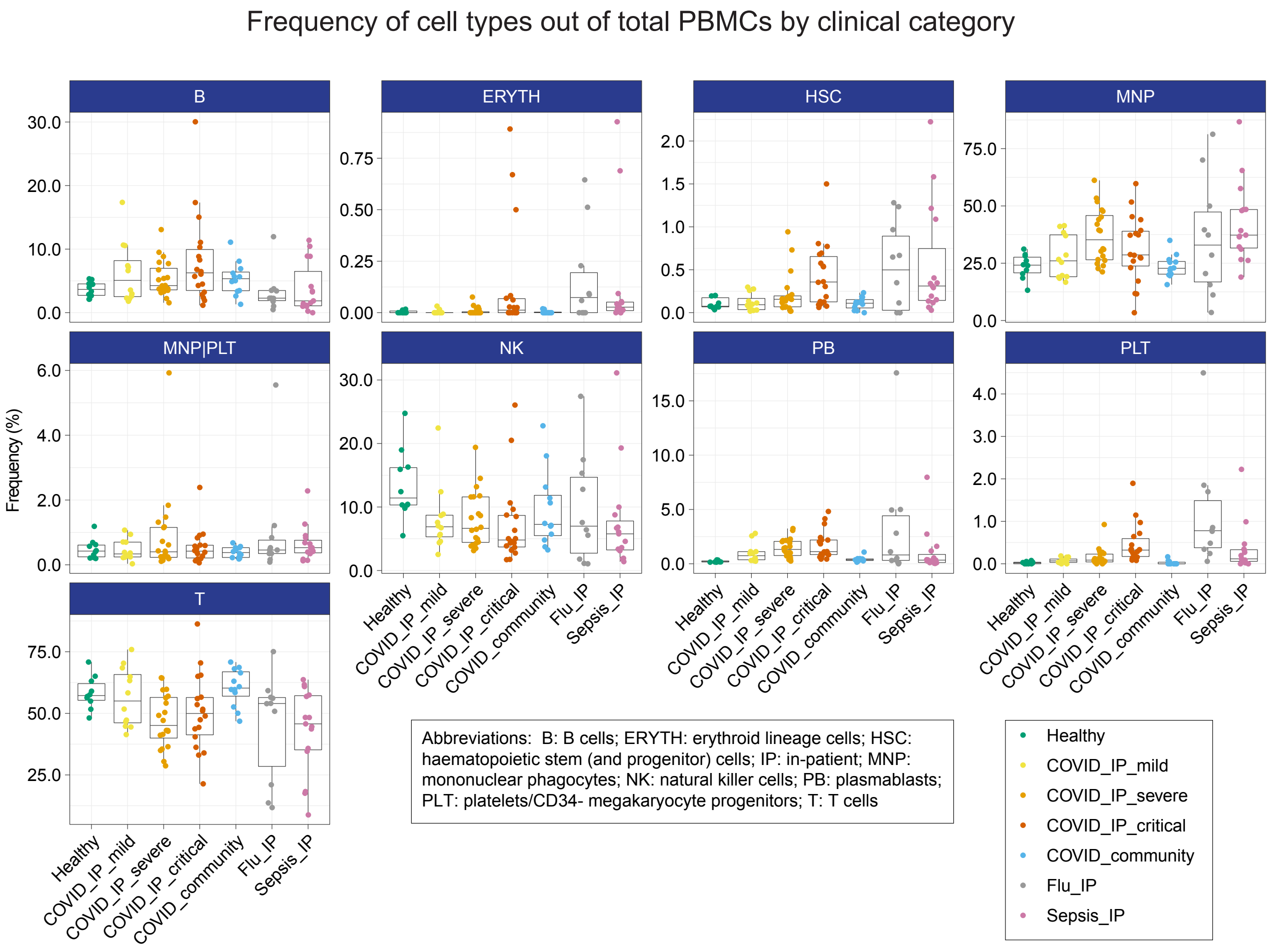

Figure 20: Cell type frequency by clinical category. The box plots show the median and the first and third quartiles; whiskers show 1.5 × the interquartile range above and below the box. Cell types are ordered alphabetically.

Cell types - PCA

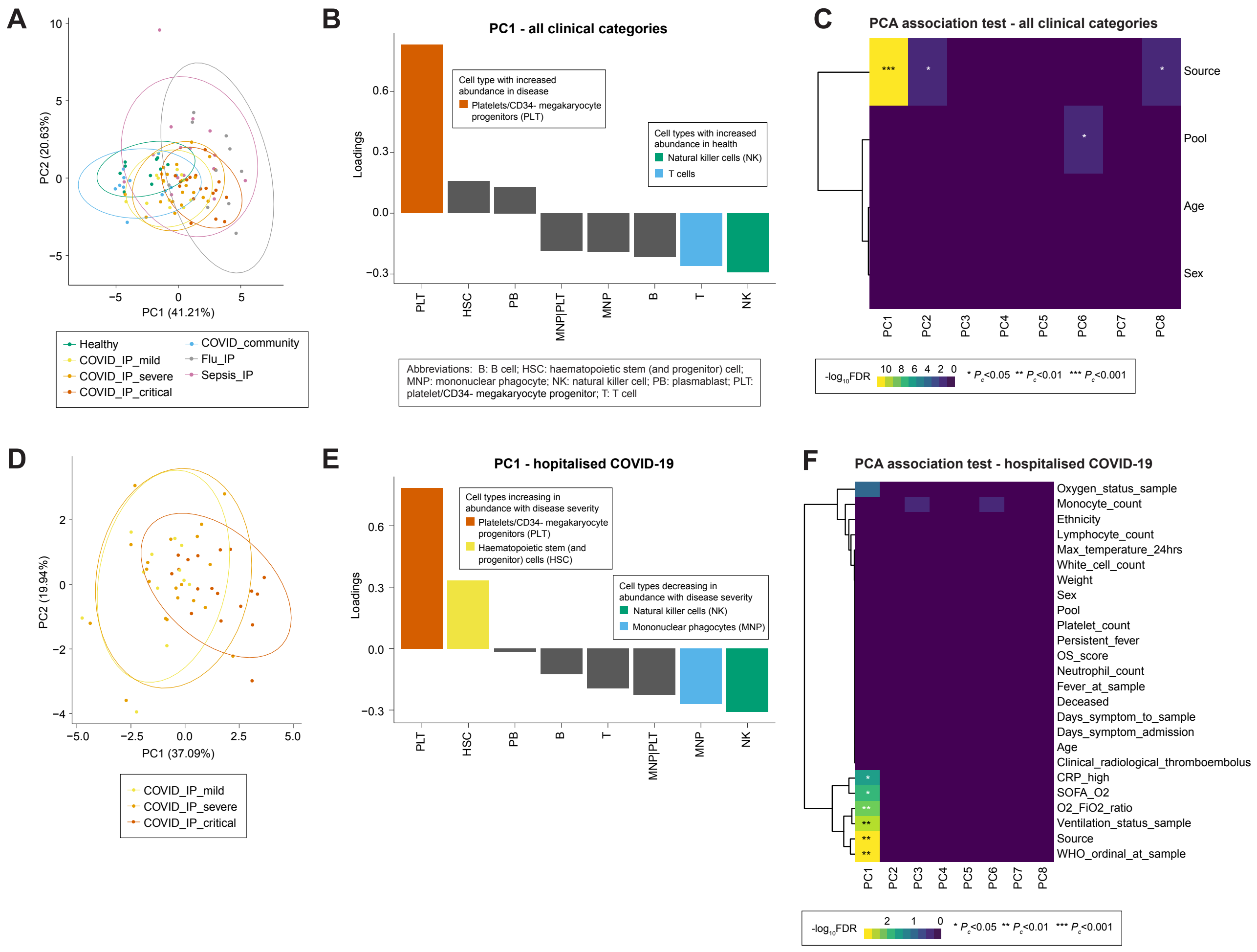

Figure 21: Cell type PCA and association tests. (A) PCA plot for all clinical categories. Percentage of variance explained by each PC is shown in the brackets. (B) Loadings of cell types on PC1 from analysis including all clinical categories. (C) Association test between top PCs and source (clinical category), age, sex, and sample pool for all clinical categories. (D) PCA plot for hospitalised COVID-19. Percentage of variance explained by each PC is shown in the brackets. (E) Loadings of cell types on PC1 from analysis of hospitalised COVID-19 cases. (F) Association test between top PCs and clinical, demographic and experimental variables for hospitalised COVID-19 cases. Significance tested using Benjamini-Hochberg-corrected ANOVA.

### Cell types - edgeR differential abundance analysis

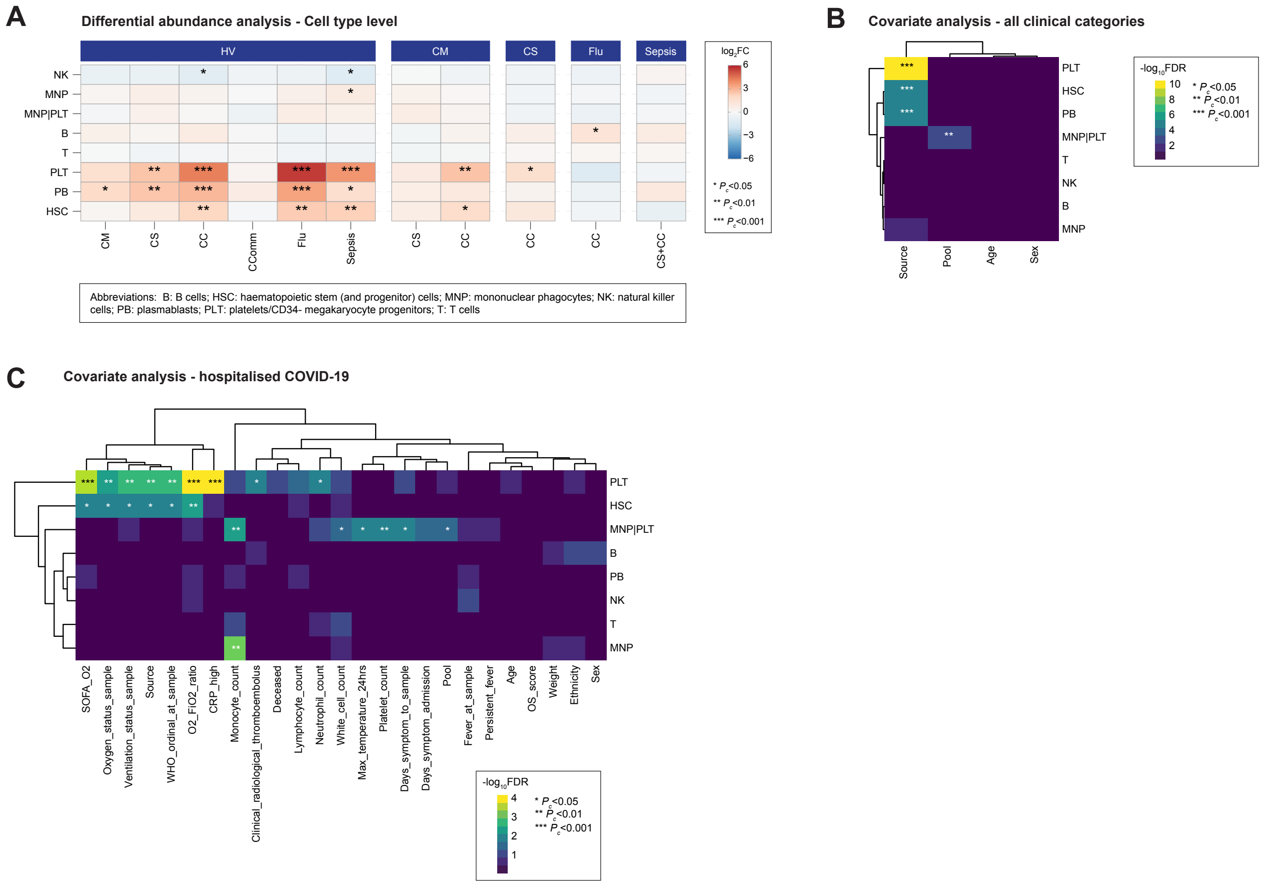

Figure 22: Cell type differential abundance analysis. (A) Cell type differential abundance analysis between clinical categories, controlling for age, sex and sample pool effects. (B) Cell type covariate analysis for source (clinical category), age, sex and sample pool for all clinical categories. (C) Cell type covariate analysis for clinical, demographic and experimental variables for hospitalised COVID-19 cases. Significance tested using Benjamini-Hochberg-corrected ANOVA.

#### Composition analysis of major cell subsets

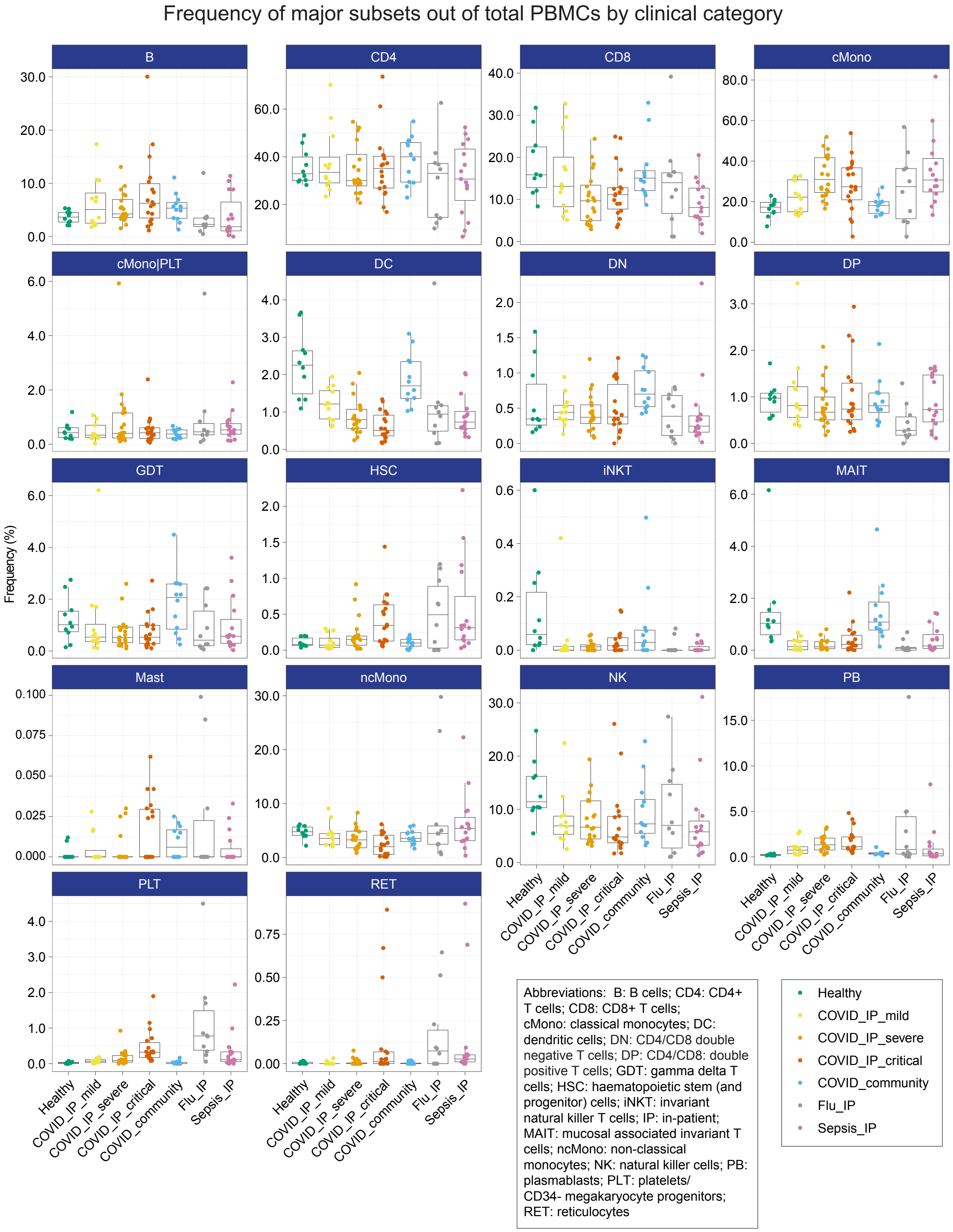

Figure 23: Major cell subset frequency by clinical category. Box plots show the median and the first and third quartiles; whiskers show 1.5 × the interquartile range above and below the box. Major subsets are ordered alphabetically.

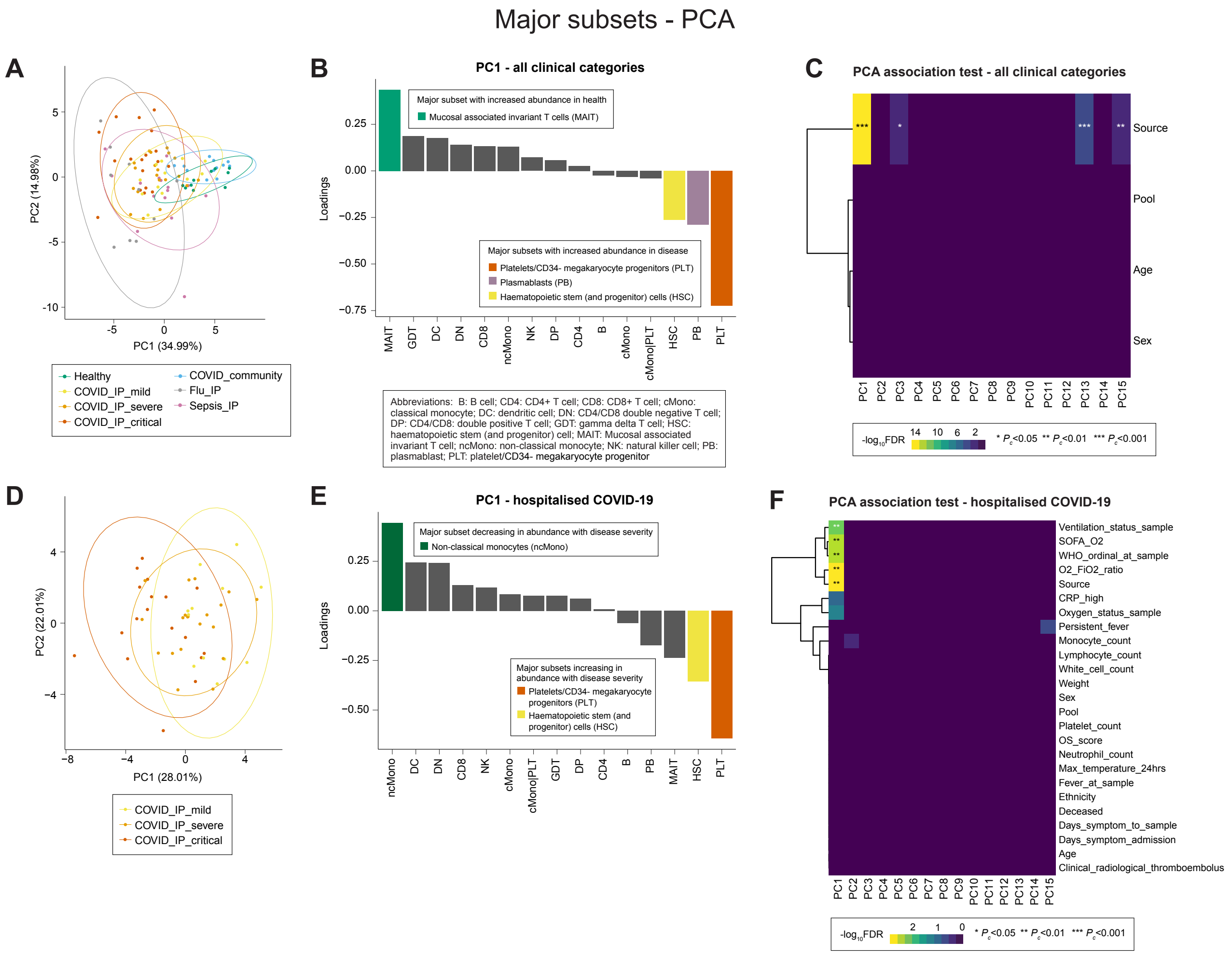

Figure 24: Major cell subset PCA and association tests. (A) PCA plot for all clinical categories. Percentage of variance explained by each PC is shown in the brackets. (B) Loadings of major subsets on PC1 from analysis including all clinical categories. (C) Association test between top PCs and source (clinical category), age, sex, and sample pool for all clinical categories. (D) PCA plot for hospitalised COVID-19. Percentage of variance explained by each PC is shown in the brackets. (E) Loadings of major subsets on PC1 from analysis of hospitalised COVID-19 cases. (F) Association test between top PCs and clinical, demographic and experimental variables for hospitalised COVID-19 cases. Significance tested using Benjamini-Hochberg-corrected ANOVA.

#### Major subsets - edgeR differential abundance analysis

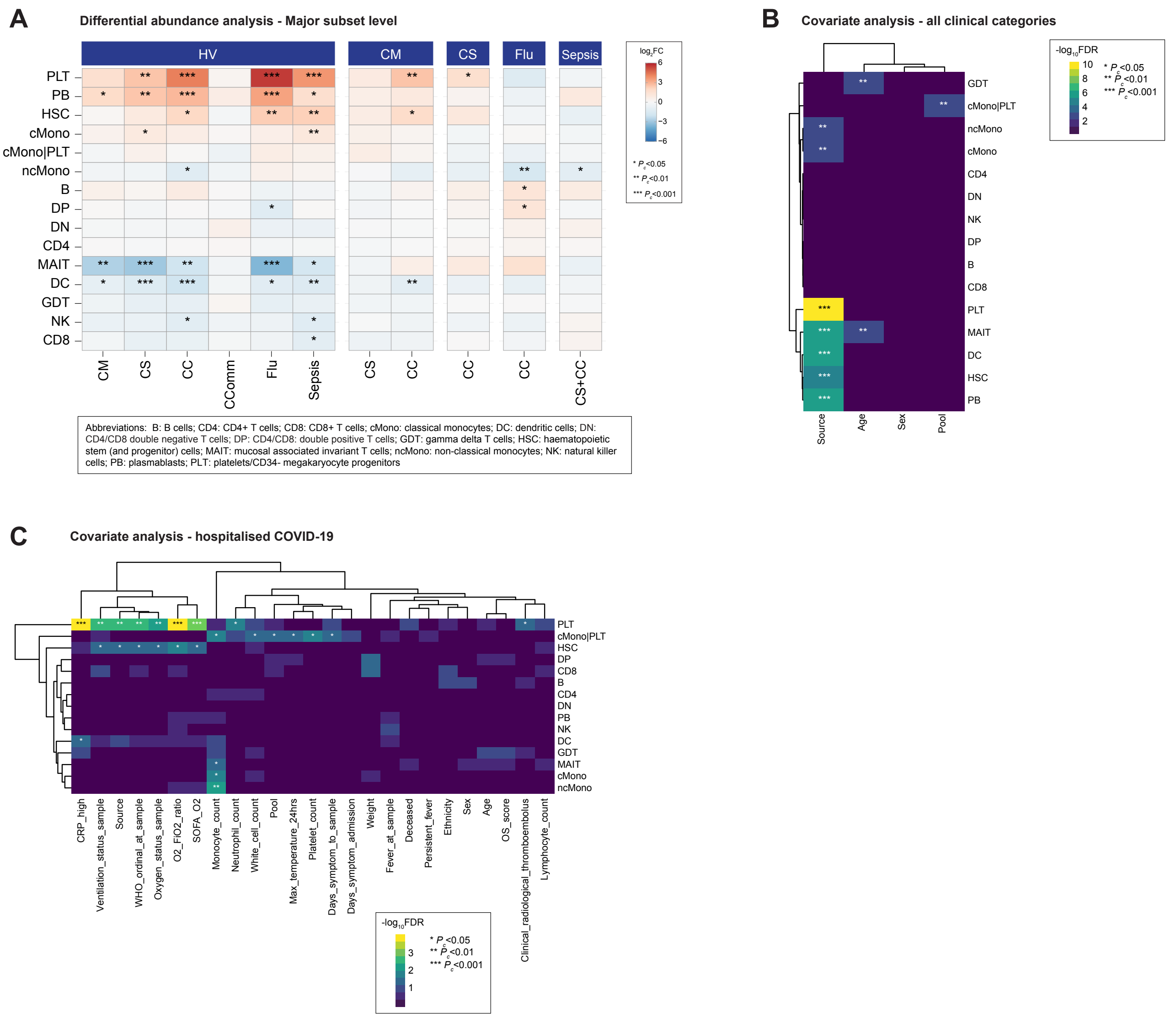

Figure 25: Major cell subset differential abundance analysis. (A) Major subset differential abundance analysis between clinical categories, controlling for age, sex and sample pool. (B) Major subset covariate analysis for source (clinical category), age, sex and pool for all clinical categories. (C) Major subset covariate analysis for clinical, demographic and experimental variables for hospitalised COVID-19 cases. Significance tested using Benjamini-Hochberg-corrected ANOVA.

#### Composition analysis of minor cell subsets

Frequency of minor subsets out of total PBMCs by clinical category

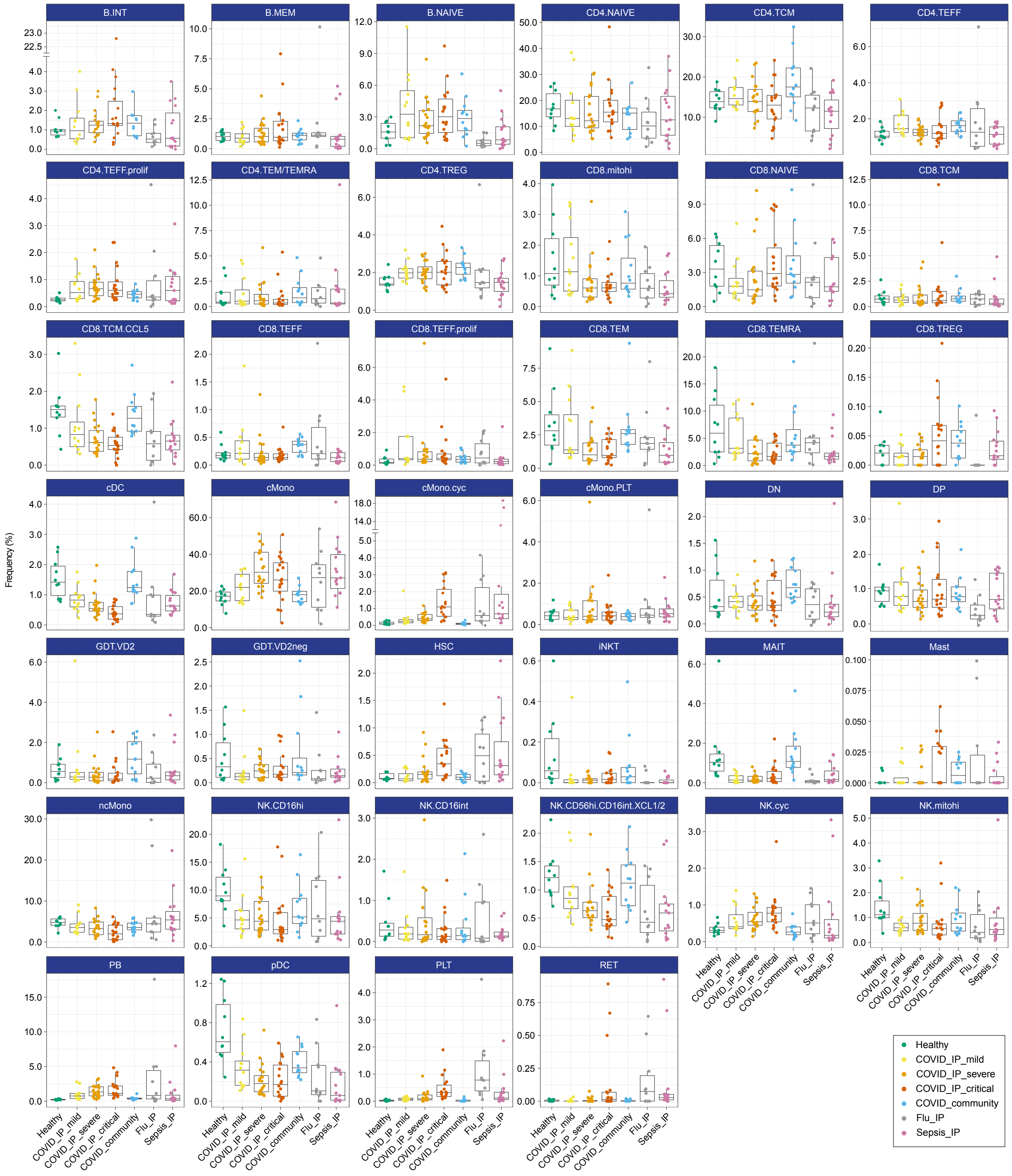

Abbreviations: B: B cell; cDC: classical dendritic cell; cMono: classical monocyte; cyc: cycling; DN: CD4/CD8 double negative T cell; DP: CD4/CD8: double positive T cell; GDT: gamma delta T cell; hi: high; HSC: haematopoietic stem (and progenitor) cell; iNKT: invariant NKT cell; INT/int: intermediate; IP: in-patient; MAIT: mucosal associated invariant T cell; MEM: memory; mito: mitochondrial; ncMono: non-classical monocyte; neg: negative; NK: natural killer cell; PB: plasmablast; pDC: plasmacytoid dendritic cell; PLT: platelet/CD34- megakaryocyte progenitor; prolif: proliferating; TCM: T central memory; TEFF: T effector; TEM(RA): T effector memory (CD45RA re-expressing); TREG: T regulatory cell; RET: reticulocytes

Figure 26: Minor cell subset frequency by clinical category. Box plots show the median and the first and third quartiles; whiskers show 1.5 × the interquartile range above and below the box. Minor subsets are ordered alphabetically.

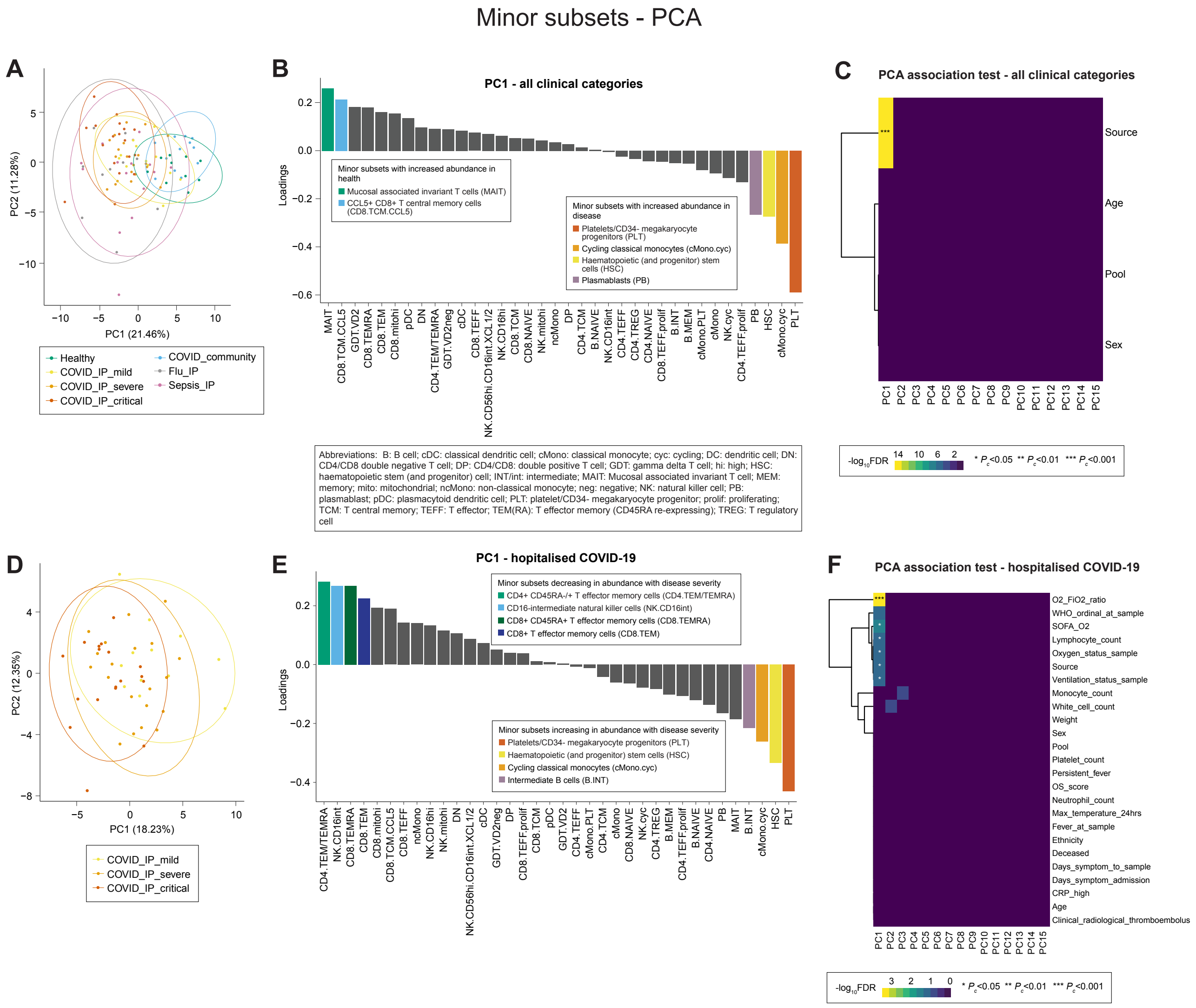

Figure 27: Minor cell subset PCA and association tests. (A) PCA plot for all clinical categories. Percentage of variance explained by each PC is shown in the brackets. (B) Loadings of minor subsets on PC1 from analysis including all clinical categories. (C) Association test between top PCs and source (clinical category), age, sex, and sample pool for all clinical categories. (D) PCA plot for hospitalised COVID-19. Percentage of variance explained by each PC is shown in the brackets. (E) Loadings of minor subsets on PC1 from analysis of hospitalised COVID-19 cases. (F) Association test between top PCs and clinical, demographic and experimental variables for hospitalised COVID-19 cases. Significance tested using Benjamini-Hochberg-corrected ANOVA.

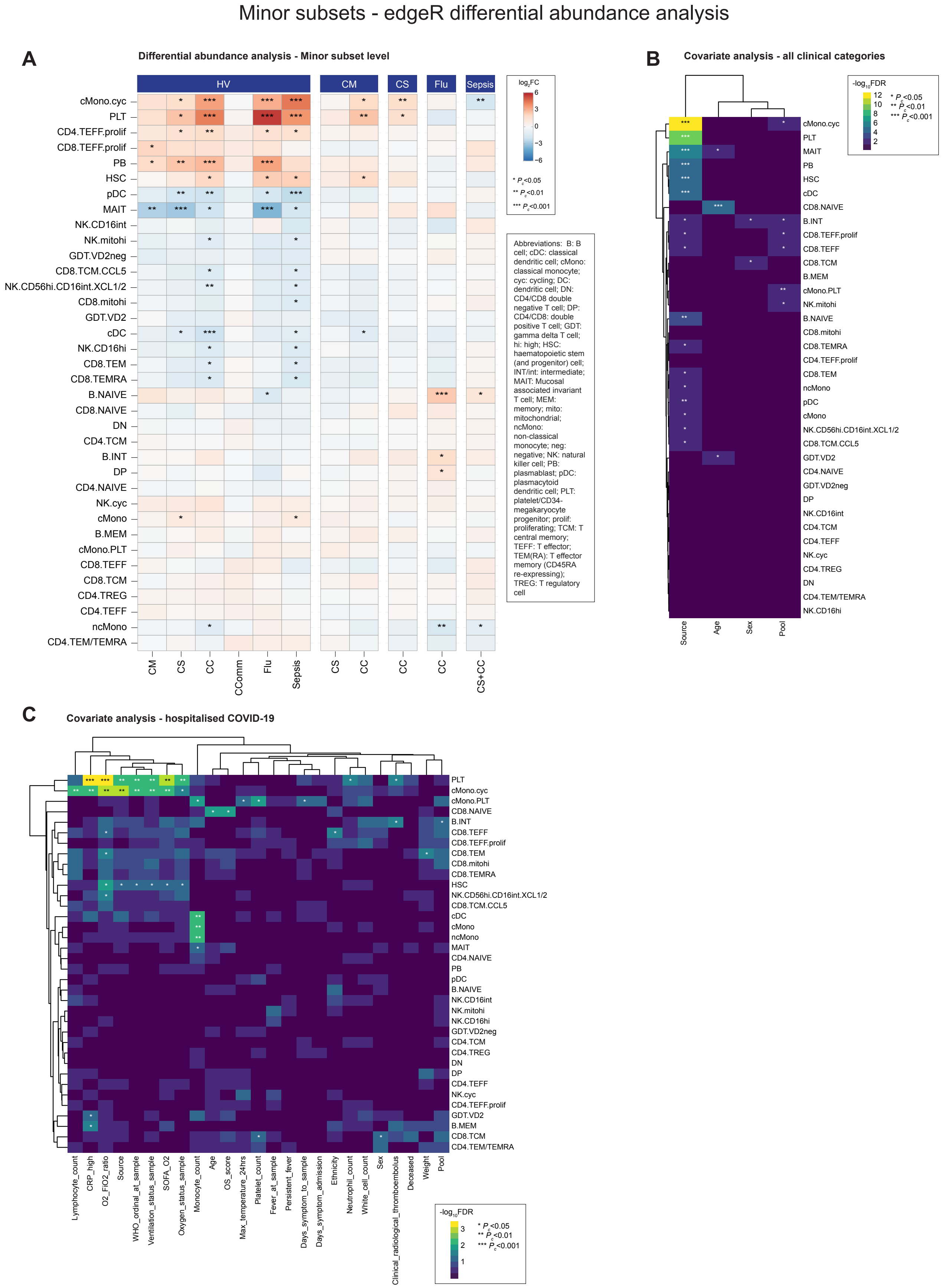

Figure 28: Minor cell subset differential abundance analysis. (A) Minor subset differential abundance analysis between clinical categories, controlling for age, sex and sample pool. (B) Minor subset covariate analysis for source (clinical category), age, sex and pool for all clinical categories. (C) Minor subset covariate analysis for clinical, demographic and experimental variables for hospitalised COVID-19 cases. Significance tested using Benjamini-Hochberg-corrected ANOVA.

#### Composition analysis of T and natural killer cell clusters

Frequency of clusters out of total T and natural killer cells by clinical category (1)

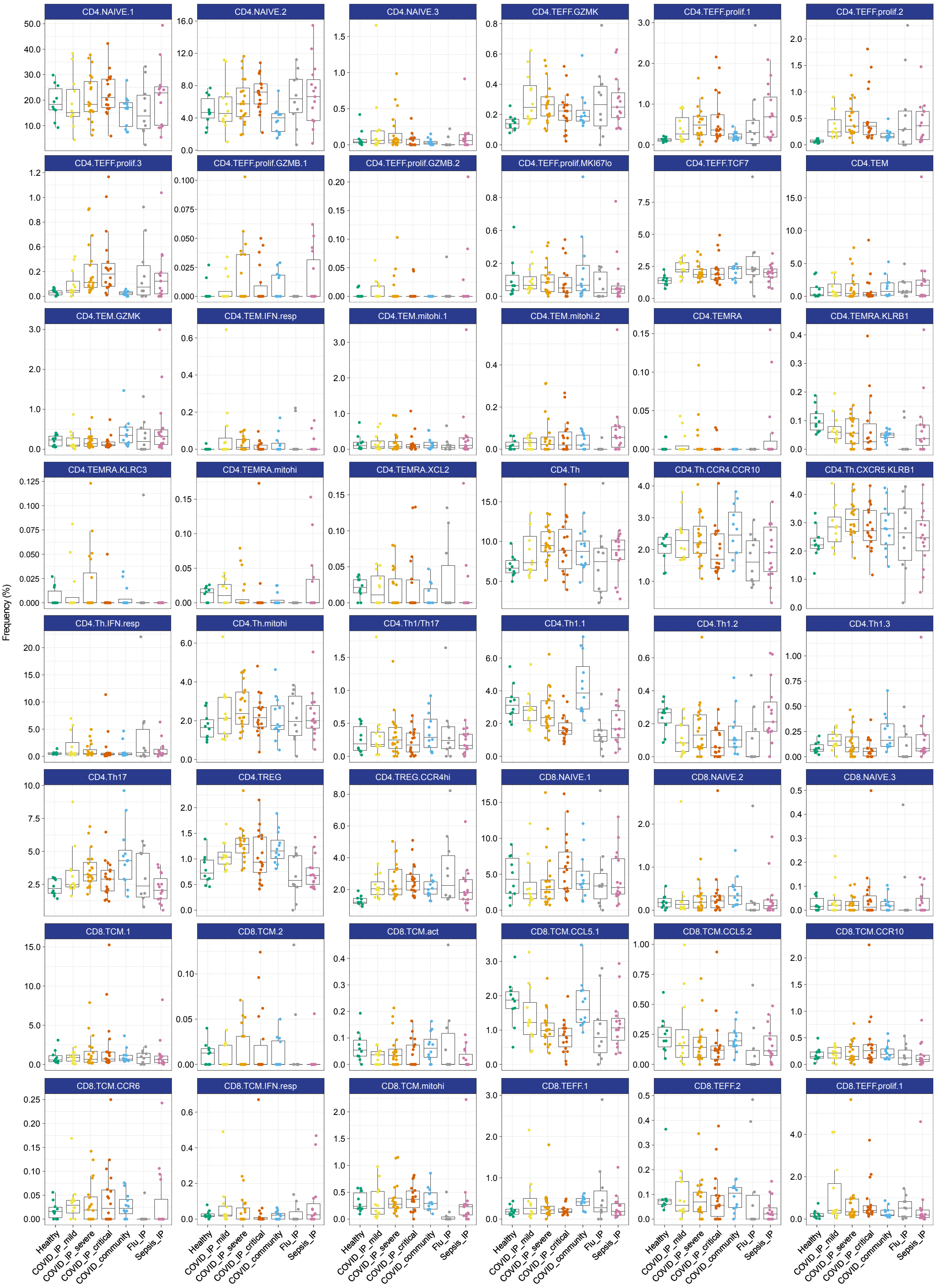

Abbreviations: act: activated; hi: high; IP: in-patient; lo: low; mito: mitochondrial; prolif: proliferating; resp: responsive; TCM: T central memory; TEFF: T effector; TEM(RA): T effector memory (CD45RA re-expressing); Th: T helper; TREG: T regulatory

- Healthy
- COVID_IP_mild
- COVID_IP_severe
- COVID_IP_critical
- COVID_community
- Flu_IP
- Sepsis_IP

### Frequency of clusters out of total T and natural killer cells by clinical category (2)

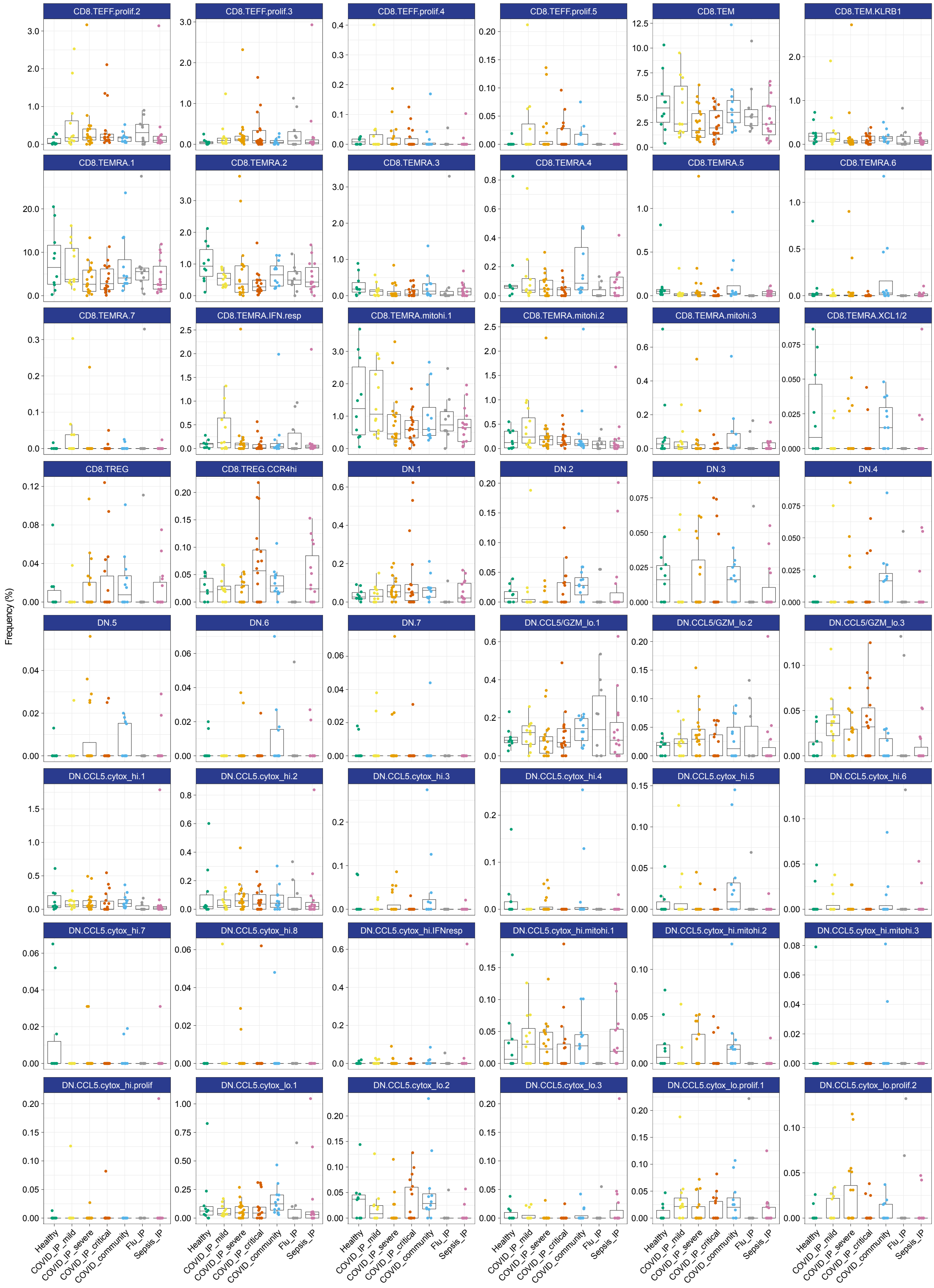

Abbreviations: cytox: cytotoxic; DN: CD4/CD8 double negative; GZM: granzyme; hi: high; IP: in-patient; lo: low; mito: mitochondrial; prolif: proliferating; resp: responsive; TEFF: T effector; TEM(RA): T effector memory (CD45RA re-expressing); TREG: T regulatory

- Healthy
- COVID_IP_mild
- COVID_IP_severe
- COVID_IP_critical
- COVID_community
- Flu_IP
- Sepsis_IP

### Frequency of clusters out of total T and natural killer cells by clinical category (3)

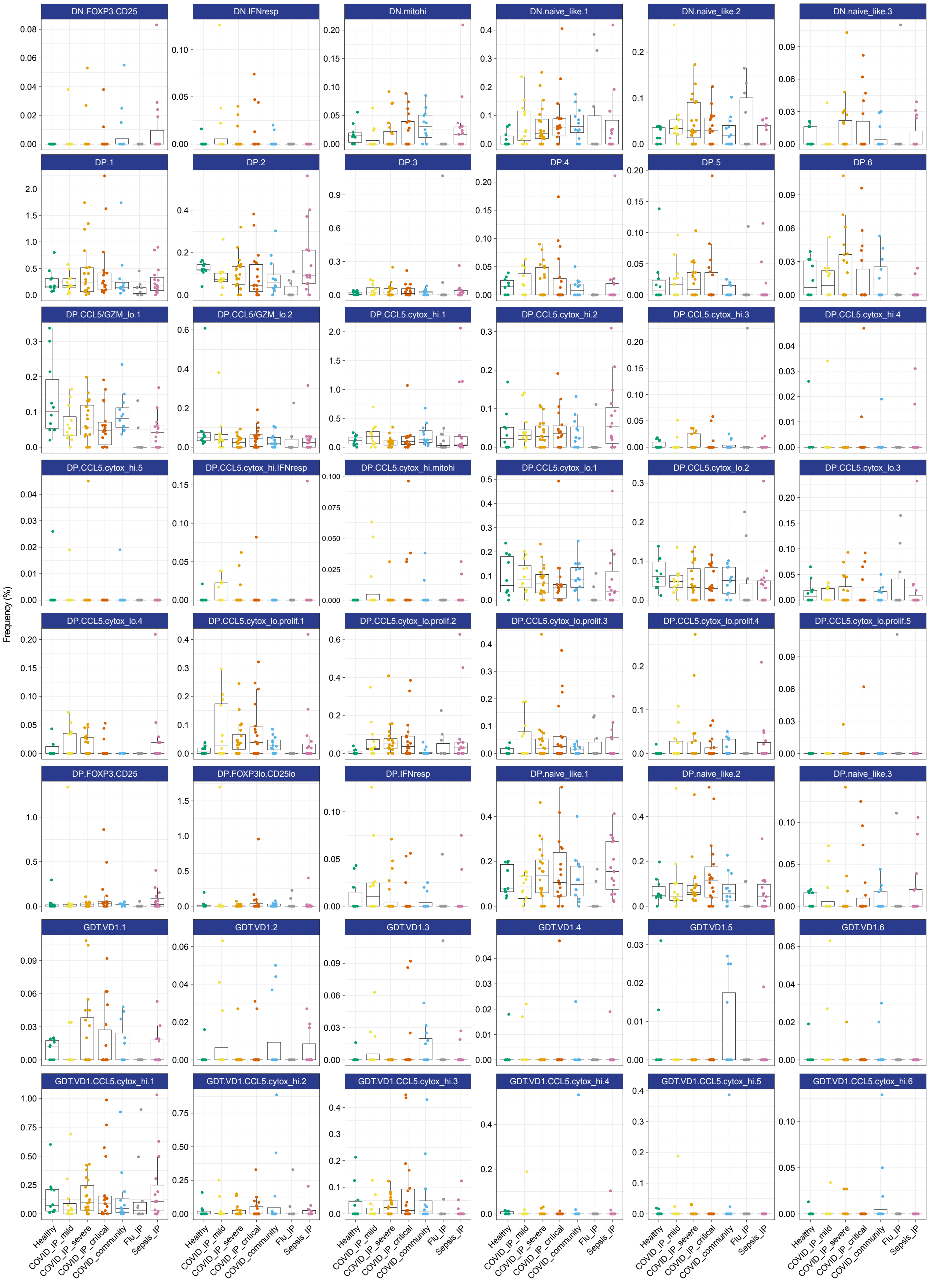

Abbreviations: cytox: cytotoxic; DN: CD4/CD8 double negative; DP: CD4/CD8: double positive; GDT: gamma delta T; GZM: granzyme; hi: high; IP: in-patient; lo: low; mito: mitochondrial; prolif: proliferating; resp: responsive

- Healthy
- COVID_IP_mild
- COVID_IP_severe
- COVID_IP_critical
- COVID_community
- Flu_IP
- Sepsis_IP

### Frequency of clusters out of total T and natural killer cells by clinical category (4)

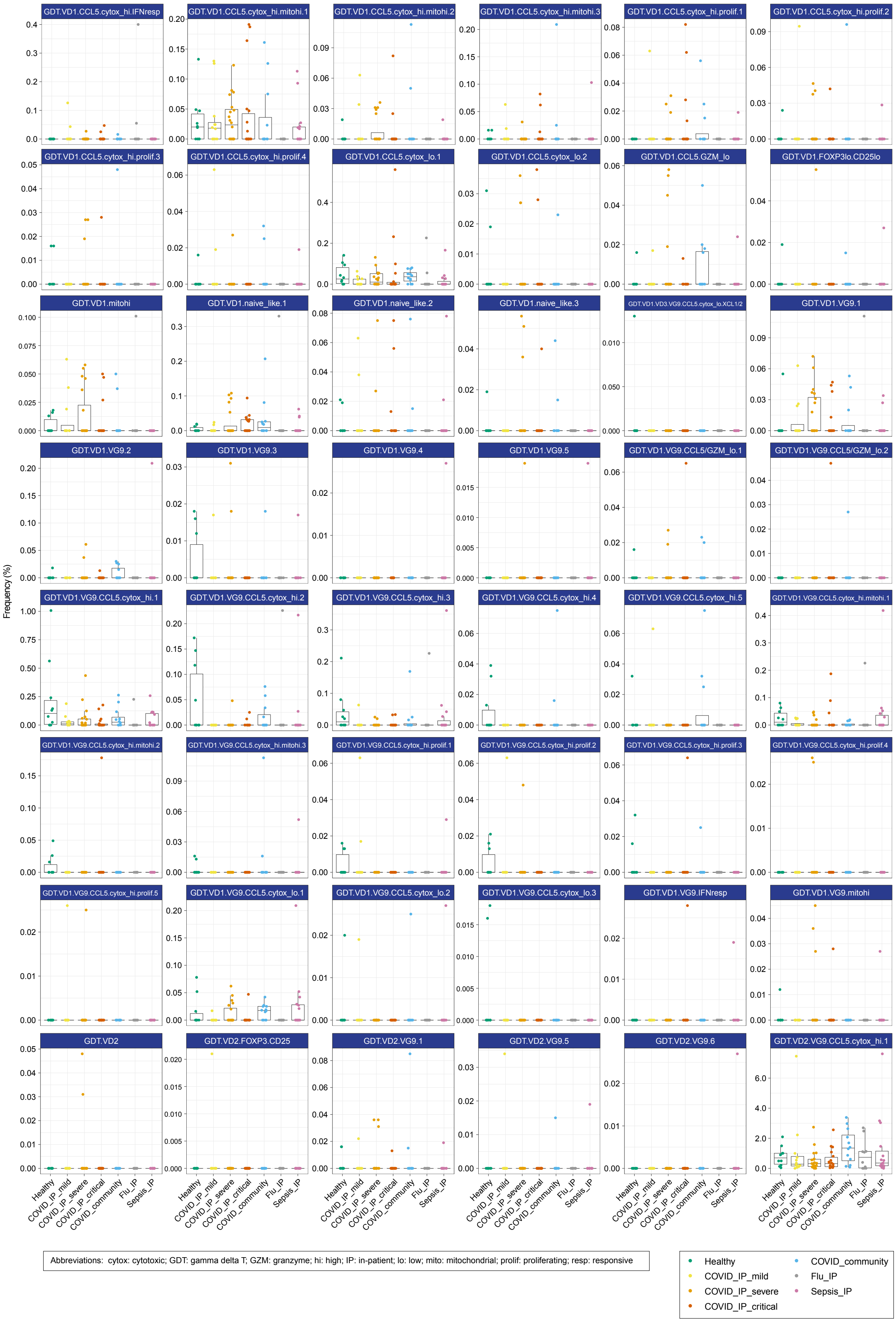

Frequency of clusters out of total T and natural killer cells by clinical category (5)

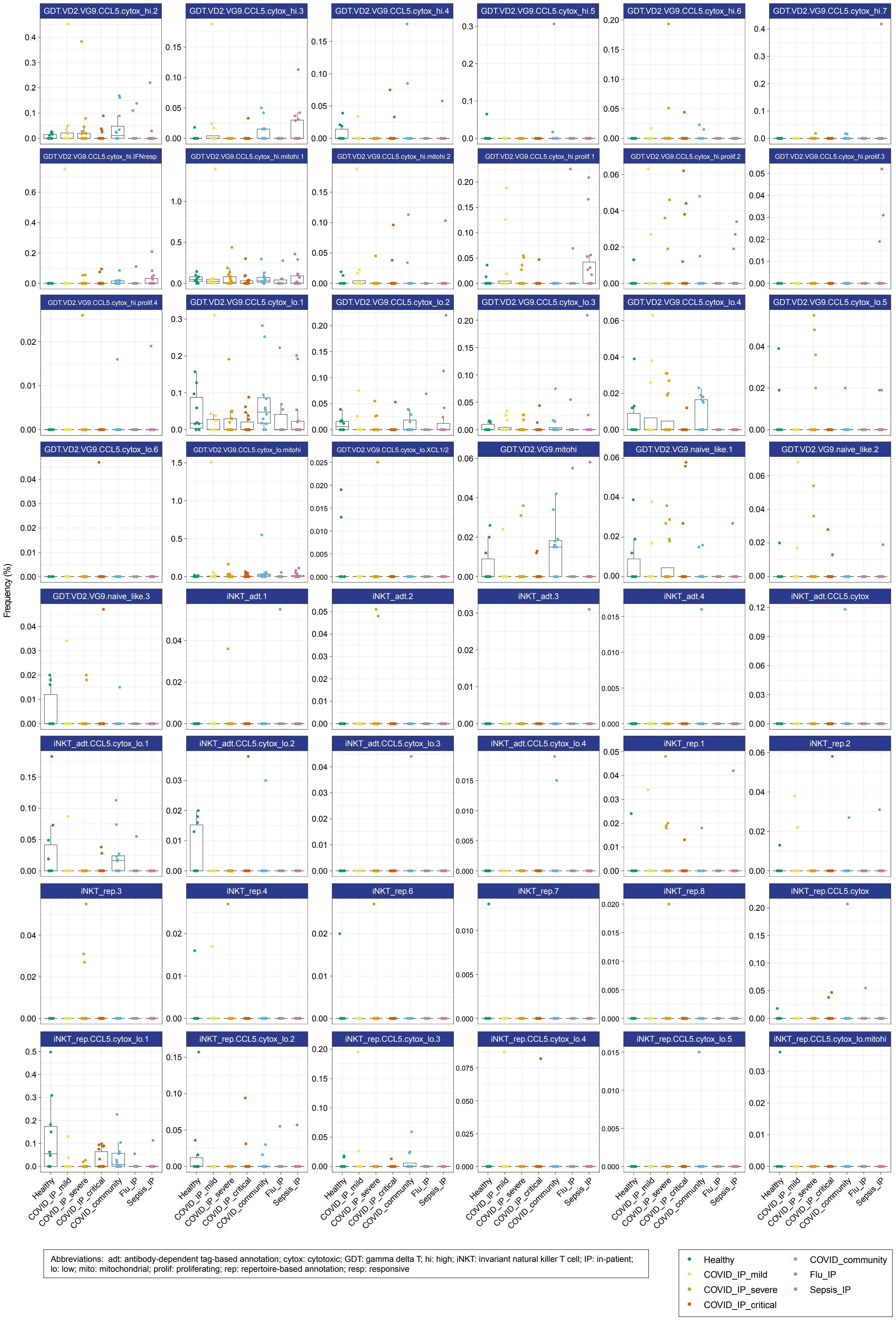

Abbreviations: adt: antibody-dependent tag-based annotation; cytox: cytotoxic; GDT: gamma delta T; hi: high; iNKT: invariant natural killer T cell; IP: in-patient; lo: low; mito: mitochondrial; prolif: proliferating; rep: repertoire-based annotation; resp: responsive

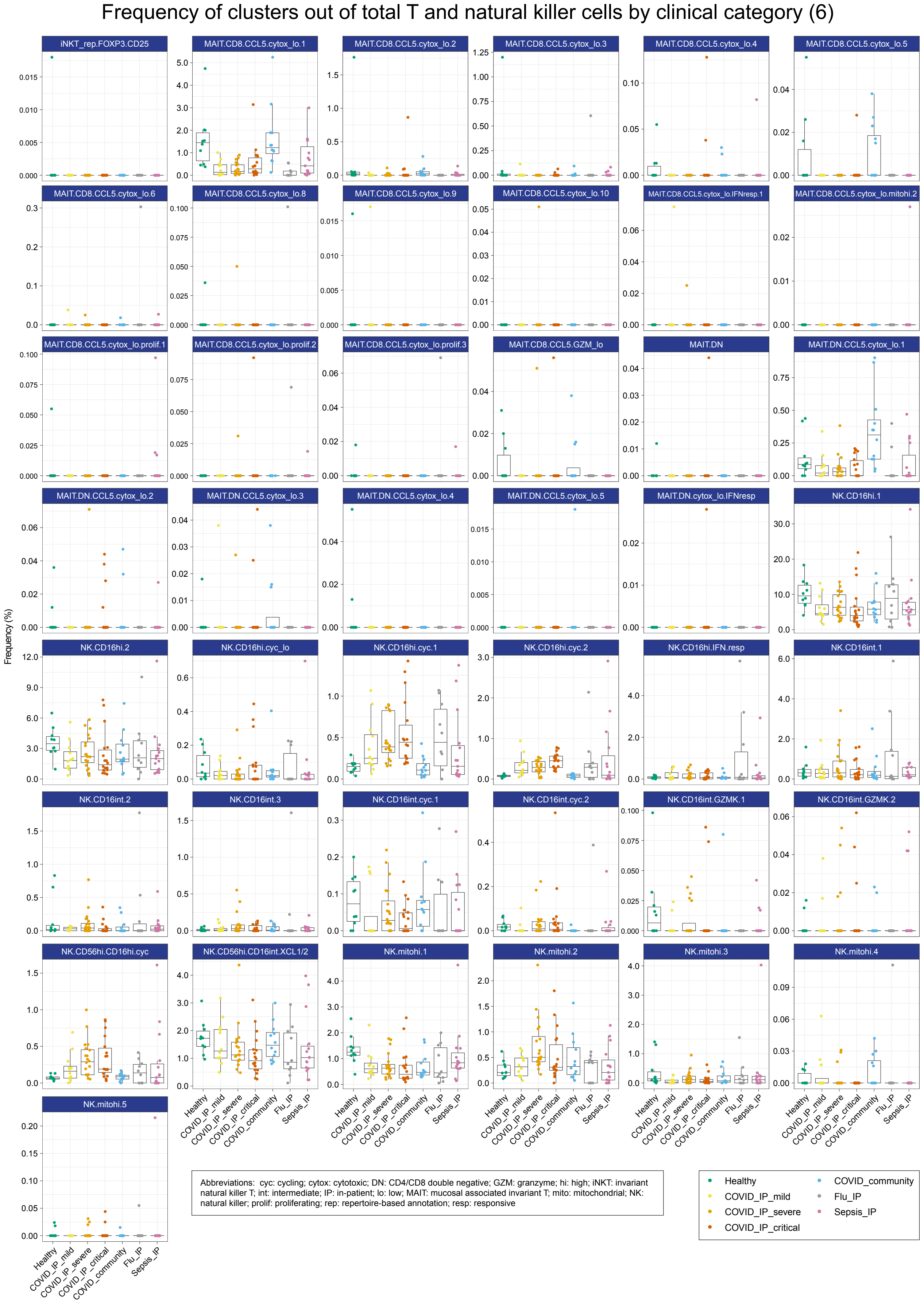

Figure 29: T and NK cluster frequency by clinical category. Box plots show the median and the first and third quartiles; whiskers show 1.5 × the interquartile range above and below the box. Clusters are ordered alphabetically.

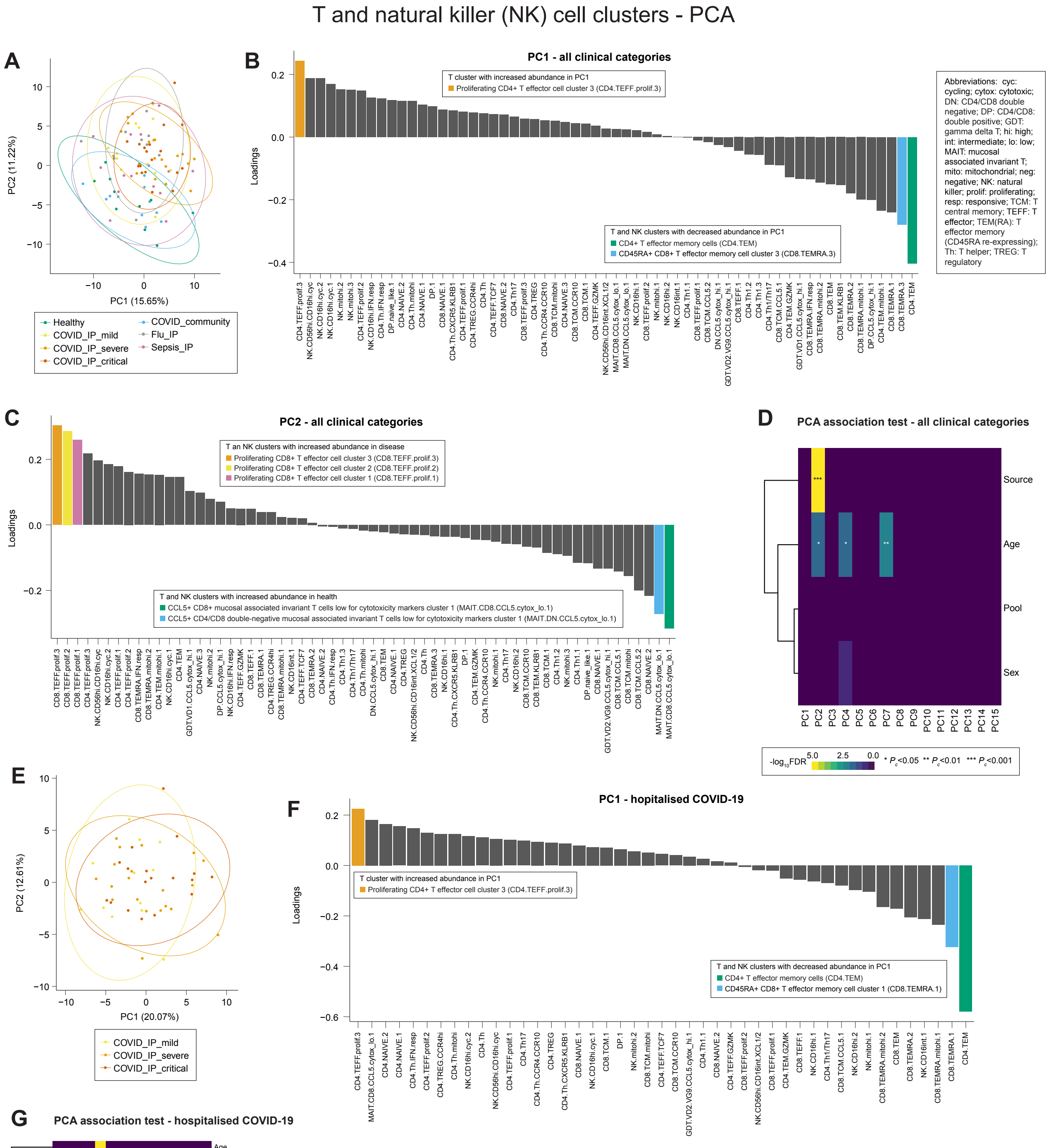

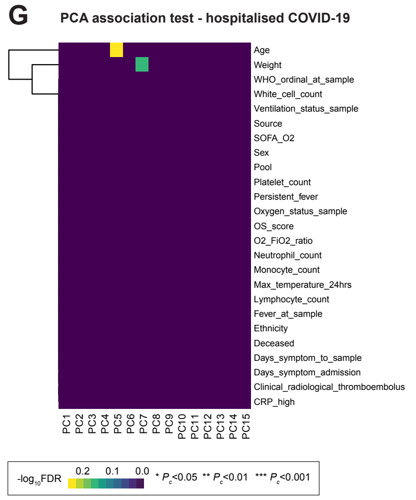

Figure 30: T and NK cluster PCA and association tests.

(A) PCA plot for all clinical categories. Percentage of variance explained by each PC is shown in the brackets. (B) Loadings of T and NK clusters on PC1 from analysis including all clinical categories. (C) Loadings of T and NK clusters on PC2 (associated with source as shown in (D)), from analysis including all clinical categories. (D) Association test between top PCs and source (clinical category), age, sex, and sample pool for all clinical categories. (E) PCA plot for hospitalised COVID-19. Percentage of variance explained by each PC is shown in the brackets. (F) Loadings of T and NK clusters on PC1 from analysis of hospitalised COVID-19 cases. (G) Association test between top PCs and clinical, demographic and experimental variables for hospitalised COVID-19 cases. Significance tested using Benjamini-Hochberg-corrected ANOVA.

T and natural killer (NK) cell clusters - edgeR differential abundance analysis

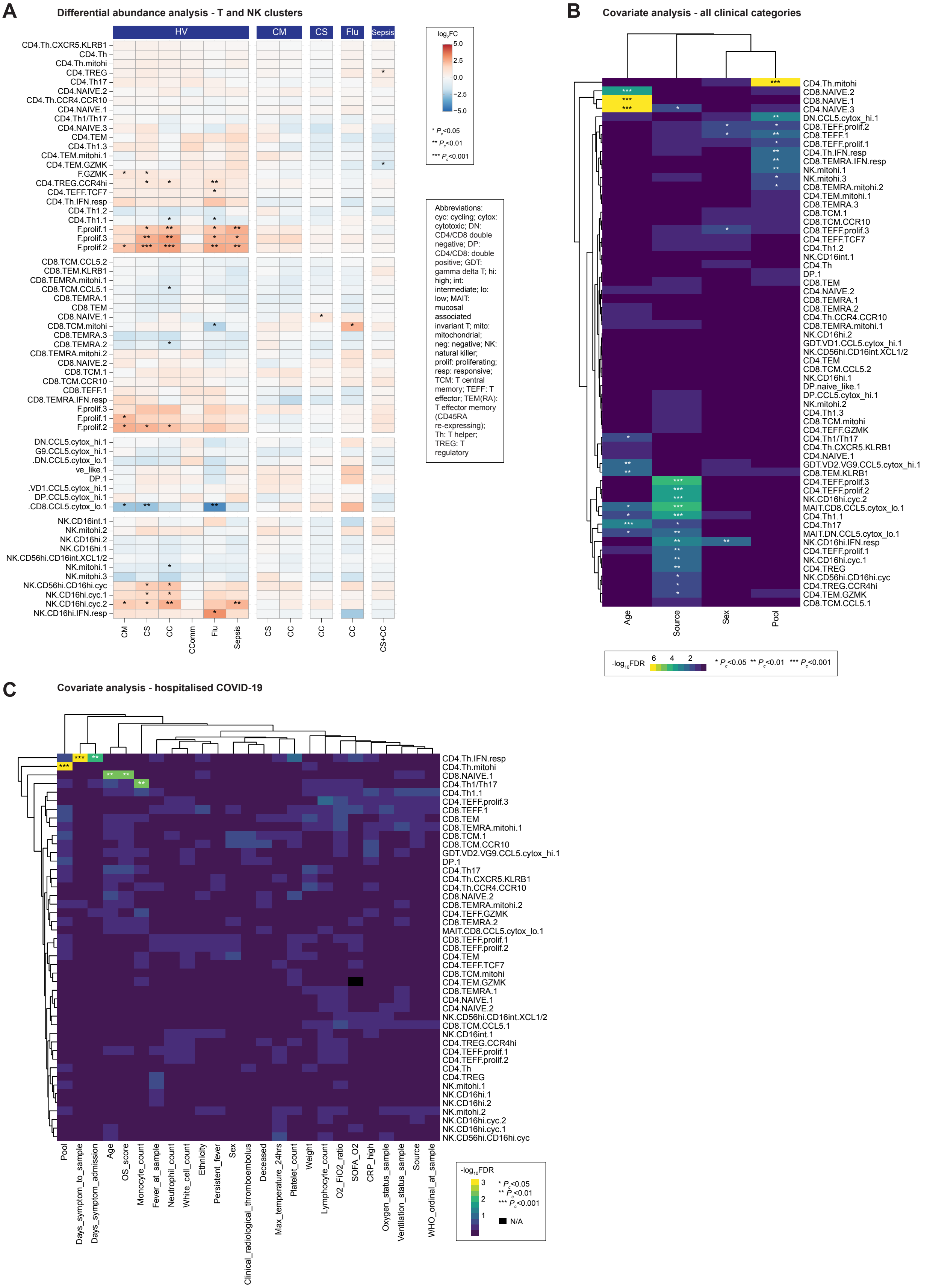

Figure 31: T and NK cluster differential abundance analysis. (A) T and NK cluster differential abundance analysis between clinical categories, controlling for age, sex, and sample pool. (B) T and NK cluster covariate analysis for source (clinical category), age, sex and pool for all clinical categories. (C) T and NK cluster covariate analysis for clinical, demographic and experimental variables for hospitalised COVID-19 cases. Significance tested using Benjamini-Hochberg-corrected ANOVA.

#### Composition analysis of B and plasmablast cell clusters

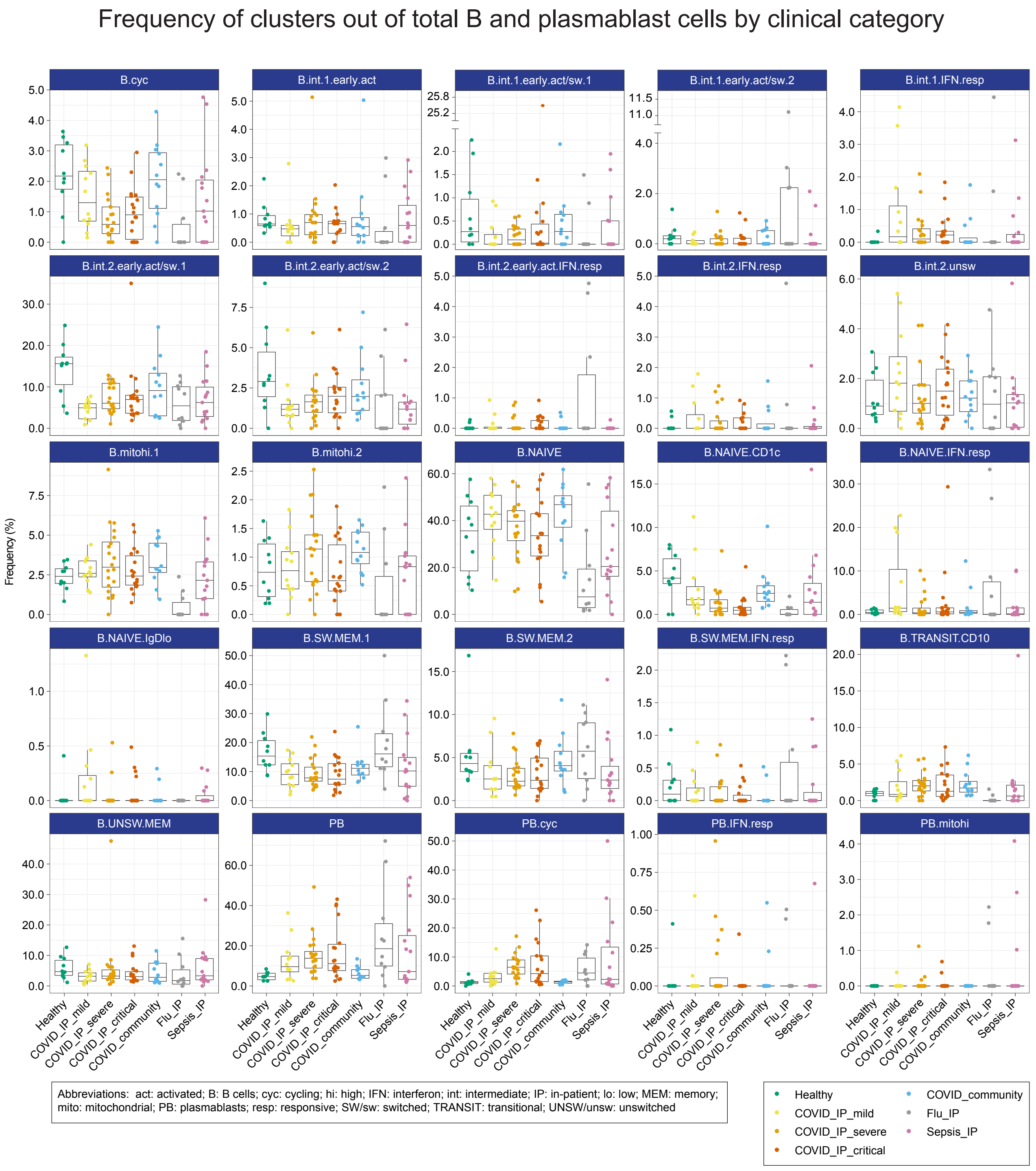

Figure 32: B and PB cluster frequency by clinical category. Box plots show the median and the first and third quartiles; whiskers show 1.5 × the interquartile range above and below the box. Clusters are ordered alphabetically.

B and plasmablast (PB) cell clusters - PCA

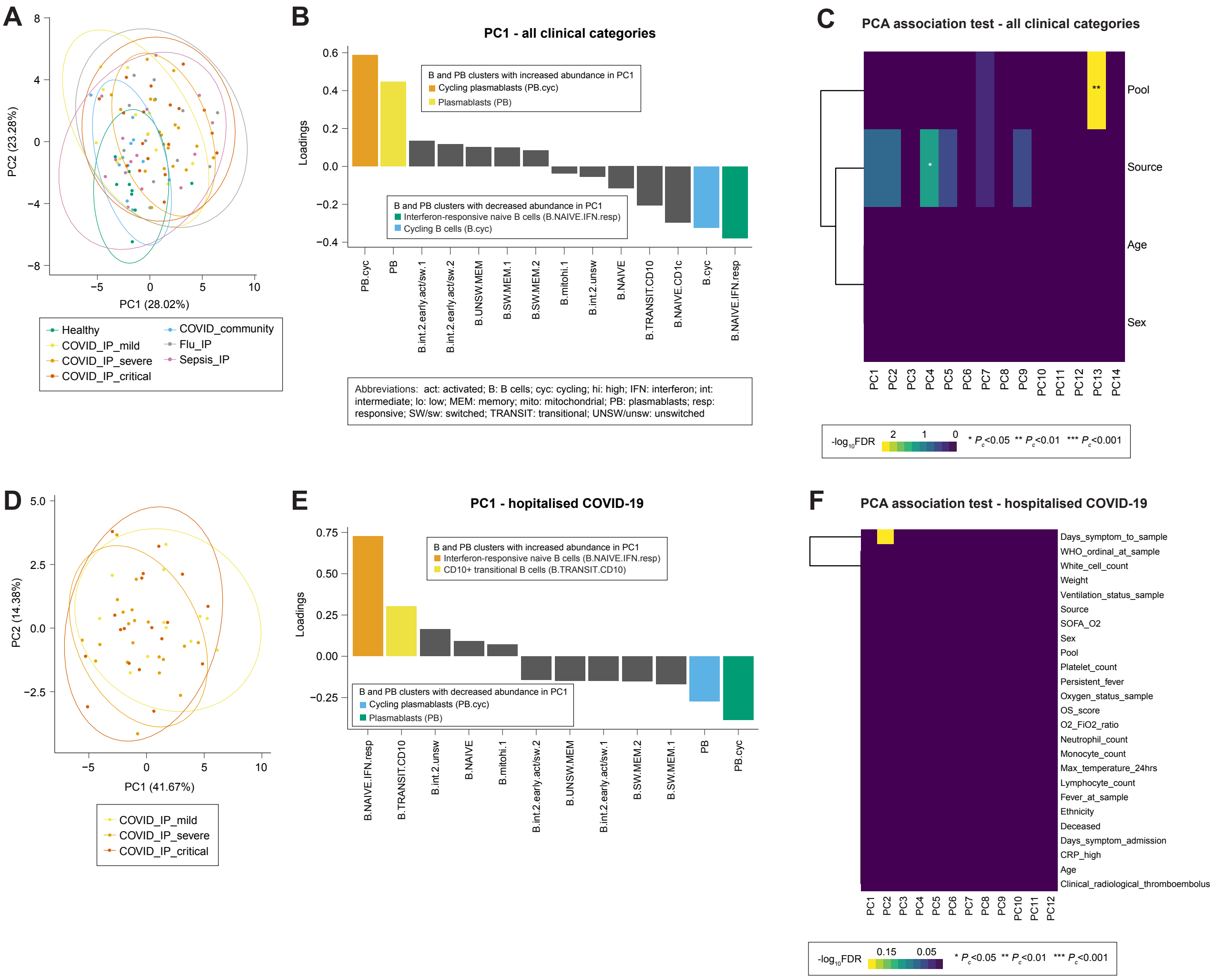

Figure 33: B and PB PCA and association tests. (A) PCA plot for all clinical categories. Percentage of variance explained by each PC is shown in the brackets. (B) Loadings of B and PB clusters on PC1 from analysis including all clinical categories. (C) Association test between top PCs and source (clinical category), age, sex, and sample pool for all clinical categories. (D) PCA plot for hospitalised COVID-19. Percentage of variance explained by each PC is shown in the brackets. (E) Loadings of B and PB clusters on PC1 from analysis of hospitalised COVID-19 cases. (F) Association test between top PCs and clinical, demographic and experimental variables for hospitalised COVID-19 cases. Significance tested using Benjamini-Hochberg-corrected ANOVA.

B and plasmablast (PB) cell clusters - edgeR differential abundance analysis

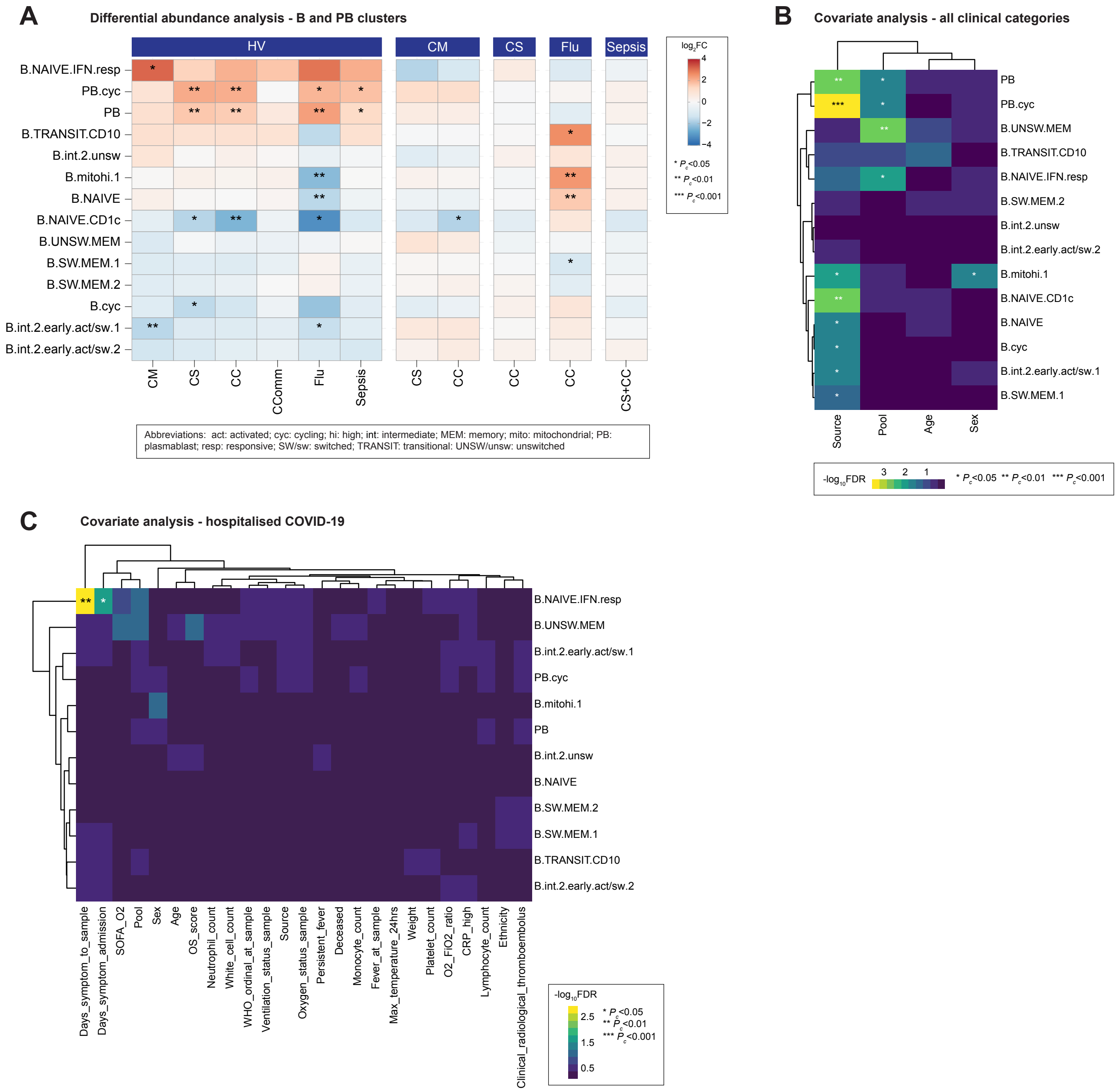

Figure 34: B and PB cluster differential abundance analysis. (A) B and PB cluster differential abundance analysis between clinical categories, controlling for age, sex and sample pool. (B) B and PB cluster covariate analysis for source (clinical category), age, sex and pool for all clinical categories. (C) B and PB cluster covariate analysis for clinical, demographic and experimental variables for hospitalised COVID-19 cases. Significance tested using Benjamini-Hochberg-corrected ANOVA.

#### Composition analysis of mononuclear phagocyte clusters

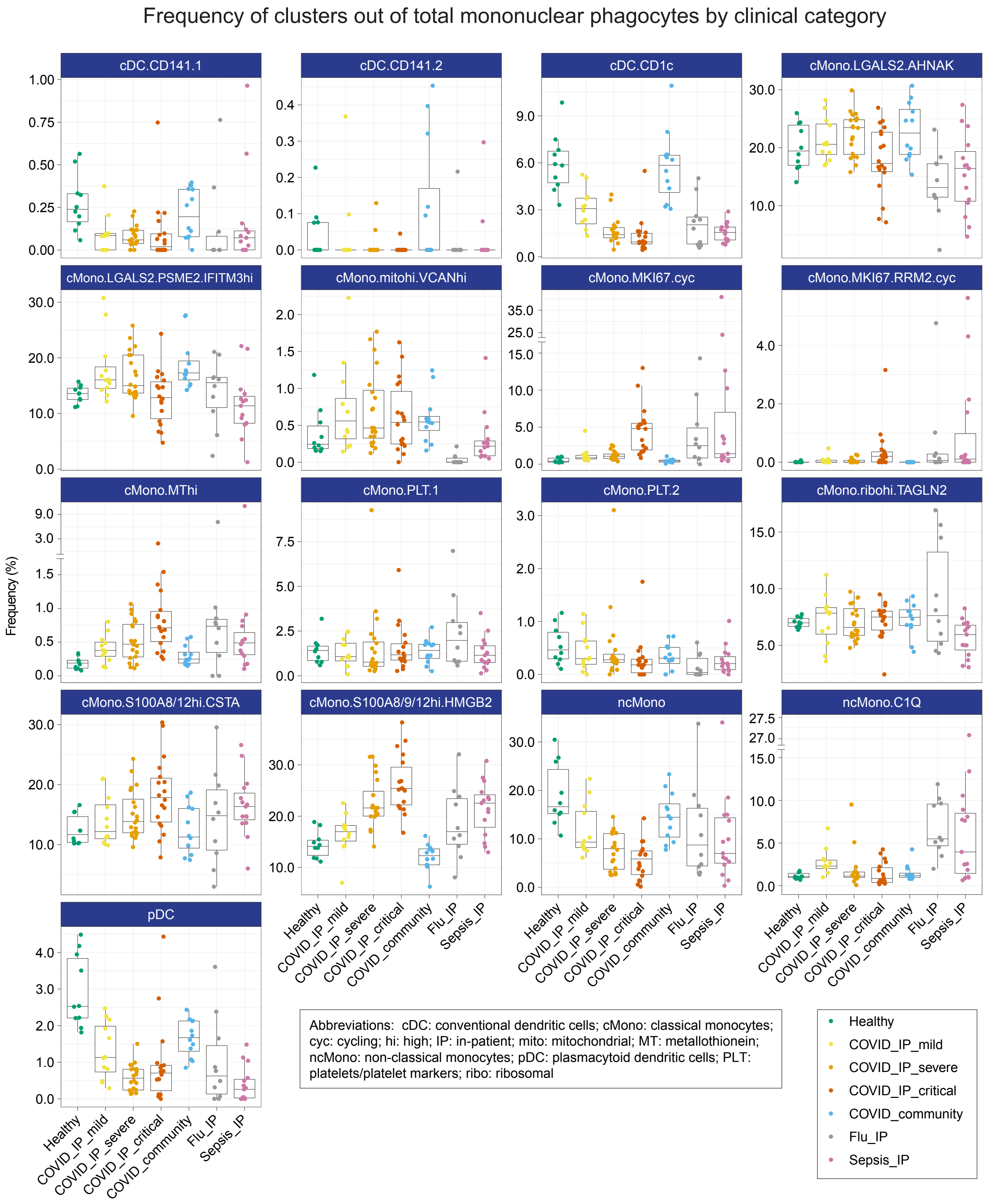

Figure 35: MNP cluster frequency by clinical category. Box plots show the median and the first and third quartiles; whiskers show 1.5 × the interquartile range above and below the box. Clusters are ordered alphabetically.

Mononuclear phagocyte (MNP) clusters - PCA

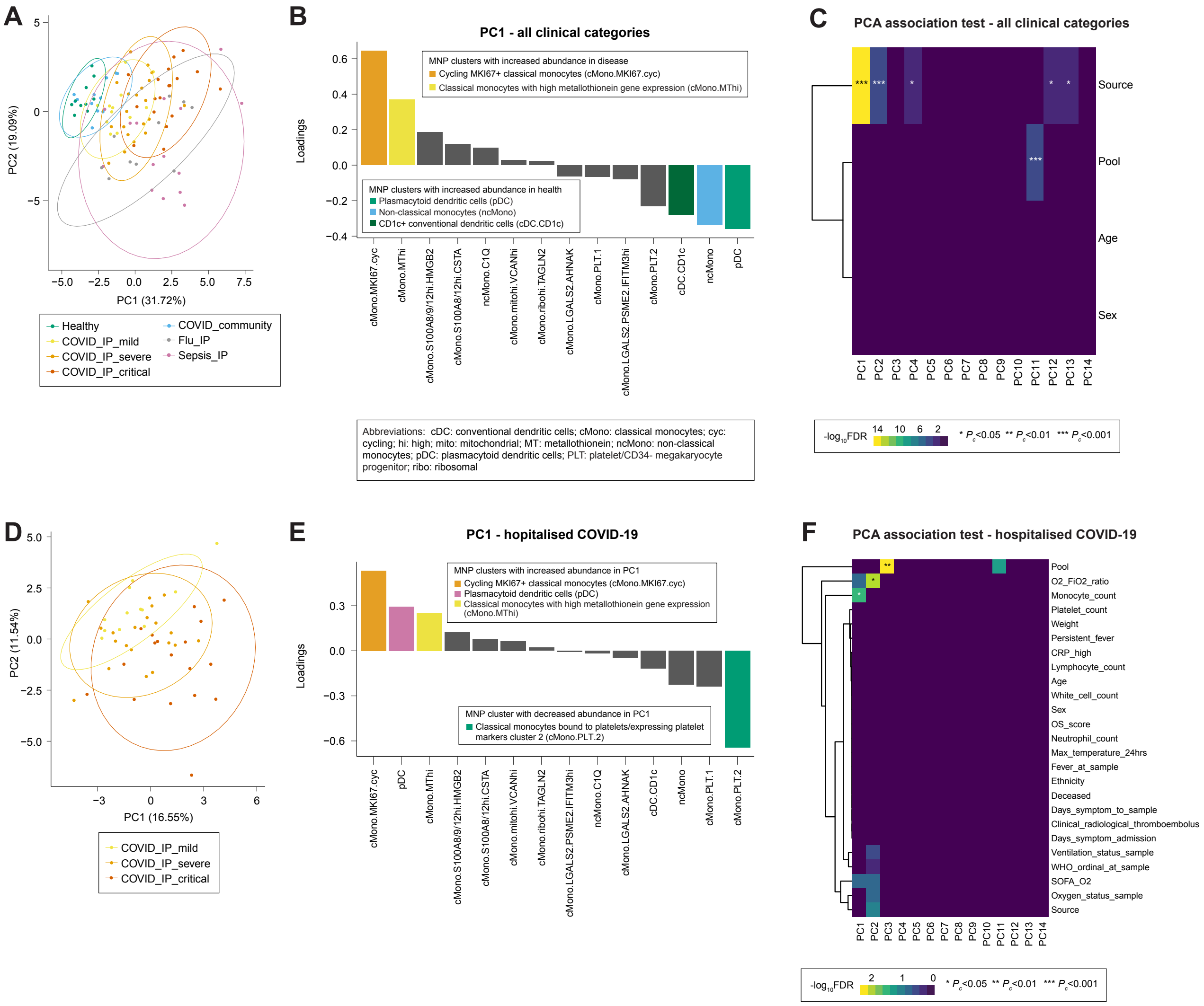

Figure 36: MNP cluster PCA and association tests. (A) PCA plot for all clinical categories. Percentage of variance explained by each PC is shown in the brackets. (B) Loadings of MNP clusters on PC1 from analysis including all clinical categories. (C) Association test between top PCs and source (clinical category), age, sex, and sample pool for all clinical categories. (D) PCA plot for hospitalised COVID-19. Percentage of variance explained by each PC is shown in the brackets. (E) Loadings of MNP clusters on PC1 from analysis of hospitalised COVID-19 cases. (F) Association test between top PCs and clinical, demographic and experimental variables for hospitalised COVID-19 cases. Significance tested using Benjamini-Hochberg-corrected ANOVA.

Mononuclear phagocyte (MNP) clusters - edgeR differential abundance analysis

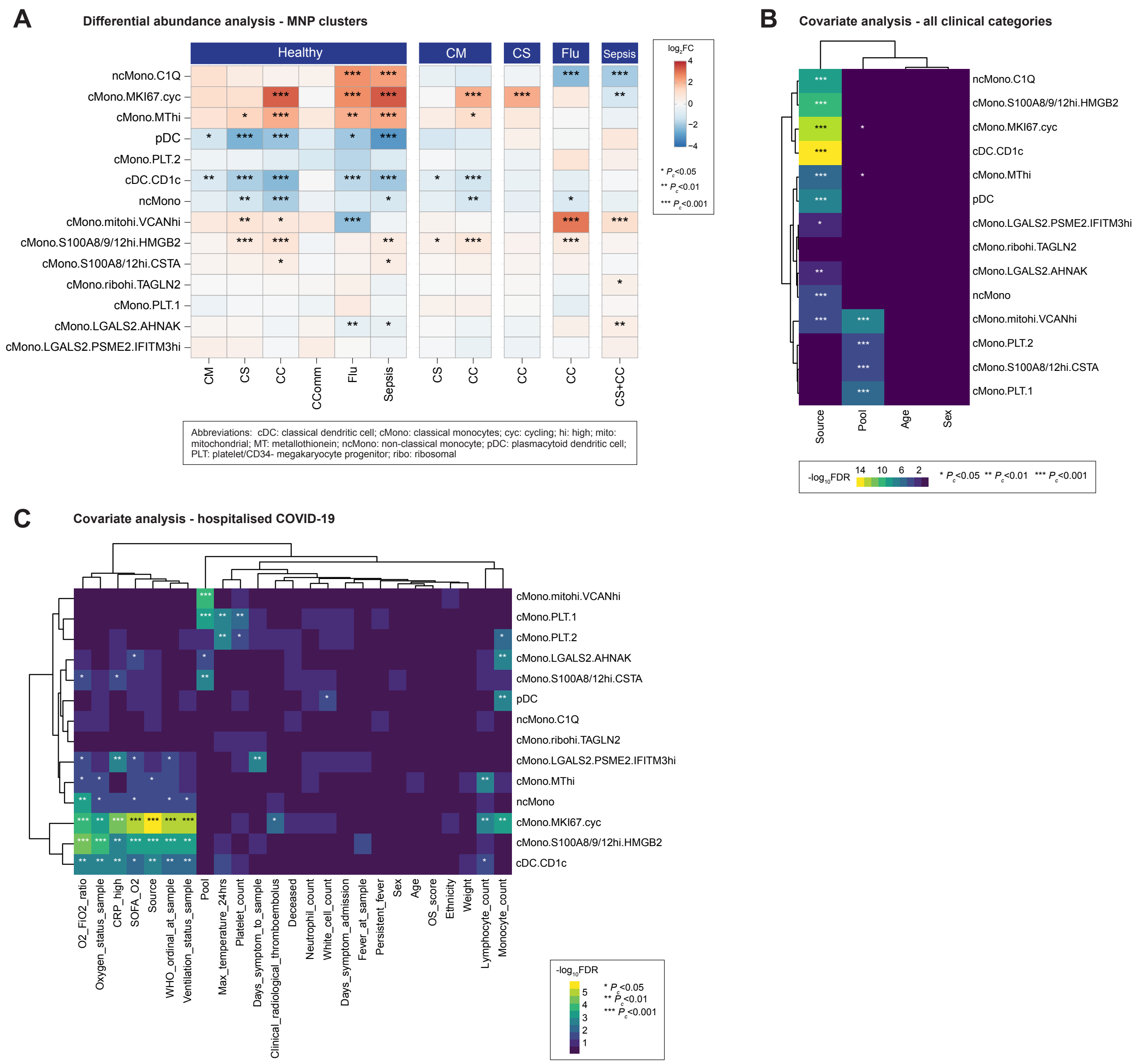

Figure 37: MNP cluster differential abundance analysis. (A) MNP cluster differential abundance analysis between clinical categories, controlling for age, sex and sample pool. (B) MNP cluster covariate analysis for source (clinical category), age, sex and pool for all clinical categories. (C) MNP cluster covariate analysis for clinical, demographic and experimental variables for hospitalised COVID-19 cases. Significance tested using Benjamini-Hochberg-corrected ANOVA.

### Principle Components Analysis

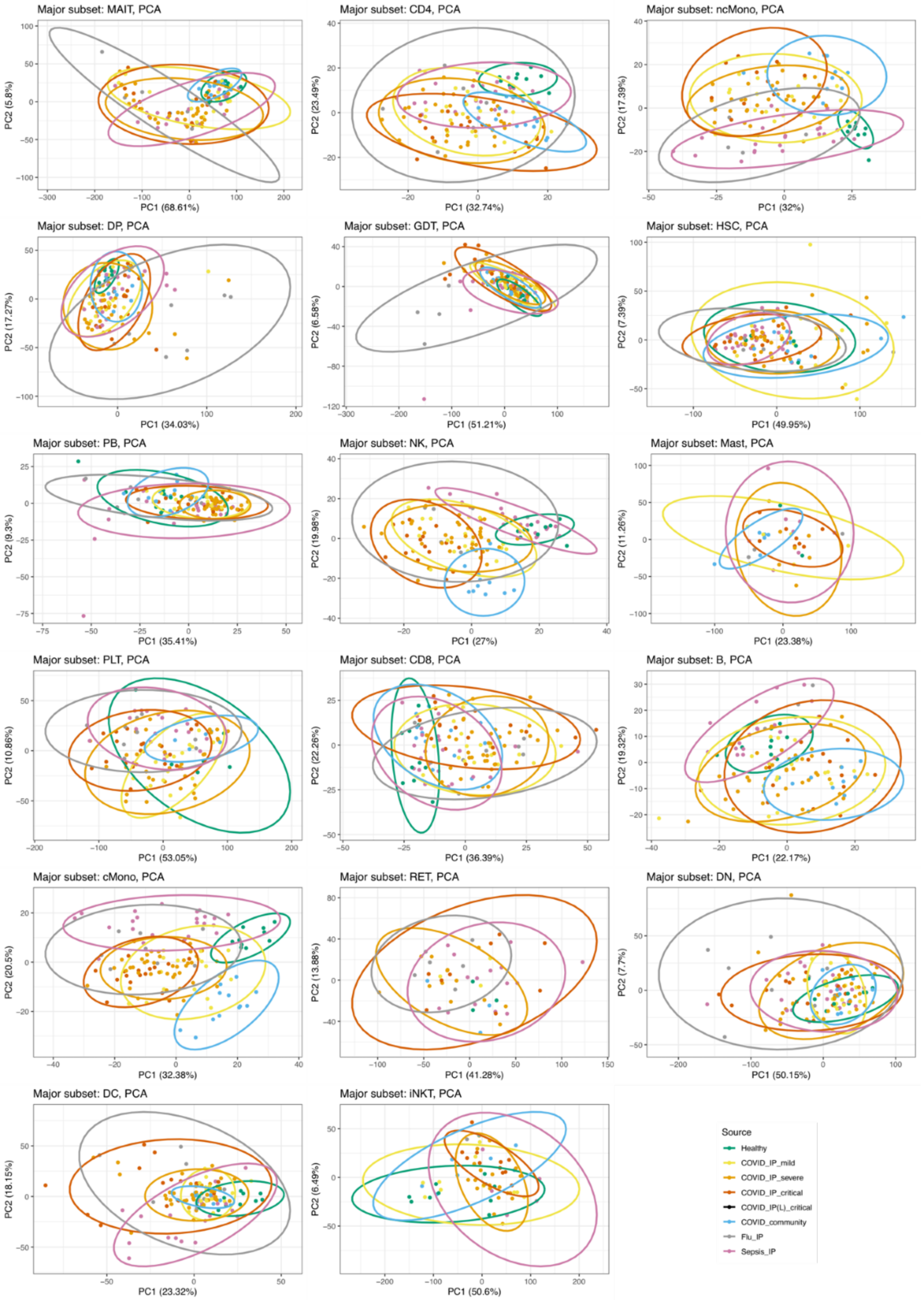

Figure 38: The first two principle components of pseudobulk gene expression for each of the major subsets, coloured by sample source.

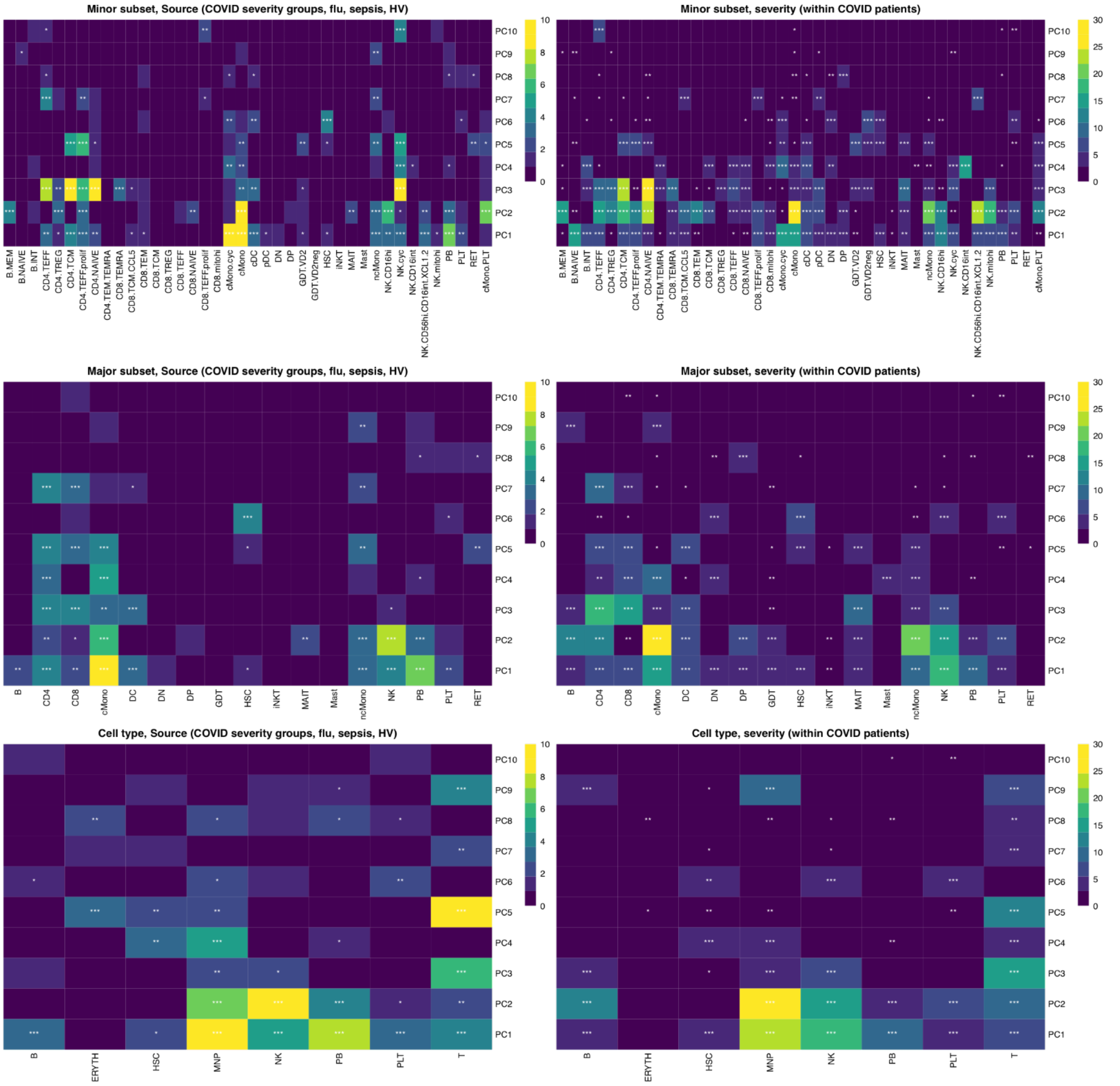

Figure 39: Association with Source and COVID Severity for each of the first 10 PCs in each cluster, grouped by minor subset, major subset and cell type. The colour shows the -log10 transformation of the Benjamini-Hochberg corrected omnibus p-value, capped at 10 (for Source) and 30 (for severity). *** p < 0.001, ** p < 0.01, * p < 0.05.

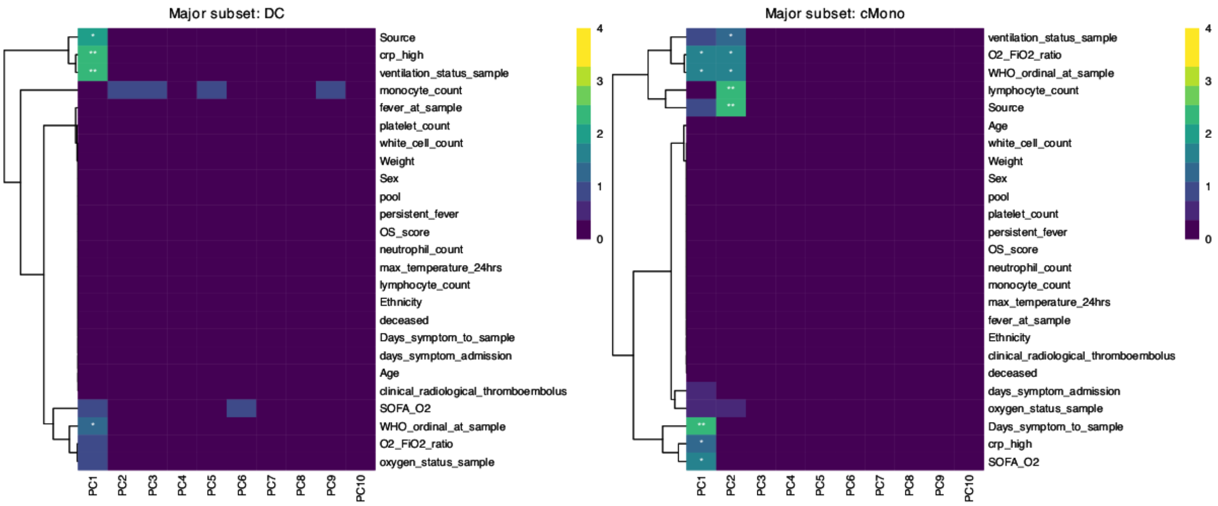

Figure 40: Association of principle components with clinical within hospitalized patients. The colour shows the -log10 transformation of the Benjamini-Hochberg corrected omnibus p-value. *** $p < 0.001$, ** $p < 0.01$, * $p < 0.05$.

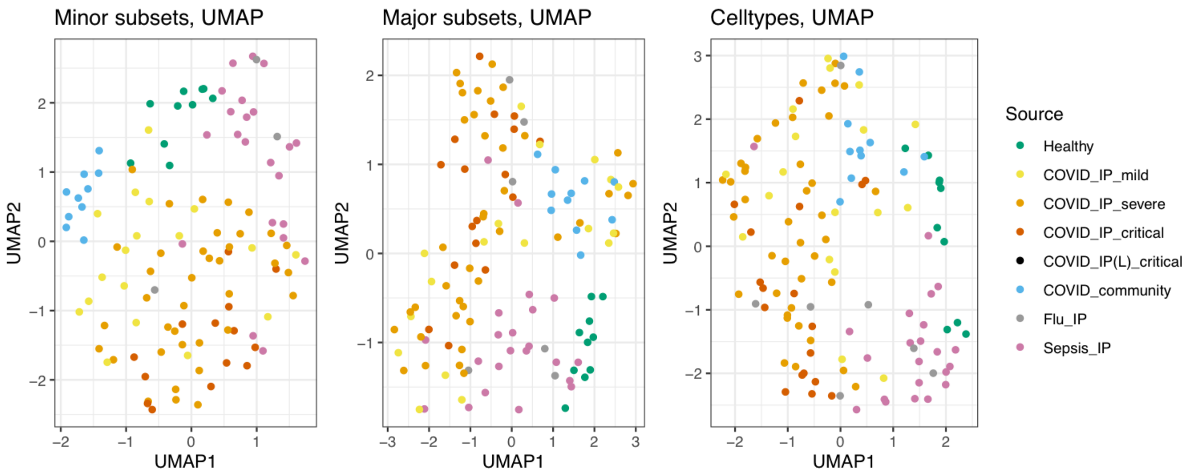

Figure 41: UMAP plots of gene expression, based on combining PCs across all clusters at the minor subset, major subset or cell type levels.

#### Differential Expression Analysis

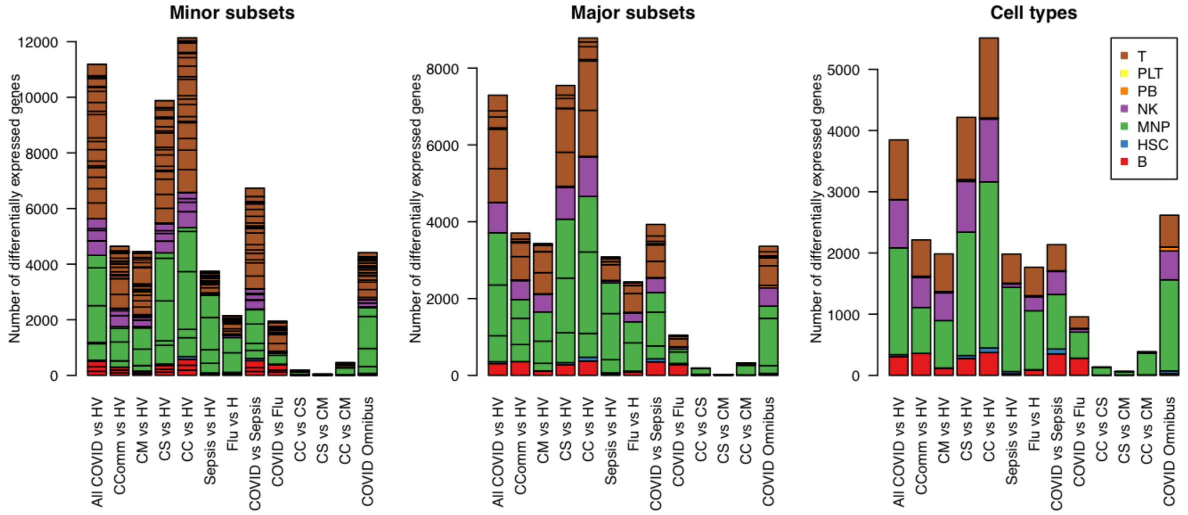

Figure 42: The numbers of differentially expressed genes for different contrasts by cell cluster, coloured by cell type. The size of each individual rectangle gives the number of differentially expressed genes (FDR < 0.01, FC > 2) for a particular cell type for a given contrast, with cell types grouped and coloured by cell lineage.

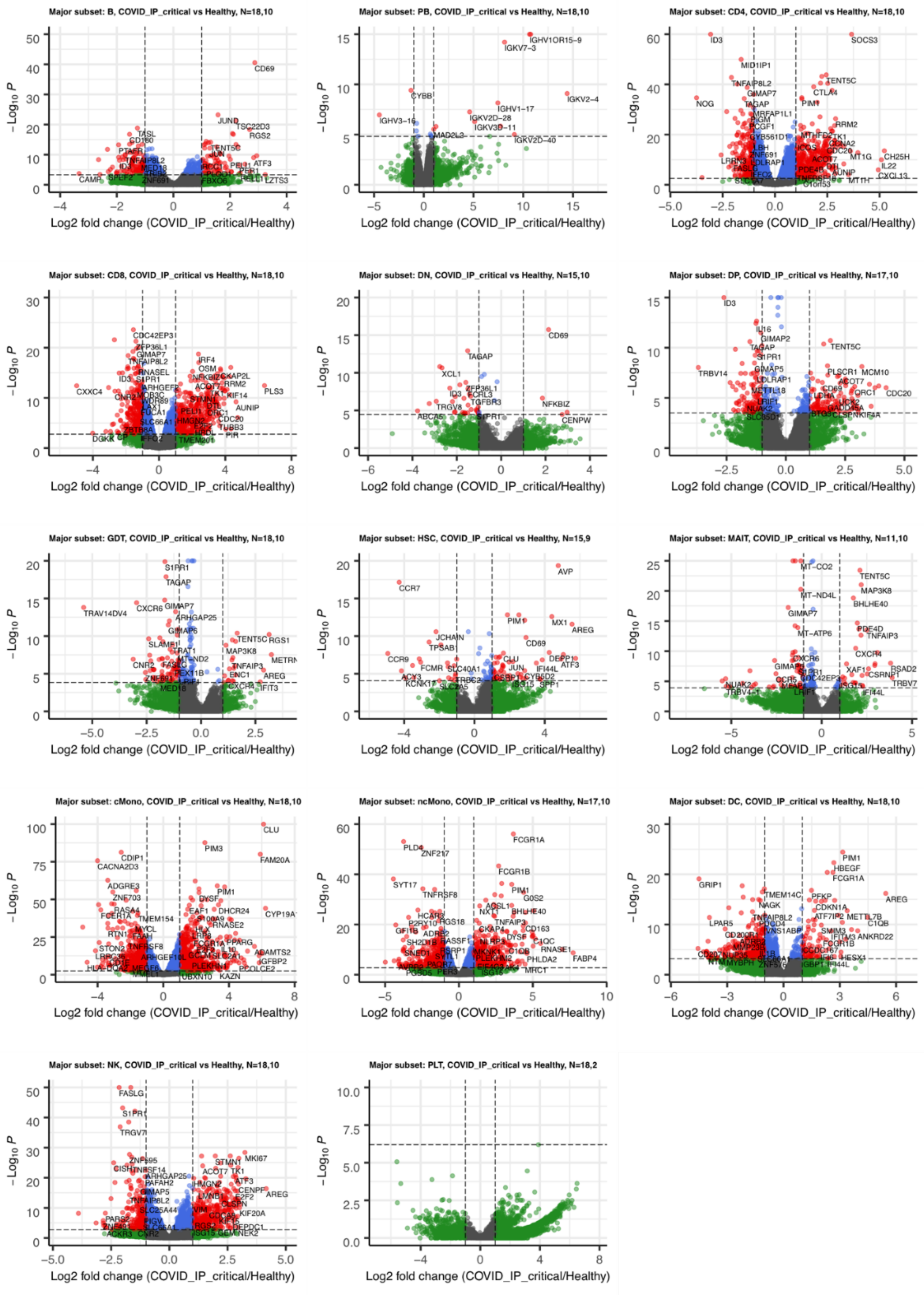

Figure 43: Volcano plots of differential expression for critical COVID vs heathy controls for each of the major subsets. The N given in each title refers to the number COVID patients and number of controls with a cell count high enough to be included in each analysis.

**All COVID vs Healthy Volunteers:**

| HGNC.symbol | Ensembl.gene.ID | celltype | logFC | PValue | QValue |
|---|---|---|---|---|---|
| HBB | ENSG00000244734 | cMono_PLT_minor | -3.68 | 8.52e-148 | 1.1e-143 |
| MT-CO1 | ENSG00000198804 | CD8_TEM_minor | 0.723 | 1.68e-120 | 2.24e-116 |
| GNLY | ENSG00000115523 | CD8_TEFF_minor | -2.01 | 3.83e-109 | 4.63e-105 |
| ID3 | ENSG00000117318 | CD4_major | 2.86 | 5.32e-66 | 6.43e-62 |
| ID3 | ENSG00000117318 | T_celltype | 2.71 | 5.83e-62 | 7.04e-58 |
| ID3 | ENSG00000117318 | CD4_TCM_minor | 2.99 | 6.11e-61 | 7.69e-57 |
| S1PR1 | ENSG00000170989 | NK_mitohi_minor | 2.25 | 2.25e-59 | 1.45e-55 |
| MT-CO1 | ENSG00000198804 | GDT_VD2_minor | 0.751 | 3.68e-59 | 4.64e-55 |
| ZNF217 | ENSG00000171940 | MNP_celltype | 1.72 | 5.97e-58 | 7.35e-54 |
| MID1IP1 | ENSG00000165175 | CD4_NAIVE_minor | 1.56 | 7.74e-54 | 9.8e-50 |

**COVID_community vs Healthy:**

| HGNC.symbol | Ensembl.gene.ID | celltype | logFC | PValue | QValue |
|---|---|---|---|---|---|
| IGHV3-69-1 | ENSG00000282600 | PB_celltype | 7.93 | 2.35e-96 | 2.76e-92 |
| IGHV3-69-1 | ENSG00000282600 | PB_minor | 7.85 | 1.64e-92 | 1.92e-88 |
| IGHV3-69-1 | ENSG00000282600 | PB_major | 7.85 | 1.84e-92 | 2.16e-88 |
| IGHV1-14 | ENSG00000253709 | PB_celltype | -15.5 | 3e-87 | 1.76e-83 |
| IGHV1-14 | ENSG00000253709 | PB_major | -15.5 | 3.06e-87 | 1.8e-83 |
| IGHV1-14 | ENSG00000253709 | PB_minor | -15.5 | 3.07e-87 | 1.81e-83 |
| NR4A1 | ENSG00000123358 | cDC_minor | -3.35 | 2.6e-54 | 3.2e-50 |
| NR4A1 | ENSG00000123358 | DC_major | -3.55 | 8.98e-46 | 1.13e-41 |
| PMAIP1 | ENSG00000141682 | B_celltype | -2.42 | 2.31e-41 | 2.87e-37 |
| PMAIP1 | ENSG00000141682 | B_major | -2.42 | 2.47e-41 | 3.08e-37 |

**COVID_IP_mild vs Healthy:**

| HGNC.symbol | Ensembl.gene.ID | celltype | logFC | PValue | QValue |
|---|---|---|---|---|---|
| MT-CO1 | ENSG00000198804 | CD8_TEM_minor | 0.7 | 0 | 0 |
| ID3 | ENSG00000117318 | CD4_NAIVE_minor | 2.81 | 1.61e-109 | 1.89e-105 |
| ID3 | ENSG00000117318 | CD4_major | 2.71 | 4.21e-96 | 4.9e-92 |
| ID3 | ENSG00000117318 | CD4_TCM_minor | 2.86 | 4.73e-82 | 5.58e-78 |
| CD69 | ENSG00000110848 | CD4_NAIVE_minor | -2.01 | 4.57e-69 | 2.69e-65 |
| G0S2 | ENSG00000123689 | ncMono_major | -4.56 | 2.89e-52 | 3.52e-48 |
| G0S2 | ENSG00000123689 | ncMono_minor | -4.56 | 2.89e-52 | 3.52e-48 |
| PIM3 | ENSG00000198355 | MNP_celltype | -2.21 | 1.49e-50 | 1.76e-46 |
| PRDM1 | ENSG00000057657 | CD4_TCM_minor | -1.38 | 1.9e-47 | 1.12e-43 |
| PIM3 | ENSG00000198355 | cMono_minor | -1.99 | 4.96e-46 | 5.84e-42 |

**COVID_IP_severe vs Healthy:**

| HGNC.symbol | Ensembl.gene.ID | celltype | logFC | PValue | QValue |
|---|---|---|---|---|---|
| MT-CO3 | ENSG00000198938 | CD8_TCM_CCL5_minor | 1 | 0 | 0 |
| RPS26 | ENSG00000197728 | CD8_TEM_minor | 0.701 | 0 | 0 |
| GAPDH | ENSG00000111640 | CD8_TEFF_prolif_minor | -0.819 | 2.57e-162 | 3.11e-158 |
| ID3 | ENSG00000117318 | CD4_NAIVE_minor | 3.5 | 4.99e-142 | 6e-138 |
| RPL29 | ENSG00000162244 | MAIT_minor | 0.707 | 2.56e-137 | 2.95e-133 |
| RPL29 | ENSG00000162244 | MAIT_major | 0.707 | 2.62e-137 | 3.02e-133 |
| ID3 | ENSG00000117318 | CD4_major | 3.07 | 3.9e-127 | 4.59e-123 |
| MT-CO1 | ENSG00000198804 | CD8_TEMRA_minor | 0.87 | 8.21e-127 | 3.39e-123 |
| ID3 | ENSG00000117318 | CD4_TCM_minor | 3.07 | 3.32e-122 | 3.99e-118 |
| MT-CO1 | ENSG00000198804 | CD4_TEM_TEMRA_minor | 0.695 | 3.39e-107 | 4.15e-103 |

**COVID_IP_critical vs Healthy:**

| HGNC.symbol | Ensembl.gene.ID | celltype | logFC | PValue | QValue |
|---|---|---|---|---|---|
| IFITM1 | ENSG00000185885 | CD4_TEFF_minor | -0.887 | 0 | 0 |
| CD74 | ENSG00000019582 | cMono_PLT_minor | 1.5 | 0 | 0 |
| MT-CO3 | ENSG00000198938 | MAIT_major | 1.58 | 2e-266 | 2.33e-262 |
| MT-CO3 | ENSG00000198938 | MAIT_minor | 1.58 | 6.41e-266 | 7.47e-262 |
| TXNIP | ENSG00000265972 | B_NAIVE_minor | -1.25 | 1.18e-200 | 1.47e-196 |
| MT-CYB | ENSG00000198727 | MAIT_minor | 1.46 | 1.46e-145 | 8.52e-142 |
| MT-CYB | ENSG00000198727 | MAIT_major | 1.46 | 2.75e-145 | 1.6e-141 |
| TUBA1B | ENSG00000123416 | CD4_TEFF_prolif_minor | -0.873 | 6.6e-133 | 8.21e-129 |
| ID3 | ENSG00000117318 | CD4_NAIVE_minor | 3.57 | 8.5e-119 | 1e-114 |
| ID3 | ENSG00000117318 | CD4_major | 3.08 | 1.21e-116 | 1.42e-112 |

**Sepsis_IP vs Healthy:**

| HGNC.symbol | Ensembl.gene.ID | celltype | logFC | PValue | QValue |
|---|---|---|---|---|---|
| HBB | ENSG00000244734 | cMono_PLT_minor | -1.1 | 9.22-310 | 1.1e-305 |
| FAM20A | ENSG00000108950 | cMono_minor | 6 | 3.36e-72 | 3.98e-68 |
| FAM20A | ENSG00000108950 | cMono_major | 5.93 | 5.47e-70 | 6.51e-66 |
| SCGB3A1 | ENSG00000161055 | CD4_TCM_minor | -7.3 | 4.94e-64 | 5.97e-60 |
| CLU | ENSG00000120885 | cMono_minor | 5.78 | 1.67e-59 | 9.91e-56 |
| CLU | ENSG00000120885 | cMono_major | 5.76 | 5.19e-59 | 3.09e-55 |
| SCGB3A1 | ENSG00000161055 | CD8_NAIVE_minor | -8.8 | 3.05e-56 | 3.72e-52 |
| CLDN5 | ENSG00000184113 | CD4_NAIVE_minor | 5.85 | 6.29e-51 | 7.27e-47 |
| CLU | ENSG00000120885 | MNP_celltype | 5.78 | 3.33e-46 | 4e-42 |
| METTL7B | ENSG00000170439 | cMono_minor | 6.74 | 4.64e-45 | 1.83e-41 |

**Flu_IP vs Healthy:**

| HGNC.symbol | Ensembl.gene.ID | celltype | logFC | PValue | QValue |
|---|---|---|---|---|---|
| ADAMTS2 | ENSG00000087116 | cMono_major | -6.87 | 1.53e-90 | 1.81e-86 |
| ADAMTS2 | ENSG00000087116 | cMono_minor | -6.86 | 4.37e-90 | 5.18e-86 |
| IGKV6-21 | ENSG00000211611 | PB_minor | -7.94 | 2.85e-83 | 3.2e-79 |
| IGKV6-21 | ENSG00000211611 | PB_major | -7.94 | 2.93e-83 | 3.29e-79 |
| IGKV6-21 | ENSG00000211611 | PB_celltype | -7.94 | 2.96e-83 | 3.33e-79 |
| ADAMTS2 | ENSG00000087116 | MNP_celltype | -6.64 | 9.06e-75 | 1.1e-70 |
| VSIG4 | ENSG00000155659 | ncMono_minor | -6.54 | 7.41e-71 | 8.85e-67 |
| VSIG4 | ENSG00000155659 | ncMono_major | -6.54 | 7.42e-71 | 8.85e-67 |
| HLA-DQA1 | ENSG00000196735 | cMono_major | 3.68 | 4.72e-66 | 2.8e-62 |
| HLA-DQA1 | ENSG00000196735 | cMono_minor | 3.63 | 4.33e-58 | 2.57e-54 |

**COVID_IP_critical+COVID_IP_mild vs Sepsis_IP:**

| HGNC.symbol | Ensembl.gene.ID | celltype | logFC | PValue | QValue |
|---|---|---|---|---|---|
| H3-3B | ENSG00000132475 | NK_CD56hi_CD16int_XCL1_2_minor | -1.08 | 5.35e-159 | 6.94e-155 |
| BCDIN3D | ENSG00000186666 | CD4_major | 1.12 | 2.58e-51 | 3.16e-47 |
| BCDIN3D | ENSG00000186666 | T_celltype | 1.16 | 5.67e-46 | 6.91e-42 |
| EEF1A1 | ENSG00000156508 | CD8_mitohi_minor | 0.7 | 3.22e-45 | 2.12e-41 |
| RPS26 | ENSG00000197728 | DN_major | -0.733 | 6.66e-44 | 8.4e-40 |
| RPS26 | ENSG00000197728 | DN_minor | -0.733 | 6.71e-44 | 8.47e-40 |
| GIMAP7 | ENSG00000179144 | CD4_NAIVE_minor | 1.06 | 1.24e-39 | 1.09e-35 |
| GEMIN6 | ENSG00000152147 | CD4_NAIVE_minor | 1.23 | 1.71e-39 | 1.09e-35 |
| BCDIN3D | ENSG00000186666 | CD4_NAIVE_minor | 1.05 | 8.54e-38 | 3.62e-34 |
| CCR7 | ENSG00000126353 | CD4_NAIVE_minor | 1.23 | 9.77e-37 | 3.1e-33 |

**COVID_IP_critical vs flu_IP:**

| HGNC.symbol | Ensembl.gene.ID | celltype | logFC | PValue | QValue |
|---|---|---|---|---|---|
| RPL30 | ENSG00000156482 | DP_minor | -0.698 | 6.45e-67 | 7.83e-63 |
| HLA-DRA | ENSG00000204287 | B_NAIVE_minor | -0.702 | 5.02e-66 | 5.97e-62 |
| MT-CO1 | ENSG00000198804 | NK_CD16int_minor | 2.07 | 8.74e-54 | 8.56e-50 |
| TXNIP | ENSG00000265972 | B_NAIVE_minor | -1.2 | 5.06e-46 | 1.51e-42 |
| RPS15A | ENSG00000134419 | B_NAIVE_minor | -0.787 | 5.3e-45 | 1.26e-41 |
| RPL29 | ENSG00000162244 | B_NAIVE_minor | -0.71 | 3.76e-37 | 7.46e-34 |
| JUN | ENSG00000177606 | T_celltype | -1.89 | 8.24e-32 | 9.89e-28 |
| IGLV7-46 | ENSG00000211649 | B_INT_minor | -14.5 | 1.47e-31 | 1.75e-27 |
| RPL32 | ENSG00000144713 | DP_minor | -0.741 | 2.79e-31 | 1.69e-27 |
| IGLV2-34 | ENSG00000253120 | PB_major | 6.33 | 1.06e-30 | 1.21e-26 |

**COVID_IP_critical vs COVID_IP_severe:**

| HGNC.symbol | Ensembl.gene.ID | celltype | logFC | PValue | QValue |
|---|---|---|---|---|---|
| IGKV6D-21 | ENSG00000225523 | B_MEM_minor | -5.19 | 9.55e-32 | 1.22e-27 |
| TRBV10-2 | ENSG00000229769 | CD8_TEFF_prolif_minor | 6.16 | 5.62e-26 | 6.95e-22 |
| TRAV38-1 | ENSG00000211816 | CD8_TEFF_minor | 11.4 | 1.58e-21 | 1.75e-17 |
| TRBV6-6 | ENSG00000211724 | GDT_major | 8.03 | 1.11e-17 | 1.34e-13 |
| TRBV25-1 | ENSG00000282499 | CD8_TEFF_minor | -11.6 | 1.71e-16 | 9.43e-13 |
| TRBV4-1 | ENSG00000211710 | CD4_TEM_TEMRA_minor | 4.88 | 3.83e-16 | 4.66e-12 |
| TRBV20-1 | ENSG00000211747 | CD8_TEFF_minor | 11.9 | 8.99e-16 | 3.31e-12 |
| HSPA1B | ENSG00000204388 | NK_CD16hi_minor | -3.2 | 1.77e-12 | 1.15e-08 |
| GGH | ENSG00000137563 | cMono_minor | -1.72 | 5.13e-12 | 6.08e-08 |
| GGH | ENSG00000137563 | cMono_major | -1.83 | 1.14e-11 | 7.56e-08 |

**COVID_IP_severe vs COVID_IP_mild:**

| HGNC.symbol | Ensembl.gene.ID | celltype | logFC | PValue | QValue |
|---|---|---|---|---|---|
| SCGB3A1 | ENSG00000161055 | CD4_TREG_minor | -5.09 | 5.11e-29 | 6.55e-25 |
| IGHV1OR16-1 | ENSG00000261704 | PB_minor | -6.4 | 7.52e-17 | 8.88e-13 |
| IGHV1OR16-1 | ENSG00000261704 | PB_celltype | -6.4 | 7.52e-17 | 8.88e-13 |

| | | | | | |
|---|---|---|---|---|---|
| IGHV1OR16-1 | ENSG00000261704 | PB_major | -6.4 | 8.73e-17 | 1.03e-12 |
| FCER1A | ENSG00000179639 | cMono_PLT_minor | -4.04 | 9.45e-15 | 9.83e-11 |
| HBB | ENSG00000244734 | cMono_PLT_minor | 6.64 | 1.62e-14 | 9.83e-11 |
| TRBV5-4 | ENSG00000230099 | CD4_TEM_TEMRA_minor | 8.04 | 3.37e-13 | 4.13e-09 |
| TRBV12-5 | ENSG00000275158 | CD4_TEFF_prolif_minor | -6.23 | 3.91e-12 | 4.95e-08 |
| IGLV1-50 | ENSG00000211645 | B_celltype | 3.22 | 5.44e-12 | 7.1e-08 |
| IGLV1-50 | ENSG00000211645 | B_major | 3.22 | 5.44e-12 | 7.11e-08 |

**COVID_IP_critical vs COVID_IP_severe:**

| HGNC.symbol | Ensembl.gene.ID | celltype | logFC | PValue | QValue |
|---|---|---|---|---|---|
| TRBV3-1 | ENSG00000237702 | CD8_TEFF_minor | -18.5 | 1.75e-37 | 1.93e-33 |
| IGKV6D-21 | ENSG00000225523 | B_MEM_minor | -9.34 | 7.94e-30 | 3.3e-26 |
| TRBV13 | ENSG00000276405 | CD8_TEFF_minor | 27.9 | 4.45e-20 | 2.46e-16 |
| CYP19A1 | ENSG00000137869 | MNP_celltype | -3.34 | 1.17e-17 | 1.41e-13 |
| TPSD1 | ENSG00000095917 | HSC_celltype | 13.9 | 1.34e-17 | 1.65e-13 |
| CYP19A1 | ENSG00000137869 | cMono_major | -3.12 | 8.39e-17 | 1.01e-12 |
| SBNO2 | ENSG00000064932 | CD4_TCM_minor | -1.5 | 4.84e-16 | 5.89e-12 |
| CYP19A1 | ENSG00000137869 | cMono_minor | -3.09 | 3.06e-15 | 3.66e-11 |
| RPS26 | ENSG00000197728 | B_NAIVE_minor | 0.855 | 3.6e-15 | 1.48e-11 |
| IGLV3-12 | ENSG00000211667 | PB_minor | 2.74 | 1.23e-14 | 1.44e-10 |

**COVID omnibus test (all logFCs are given relative to COVID_community):**

| HGNC.symbol | Ensembl.gene.ID | celltype | logFC_CM | logFC_CS | logFC_CC | PValue | QValue |
|---|---|---|---|---|---|---|---|
| CCL5 | ENSG00000271503 | CD8.TCM.CCL5_minor | -0.43 | -0.74 | -1.2 | 0 | 0 |
| FTH1 | ENSG00000167996 | MAIT_major | -1.14 | -1.25 | -0.49 | 0 | 0 |
| FTH1 | ENSG00000167996 | MAIT_minor | -1.14 | -1.25 | -0.52 | 0 | 0 |
| RPS3A | ENSG00000145425 | GDT_VD2neg_minor | -0.84 | -0.45 | -0.78 | 0 | 0 |
| VIM | ENSG00000026025 | cMono.cyc_minor | 0.37 | 0.60 | 0.92 | 3.04e-79 | 3.82e-75 |
| LGALS1 | ENSG00000100097 | cMono.cyc_minor | 0.979 | 1.54 | 1.95 | 1.28e-75 | 8.04e-72 |
| SH3BGRL3 | ENSG00000142669 | cMono.cyc_minor | 0.48 | 0.75 | 0.9 | 5.39e-62 | 2.26e-58 |
| MCTP2 | ENSG00000140563 | cMono_major | 1.29 | 2.27 | 3.14 | 5.49e-53 | 6.71e-49 |
| MCTP2 | ENSG00000140563 | cMono_minor | 1.3 | 2.27 | 3.15 | 7.32e-52 | 8.91e-48 |
| MCTP2 | ENSG00000140563 | MNP_celltype | 1.2 | 2.22 | 3.11 | 1.92e-51 | 2.35e-47 |

Table 3: Top 10 differentially expressed genes (across all cell clusters) for each contrast. logFC = log fold change, QValue=Benjamini-Hochberg adjusted p-valued. Cell clusters ending in _minor are minor subsets, _major are major subsets and _celltype are celltypes.

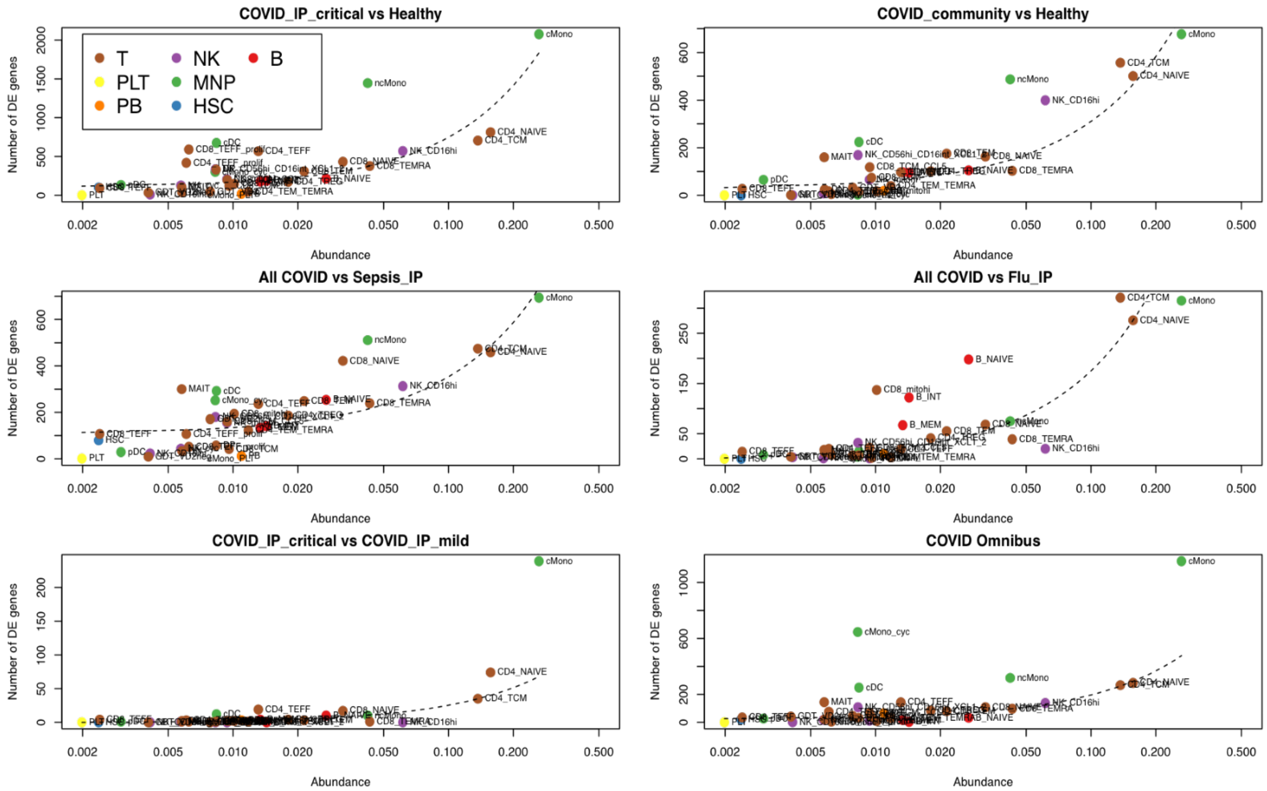

Figure 44: The relationship between mean abudance of minor subsets (i.e. the proportion of all cells that belong to this subset across all samples) and number of differentially expressed genes or a range of contrasts. Points are coloured by cell type and labelled with the minor subset name.

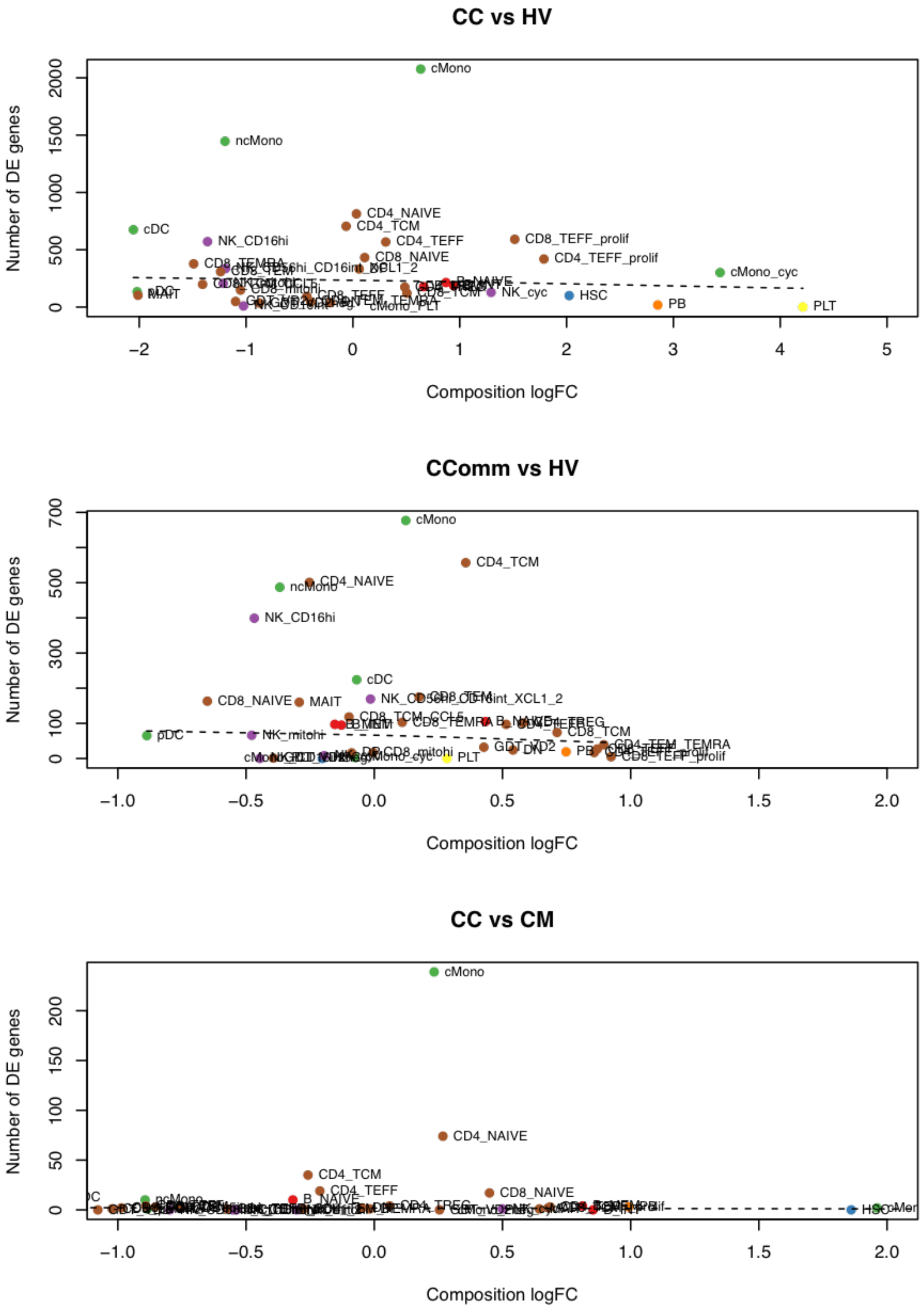

Figure 45: The relationship between differential abundance (measured by log-fold change in cell frequency between the two contrasts) and number of differentially expressed genes for each minor subset. P Points are coloured by cell type (using the same colour scheme as Figure 42) and labelled with the minor subset name.

#### Pathway Analysis

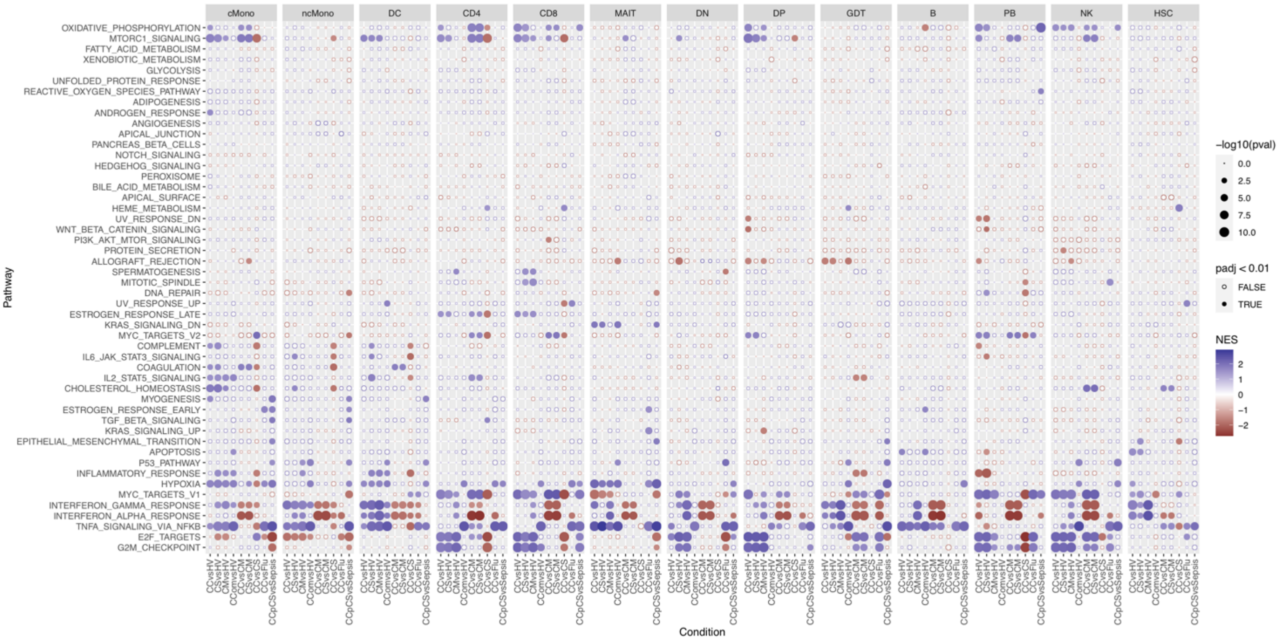

Figure 46: Hallmark gene set enrichment statistics for each pair of contrast and major cell subset. NES = Normalized effect size, padj = Benjamini-Hochberg-corrected p-value. Hallmark genesets downloaded from MSigDB.

| GO Term | pval | NES | celltype |
|---|---|---|---|
| GO_COTRANSLATIONAL_PROTEIN_TARGETING_TO_MEMBRANE | 1e-10 | 1.96 | GDT_VD2neg_minor |
| GO_CYTOSOLIC_RIBOSOME | 1e-10 | 1.96 | GDT_VD2neg_minor |
| GO_COTRANSLATIONAL_PROTEIN_TARGETING_TO_MEMBRANE | 1e-10 | 1.95 | GDT_major |
| GO_NUCLEAR_TRANSCRIBED_MRNA_CATABOLIC_PROCESS_NONSENSE_MEDIATED_DECAY | 1e-10 | 1.95 | GDT_VD2neg_minor |
| GO_CYTOSOLIC_RIBOSOME | 1e-10 | 1.94 | GDT_major |
| GO_ESTABLISHMENT_OF_PROTEIN_LOCALIZATION_TO_ENDOPLASMIC_RETICULUM | 1e-10 | 1.94 | GDT_major |
| GO_ESTABLISHMENT_OF_PROTEIN_LOCALIZATION_TO_ENDOPLASMIC_RETICULUM | 1e-10 | 1.94 | GDT_VD2neg_minor |
| GO_PROTEIN_LOCALIZATION_TO_ENDOPLASMIC_RETICULUM | 1e-10 | 1.93 | GDT_VD2neg_minor |
| GO_NUCLEAR_TRANSCRIBED_MRNA_CATABOLIC_PROCESS_NONSENSE_MEDIATED_DECAY | 1e-10 | 1.92 | GDT_major |
| GO_PROTEIN_LOCALIZATION_TO_ENDOPLASMIC_RETICULUM | 1e-10 | 1.91 | GDT_major |

| Canonical pathway | pval | NES | celltype |
|---|---|---|---|
| REACTOME_EUKARYOTIC_TRANSLATION_ELONGATION | 1e-10 | 2 | GDT_VD2neg_minor |
| KEGG_RIBOSOME | 1e-10 | 1.99 | GDT_VD2neg_minor |
| REACTOME_RESPONSE_OF_EIF2AK4_GCN2_TO_AMINO_ACID_DEFICIENCY | 1e-10 | 1.98 | GDT_VD2neg_minor |
| REACTOME_SELENOAMINO_ACID_METABOLISM | 1e-10 | 1.97 | GDT_VD2neg_minor |
| KEGG_RIBOSOME | 1e-10 | 1.96 | GDT_major |
| REACTOME_EUKARYOTIC_TRANSLATION_ELONGATION | 1e-10 | 1.96 | GDT_major |
| REACTOME_NONSENSE_MEDIATED_DECAY_NMD | 1e-10 | 1.96 | GDT_VD2neg_minor |
| REACTOME_SRP_DEPENDENT_COTRANSLATIONAL_PROTEIN_TARGETING_TO_MEMBRANE | 1e-10 | 1.96 | GDT_VD2neg_minor |
| REACTOME_RESPONSE_OF_EIF2AK4_GCN2_TO_AMINO_ACID_DEFICIENCY | 1e-10 | 1.95 | GDT_major |
| REACTOME_EUKARYOTIC_TRANSLATION_INITIATION | 1e-10 | 1.94 | GDT_VD2neg_minor |

| immunologic signature | pval | NES | celltype |
|---|---|---|---|

| | | | |
|---|---|---|---|
| GSE2405_0H_VS_24H_A_PHAGOCYTOPHILUM_STIM_NEUTROPHIL_UP | 1e-10 | 1.82 | GDT_VD2neg_minor |
| GSE2405_0H_VS_9H_A_PHAGOCYTOPHILUM_STIM_NEUTROPHIL_DN | 1e-10 | 1.81 | GDT_VD2neg_minor |
| GSE2405_0H_VS_24H_A_PHAGOCYTOPHILUM_STIM_NEUTROPHIL_UP | 1e-10 | 1.79 | GDT_major |
| GSE41978_KLRG1_HIGH_VS_LOW_EFFECTOR_CD8_TCELL_DN | 1e-10 | 1.77 | GDT_major |
| GSE41978_ID2_KO_VS_ID2_KO_AND_BIM_KO_KLRG1_LOW_EFFECTOR_CD8_TCELL_DN | 1e-10 | 1.76 | GDT_major |
| GSE2405_0H_VS_9H_A_PHAGOCYTOPHILUM_STIM_NEUTROPHIL_DN | 1e-10 | 1.75 | GDT_major |
| GSE45365_HEALTHY_VS_MCMV_INFECTION_CD11B_DC_DN | 1e-10 | 1.74 | CD8_major |
| GSE15750_DAY6_VS_DAY10_EFF_CD8_TCELL_UP | 1e-10 | 1.73 | CD8_major |
| GSE45365_WT_VS_IFNAR_KO_BCELL_DN | 1e-10 | 1.73 | CD8_major |
| GSE45365_WT_VS_IFNAR_KO_CD11B_DC_MCMV_INFECTION_DN | 1e-10 | 1.71 | CD8_major |

Table 4: Top 10 differentially expressed pairs of cell cluster and GO term, canonical pathway and immunologic signature (ranked by p-value and then NES) for critical COVID vs healthy controls. NES = normalized effect size. Further details of gene set definitions can be found on https://www.gsea-msigdb.org/gsea/msigdb/.

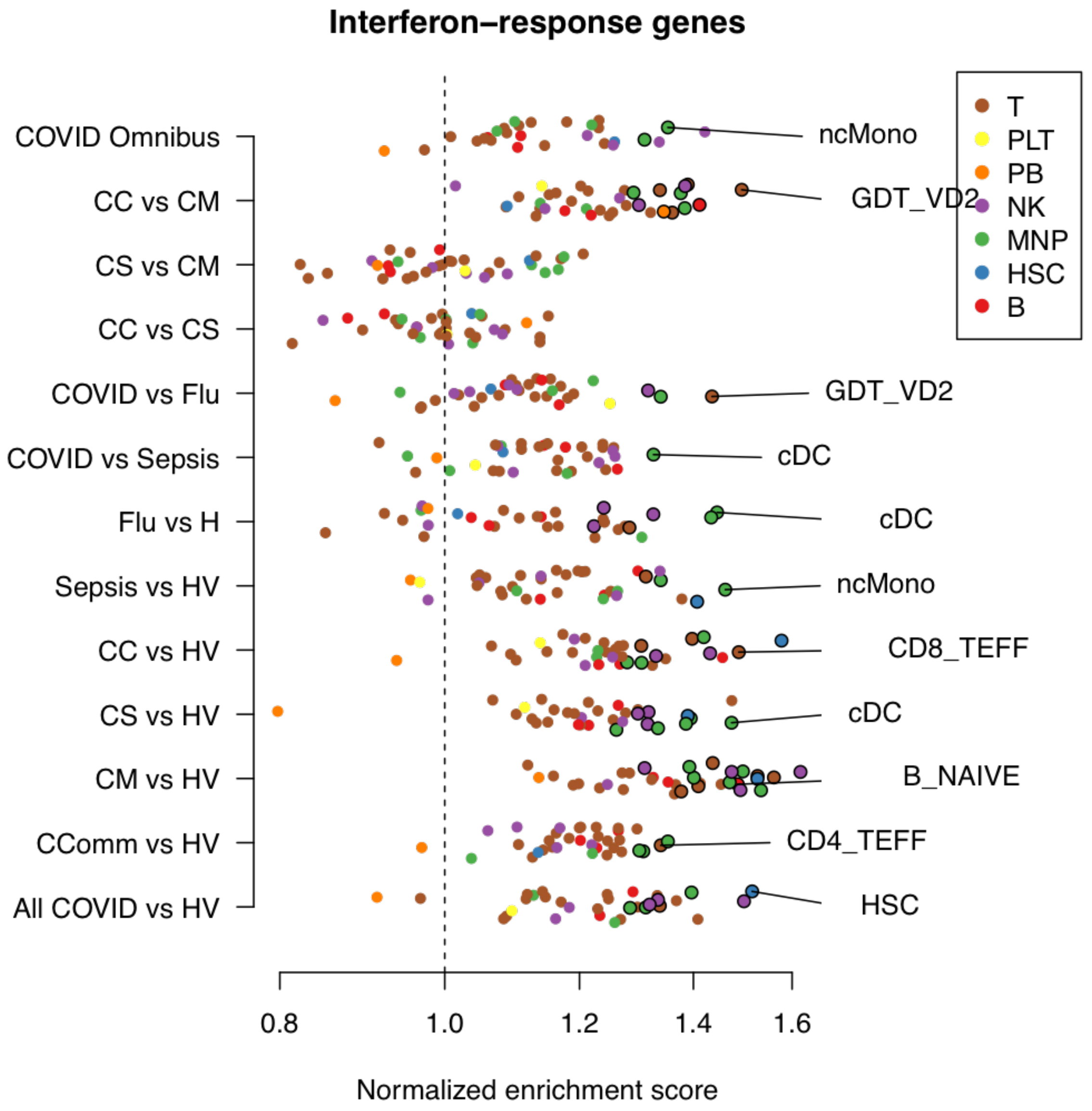

Figure 47: Enrichment of interferon-stimulated genes for each pair of minor subset and contrasts. Circled dots have p < 1e-5 (Bonferroni-corrected threshold for the number of subsets/contrasts pairs). The most significant cell subset is highlighted for each contrast

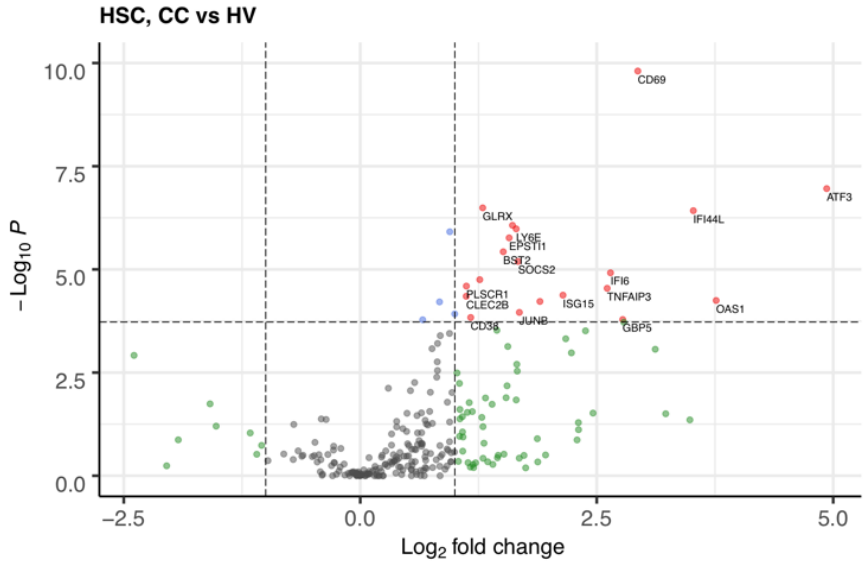

Figure 48: Volcano plot of differential expression between critical COVID and healthy controls, restricted to interferon-stimulated genes, in the HSC minor subset.

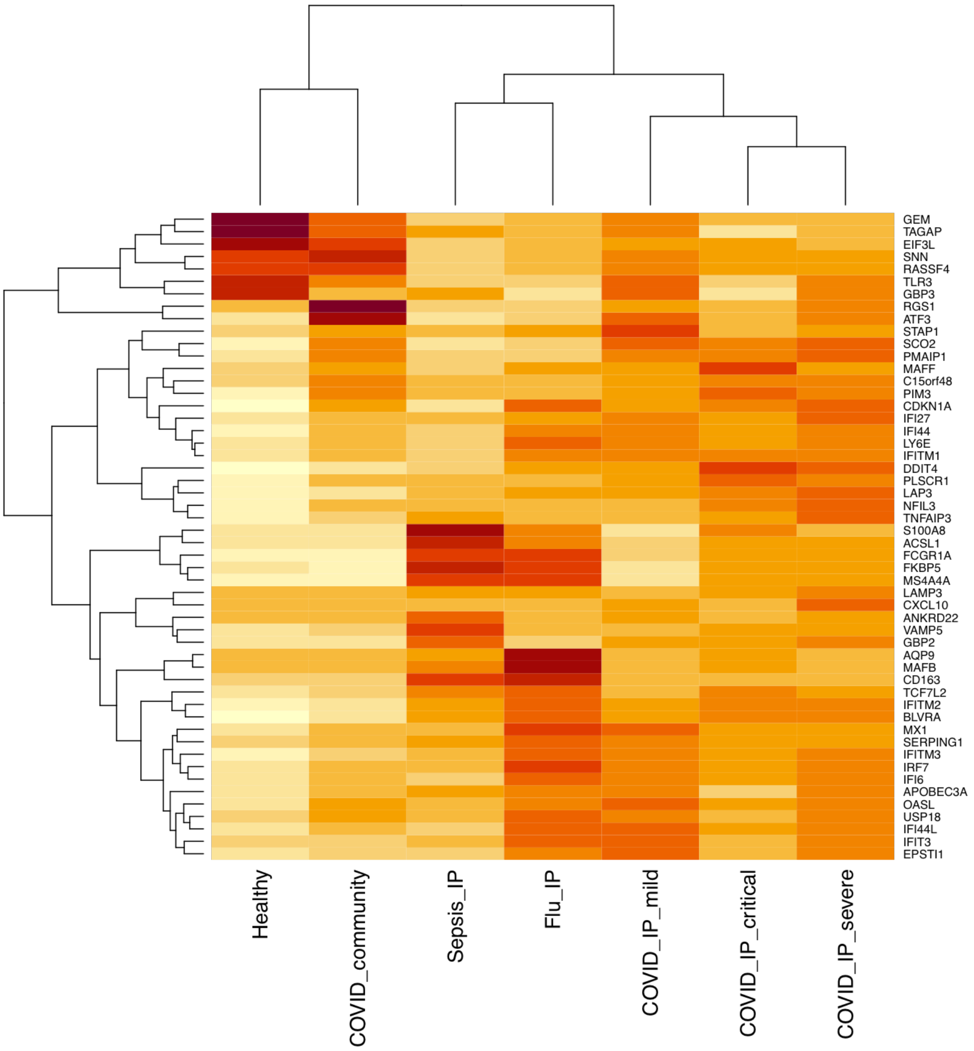

Figure 49: A hierarchically clustered heatmap of gene expression in classical dendritic cells (cDCs) of highly differentially expressed (FDR < 0.001, absolute fold change > 3) genes from the leading edges of interferon stimulated gene sets. Colour showed mean zero-centered RPM in units of standard deviations within each group.

#### WGCNA analysis

| WGCNA parameter | NK cells | cMono | ncMono | PB | B cells | CD4 T cells | CD8 T cells |
|---|---|---|---|---|---|---|---|
| network type | signed hybrid | signed hybrid | signed hybrid | signed hybrid | signed hybrid | signed hybrid | signed hybrid |
| adjacency correlation function | bicor | bicor | bicor | bicor | bicor | bicor | bicor |
| adjacency distance function | dist | dist | dist | dist | dist | dist | dist |
| TOM type | signed | signed | signed | signed | signed | signed | signed |
| minFraction (goodSamplesGenes) | 0.5 | 0.5 | 0.5 | 0.5 | 0.5 | 0.5 | 0.5 |
| minNSamples (goodSamplesGenes) | 4 | 4 | 4 | 4 | 4 | 4 | 4 |
| minNGenes (goodSamplesGenes) | 4 | 4 | 4 | 4 | 4 | 4 | 4 |
| minRelativeWeight (goodSamplesGenes) | 0.1 | 0.1 | 0.1 | 0.1 | 0.1 | 0.1 | 0.1 |
| minSize (cutreeStatic) | 10 | 10 | 10 | 10 | 10 | 10 | 10 |
| cutHeight (cutreeStatic) | 125 | 100 | 200 | 156 | 200 | 100 | 120 |
| Soft power (adjacency) | 3 | 6 | 4 | 5 | 4 | 6 | 6 |
| minimum module size (cutreeDynamic) | 50 | 50 | 50 | 50 | 50 | 50 | 50 |
| cutHeight (mergeCloseModules) | 0.25 | 0.4 | 0.25 | 0.1 | 0.25 | 0.25 | 0.25 |

Table 5: WGCNA parameters

| major cell type | module color | assigned name |
|---|---|---|
| B cell | cyan | cycling |
| B cell | greenyellow | ER_stress |
| B cell | purple | IFN_resp |
| B cell | pink | integrin_sig |
| B cell | turquoise | IRF2.ZF-C2H2 |
| B cell | green | mature.B.cell |
| B cell | midnightblue | mito_translation |
| B cell | black | OxPhos |
| B cell | blue | p38MAPK.AP1 |
| B cell | magenta | ribosomal |
| B cell | grey | unassigned_genes |
| B cell | brown | wnt.notch |
| CD4 T cell | lightgreen | ambient_rna |
| CD4 T cell | black | cycling |
| CD4 T cell | pink | IFN_resp |
| CD4 T cell | cyan | integrin_sig |
| CD4 T cell | turquoise | IRF2.ZF-C2H2 |
| CD4 T cell | lightyellow | JAK_STAT.IL_sig |
| CD4 T cell | green | p38MAPK.AP1 |
| CD4 T cell | midnightblue | ribosomal |
| CD4 T cell | grey | unassigned_genes |
| CD8 T cell | lightcyan | ambient_rna |
| CD8 T cell | blue | cycling |
| CD8 T cell | midnightblue | IFN_resp |
| CD8 T cell | yellow | IRF2.ZF-C2H2 |
| CD8 T cell | purple | mito_translation |
| CD8 T cell | pink | NK_activation |
| CD8 T cell | black | p38MAPK.AP1 |
| CD8 T cell | magenta | ribosomal |
| CD8 T cell | grey | unassigned_genes |
| CD8 T cell | cyan | vesicle_loc |
| cMono | grey60 | cycling |
| cMono | lightgreen | FKBP5.CD163 |
| cMono | black | IFN_resp |
| cMono | turquoise | IRF2.ZF-C2H2 |
| cMono | tan | OxPhos |
| cMono | lightcyan | p38MAPK.AP1 |
| cMono | green | ribosomal |
| cMono | grey | unassigned_genes |
| ncMono | yellow | cell_adhesion |
| ncMono | greenyellow | cell_growth |

| | | |
|---|---|---|
| ncMono | blue | Cilium.PECAM1 |
| ncMono | black | IC_Sig |
| ncMono | green | IFN_resp |
| ncMono | pink | IL-1_pathway |
| ncMono | purple | IRF2.ZF-C2H2 |
| ncMono | salmon | OxPhos |
| ncMono | magenta | p38MAPK.AP1 |
| ncMono | brown | ribosomal |
| ncMono | grey | unassigned_genes |
| NK cell | salmon | ambient_rna |
| NK cell | blue | cycling |
| NK cell | red | ECM_organisation |
| NK cell | purple | IFN_resp |
| NK cell | turquoise | IRF2.ZF-C2H2 |
| NK cell | green | ITK.STAT4 |
| NK cell | tan | leuk_prolif |
| NK cell | black | mito_translation |
| NK cell | brown | p38MAPK.AP1 |
| NK cell | greenyellow | ribosomal |
| PB | turquoise | cycling |
| PB | brown | IFN_resp |
| PB | blue | ribosomal |
| PB | grey | unassigned_genes |

Table 6: The identified WGCNA modules (from separate WCGNA analyses of each of the “major cell types”).
